# Hyperglycemia in Acute COVID-19 is Characterized by Adipose Tissue Dysfunction and Insulin Resistance

**DOI:** 10.1101/2021.03.21.21254072

**Authors:** Moritz Reiterer, Mangala Rajan, Nicolás Gómez-Banoy, Jennifer D. Lau, Luis G. Gomez-Escobar, Ankit Gilani, Sergio Alvarez-Mulett, Evan T. Sholle, Vasuretha Chandar, Yaron Bram, Katherine Hoffman, Alfonso Rubio-Navarro, Skyler Uhl, Alpana P. Shukla, Parag Goyal, Benjamin R. tenOever, Laura C. Alonso, Robert E. Schwartz, Edward J. Schenck, Monika M. Safford, James C. Lo

**Affiliations:** Weill Center for Metabolic Health, Cardiovascular Research Institute, Division of Cardiology, Department of Medicine, Weill Cornell Medicine, New York, NY, USA; Department of Medicine, Weill Cornell Medicine, New York, NY, USA; Division of Pulmonary and Critical Care Medicine, Department of Medicine, Weill Cornell Medicine, New York, NY, USA; Information Technologies & Services Department, Weill Cornell Medicine, New York, NY, USA; Weill Center for Metabolic Health, Division of Endocrinology, Diabetes and Metabolism, Department of Medicine, Weill Cornell Medicine, New York, NY, USA; Division of Gastroenterology and Hepatology, Department of Medicine, Weill Cornell Medicine, New York, NY, USA; Department of Microbiology, Icahn School of Medicine at Mount Sinai, New York, NY, USA; Division of Gastroenterology and Hepatology, Departments of Medicine and Physiology, Biophysics and Systems Biology, Weill Cornell Medicine, New York, NY, USA

## Abstract

COVID-19 has proven to be a metabolic disease resulting in adverse outcomes in individuals with diabetes or obesity. Patients infected with SARS-CoV-2 and hyperglycemia suffer from longer hospital stays, higher risk of developing acute respiratory distress syndrome (ARDS), and increased mortality compared to those who do not develop hyperglycemia. Nevertheless, the pathophysiological mechanism(s) of hyperglycemia in COVID-19 remains poorly characterized. Here we show that insulin resistance rather than pancreatic beta cell failure is the prevalent cause of hyperglycemia in COVID-19 patients with ARDS, independent of glucocorticoid treatment. A screen of protein hormones that regulate glucose homeostasis reveals that the insulin sensitizing adipokine adiponectin is reduced in hyperglycemic COVID-19 patients. Hamsters infected with SARS-CoV-2 also have diminished expression of adiponectin. Together these data suggest that adipose tissue dysfunction may be a driver of insulin resistance and adverse outcomes in acute COVID-19.

---

The deadly COVID-19 pandemic is underscored by the high morbidity and mortality rates seen in certain vulnerable populations, including patients with diabetes mellitus (DM), obesity, cardiovascular disease, and advanced age, with the latter associated with many chronic cardiometabolic diseases^1–4^. Hyperglycemia with or without a history of DM is a strong predictor of in-hospital adverse outcomes, portending a 7-fold higher mortality compared to patients with well-controlled blood glucose levels^5^. Hyperglycemia may be seen as a biomarker that predicts poor prognosis. A retrospective study that compared hyperglycemic patients that were treated with insulin against those who were not showed increased mortality in those receiving insulin^6^. However, it remains unclear whether insulin treatment is a surrogate for increased hyperglycemia and overall morbidity, or whether it is an actual causative factor for death. There is thus uncertainty regarding specific treatments for hyperglycemia in acute COVID-19^7^.

Despite our early recognition of the association between hyperglycemia and perilous outcomes, the pathophysiological mechanisms that underlie hyperglycemia in COVID-19 remain undefined^8,9^. Hypotheses have included a broad range of pathologies from direct infection of islets leading to beta cell failure (BCF) and to inflammation and glucocorticoids leading to insulin resistance (IR). Although COVID-19 is primarily a respiratory tract infection, SARS-CoV-2 is known to infect other cell types and often leads to extrapulmonary consequences^10,11^. *ACE2* and other entry receptors for SARS-CoV-2 can be expressed on pancreatic islet cells and endocrine cells differentiated from human pluripotent stem cells are permissive to infection^12^. Early reports of unexpected diabetic ketoacidosis (DKA) in COVID-19 patients fuelled concerns for a novel form of acute onset beta cell failure. For example, one case described a patient with new onset diabetic ketoacidosis (DKA) who was found to be autoantibody negative for type 1 DM (T1DM) but showed evidence of prior SARS-CoV-2 infection based on serology results, suggesting the possibility of pancreatic beta cell dysfunction or destruction as a result of COVID-19^13^. However, given the high rates of COVID-19 during this pandemic coupled with low background rates of new onset T1DM, the connection between these two events in this case could be “true, true, and unrelated.” Recent studies disagree on whether ACE2 is expressed on pancreatic beta cells or whether the SARS-CoV-2 virus is found in pancreatic beta cells of deceased individuals with COVID-19^14–16^. Conversely, the well-known connection between obesity and insulin resistance might lead to impaired immunity and more severe SARS-CoV-2 infection^17^. In fact, population level studies have reported higher risk of complications in obese patients with COVID-19^18–20^. Viral infection may lead to systemic insulin resistance and worsened hyperglycemia. In sum, despite much attention, the pathophysiology of hyperglycemia in COVID-19 remains unknown.

Dexamethasone substantially reduces mortality in patients with severe COVID-19 infection requiring oxygen or invasive mechanical ventilation^21^. Glucocorticoids can also provoke hyperglycemia by inducing insulin resistance and beta cell dysfunction. The widespread usage of dexamethasone in severe SARS-CoV-2 infection is sure to exacerbate both the incidence and severity of hyperglycemia in COVID-19.

In this study, we assessed the pathophysiological mechanism of hyperglycemia in acute and severe COVID-19 and analyzed protein hormones regulating glucose homeostasis. We compared COVID-19 patients to critically ill control patient groups and found striking differences in the characteristics associated with hyperglycemia, further highlighting the metabolic dysfunction seen in this disease. Our study consisted of patients hospitalized at New York-Presbyterian Hospital/Weill Cornell Medical Center and affiliated campuses at Queens and Lower Manhattan Hospital. We assessed the characteristics of 4,102 inpatients diagnosed with COVID-19 between Mar 3, 2020 and May 15, 2020 and found a clear association between hyperglycemia (glucose > 170 mg/dl) and adverse outcomes. Overall, 47% of all COVID-19 patients were hyperglycemic but disproportionately accounted for 90% of intubations and 72% of deaths (Table 1). Similarly, hyperglycemic patients had a more than 2-fold increase in the average length of hospital stay, 10 compared to 5 days for non-hyperglycemic patients.

**Table 1:**
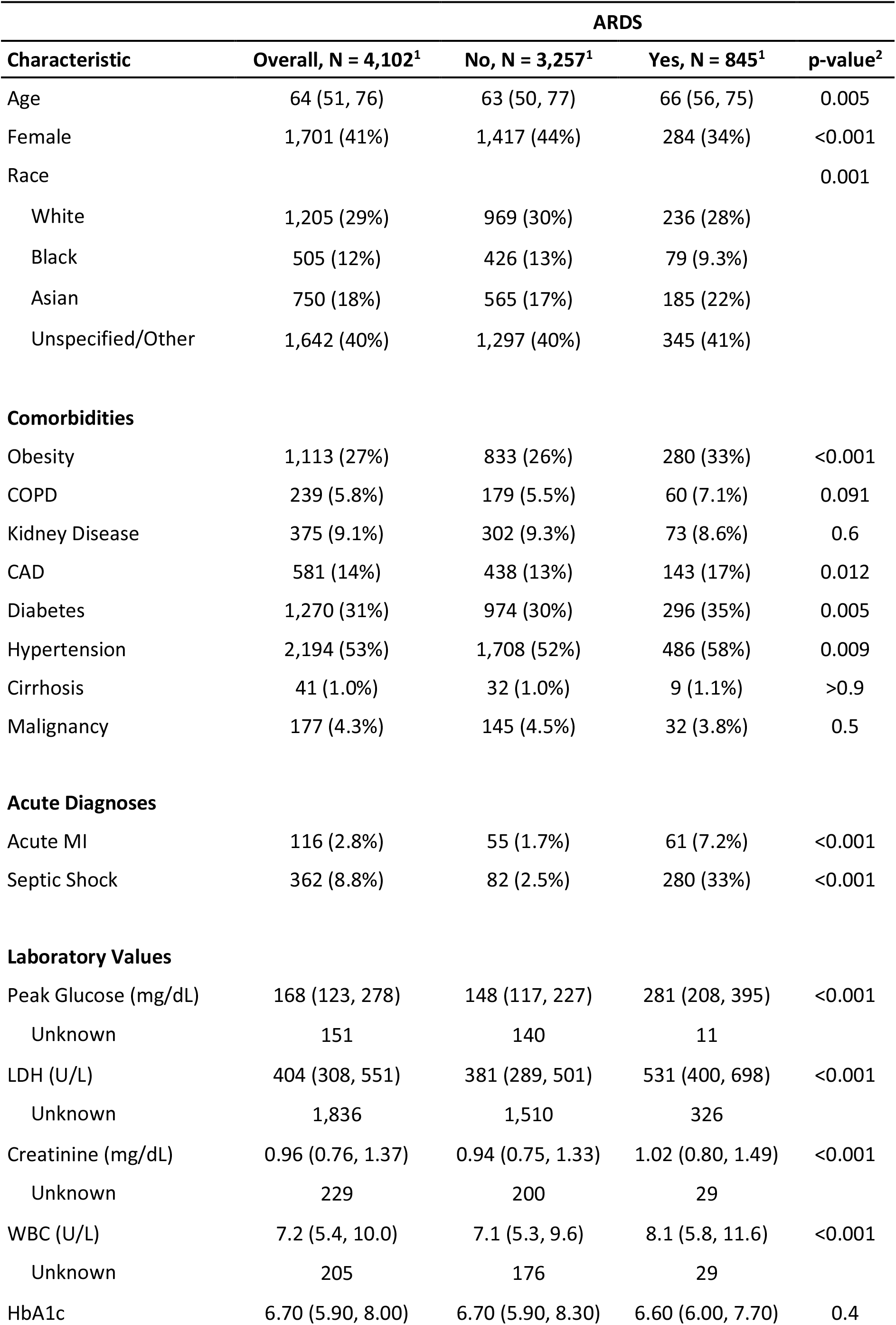
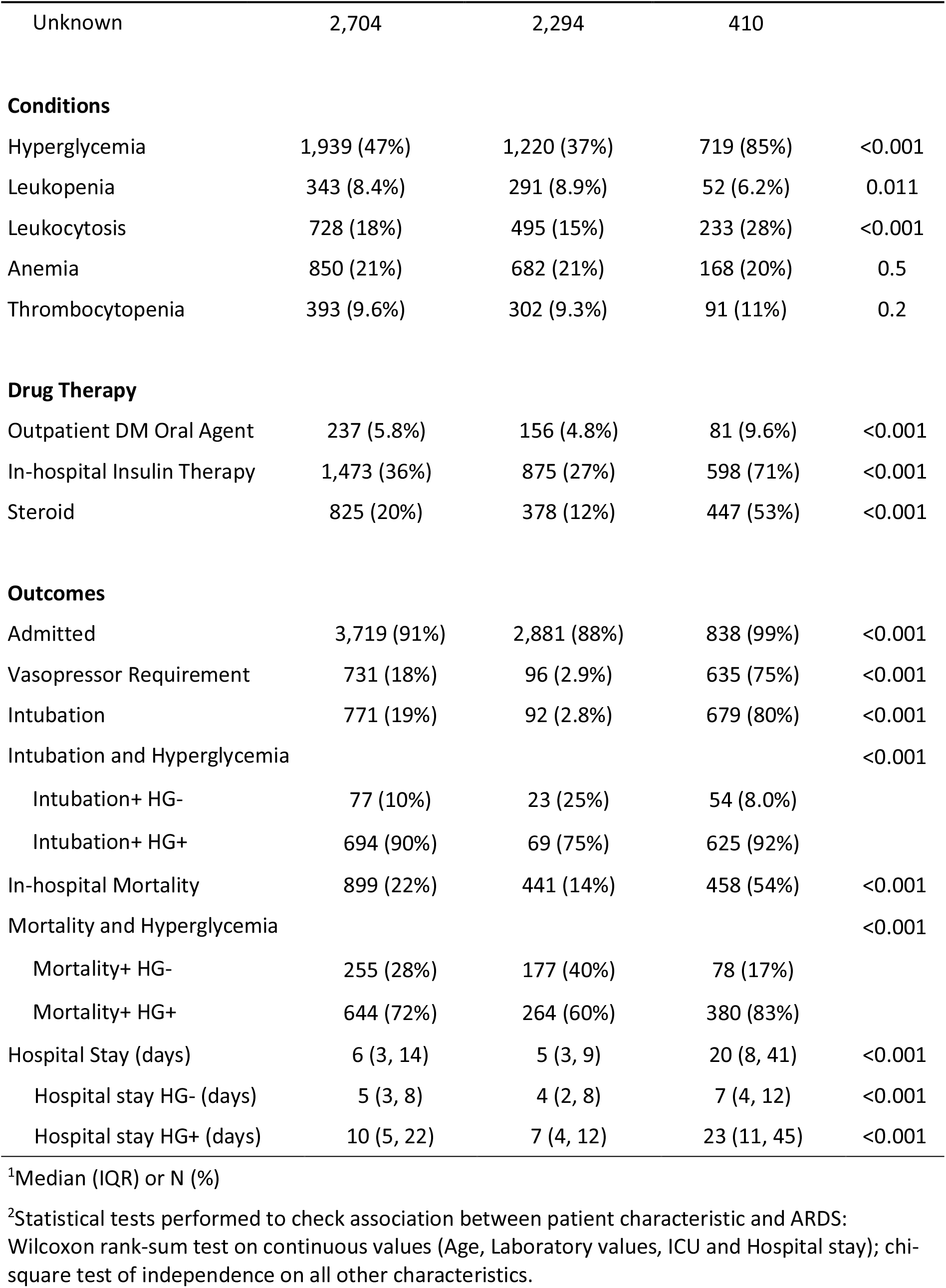
Characteristics of COVID patients, March 1, 2020 to May 15, 2020.

| Characteristic | Overall, N = 4,102<sup>1</sup> | ARDS |  | p-value<sup>2</sup> |
| --- | --- | --- | --- | --- |
|  |  | No, N = 3,257<sup>1</sup> | Yes, N = 845<sup>1</sup> |  |
| Age | 64 (51, 76) | 63 (50, 77) | 66 (56, 75) | 0.005 |
| Female | 1,701 (41%) | 1,417 (44%) | 284 (34%) | <0.001 |
| Race |  |  |  | 0.001 |
| White | 1,205 (29%) | 969 (30%) | 236 (28%) |  |
| Black | 505 (12%) | 426 (13%) | 79 (9.3%) |  |
| Asian | 750 (18%) | 565 (17%) | 185 (22%) |  |
| Unspecified/Other | 1,642 (40%) | 1,297 (40%) | 345 (41%) |  |
| **Comorbidities** |  |  |  |  |
| Obesity | 1,113 (27%) | 833 (26%) | 280 (33%) | <0.001 |
| COPD | 239 (5.8%) | 179 (5.5%) | 60 (7.1%) | 0.091 |
| Kidney Disease | 375 (9.1%) | 302 (9.3%) | 73 (8.6%) | 0.6 |
| CAD | 581 (14%) | 438 (13%) | 143 (17%) | 0.012 |
| Diabetes | 1,270 (31%) | 974 (30%) | 296 (35%) | 0.005 |
| Hypertension | 2,194 (53%) | 1,708 (52%) | 486 (58%) | 0.009 |
| Cirrhosis | 41 (1.0%) | 32 (1.0%) | 9 (1.1%) | >0.9 |
| Malignancy | 177 (4.3%) | 145 (4.5%) | 32 (3.8%) | 0.5 |
| **Acute Diagnoses** |  |  |  |  |
| Acute MI | 116 (2.8%) | 55 (1.7%) | 61 (7.2%) | <0.001 |
| Septic Shock | 362 (8.8%) | 82 (2.5%) | 280 (33%) | <0.001 |
| **Laboratory Values** |  |  |  |  |
| Peak Glucose (mg/dL) | 168 (123, 278) | 148 (117, 227) | 281 (208, 395) | <0.001 |
| Unknown | 151 | 140 | 11 |  |
| LDH (U/L) | 404 (308, 551) | 381 (289, 501) | 531 (400, 698) | <0.001 |
| Unknown | 1,836 | 1,510 | 326 |  |
| Creatinine (mg/dL) | 0.96 (0.76, 1.37) | 0.94 (0.75, 1.33) | 1.02 (0.80, 1.49) | <0.001 |
| Unknown | 229 | 200 | 29 |  |
| WBC (U/L) | 7.2 (5.4, 10.0) | 7.1 (5.3, 9.6) | 8.1 (5.8, 11.6) | <0.001 |
| Unknown | 205 | 176 | 29 |  |
| HbA1c | 6.70 (5.90, 8.00) | 6.70 (5.90, 8.30) | 6.60 (6.00, 7.70) | 0.4 |

| ARDS |  |  |  |  |
| --- | --- | --- | --- | --- |
| Characteristic | Overall, N = 4,102<sup>1</sup> | No, N = 3,257<sup>1</sup> | Yes, N = 845<sup>1</sup> | p-value<sup>2</sup> |
| Unknown | 2,704 | 2,294 | 410 |  |
| **Conditions** |  |  |  |  |
| Hyperglycemia | 1,939 (47%) | 1,220 (37%) | 719 (85%) | <0.001 |
| Leukopenia | 343 (8.4%) | 291 (8.9%) | 52 (6.2%) | 0.011 |
| Leukocytosis | 728 (18%) | 495 (15%) | 233 (28%) | <0.001 |
| Anemia | 850 (21%) | 682 (21%) | 168 (20%) | 0.5 |
| Thrombocytopenia | 393 (9.6%) | 302 (9.3%) | 91 (11%) | 0.2 |
| **Drug Therapy** |  |  |  |  |
| Outpatient DM Oral Agent | 237 (5.8%) | 156 (4.8%) | 81 (9.6%) | <0.001 |
| In-hospital Insulin Therapy | 1,473 (36%) | 875 (27%) | 598 (71%) | <0.001 |
| Steroid | 825 (20%) | 378 (12%) | 447 (53%) | <0.001 |
| **Outcomes** |  |  |  |  |
| Admitted | 3,719 (91%) | 2,881 (88%) | 838 (99%) | <0.001 |
| Vasopressor Requirement | 731 (18%) | 96 (2.9%) | 635 (75%) | <0.001 |
| Intubation | 771 (19%) | 92 (2.8%) | 679 (80%) | <0.001 |
| Intubation and Hyperglycemia |  |  |  | <0.001 |
| Intubation+ HG- | 77 (10%) | 23 (25%) | 54 (8.0%) |  |
| Intubation+ HG+ | 694 (90%) | 69 (75%) | 625 (92%) |  |
| In-hospital Mortality | 899 (22%) | 441 (14%) | 458 (54%) | <0.001 |
| Mortality and Hyperglycemia |  |  |  | <0.001 |
| Mortality+ HG- | 255 (28%) | 177 (40%) | 78 (17%) |  |
| Mortality+ HG+ | 644 (72%) | 264 (60%) | 380 (83%) |  |
| Hospital Stay (days) | 6 (3, 14) | 5 (3, 9) | 20 (8, 41) | <0.001 |
| Hospital stay HG- (days) | 5 (3, 8) | 4 (2, 8) | 7 (4, 12) | <0.001 |
| Hospital stay HG+ (days) | 10 (5, 22) | 7 (4, 12) | 23 (11, 45) | <0.001 |

<sup>1</sup>Median (IQR) or N (%)
<sup>2</sup>Statistical tests performed to check association between patient characteristic and ARDS: Wilcoxon rank-sum test on continuous values (Age, Laboratory values, ICU and Hospital stay); chi-square test of independence on all other characteristics.

Patients severely infected with SARS-CoV-2 develop pneumonia, ARDS, and require mechanical ventilation^22^. To further analyze the impact of hyperglycemia on critically ill COVID-19 patients, we divided our cohort based on the development of ARDS. Among COVID-19 patients with ARDS, hyperglycemia was considerably more prevalent than in patients without ARDS (85% compared to 37%, Table 1). This corresponded to a higher proportion of hyperglycemic patients among intubations (92% vs 75%) and deaths (83% vs 60%). Most strikingly, the median hospital stay of hyperglycemic COVID-19 ARDS patients was more than three times longer than for non-hyperglycemic individuals with the same condition (7 days vs 23 days). Among COVID-19 patients without ARDS, hyperglycemia was only associated with a 75% increase in median hospital stay (4 days vs 7 days).

To determine the mechanism behind hyperglycemia in patients with a severe acute SARS-CoV-2 infection, we performed a multiplex metabolic protein array targeting hormones known to modulate blood glucose homeostasis. We analyzed plasma samples from COVID-19 patients and ICU control groups from patients enrolled in our Biobank of the Critically Ill (BOCI)^23–25^. The characteristics of this cohort and the full list of targets including incretins, islet hormones and adipokines are shown in Extended Data Table 2 and described below. The hyperglycemia incidence within the profiled cohort is comparable to what was observed in the parental cohorts (Table 1 and Extended Data Tables 1, 2). COVID-19 ARDS patients showed a significantly higher rate of hyperglycemia than COVID-negative ARDS patients (Extended Data Fig. 1a). Pre-existing diabetes and body mass index (BMI) were not significantly different between the three groups (Extended Data Fig. 1b,c). Since glucocorticoids are part of the treatment for severe COVID-19^21,26^ and known to cause or exacerbate hyperglycemia, we assessed the rate of glucocorticoid treatment among the profiled cohort. There was no difference between COVID-19 ARDS patients and ARDS controls (80% vs 68%, p=0.35, Extended Data Fig. 1d). None of the profiled ARDS-controls received glucocorticoids. We also charted when glucocorticoids were administered to patients along with daily glucose values and insulin injections (Extended Data Fig. 2a-d and Supplementary Data File 1). Some patients were persistently hyperglycemic with or without glucocorticoids (Extended Data Fig. 2b). Others were initially hyperglycemic but their glucose normalized and remained below the hyperglycemia threshold until hospital discharge (Extended data Fig 2c). Similarly, some patients were not hyperglycemic for several days following admission but then experienced a rise in blood glucose which in some cases was associated with glucocorticoids (Extended data Fig. 2d). We quantified the incidence of each of these patterns within the three patient cohorts and found no significant difference in their distribution (Extended Data Fig. 2e,f).

Hyperglycemia can be caused by two distinct mechanisms: insulin resistance (IR) or beta cell failure (BCF) due to diminished beta cell mass and/or insufficient insulin secretion from failing beta cells^27^. IR is characterized by elevated blood insulin levels and hyperglycemia, as the pancreatic beta cells are still functional and attempt to overcome hyperglycemia by increasing insulin secretion. Individuals experiencing BCF on the other hand, are no longer able to secrete appropriate amounts of insulin, resulting in insulinopenia and severe hyperglycemia. To divide the hyperglycemic patients into these two subgroups, we assessed their serum C-peptide levels. As a by-product of insulin secretion by the pancreas, C-peptide is a direct indicator of endogenous insulin production and has a longer half-life compared to insulin. Insulin levels were also assessed but our assay could not distinguish between endogenous and therapeutically administered exogenous insulin. Many of the patients involved in this study received insulin treatment (Extended Data Fig. 2b-d and Supplementary Data File 1), confounding the insulin measurement results. We observed that COVID-19 ICU patients displayed significantly elevated levels of plasma C-peptide compared to either ARDS+ and ARDS-controls, who did not differ from each other (Fig. 1a and Extended Data Fig. 3a). This was independent of the higher rate of hyperglycemia in COVID-19 patients since it persisted even if only hyperglycemic patients from each cohort were compared (Fig. 1b and Extended Data Fig. 3b). Similarly, it was also independent of glucocorticoid treatment (Extended Data Fig. 3c). We also found an increase in amylin among COVID-19 patients (Fig 2d). Like C-peptide, this protein is co-secreted with insulin from pancreatic beta cells, further confirming the beta cell hyper-secretory phenotype present in a majority of COVID-19 ARDS+ patients.

**Fig 1.**
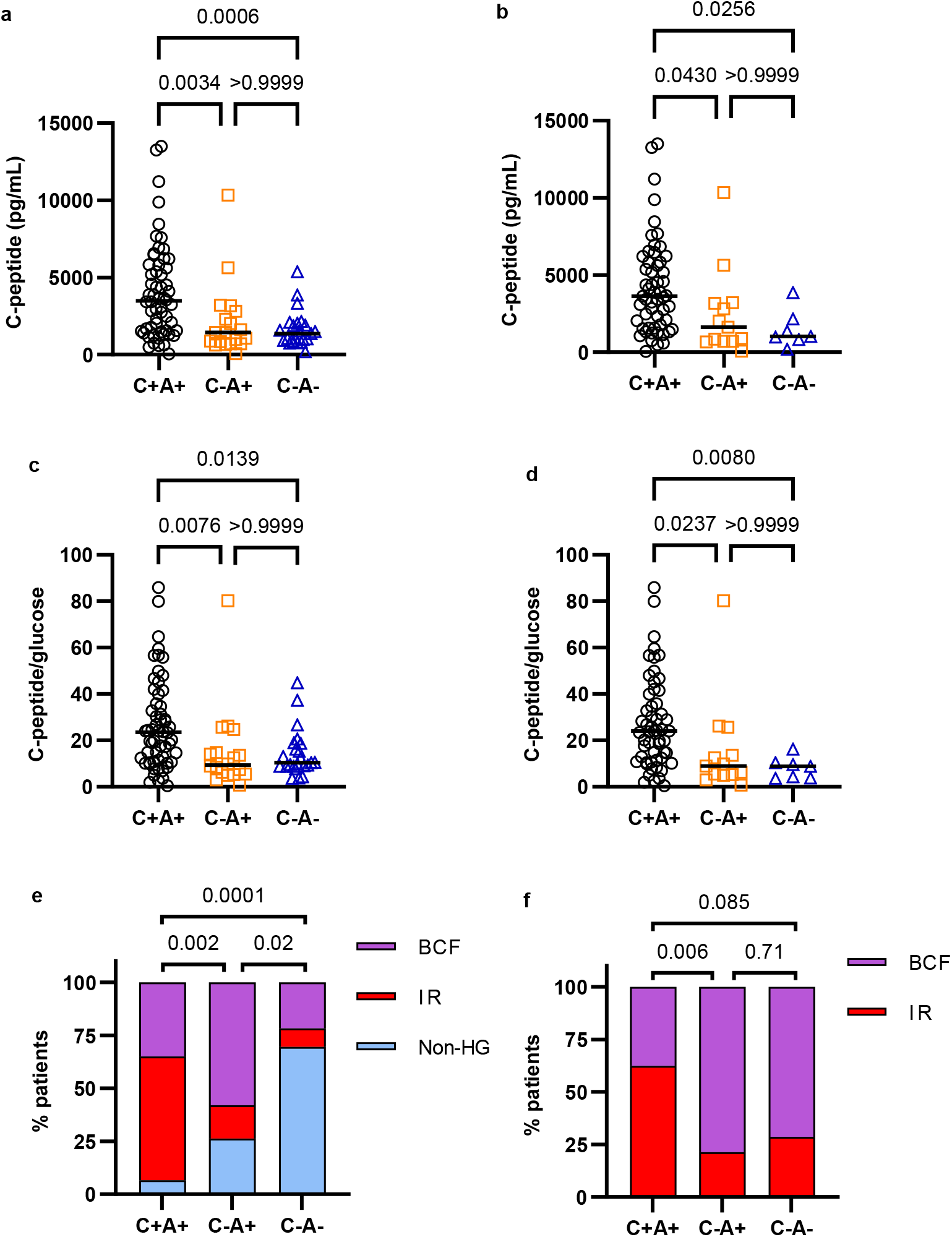
Hyperglycemia in COVID-19 presents predominantly through insulin resistance. **a**, C-peptide concentration in the plasma of COVID-19 ARDS patients (C+A+ n=59), ARDS+ ICU controls (C-A+ n=19), and ARDS-ICU controls (C-A-n=23), measured by Multiple Protein Array. Data were analyzed using Kruskal-Wallis and Dunn’s tests with multiple comparison correction. **b**, C-peptide concentration in the plasma of hyperglycemic patients and controls (C+A+ n=55, C-A+ n=13, C-A-n=7), measured by Multiple Protein Array. Data were analyzed using Kruskal-Wallis and Dunn’s tests with multiple comparison correction. **c, d**, C-peptide/glucose ratio of all (**c**) or hyperglycemic only (**d**) Covid-19 ARDS and control patients, C-peptide measured by Multiple Protein Array. Data were analyzed using Kruskal-Wallis and Dunn’s tests with multiple comparison correction. **e, f**, Subcategorization of Covid-19 ARDS and control patients as non-hyperglycemic (non-HG), insulin resistant (IR), beta cell failure (BCF). **e** shows the incidence of non-HG, IR, and BCF with respect to all patients. **f** shows the incidence of IR and BCF among HG patients. Data were analyzed using two-sided Chi-squared (**e**) or Fisher’s exact (**f**) test with Bonferroni-Holm’s correction for multiple comparisons.

**Fig 2.**
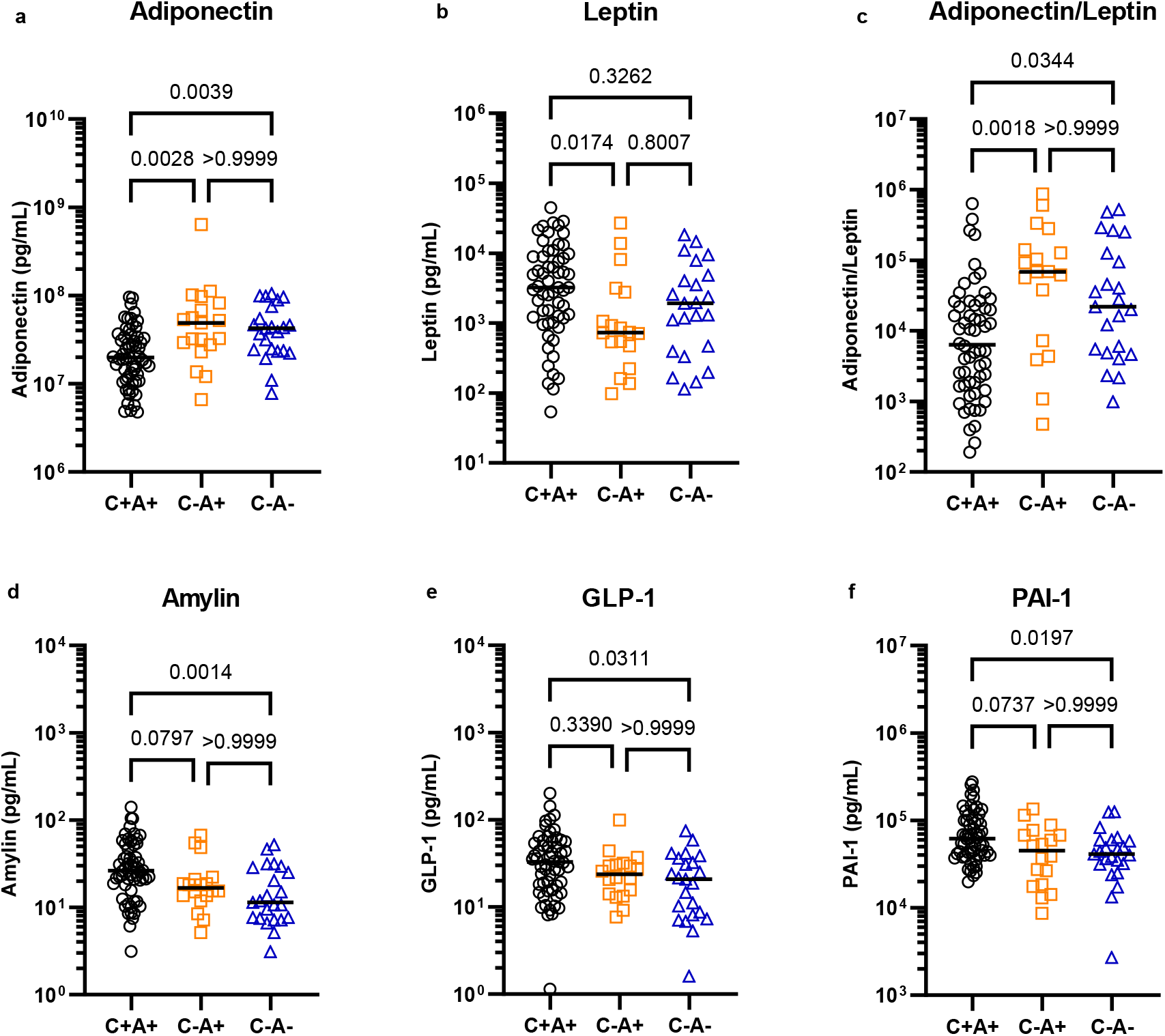
Altered metabolic hormone profiles in COVID-19. Plasma from COVID-19 ARDS (C+A+, n=59) and control ICU patients with ARDS (C-A+, n=19) and without ARDS (C-A-n=23) was analysed by multiplex metabolic protein array, targeting hormones known to modulate blood glucose homeostasis. The data were analyzed using Kruskal-Wallis tests. All targets with p<0.05 are shown here and were subjected to Dunn’s tests with multiple comparison correction.

C-peptide values were compared against glucose levels at or close to the plasma collection time to determine C-peptide/glucose ratios, which are an established marker of beta cell function^28–30^. An elevated C-peptide/glucose ratio coupled with hyperglycemia reflects insulin resistance. Similar to the raw C-peptide values, COVID-19 patients showed higher C-peptide/glucose ratios compared to ARDS+ and ARDS-controls, which did not differ significantly (Fig. 1c and Extended Data Fig. 3d). Again, this trend persisted even after restricting the comparison to only hyperglycemic patients within each cohort and was independent of glucocorticoid treatment (Fig. 1d and Extended Data Fig. 3e,f).

To dissect the mechanism of hyperglycemia in COVID-19 patients, hyperglycemic subjects were divided into subtypes reflecting a predominantly IR or BCF phenotype (Extended Data Fig. 3g-i). The proportion of subjects with IR was more than three times greater among COVID-19 patients than ARDS+ controls, and more than six times greater than ARDS-controls (Fig. 1e,f). Glucocorticoids are known to cause hyperglycemia, by inducing either IR or beta cell damage^31^. We found no difference in the incidence of glucocorticoid treatments among the IR and BCF subgroups within the COVID-19 or ARDS+ control patients (Extended Data Fig. 4). None of the patients assayed in the ARDS-control group received glucocorticoids. There was also no difference in the prevalence of obesity between BCF and IR subtypes in any of the three patient groups. A pre-existing diagnosis of type 2 diabetes on the other hand was significantly associated with BCF over IR in both the COVID-19 and ARDS+ control groups (Fisher’s exact test, p=0.01 and 0.03 respectively). This suggests that BCF occurs mostly in people with DM prior to COVID rather than developing *de novo* as a result of the viral disease. Given the higher incidence of glucocorticoid treatments in the COVID-19 ARDS+ group and DM among ARDS+ control patients (Extended Data Fig. 1b and Extended Data Fig.4), we constructed a logistic regression model that adjusted for these differences. We still found a significantly higher rate of IR in COVID-19 patients compared to both ARDS+ (OR 8.3, p=0.009) and ARDS-controls (OR 15.9, p<0.0001) (Extended Data Fig. 5). No difference was observed in the proportion of IR between the two controls, or in the distribution of BCF between any of the groups.

Beta cell function and insulin sensitivity are conditioned by endocrine stimuli between tissues. Over the last two decades, it has become increasingly clear that adipose tissue (AT) acts as a major source of such stimuli. Proteins/cytokines secreted from AT into the bloodstream, collectively called adipokines, have been shown to regulate beta cell function and mass and to modulate insulin sensitivity in peripheral tissues^32^. Dysfunctional adipokine secretion has been associated with metabolic disorders such as obesity, metabolic syndrome, and type 2 diabetes.

We therefore tested if hyperglycemia in COVID-19 patients is associated with aberrations in glucoregulatory hormones such as those arising from the islets, incretins, and adipokines. Indeed, the adipokine adiponectin was found to be decreased by 50-60% in the serum of patients suffering from severe COVID-19, compared to ICU controls with or without ARDS (Fig. 2a). The two controls did not differ from each other. Adiponectin is associated with anti-diabetic, anti-atherogenic, and anti-inflammatory properties and its levels are known to be lower in individuals suffering from obesity or type 2 diabetes. However, we found that the two control groups contained slightly higher percentages of both obese and diabetic patients than the COVID-19 group, suggesting that the decrease in adiponectin associated with COVID-19 may even be underestimated. We found no difference in the abundance of adiponectin among the IR and BCF hyperglycemic subgroups within COVID-19 patients (Extended Data Fig. 7g). Adipsin, an adipokine known to promote beta-cell survival and insulin secretion^33^, was also decreased among the COVID-19 patient group compared to ICU controls, although this difference was not statistically significant (overall p=0.06, Extended Data Fig. 6h). Leptin, an adipokine regulator of appetite and energy expenditure, was increased with respect to ARDS+ controls but not with respect to ARDS-(Fig. 2b). The adiponectin-leptin ratio, a biomarker of metabolic health and adipose function, was severely depressed in COVID-19 patients by 10-fold compared to ARDS+ and 6-fold compared to ARDS-control patients (Fig. 2c)^34^. PAI-1, an adipokine which acts as an inhibitor of fibrinolysis, was also found to be significantly increased with respect to ARDS-but not ARDS+ controls (Fig. 2f), as was reported previously for severe COVID-19 patients^35^. However, increased PAI-1 levels did not correlate with hyperglycemia (Extended Data Fig. 6g). We also found GLP-1, a stimulator of insulin secretion produced by intestinal L-cells, to be significantly increased in COVID-19 patients compared to ARDS-but not ARDS+ controls (Fig. 2e). Of note, IL-6, ghrelin, glucagon, GIP, pancreatic polypeptide (PP), PYY, resistin, and lipocalin-2 did not differ between the three groups (Extended Data Fig. 6,7). These results raise the possibility that COVID-19 causes adipocyte dysfunction, which may occur as a direct result of SARS-CoV-2 infecting adipocytes or other cells within the AT, or through other indirect mechanisms.

To test whether SARS-CoV-2 can cause direct adipocyte dysfunction, we measured the mRNA levels of adipokines within the AT of Syrian Hamsters following infection^36,37^. Indeed, *Adipoq* was robustly decreased by 80% in the subcutaneous adipose tissue (SAT) of SARS-CoV-2-infected hamsters compared to controls (Fig. 3a); in visceral adipose tissue (VAT), no significant difference was observed (Extended Data Fig. 8). *Lep* mRNA was also decreased in hamster fat after infection with SARS-CoV-2, a trend which was not mirrored by the human blood samples (Fig. 3a and 2b). Adiponectin protein was decreased with SARS-CoV-2 infection in subcutaneous and visceral adipose tissues, and serum compared to mock-infected controls (Fig. 3b-e). To determine whether the SARS-CoV-2 virus might infect the adipose tissue, we performed qPCR for the replicating strand of the viral nucleocapsid gene and found elevated levels of SARS-CoV-2 viral RNA in the fat of infected hamsters compared to mock controls (Fig. 3e). This suggests that cells within the adipose tissue may be permissive to viral infection and replication; which cell type remains to be determined.

**Fig 3.**
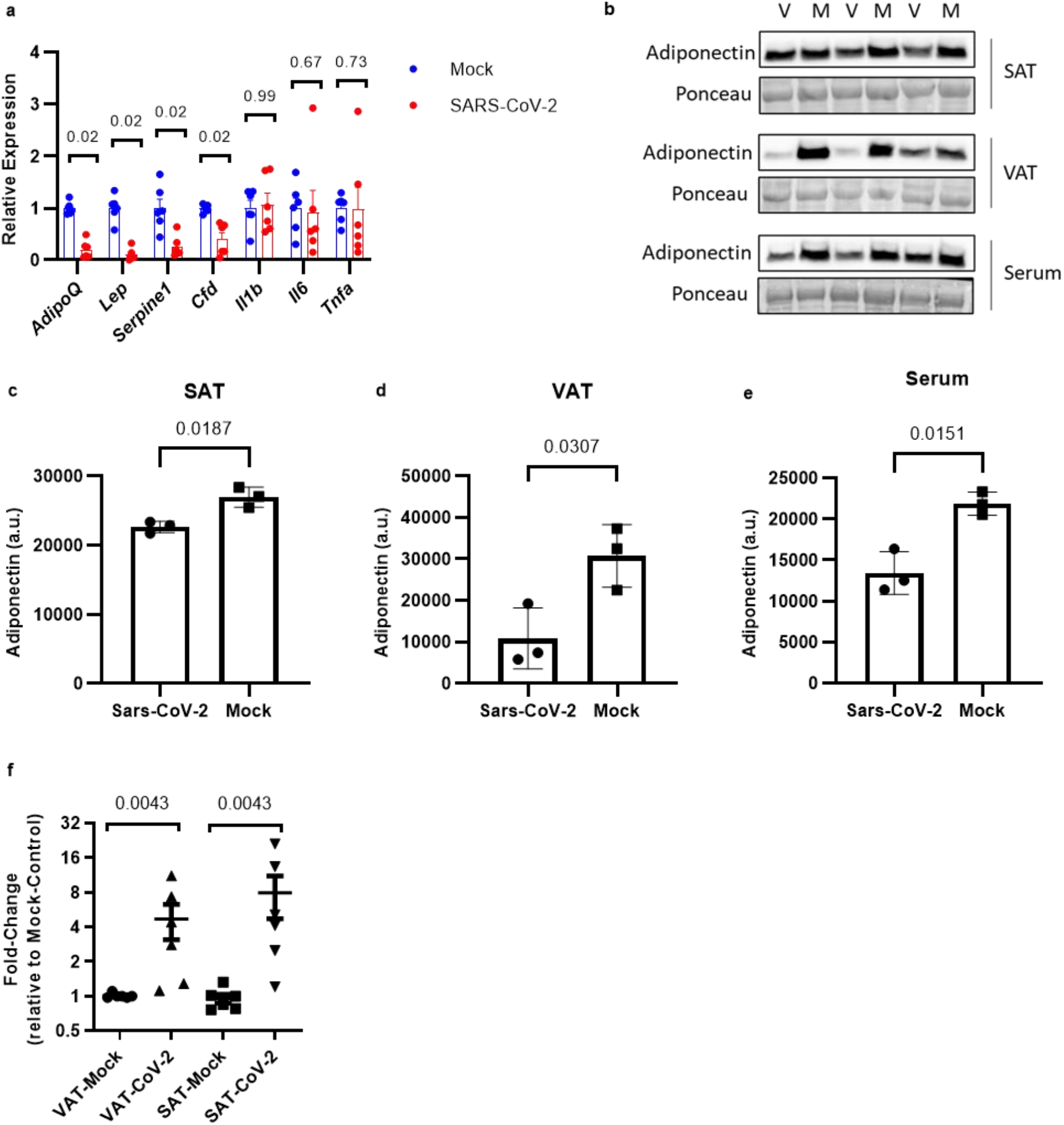
Sars-CoV-2 replicates in hamster adipose tissue and alters adipokine expression. **a**, RT-qPCR of mRNA from subcutaneous fat tissue of Syrian hamsters infected with Sars-CoV-2 or mock virus. Data were analyzed using Mann-Whitney tests with Holm-Sidak’s multiple comparisons correction (n=6 hamsters per group). **b**, Western blot for adiponectin within subcutaneous adipose tissue (SAT), visceral adipose tissue (VAT), or serum from Syrian hamsters infected with Sars-CoV-2 (V) or mock virus (M) (n=3 hamsters per group). **c-e**, quantification of the western blots shown in **b**. The data were analyzed using Welch’s t-test. **f** RT-qPCR for the replicating strand of the Sars-Cov-2 nucleocapsid gene using RNA from SAT and VAT from Syrian hamsters infected with Sars-CoV-2 or mock virus. Data were analyzed using Mann-Whitney tests with Holm-Sidak’s multiple comparisons correction (n=6 hamsters per group)

In conclusion, hyperglycemia is a clear and strong poor prognostic factor in COVID-19, borne out in multiple studies^38–40^. Yet the causes of hyperglycemia in this population remain unclear. To address this gap, we studied hyperglycemic patients suffering from severe SARS-CoV-2 infection as this group is expected to show the most dramatic metabolic effects from acute viral infection. Insulin resistance was the predominant mechanism of hyperglycemia in severe COVID-19, even after accounting for glucocorticoid use. While we did observe beta cell failure in a minority of patients, many of them had pre-existing advanced diabetes marked by the use of 2 or more anti-hyperglycemic medications or insulin, or an elevated hemoglobin A1c (HbA1c), signifying poor blood glucose control. Perhaps classifying the mechanism of hyperglycemia as IR or BCF in an individual infected with SARS-CoV-2 by insulin, C-peptide, and glucose measurements may guide glucose lowering therapy. Insulin is the accepted treatment for hyperglycemia in hospitalized patients. Our study raises the question for hyperglycemic patients with severe COVID-19 who display an IR phenotype, whether adding an insulin sensitizing medication, such as a thiazolidinedione or metformin, may increase glucose metabolism and spare insulin usage. This would need to be tested for safety and efficacy before clinical recommendations are made. Conversely, hyperglycemic COVID-19 patients with BCF would be treated with insulin as the mainstay, avoiding insulin sensitizing agents due to the potential for adverse effects.

Our data suggest that adipose dysfunction is a feature of COVID-19 that may drive hyperglycemia. Adiponectin and adiponectin/leptin ratios are markedly reduced in COVID-19 patients. Hamsters infected with SARS-CoV-2 show presence of SARS-CoV-2 viral RNA in the adipose tissue along with substantial decreases in *Adipoq*. Collectively our results implicate direct viral infection of adipose tissues as one potential mechanism for adipose tissue dysfunction and insulin resistance. It remains to be determined which cell(s) within the adipose tissue are infected. Diagnosing adipose dysfunction in COVID-19 by assessing circulating adipokine levels has the potential to be clinically actionable in the future since medications such as thiazolidinediones improve adipose function and insulin sensitivity.

In this study, we classified hyperglycemic patients as predominantly IR or BCF based on plasma samples collected early, generally during the first 2 days of ICU admission. IR and BCF are not mutually exclusive and hyperglycemia can be dynamic, especially in acutely ill patients who may be experiencing septic shock, evolving through different phases of acute viral infection, and being treated with glucocorticoids. Our data in critically ill patients precludes extensive physiological assays of insulin secretion and insulin sensitivity. Future studies with longitudinal assessment of IR and BCF over the course of acute SARS-CoV-2 infection may help to better understand the dynamic nature of hyperglycemia. This study is not powered to detect rare events nor does it rule out potential SARS-CoV-2 infection of pancreatic islet cells but suggests that it is not a major etiology of hyperglycemia in the majority of COVID-19 patients. Viral infection of beta cells may be subclinical or uncommon. Notwithstanding, follow up studies on COVID survivors and those with “Long COVID” are needed to monitor for future IR and BCF. It remains to be determined if patients who recovered from hyperglycemia prior to discharge will have an increased future risk of developing diabetes.

## Materials and Methods

### Study design and cohort description

Patients were classified into three groups, COVID positive and ARDS positive (C+A+), COVID negative and ARDS positive (C-A+), COVID negative and ARDS negative (C-A-) for analysis. The COVID-19 positive component of the study cohort was sourced from multiple institutional resources designed to facilitate COVID research. These resources are aggregated and integrated into a database called the COVID Institutional Data Repository (COVID-IDR). The COVID-IDR contains data extracted automatically from electronic health record (EHR) systems, as well as data abstracted manually, using REDCap^41^, an established research data collection tool, for patients testing positive for SARS-CoV-2 via RT-PCR who were admitted or seen in the emergency department at three hospitals within the NewYork-Presbyterian (NYP) healthcare system between March 3, 2020 and May 15, 2020. Some variables (including comorbidities and outcomes) were derived from the REDCap project via manual abstraction, while others (e.g. in-hospital medication usage) were derived from automatically extracted EHR data.

The control population was drawn from an existing data base of patients with an ICU admission from 2014-2018 at NYP-WCMC, the Weill Cornell-Critical Care Database for Advanced Research (WC-CEDAR), which aggregates data from institutional EHR’s for all patients managed in intensive care units at NYP-WCMC and NYP-LMH^42^.

For both cases and controls we extracted demographics (age, sex), in-hospital laboratory tests (glucose, HbA1c, LDH, creatinine and WBC), medication use (glucocorticoids, insulin, dextrose) and comorbidity information (history of diabetes, hypertension, coronary artery disease, obesity [BMI ≥30], kidney disease, liver disease, chronic obstructive pulmonary disease [COPD], and cancer). Acute conditions, such as ARDS, acute MI, septic shock and vasopressor requirements were also available.

For COVID-19 patients, data pertaining to BMI, diabetes status, and HbA1c were manually extracted by trained and quality controlled medical student reviewers. BMI was determined from height and weight recorded during hospital admission or from an outpatient encounter within the past 3 months. Diabetes status was defined by physician documentation of type 1 or type 2 DM in the medical history/diagnosis codes or by HbA1c ≥ 6.5%. HbA1c levels recorded within 10 days of hospitalization or within 90 days prior to admission were included. Diabetes medications were abstracted by manual review of the electronic health records. Hyperglycemia was defined as having a peak glucose value of >170 mg/dL. Peak glucose was defined as the highest glucose value during the entire hospitalization. Glucose measurements were extracted from the electronic health record. Patients without glucose values were excluded from the analysis. For study subjects with plasma profiled samples, two separate glucose values > 170 mg/dL were required to be classified as hyperglycemic. In addition, dextrose infusions were assessed to ensure that iatrogenic hyperglycemia was not being identified.

### Sample collection and ELISA

Since 2014 investigators have prospectively consented patients admitted to any ICU at NYP-WCMC to participate in a registry involving collection of biospecimens and clinical data (Weill Cornell Medicine Registry and Biobank of Critically Ill Patients). Additionally, during the COVID-19 pandemic, an electronic informed consent was obtained from all SARS-CoV-2 positive subjects or their surrogates for inclusion. For each participant, whole blood (6-10 mL) was obtained. Briefly, whole blood samples were drawn into EDTA-coated blood collection tubes (BD Pharmingen, San Jose, CA). Samples were stored at 4°C and centrifuged within 4 hours of collection. Plasma was separated and divided into aliquots and kept at −80°C. Plasma insulin and C-peptide were measured by ELISA and values determined by a standard curve according to manufacturer’s instructions (Mercodia, Uppsala, Sweden).

The registry was approved by the institutional review board of WCMC (1405015116, 20-05022072, 20-03021681).

### Multiplex Metabolic Protein Array

From the COVID and control populations, 101 patients (C+A+ n=59, C-A+ n=19, C-A-n=23) with plasma samples were selected and analysed using the MILLIPLEX MAP Human Adipokine and Human Metabolic Hormone Magnetic Bead Panels (EMD Millipore), according to manufacturer’s instructions. Undiluted plasma was used for the Metabolic Hormone Panel. For the Adipokine Panel, plasma samples were diluted 1:800. The targets of the Metabolic Hormone Panel were: C-peptide, Ghrelin, GIP, GLP-1 (active), Glucagon, IL-6, Insulin, Leptin, PP, PYY, Amylin (active). The targets of the Adipokine panel were: Adiponectin, Adipsin, Resistin, Lipocalin 2, PAI-1.

### Glycemic categorization of plasma-sampled patients

Patients classed as hyperglycemic were subdivided into IR or BCF. The C-peptide reading from the Multiplex Metabolic Protein Array (pg/mL) was divided by the glucose measurement closest to the time of plasma collection (mg/dL) to calculate C-peptide/glucose ratios. Patients who had a ratio of less than 20.5 and were either hyperglycemic or receiving insulin treatment at sampling time were classed as BCF. Hyperglycemic patients with a C-peptide/glucose ratio > 20.5 were classed as IR. The cutoff ratio of 20.5 was determined empirically, based on clinical review of patients showing consistency with BCF phenotype, such as moderate to severe hyperglycemia and extensive insulin dosing, confirming endogenous insulin deficiency. Additionally, a ratio of 20.2 has been previously associated with poor beta cell reserve and future need for insulin therapy^29,30^. The subcategorization was confirmed using C-peptide values obtained by ELISA.

### Data/Statistical Analysis

The COVID and control populations were profiled and tabulated. The subsample of patients with the multiplex arrays were profiled and tabulated and statistical tests were conducted to determine group differences (C+A+, C-A+, C-A-). Models to determine if group membership predicted the outcome of euglycemia, insulin resistance, or beta cell failure were determined from the assays. The outcome was assumed to be ordinal; ranks of 0, 1, and 2 were assigned to euglycemic, insulin resistant, and beta cell failure patients respectively. As such, ordinal logistic regression models with a cumulative logit link function was employed. Models were tested to meet the assumption of proportional odds. When the assumption was violated, we used the unequal slopes option where the model assumes separate slopes for each level of the outcome.

### Propagation and titration of SARS-CoV-2

SARS-CoV-2 isolate USA-WA1/2020 (NR-52281) was provided by the Center for Disease Control and Prevention and obtained through BEI Resources, NIAID, NIH. SARS-CoV-2 was propagated in Vero E6 cells in DMEM supplemented with 2% FBS, 4.5 g/L D-glucose, 4 mM L-glutamine, 10 mM Non-Essential Amino Acids, 1 mM Sodium Pyruvate and 10 mM HEPES using a passage-2 stock of virus. Three days after infection, supernatant containing propagated virus was filtered through an Amicon Ultra 15 (100 kDa) centrifugal filter (Millipore Sigma) at ∼4000 rpm for 20 minutes. Flow through was discarded and virus was resuspended in DMEM supplemented as described above. Infectious titers of SARS-CoV-2 were determined by plaque assay in Vero E6 cells in Minimum Essential Media supplemented with 2% FBS, 4 mM L-glutamine, 0.2% BSA, 10 mM HEPES and 0.12% NaHCO3 and 0.7% agar. All work involving live SARS-CoV-2 was performed in the CDC/USDA-approved BSL-3 facility of the Icahn School of Medicine at Mount Sinai in accordance with institutional biosafety requirements.

### SARS-CoV-2 infections of Hamsters

3-5-week-old male Golden Syrian hamsters (Mesocricetus auratus) were obtained from Jackson Laboratories. Hamsters were acclimated to the CDC/USDA-approved BSL-3 facility of the Global Health and Emerging Pathogens Institute at the Icahn School of Medicine at Mount Sinai for 2-4 days. Before intranasal infection, hamsters were anesthetized by intraperitoneal injection with a ketamine HCl/xylazine solution (4:1). Hamsters were intranasally inoculated with 100 pfu of SARS-CoV-2 in PBS (or PBS only as a control) in a total volume of 100 μl. Two days post-infection hamsters were euthanized and fat tissue depots were collected^43^. All animal experiments were performed on at least two separate occasions. All animal experiment procedures, breeding, and ethical use were performed in accordance with the guidelines set by the Institutional Animal Care and Use Committee at Mount Sinai School of Medicine.

### RNA extraction and qPCR analysis

RNA isolation from hamster fat samples was performed using the RNeasy Mini kit (Qiagen) as per manufacturer’s protocol. cDNA was synthetized through reverse transcription using a cDNA synthesis kit (Thermo). cDNA was analyzed by real-time PCR using specific gene primers and a SYBR Green Master Mix (Quanta). To quantify viral replication, measured by the expression of sgRNA transcription of the viral N gene, one-step quantitative real-time PCR was performed using SuperScript III Platinum SYBR Green One-Step qRT-PCR Kit (Invitrogen) with primers specific for the TRS-L and TRS-B sites for the N gene as well as ACTB as an internal reference. Quantitative real-time PCR reactions were performed on an Biorad Real-Time PCR Instrument (ABI). Delta-delta-cycle threshold (ΔΔCT) was determined relative to ACTB levels and normalized to mock infected samples. Primer sequences were as follows

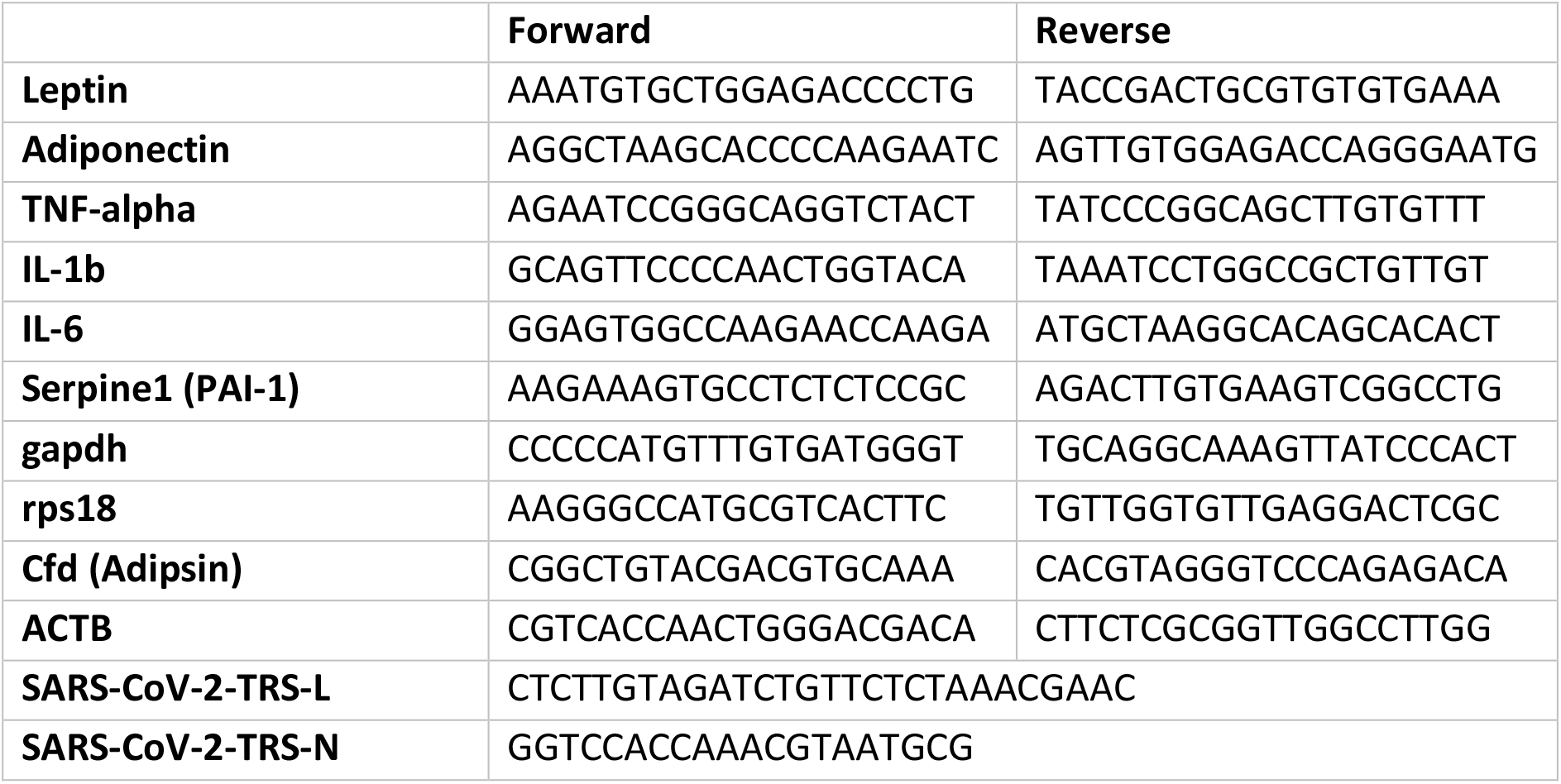

### Western blot

4-12% Bis-Tris Bolt gels (Thermo) were loaded with 20 μg of tissue protein extract from hamster SAT or VAT, or 1 μL of hamster serum. The protein was transferred to a PVDF membrane and probed with primary antibody for adiponectin (EMD Millipore, MAB3608, 1:1000) overnight at 4 °C, followed by an incubation with horseradish peroxidase-conjugated secondary antibody. Signal detection was carried out using SuperSignal West Pico detection reagent (Thermo). Ponceau stain was used as a loading control. Band density was quantified using ImageJ.

## Data Availability

All requests for raw and analyzed data will be reviewed to verify if the request is subject to any intellectual property or confidentiality obligations. Any data and materials that can be shared will be released via a Material Transfer Agreement.

## Acknowledgements

This work was supported by the following grants: NIH R01 DK121140 (J.C.L.) and R01 DK121844 (J.C.L.), NCI R01 CA234614 (R.E.S), NIAID 2R01 AI107301 (R.E.S), and NIDDK R01 DK121072 (R.E.S). R.E.S. is supported as Irma Hirschl Trust Research Award Scholars. In addition, this study received support from NewYork-Presbyterian Hospital (NYPH) and Weill Cornell Medical College (WCMC), including the Clinical and Translational Science Center (CTSC) (UL1 TR 0002384). The views expressed in this manuscript are those of the authors and do not necessarily represent the official views of the National Institute of Diabetes and Digestive and Kidney Diseases or the National Institutes of Health.

---

**Extended Data Fig.1.**
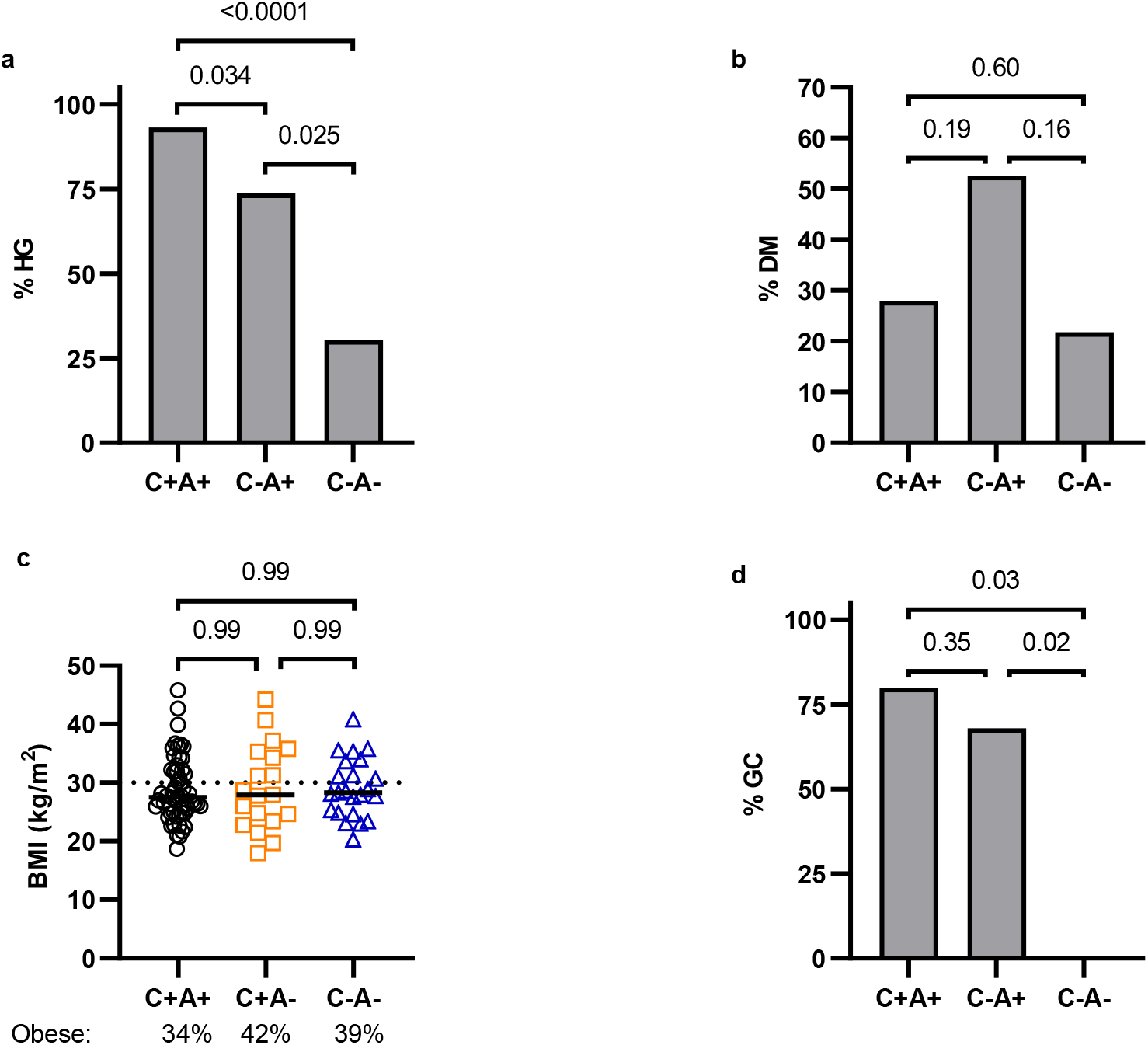
Characteristics of the plasma-sampled sub-cohorts. **a, b**, Percentage of hyperglycemic/diabetic patients among plasma sampled COVID-19 ARDS+ (C+A+, n=59) patients and ICU controls with (C-A+, n=19) or without ARDS (C-A-, n=23). Data were analyzed by two-sided Fisher’s exact test with Bonferroni-Holm’s correction for multiple comparisons. **c**, BMI of COVID-19 ARDS+ (C+A+, n=59) patients and ICU controls with (C-A+, n=19) or without ARDS (C-A-, n=23). Data were analyzed by Kruskal-Wallis test with Dunn’s test corrected for multiple comparisons. **d**, Percentage of patients who received glucocorticoid (GC) treatment among plasma sampled COVID-19 ARDS+ (C+A+, n=59) patients and ICU controls with (C-A+, n=19) or without ARDS (C-A-, n=23). Data were analyzed by two-sided Fisher’s Exact Test with Bonferroni-Holm’s correction for multiple comparisons.

**Extended Data Fig.2.**
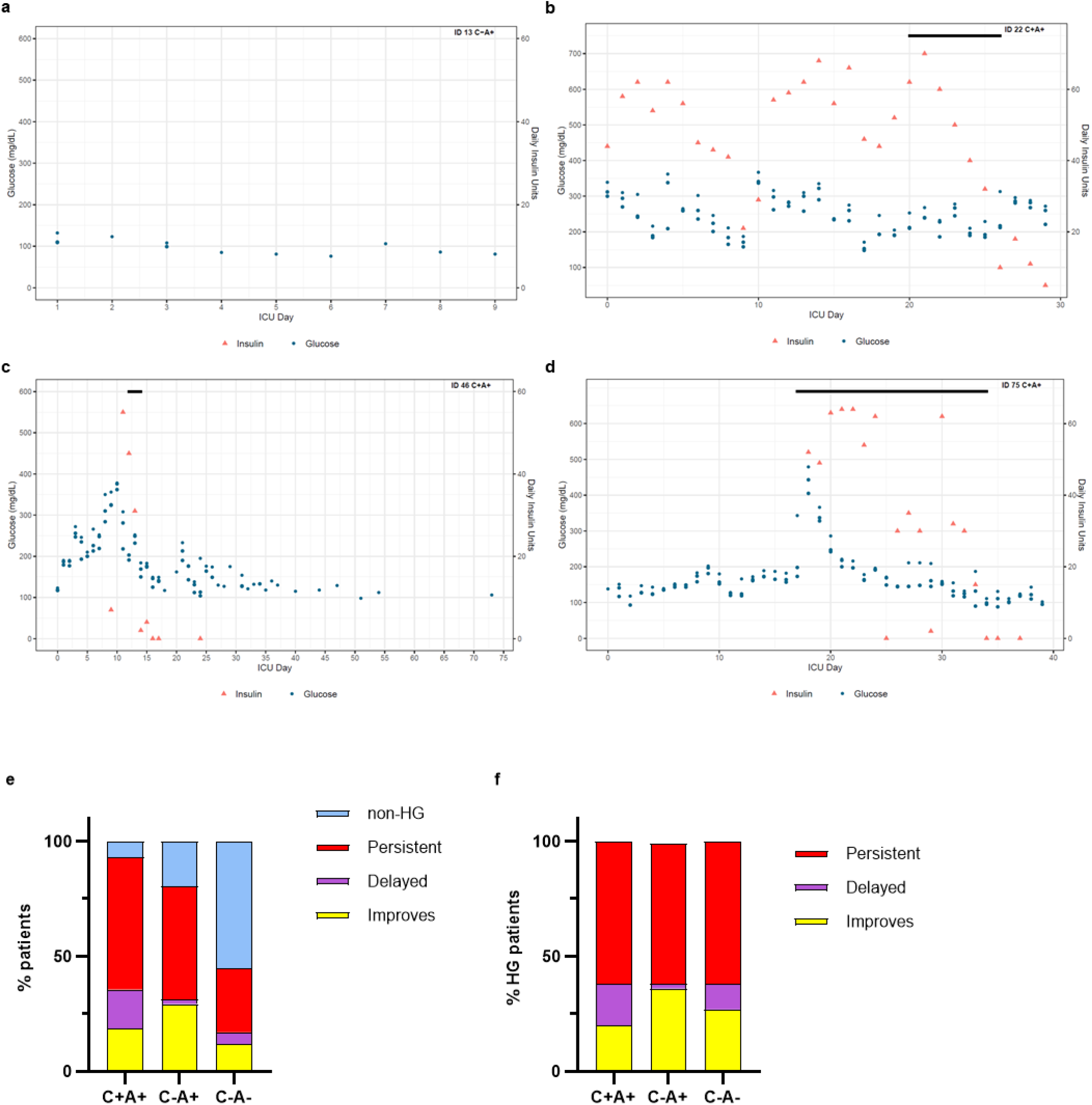
Example graphs showing the glucose values, insulin and glucocorticoid treatments of patients involved in the study. The 3 highest glucose readings for each day are plotted. The black bar indicates the days when glucocorticoids were administered. The graphs illustrate a non-hyperglycemic patient (a), a patient who remained persistently hyperglycemic (b), a patient who was initially hyperglycemic but improved over time (c), and a patient who was not hyperglycemic upon admission but developed HG over time which was further exacerbated by glucocorticoid treatment (d). e, Change in hyperglycemia over time: Percentage of patients who are never HG, HG throughout their hospital stay (“persistent”), HG developing after at least one week without HG (“delayed”), HG initially but normal for at least a week before discharge (“improves”) (C+A+, n=59, C-A+ n=51, C-A-n=201 patients) f, Data from d considering only hyperglycemic patients.

**Extended Data Fig.3.**
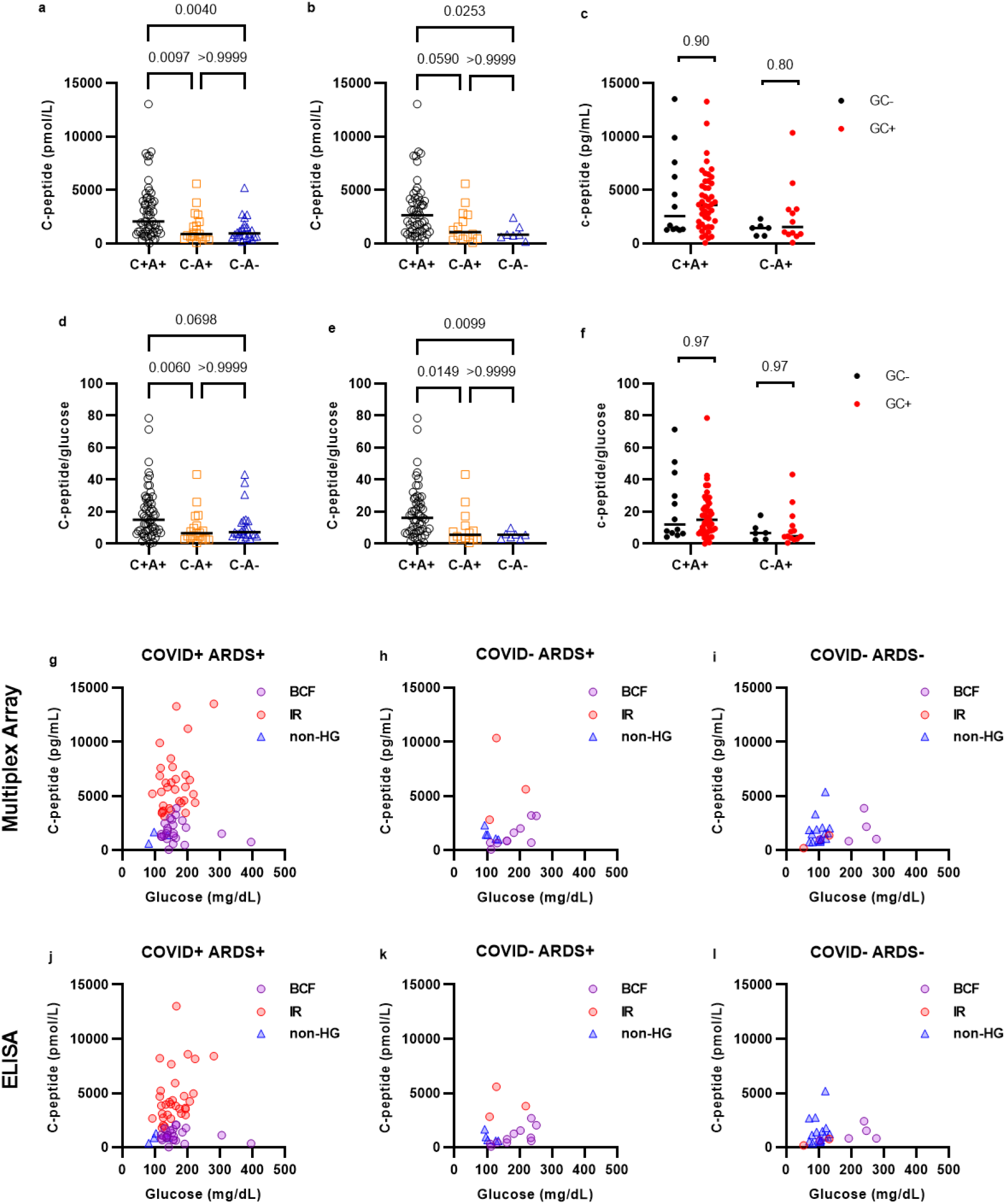
Distribution of C-peptide and glucose readings. **a, b**, C-peptide concentration in the plasma of COVID-19 ARDS patients (C+A+ n=59), ARDS+ ICU controls (C-A+ n=19), and ARDS-ICU controls (C-A-n=23), measured by ELISA. Data were analyzed using Kruskal-Wallis and Dunn’s tests with multiple comparison correction. **b**, C-peptide concentration in the plasma of hyperglycemic patients and controls (C+A+ n=55, C-A+ n=13, C-A-n=7), measured by ELISA. Data were analyzed using Kruskal-Wallis and Dunn’s tests with multiple comparison correction. **c**, C-peptide concentration in the plasma of C+A+ patients (n=59) and C-A+ controls (n=19), subdivided according to whether they received glucocortidoids (GC) during their hospital stay, measured by Multiplex Protein Array. Data were analyzed using Mann-Whitney’s tests with Holm-Šídák’s multiple comparison correction. **d, e**, C-peptide/glucose ratio of all (**c**) or hyperglycemic only (**d**) Covid-19 ARDS and control patients, C-peptide measured by ELISA. Data were analyzed using Kruskal-Wallis and Dunn’s tests with multiple comparison correction. **f**, C-peptide/glucose ratio of COVID-19 ARDS patients (n=59) and ARDS+ controls (n=19), subdivided according to whether they received glucocortidoids (GC) during their hospital stay, C-peptide measured by Multiplex Protein Array. Data were analyzed using Mann-Whitney’s tests with Holm-Šídák’s multiple comparison correction. **g-l**, Glucose at plasma collection time was plotted against C-peptide concentration in the plasma, measured by Multiplex Protein Array (**g-i**) or ELISA (**j-l**), for COVID-19 ARDS patients (**g** and **j**, n=59), ARDS+ controls (**h** and **i**, n=19), and ARDS-controls (**j** and **l**, n=23). Each data point is colored according to their glycemic subgroup: non-hyperglycemic (non-HG), insulin resistant (IR), beta cell failure (BCF).

**Extended Data Fig.4.**
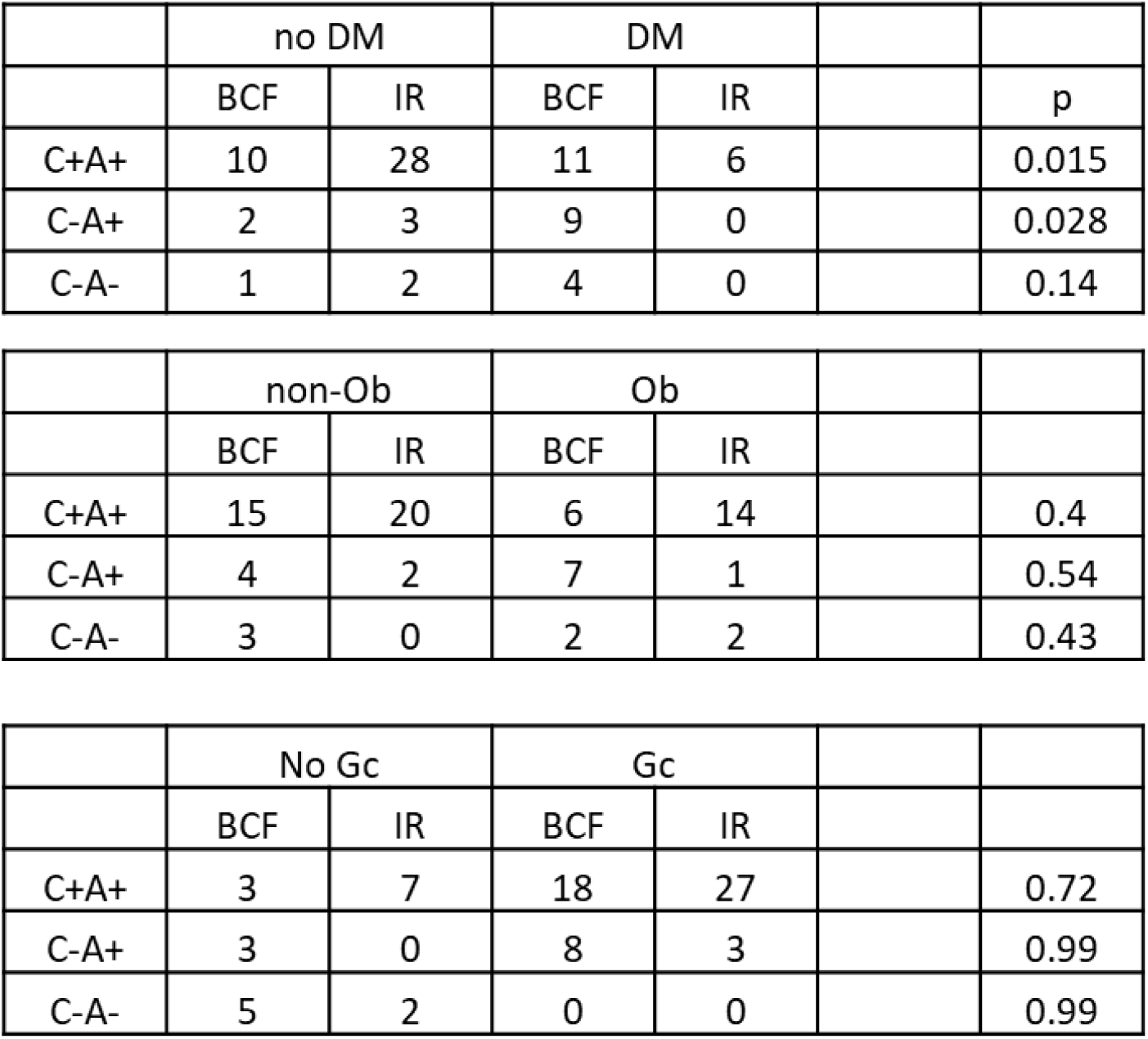
Association between insulin resistance (IR)/beta cell failure (BCF) and diabetes (DM), obesity (Ob), and glucocorticoid treatment (Gc). Association was tested using Fisher’s exact test for individuals classed as either IR or BCF among COVID-19 ARDS patients (C+A+, n=55), ARDS+ ICU controls (C-A+, n=14), and ARDS-ICU controls (C-A-, n=7).

**Extended Data Fig.5.**
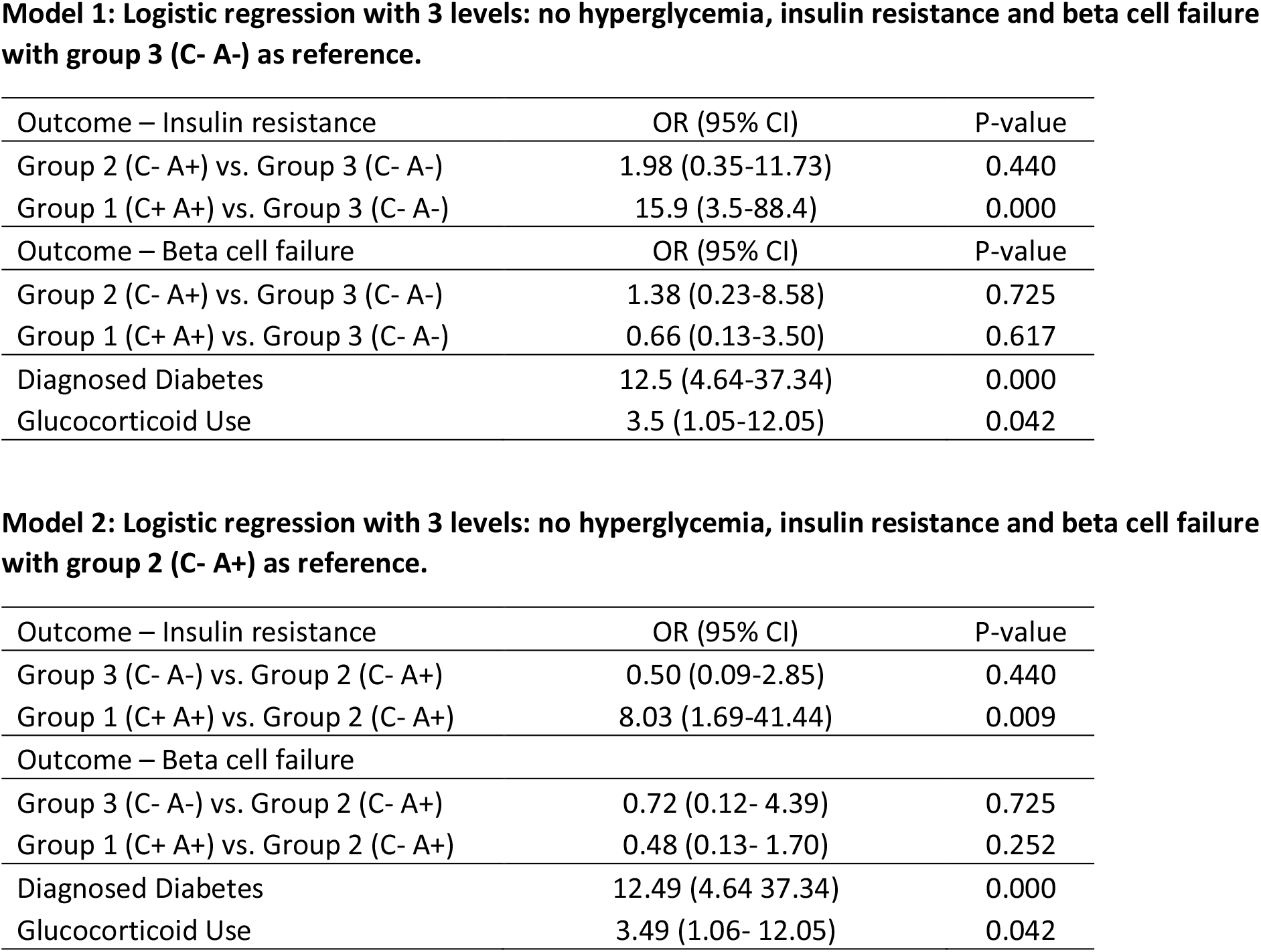
Logistic regression adjusting for diagnosed diabetes and glucocorticoid use. Models with a three level ordinal outcome (0 – no hyperglycemia, 1=insulin resistance and 2=beta cell failure). We fitted a partial proportional odds model assuming given nonproportionality only in the group variable and proportionality of odds for all the additional covariates such as Diagnosed Diabetes and Glucocorticoid use.

**Extended Data Fig.6.**
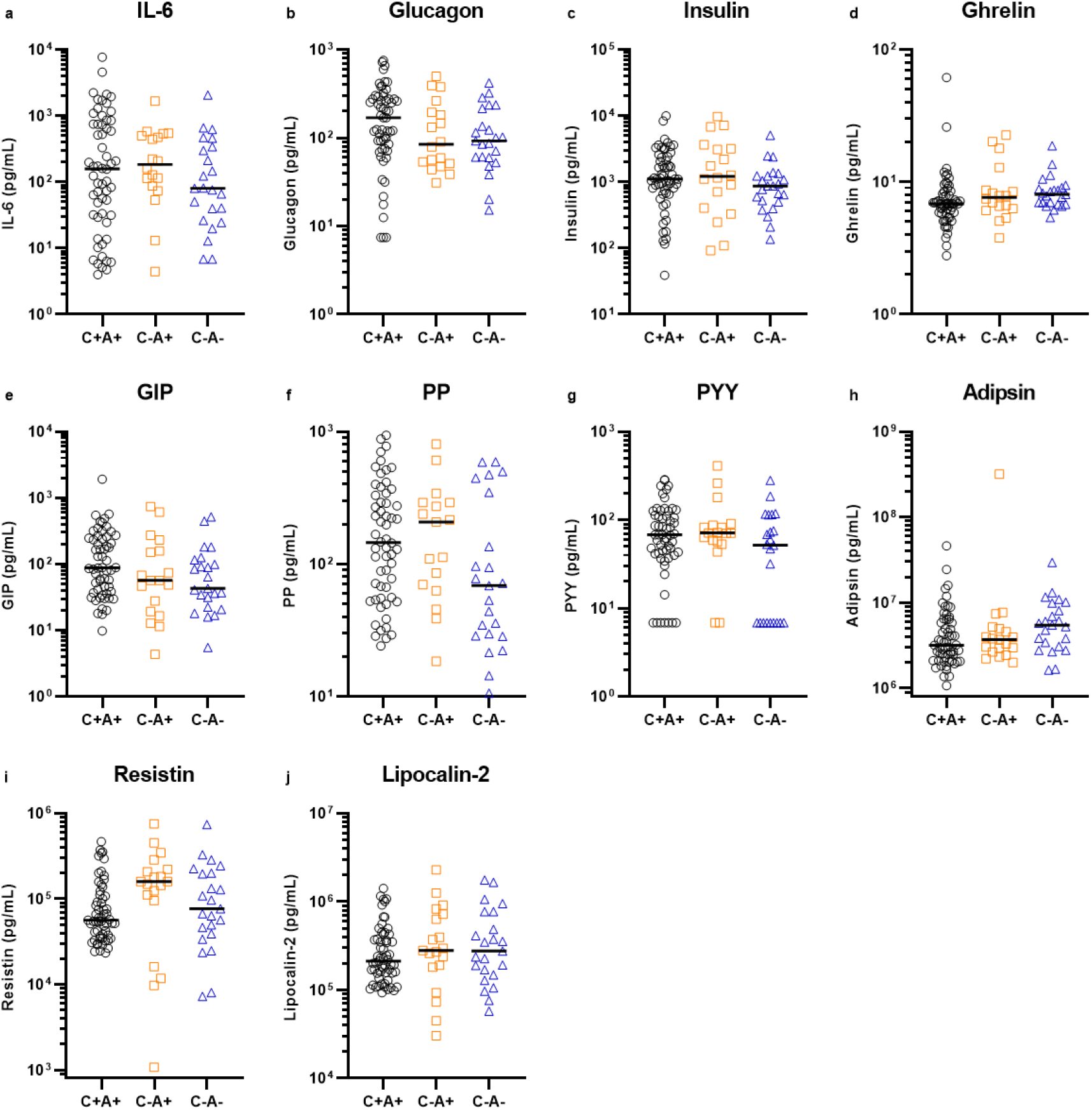
Glucoregulatory hormone profile in COVID-19. Plasma from COVID-19 ARDS (C+A+, n=59) and ICU control patients with ARDS (C-A+, n=19) and without ARDS (C-A-n=23) was analysed by multiplex metabolic protein array, targeting hormones known to modulate blood glucose homeostasis. The data were analyzed using Kruskal-Wallis tests. All targets with p>0.05 are shown here.

**Extended Data Fig.7.**
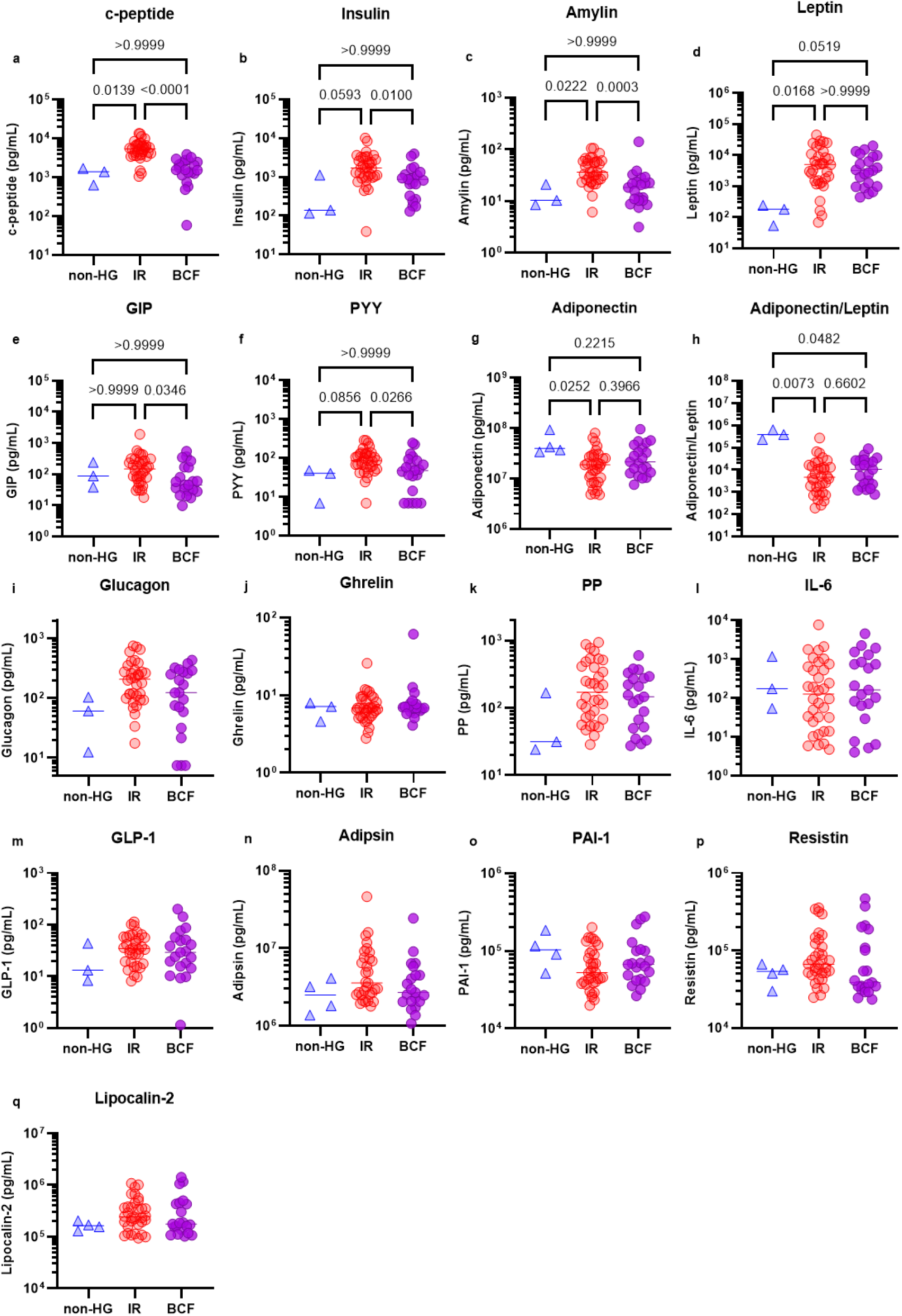
Metabolic hormone analysis of COVID-19 patients split by glycemic subgroup. Metabolic hormone readings of COVID-19 patients were subdivided according to the glycemic subcategory assigned to each patient. The data were analyzed using Kruskal-Wallis tests. All targets with p<0.05 (**a-h**) were subjected to Dunn’s tests with multiple comparison correction.

**Extended Data Fig.8.**
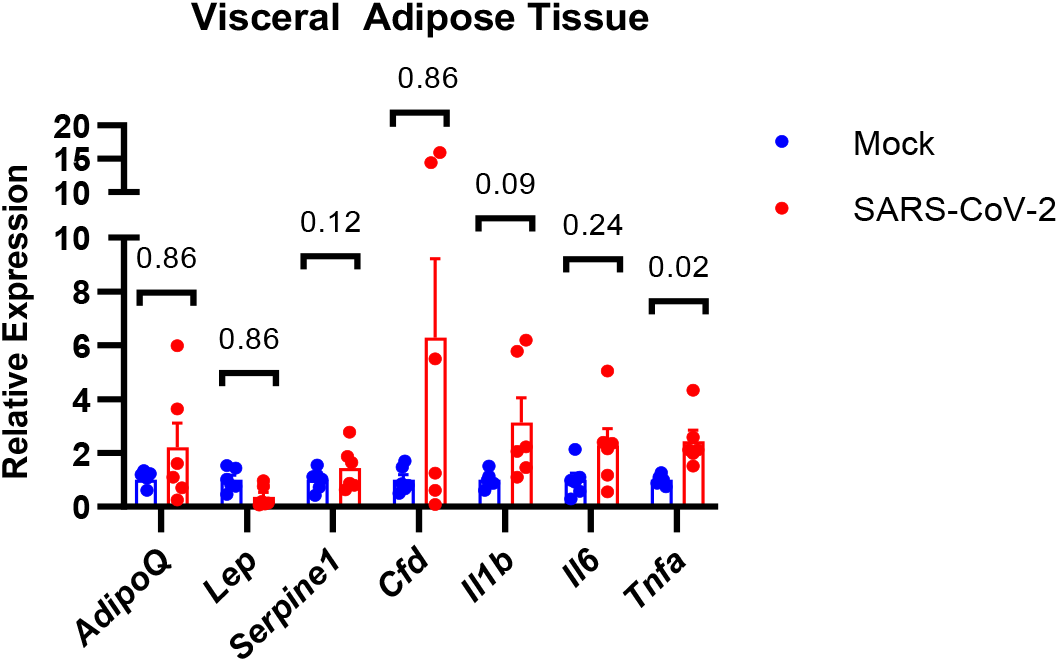
Altered gene expression in hamster visceral fat tissue following infection with Sars-CoV-2. RT-qPCR of mRNA from perigonadal fat tissue of Syrian hamsters infected with Sars-CoV-2 or mock virus. Data were analyzed using Mann-Whitney tests with Holm-Sidak’s multiple comparisons correction (n=6 hamsters per group), error bars show SEM.

**Extended Data Table 1:**
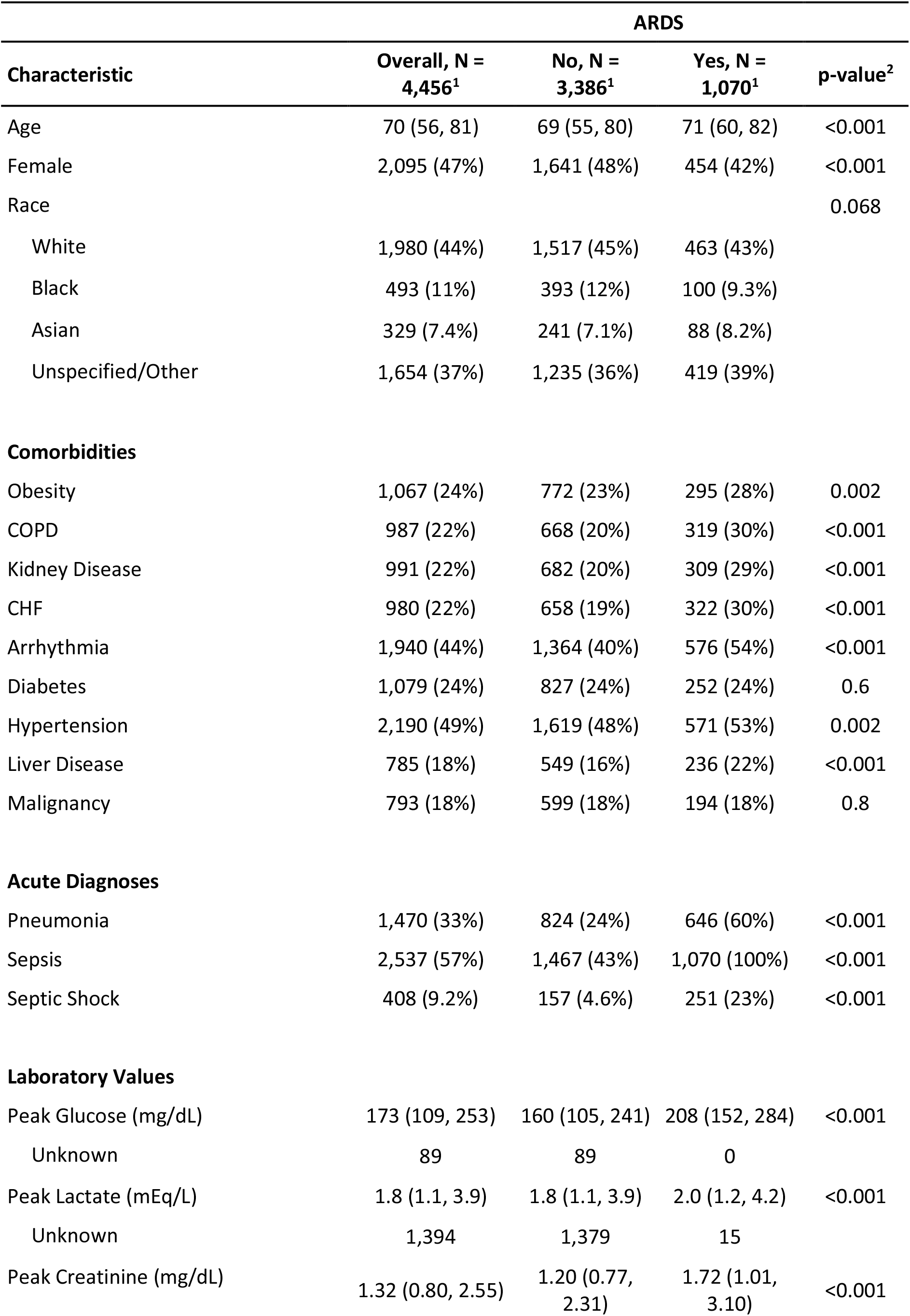
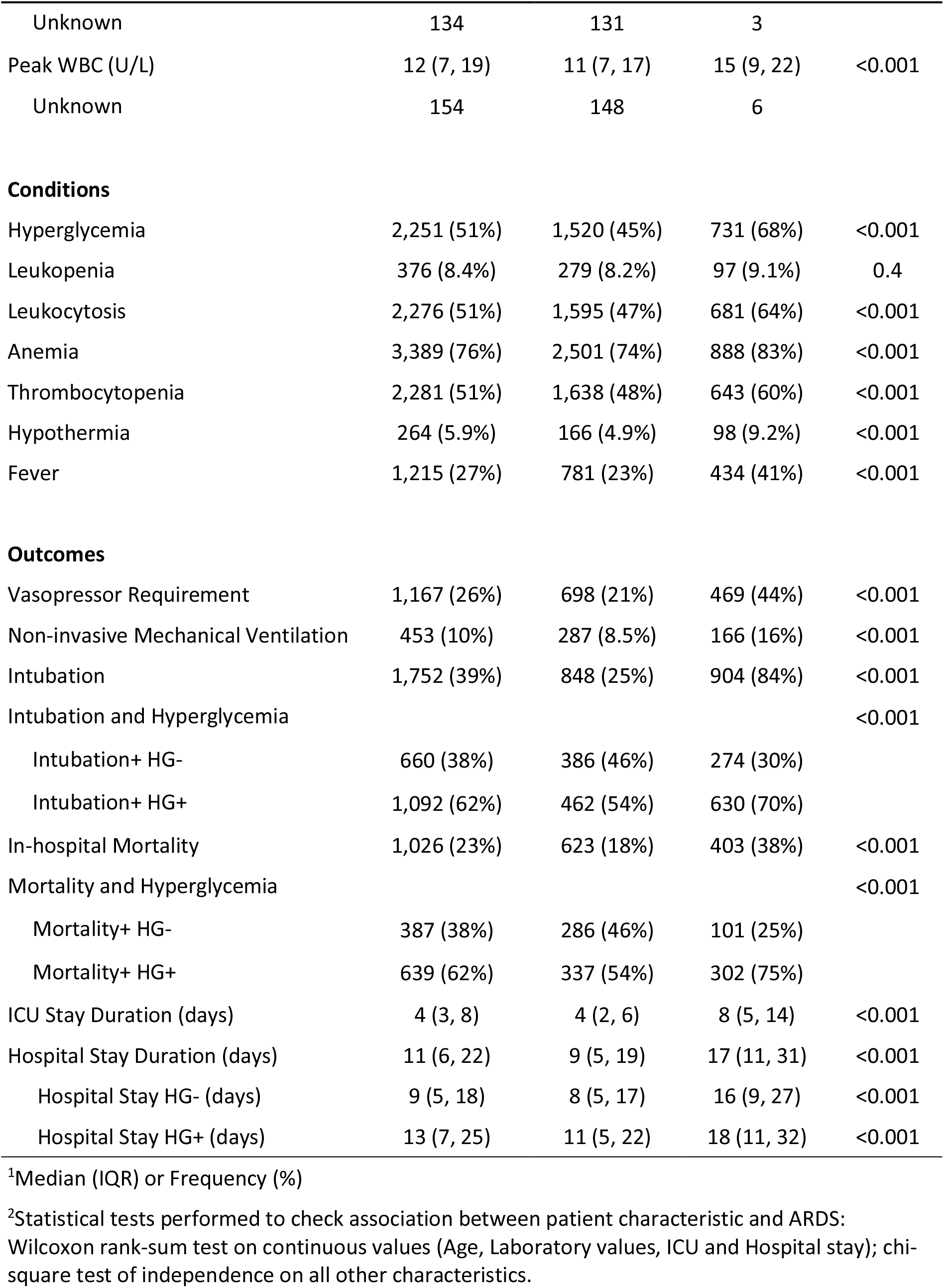
Characteristics of adult MICU patients, 2014-2018.

**Extended Data Table 2:** Demographics and clinical characteristics of plasma-sampled subset by COVID and ARDS status, COVID-19 patients with ARDS (C+A+), control patients with ARDS (C-A+), control patients without ARDS (C-A-), n=101.

| Characteristic | Overall, N = 101<sup>1</sup> | C-A-, N = 23<sup>1</sup> | C-A+, N = 19<sup>1</sup> | C+A+, N = 59<sup>1</sup> | p-value<sup>2</sup> |
| --- | --- | --- | --- | --- | --- |
| Age | 59 (50, 71) | 69 (50, 79) | 59 (52, 66) | 58 (49, 68) |  |
| Unknown | 2 | 2 | 0 | 0 |  |
| Female | 28 (27.7%) | 10 (43.5%) | 5 (26.3%) | 13 (22.0%) |  |
| Race |  |  |  |  |  |
| White | 35 (34.7%) | 9 (39.1%) | 8 (42.1%) | 18 (30.5%) |  |
| Black | 13 (12.9%) | 4 (17.4%) | 2 (10.5%) | 7 (11.9%) |  |
| Asian | 6 (5.9%) | 0 (0.0%) | 0 (0.0%) | 6 (10.2%) |  |
| Unspecified/Other | 47 (46.5%) | 10 (43.5%) | 9 (47.4%) | 28 (47.5%) |  |
| **Comorbidities** |  |  |  |  |  |
| Obesity | 37 (36.6%) | 9 (39.1%) | 8 (42.1%) | 20 (33.9%) |  |
| COPD | 5 (5.0%) | 1 (4.3%) | 0 (0.0%) | 4 (6.8%) |  |
| Kidney Disease | 5 (5.0%) | 1 (4.3%) | 1 (5.3%) | 3 (5.1%) |  |
| Heart Disease | 11 (10.9%) | 5 (21.7%) | 1 (5.3%) | 5 (8.5%) |  |
| Diabetes | 32 (31.7%) | 5 (21.7%) | 10 (52.6%) | 17 (28.8%) |  |
| Hypertension | 47 (46.5%) | 11 (47.8%) | 10 (52.6%) | 26 (44.1%) |  |
| Liver Disease | 9 (8.9%) | 4 (17.4%) | 2 (10.5%) | 3 (5.1%) |  |
| Malignancy | 23 (22.8%) | 8 (34.8%) | 10 (52.6%) | 5 (8.5%) |  |
| **Acute Diagnoses** |  |  |  |  |  |
| Acute MI | 10 (9.9%) | 3 (13.0%) | 2 (10.5%) | 5 (8.5%) |  |
| Septic Shock | 41 (40.6%) | 7 (30.4%) | 13 (68.4%) | 21 (35.6%) |  |
| Vasopressor | 83 (82.2%) | 12 (52.2%) | 16 (84.2%) | 55 (93.2%) |  |
| **Drug Therapy** |  |  |  |  |  |
| Steroids | 60 (59.4%) | 0 (0.0%) | 13 (68.4%) | 47 (79.7%) |  |
| DM Outpt Meds | 18 (17.8%) | 4 (17.4%) | 6 (31.6%) | 8 (13.6%) |  |
| ICU Insulin | 66 (65.3%) | 7 (30.4%) | 10 (52.6%) | 49 (83.1%) |  |
| **Glycemia Category** |  |  |  |  |  |
| Beta Cell Failure | 37 (36.6%) | 5 (21.7%) | 11 (57.9%) | 21 (35.6%) |  |
| Insulin Resistance | 39 (38.6%) | 2 (8.7%) | 3 (15.8%) | 34 (57.6%) |  |
| No Hyperglycemia | 25 (24.8%) | 16 (69.6%) | 5 (26.3%) | 4 (6.8%) |  |
| **In-hospital mortality** |  |  |  |  |  |
| Overall | 30 (29.7%) | 7 (30.4%) | 6 (31.6%) | 17 (28.8%) |  |
| Beta Cell Failure | 13 (43.3%) | 2 (28.6%) | 5 (83.3%) | 6 (35.3%) |  |
| Insulin Resistance | 10 (33.3%) | 0 (0%) | 0 (0%) | 10 (58.8%) |  |
| No Hyperglycemia | 7 (23.3%) | 5 (71.4%) | 1 (16.6%) | 1 (5.9%) |  |
| **Laboratory Values** |  |  |  |  |  |
| C-peptide | 2,087 (1,103, 4,367) | 1,369 (909, 1,975) | 1,441 (858, 2,689) | 3,487 (1,552, 5,756) | <0.001 |
| Ghrelin | 8.7 (6.8, 13.7) | 8.1 (6.9, 9.0) | 8.4 (7.1, 13.7) | 10.3 (6.5, 13.7) | 0.3 |
| GIP | 79 (34, 192) | 43 (27, 107) | 56 (21, 155) | 88 (39, 245) | 0.10 |
| GLP-1 active | 27 (15, 43) | 21 (8, 34) | 24 (14, 30) | 33 (16, 56) | 0.024 |
| Glucagon | 119 (61, 262) | 92 (60, 176) | 84 (52, 192) | 169 (78, 276) | 0.12 |
| IL-6 | 127 (37, 541) | 81 (33, 321) | 184 (94, 495) | 158 (32, 748) | 0.5 |
| Insulin | 1,088 (622, 2,150) | 873 (513, 1,234) | 1,213 (483, 3,153) | 1,112 (700, 2,292) | 0.2 |
| Leptin | 2,386 (718, 7,790) | 1,921 (430, 4,443) | 732 (489, 2,339) | 3,226 (1,186, 8,931) | 0.015 |
| PP | 118 (52, 292) | 69 (29, 242) | 160 (65, 287) | 145 (59, 321) | 0.11 |
| PYY | 68 (25, 109) | 52 (14, 95) | 71 (57, 86) | 68 (40, 112) | 0.2 |
| Amylin active | 21 (11, 35) | 10 (6, 27) | 17 (14, 20) | 26 (18, 50) | 0.003 |
| Adiponectin | 27,620,432 (15,716,992, 46,898,666) | 42,544,215 (23,948,160, 65,120,377) | 48,762,329 (28,519,584, 75,307,999) | 19,876,250 (11,564,496, 33,947,040) | <0.001 |
| Lipocalin2 | 239,544 (155,865, 447,157) | 276,130 (158,870, 630,597) | 279,851 (185,878, 677,894) | 212,271 (155,368, 403,378) | 0.6 |
| Resistin | 67,348 (38,703, 160,153) | 76,852 (42,781, 198,669) | 160,149 (103,462, 214,909) | 56,684 (37,270, 109,979) | 0.073 |
| Adipsin | 3,648,674 (2,522,890, 6,361,170) | 5,461,347 (3,260,265, 8,978,402) | 3,699,044 (2,793,774, 4,767,822) | 3,176,383 (2,205,467, 6,101,886) | 0.065 |
| PAI-1 | 51,739<br>(36,695, 84,340) | 41,070<br>(31,776, 58,728) | 45,221 (20,483, 67,740) | 61,900<br>(41,035, 99,918) | 0.007 |
| Adiponectin/Leptin | 11,107 (2,217, 45,859) | 21,900<br>(5,197, 110,137) | 58,967 (4,027, 100,625) | 5,247<br>(1,547, 21,360) | 0.008 |

<sup>1</sup>Median (IQR) or N (%)
<sup>2</sup>Statistical tests performed: Kruskal-Wallis test

