## Supplementary Data File 1 for "Hyperglycemia in Acute COVID-19 is Characterized by Adipose Tissue Dysfunction and Insulin Resistance"

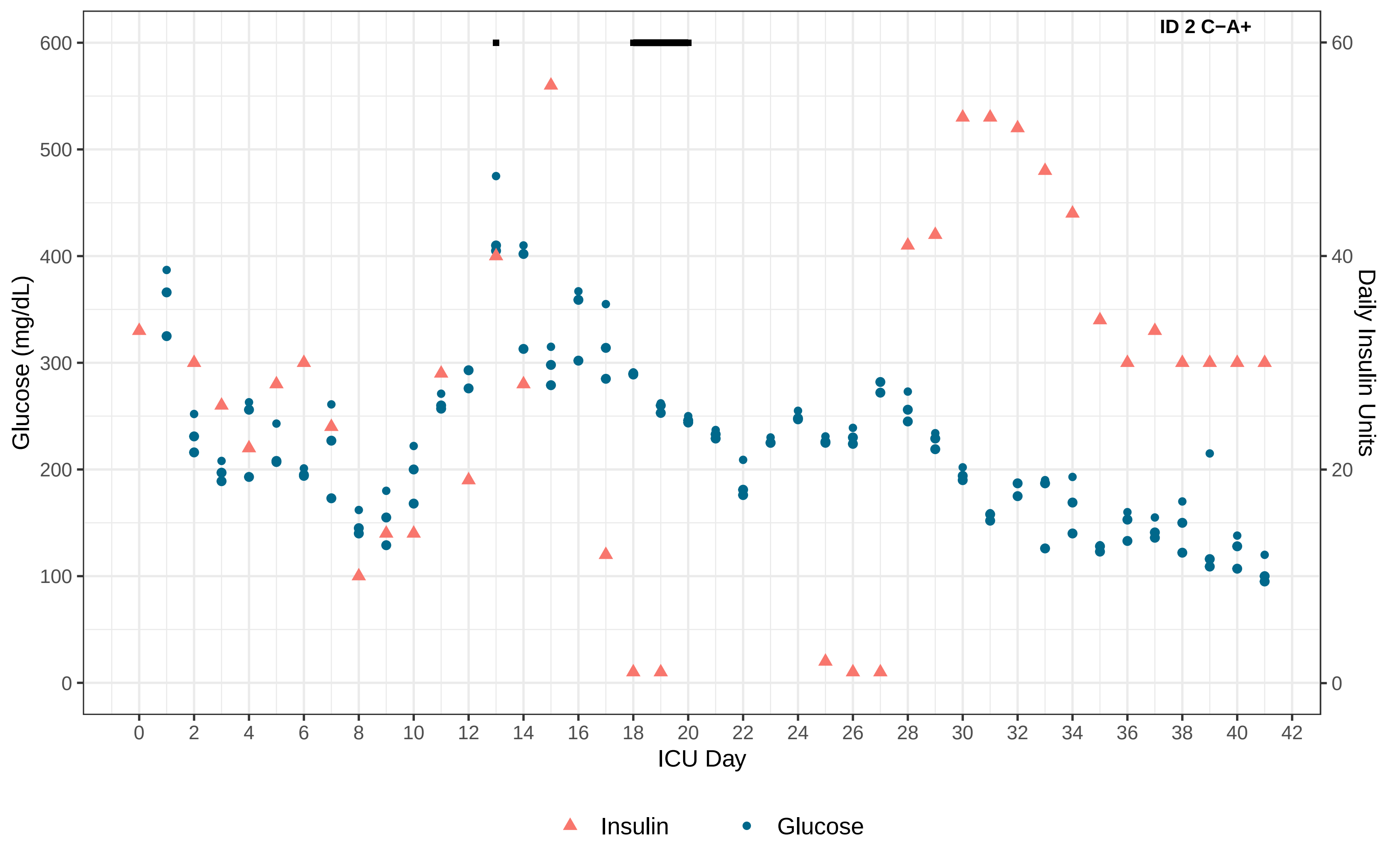

ID 2 C−A+
Glucose (mg/dL)
Daily Insulin Units
ICU Day
0
100
200
300
400
500
600
0
20
40
60
0 2 4 6 8 10 12 14 16 18 20 22 24 26 28 30 32 34 36 38 40 42
Insulin
Glucose

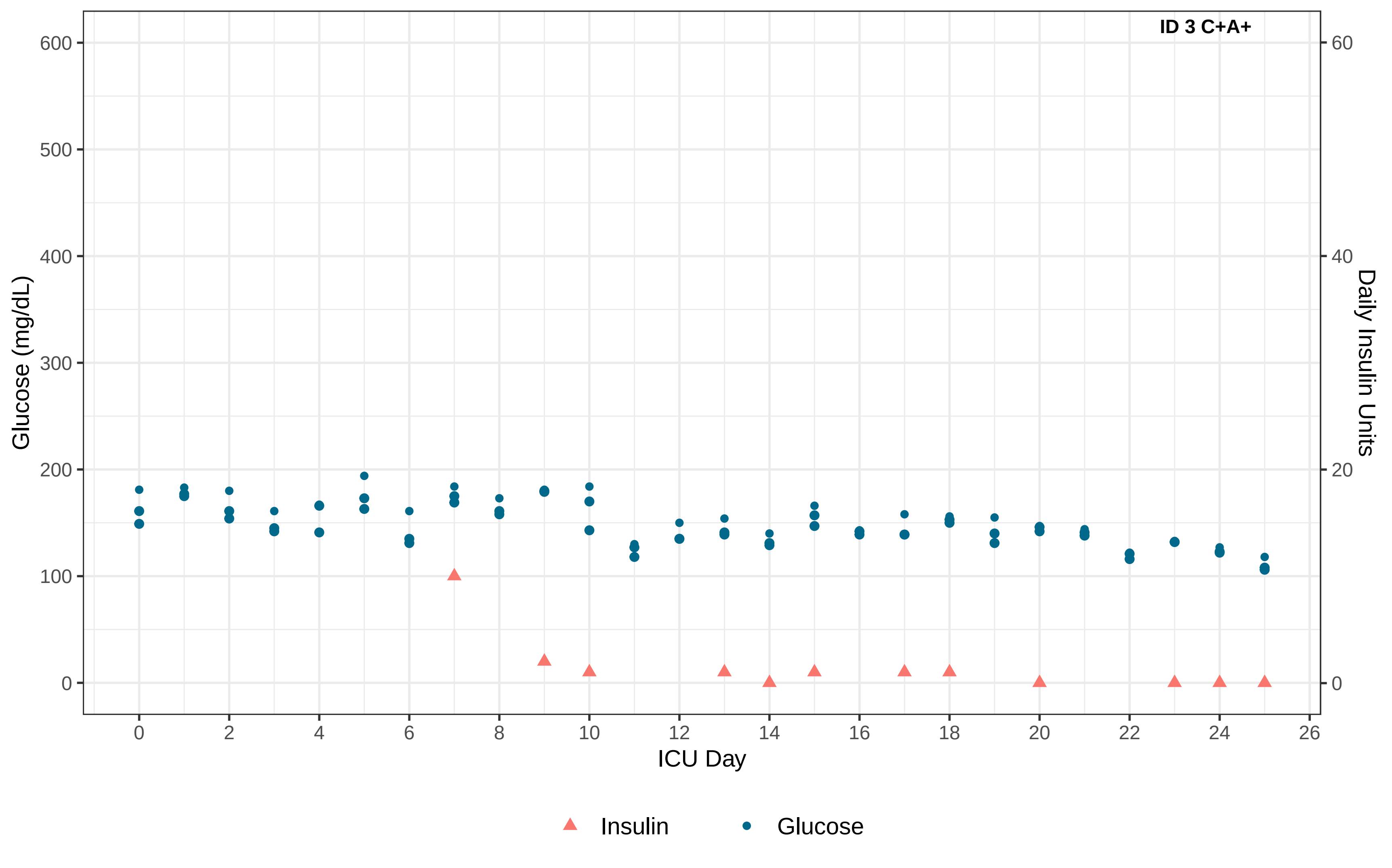

ID 3 C+A+
Glucose (mg/dL)
Daily Insulin Units
ICU Day
0
100
200
300
400
500
600
0
20
40
60
0
2
4
6
8
10
12
14
16
18
20
22
24
26
Insulin
Glucose

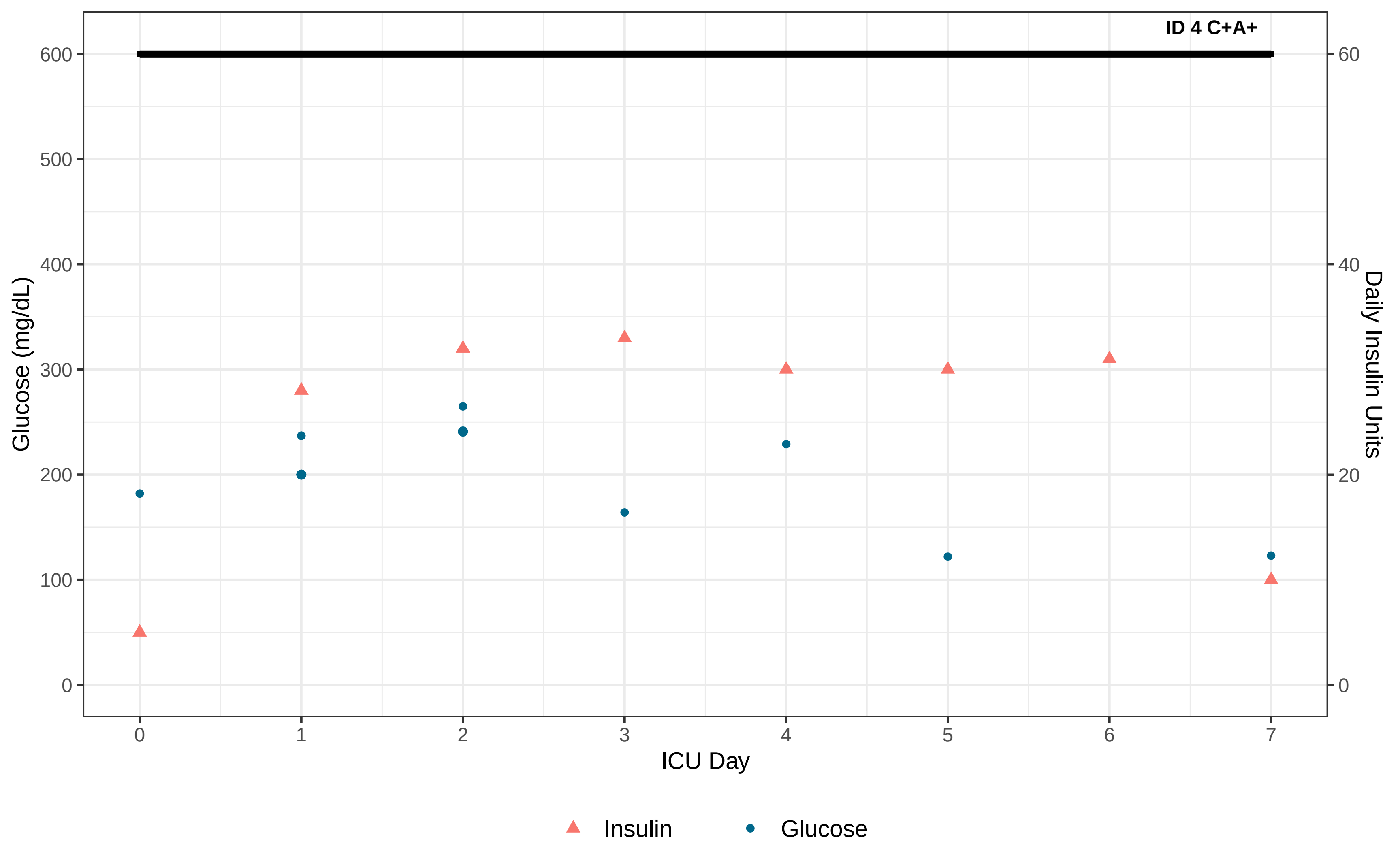
ID 4 C+A+
Glucose (mg/dL)
Daily Insulin Units
ICU Day
0
100
200
300
400
500
600
0
20
40
60
0
1
2
3
4
5
6
7
Insulin
Glucose

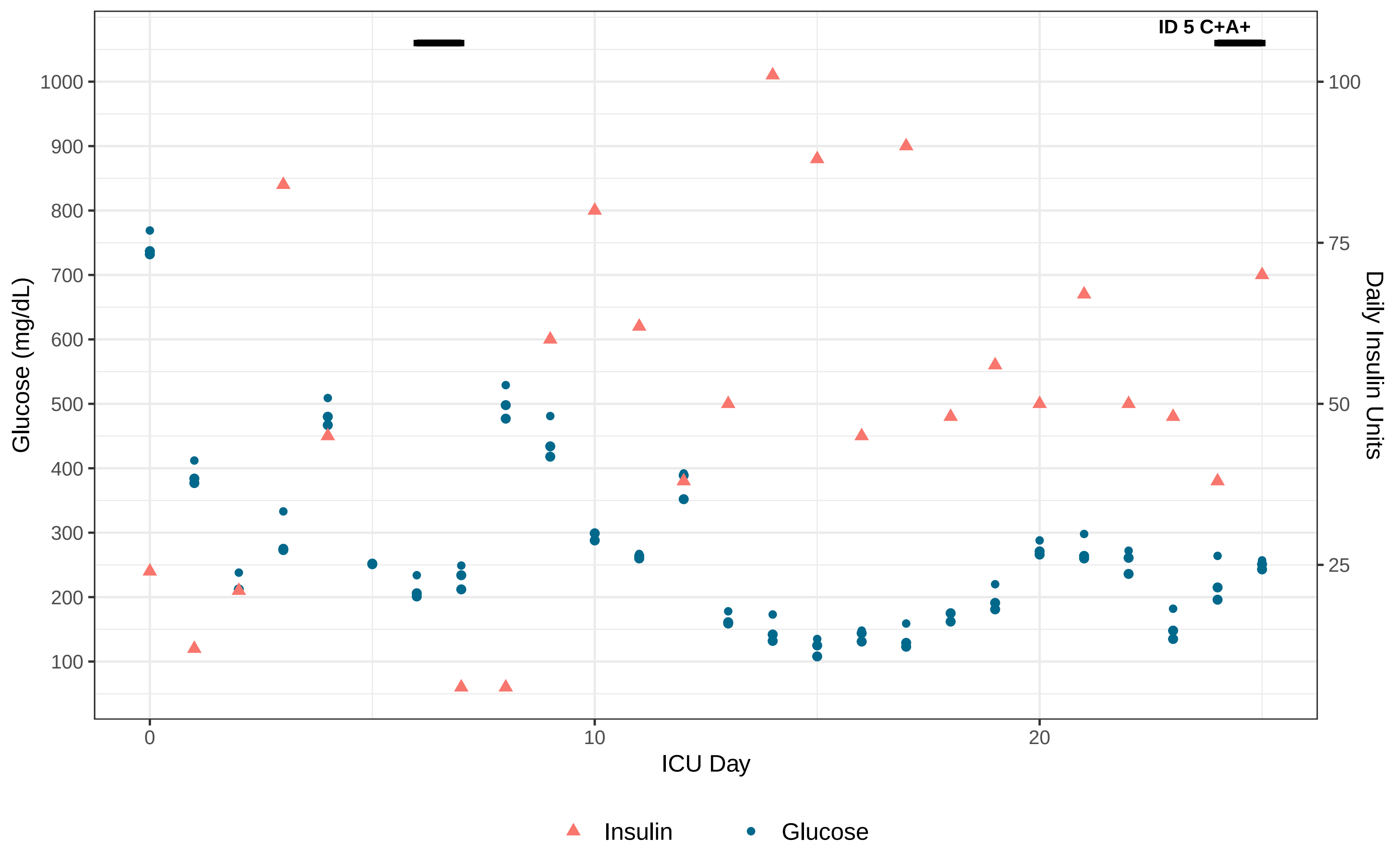

ID 5 C+A+
Glucose (mg/dL)
1000
900
800
700
600
500
400
300
200
100
Daily Insulin Units
100
75
50
25
ICU Day
0
10
20
Insulin
Glucose

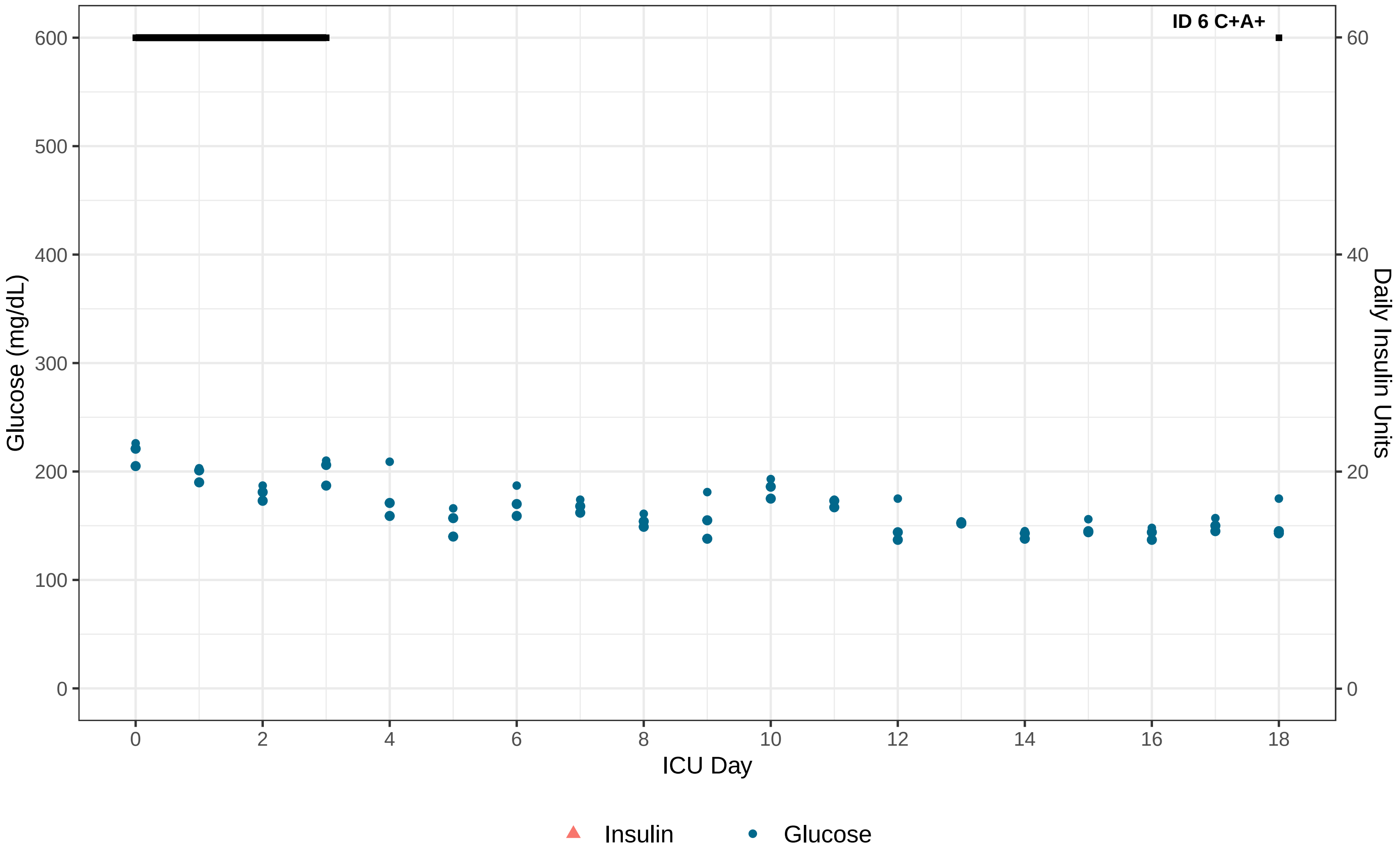
ID 6 C+A+
Glucose (mg/dL)
Daily Insulin Units
ICU Day
0
100
200
300
400
500
600
0
20
40
60
0
2
4
6
8
10
12
14
16
18
Insulin
Glucose

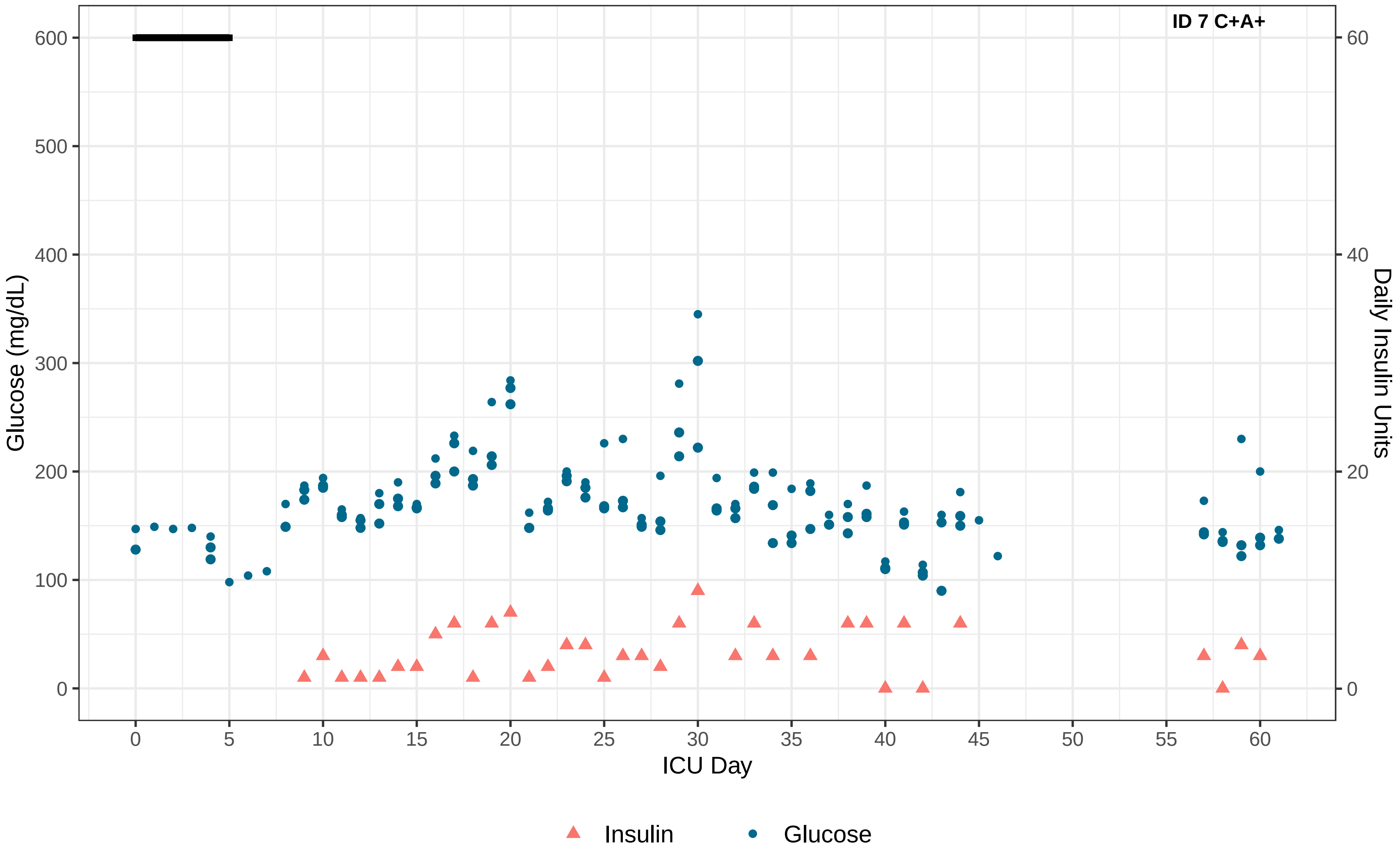

ID 7 C+A+
Glucose (mg/dL)
Daily Insulin Units
ICU Day
0
100
200
300
400
500
600
0
20
40
60
0
5
10
15
20
25
30
35
40
45
50
55
60
Insulin
Glucose

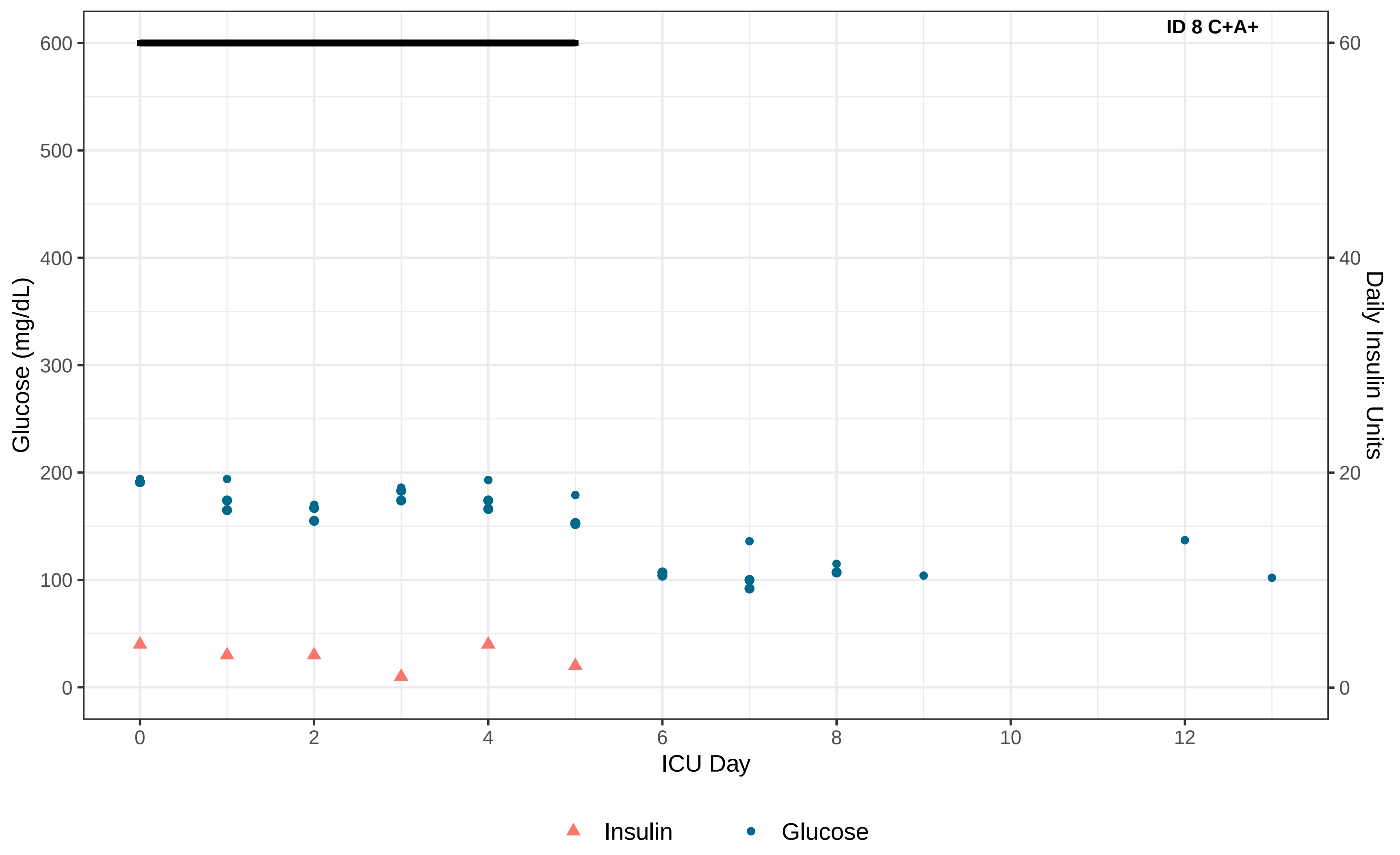
ID 8 C+A+
Glucose (mg/dL)
Daily Insulin Units
ICU Day
0
100
200
300
400
500
600
0
20
40
60
0
2
4
6
8
10
12
Insulin
Glucose

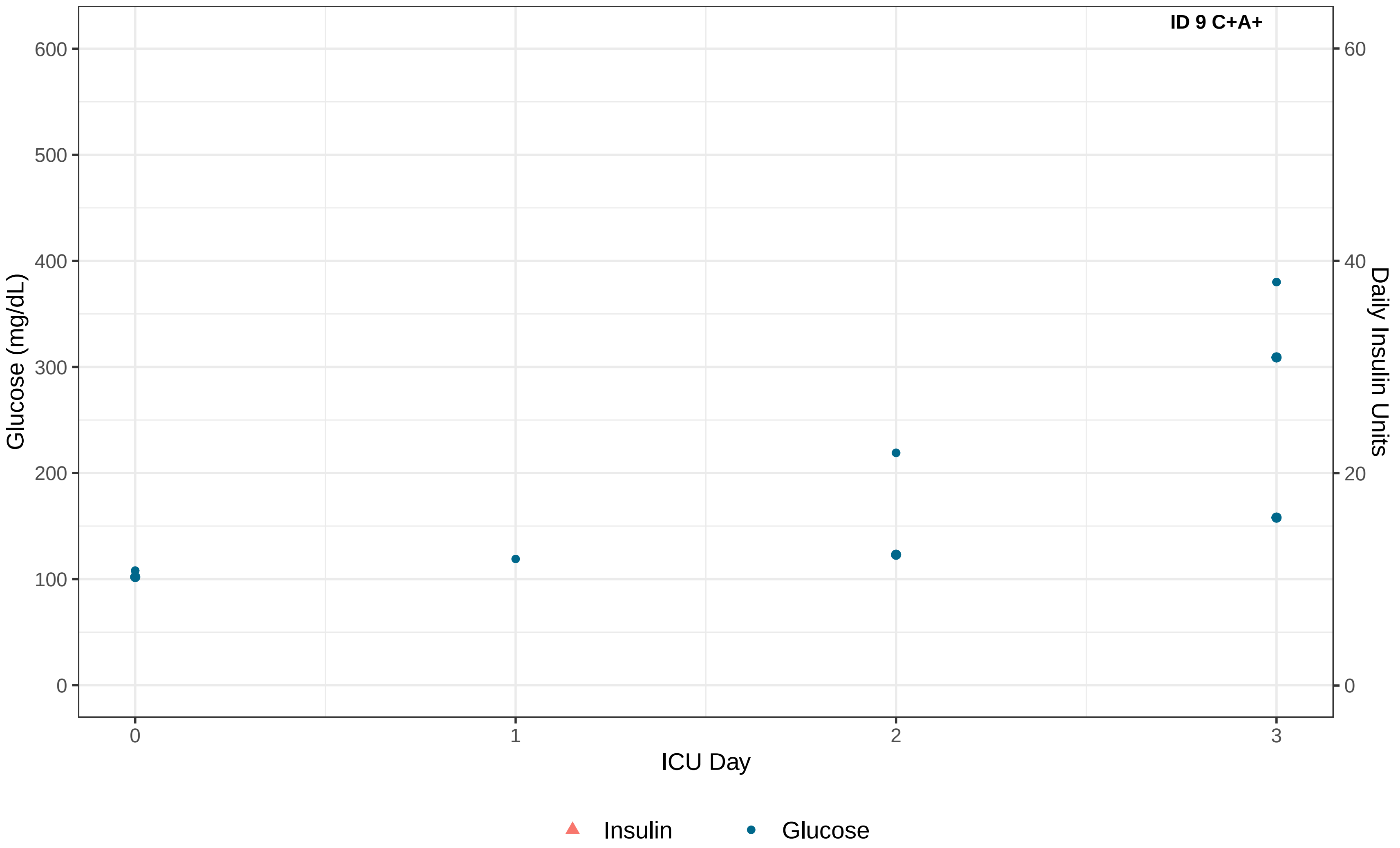

ID 9 C+A+
Glucose (mg/dL)
Daily Insulin Units
ICU Day
0
100
200
300
400
500
600
0
20
40
60
0
1
2
3
Insulin
Glucose

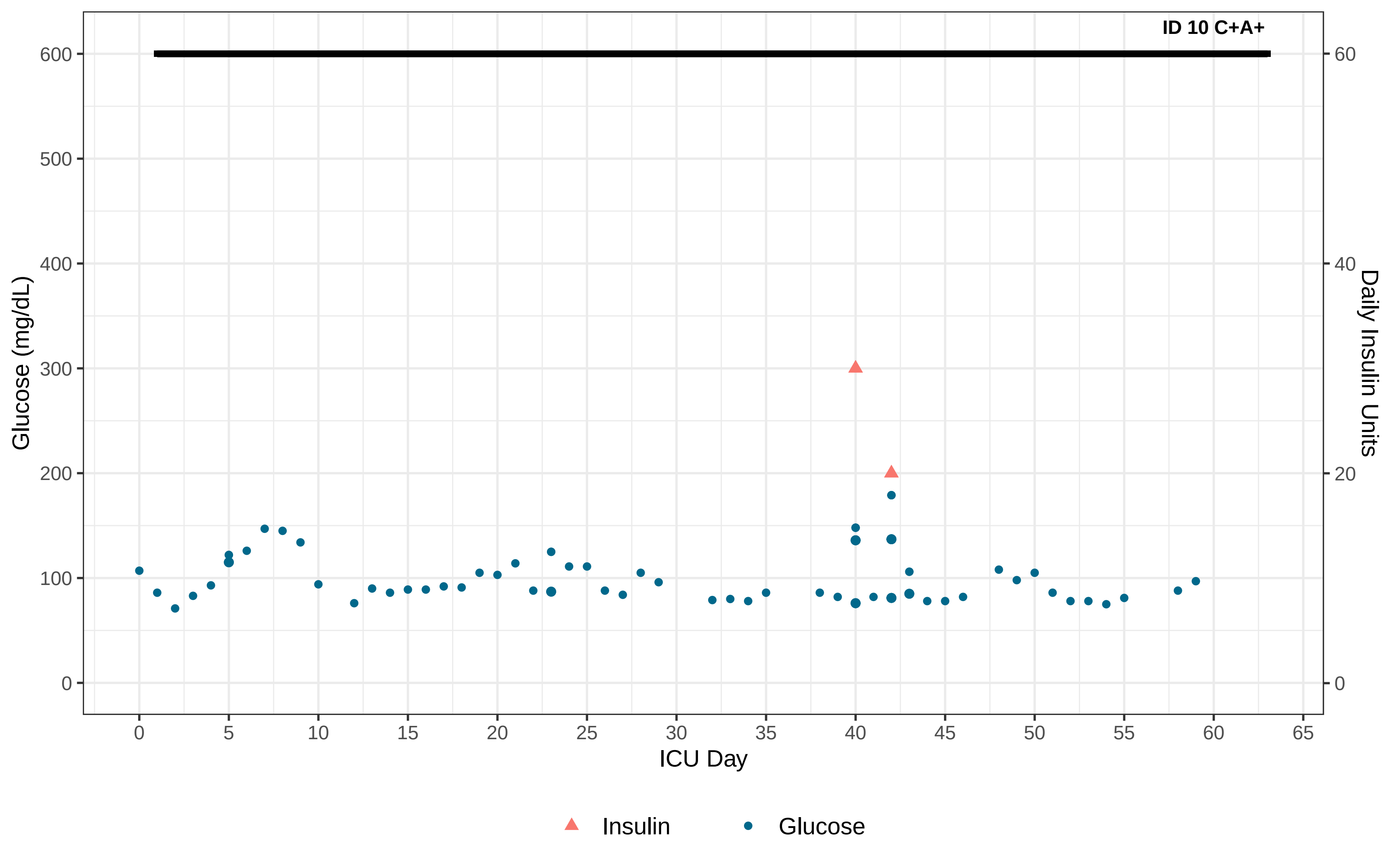

ID 10 C+A+
Glucose (mg/dL)
Daily Insulin Units
ICU Day
0
100
200
300
400
500
600
0
20
40
60
0
5
10
15
20
25
30
35
40
45
50
55
60
65
Insulin
Glucose

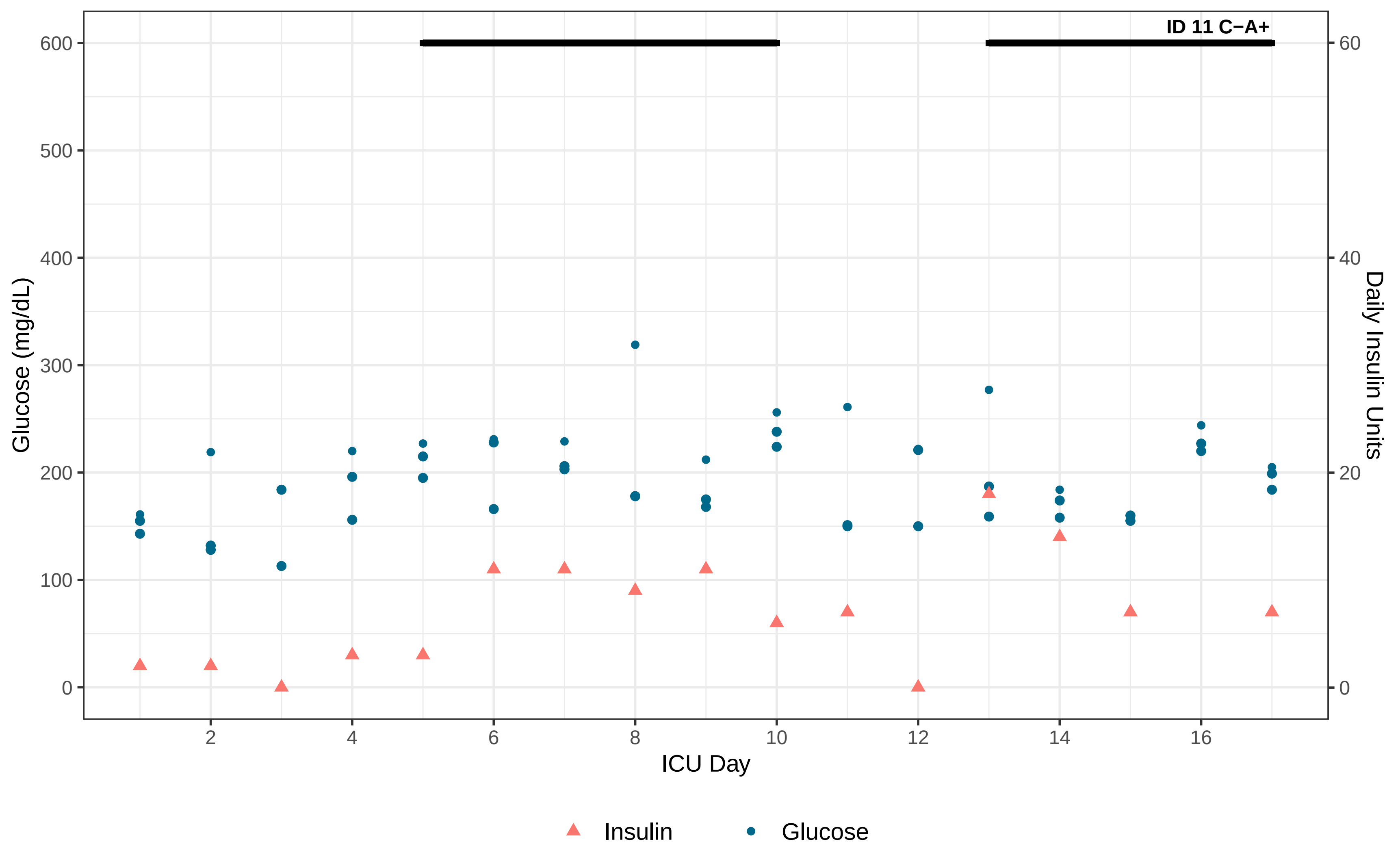
ID 11 C−A+
Glucose (mg/dL)
600
500
400
300
200
100
0
Daily Insulin Units
60
40
20
0
2
4
6
8
10
12
14
16
ICU Day
Insulin
Glucose

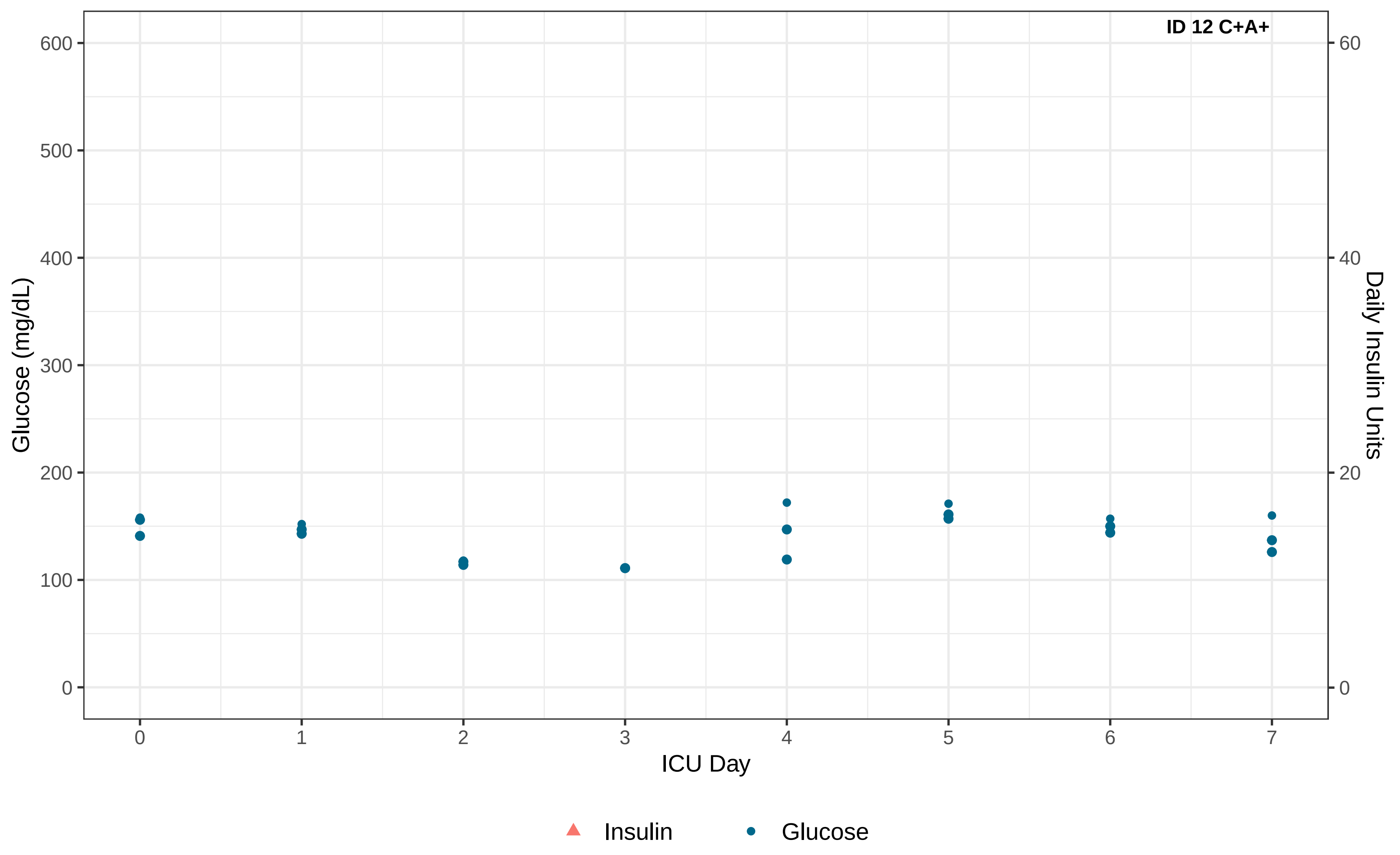

ID 12 C+A+
Glucose (mg/dL)
Daily Insulin Units
ICU Day
0
100
200
300
400
500
600
0
20
40
60
0
1
2
3
4
5
6
7
Insulin
Glucose

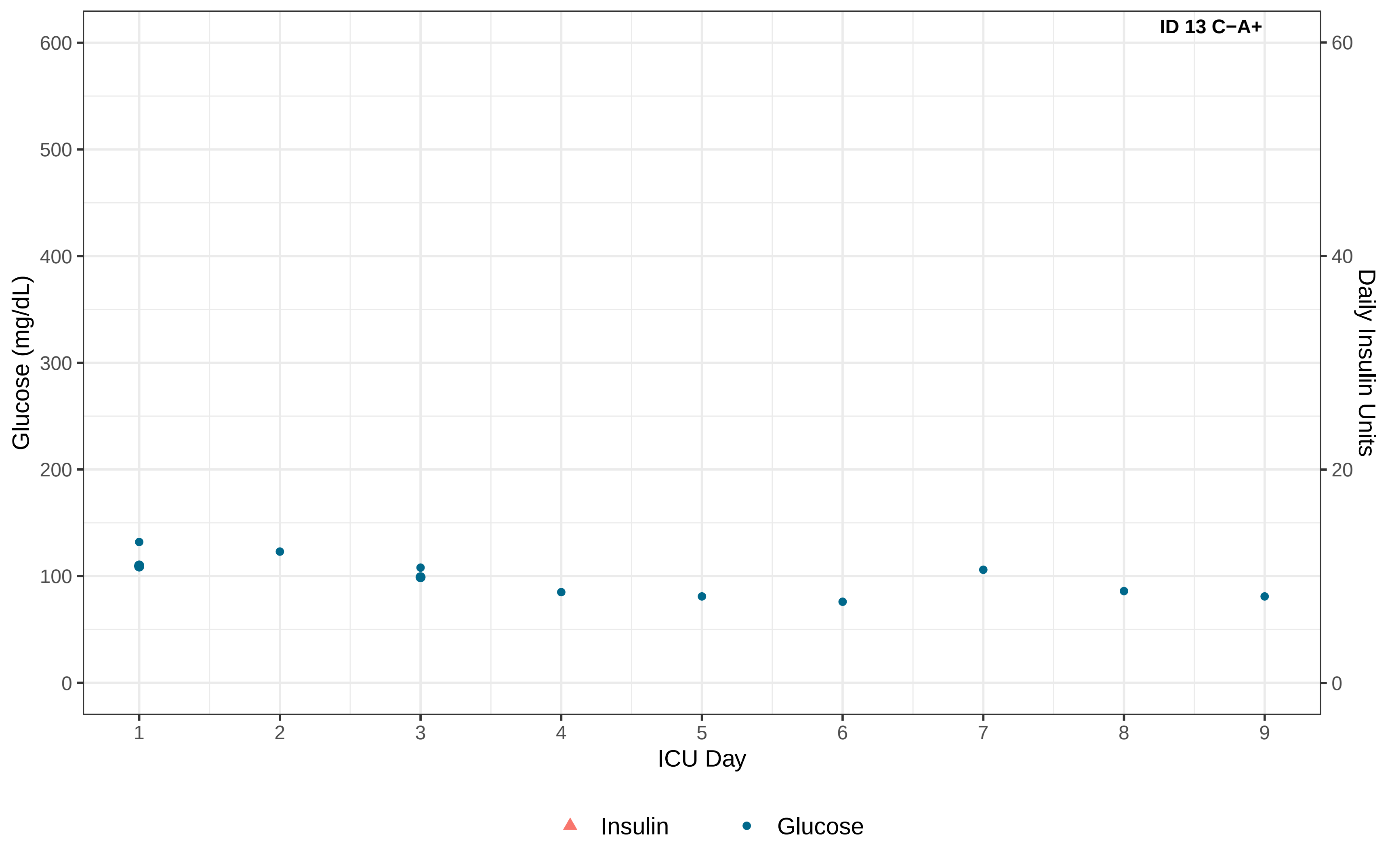

ID 13 C−A+
Glucose (mg/dL)
Daily Insulin Units
ICU Day
0
100
200
300
400
500
600
0
20
40
60
1
2
3
4
5
6
7
8
9
Insulin
Glucose

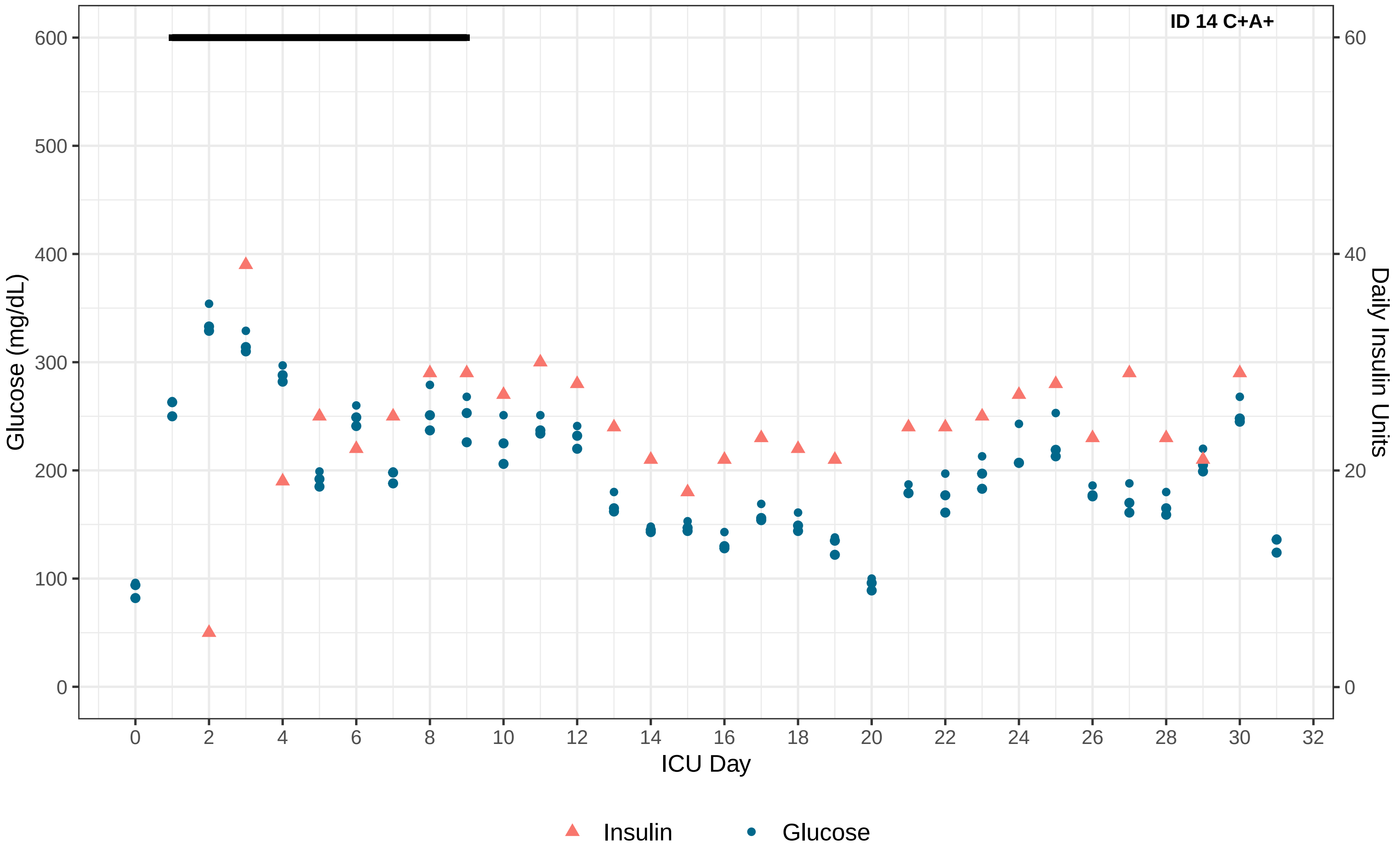

ID 14 C+A+
Glucose (mg/dL)
Daily Insulin Units
ICU Day
0
100
200
300
400
500
600
0
20
40
60
0 2 4 6 8 10 12 14 16 18 20 22 24 26 28 30 32
Insulin
Glucose

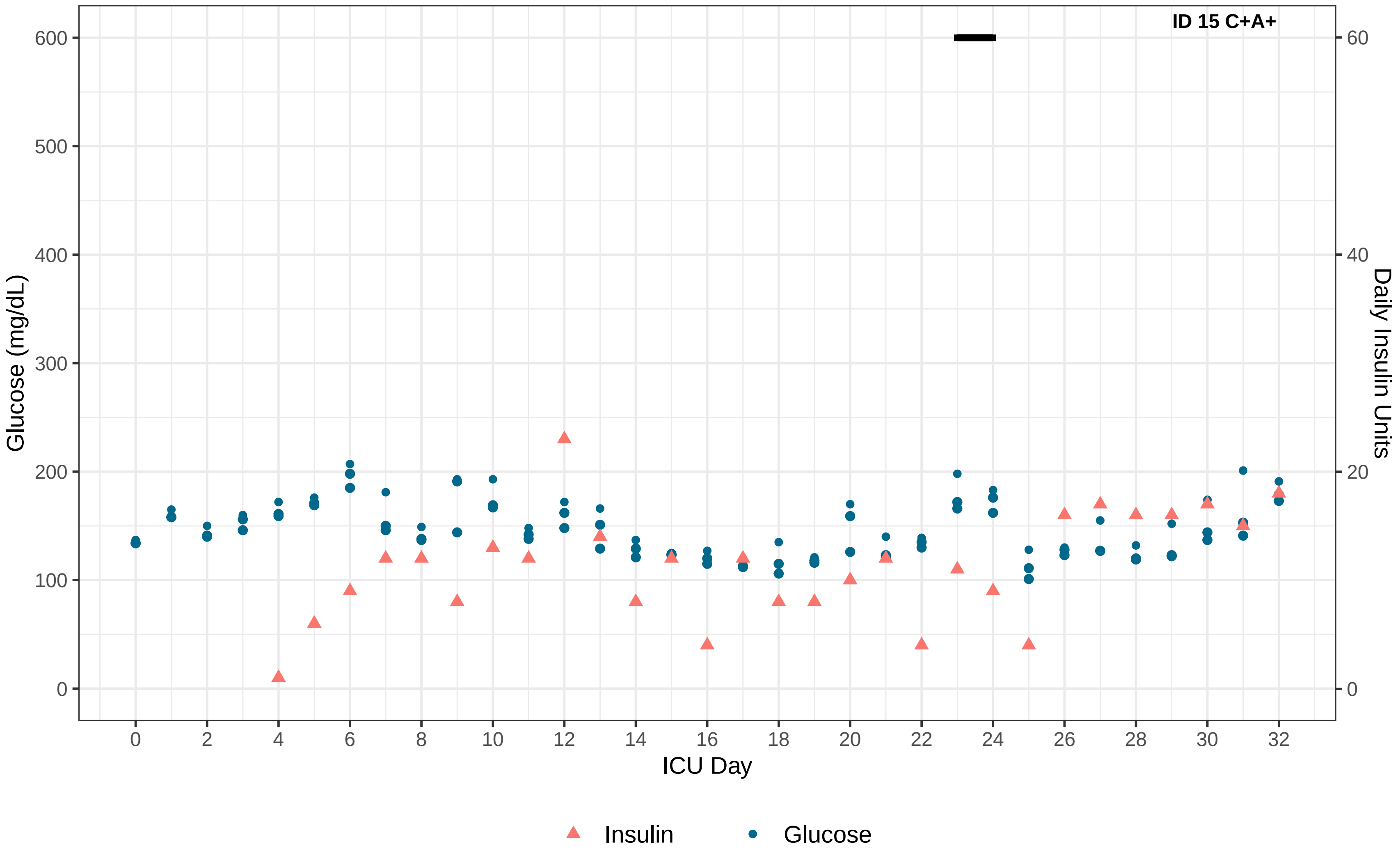
ID 15 C+A+
Glucose (mg/dL)
Daily Insulin Units
ICU Day
0
100
200
300
400
500
600
0
20
40
60
0
2
4
6
8
10
12
14
16
18
20
22
24
26
28
30
32
Insulin
Glucose

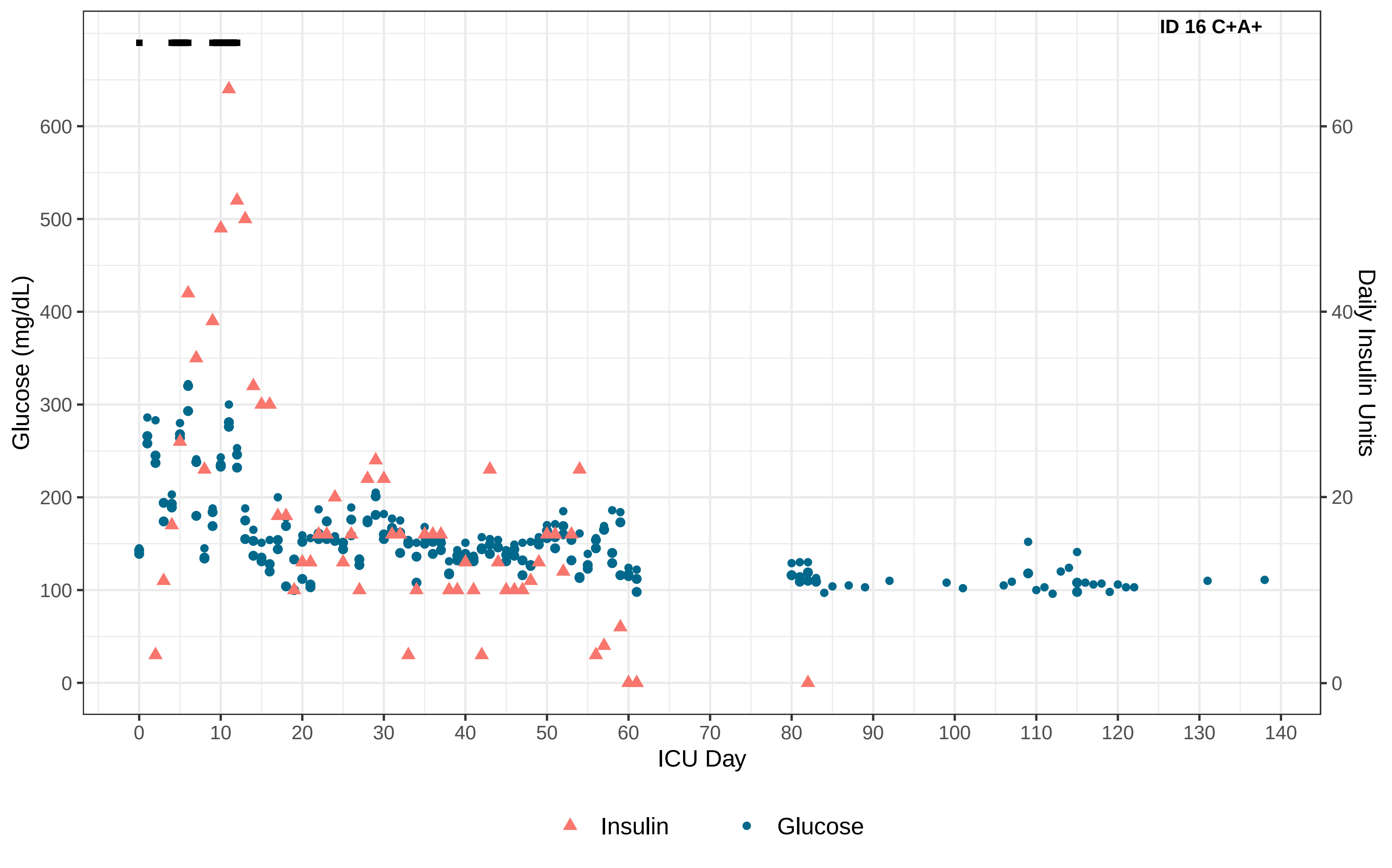

ID 16 C+A+
Glucose (mg/dL)
Daily Insulin Units
ICU Day
0
100
200
300
400
500
600
0
20
40
60
0
10
20
30
40
50
60
70
80
90
100
110
120
130
140
Insulin
Glucose

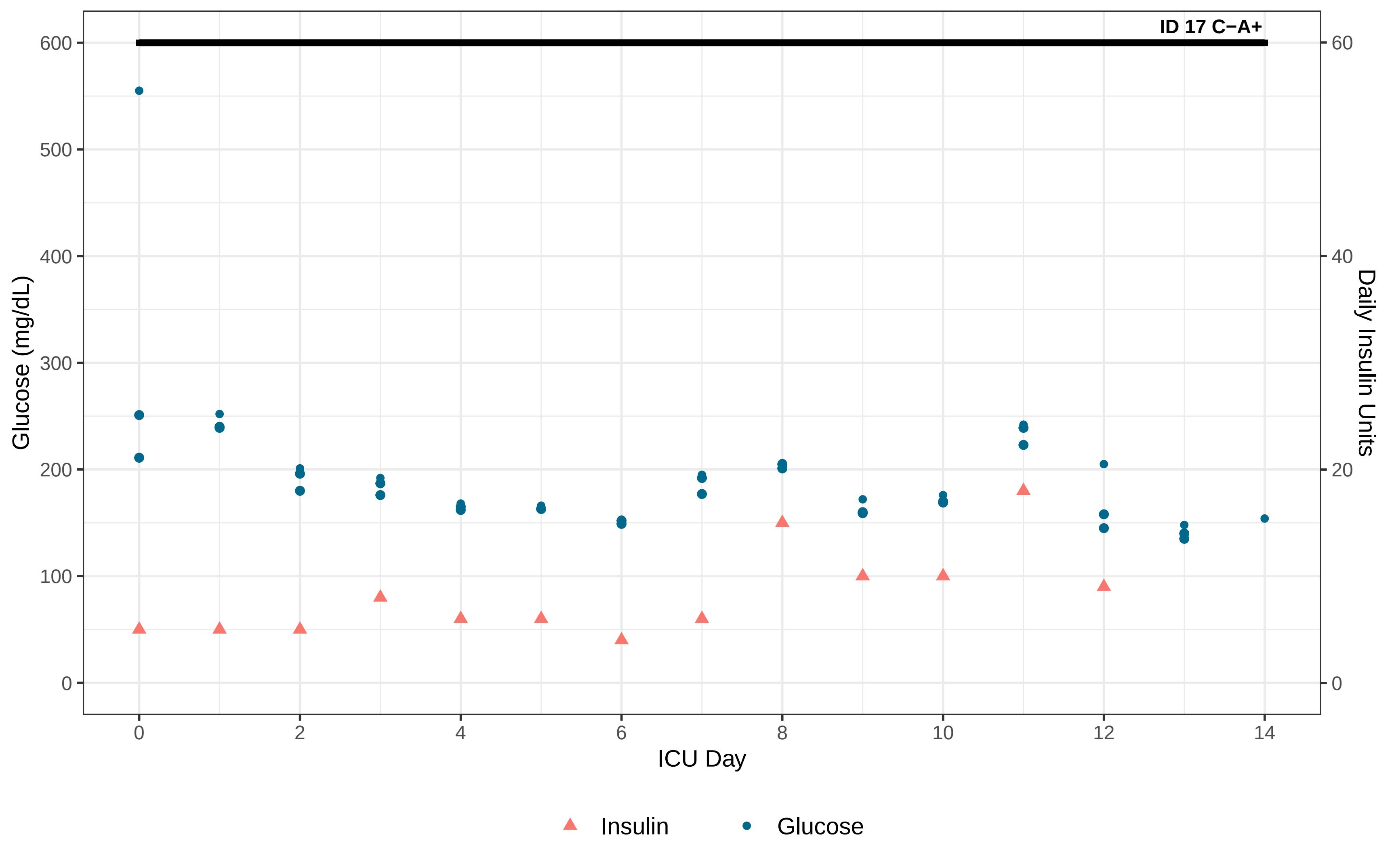

ID 17 C−A+
Glucose (mg/dL)
Daily Insulin Units
ICU Day
0
100
200
300
400
500
600
0
20
40
60
0
2
4
6
8
10
12
14
Insulin
Glucose

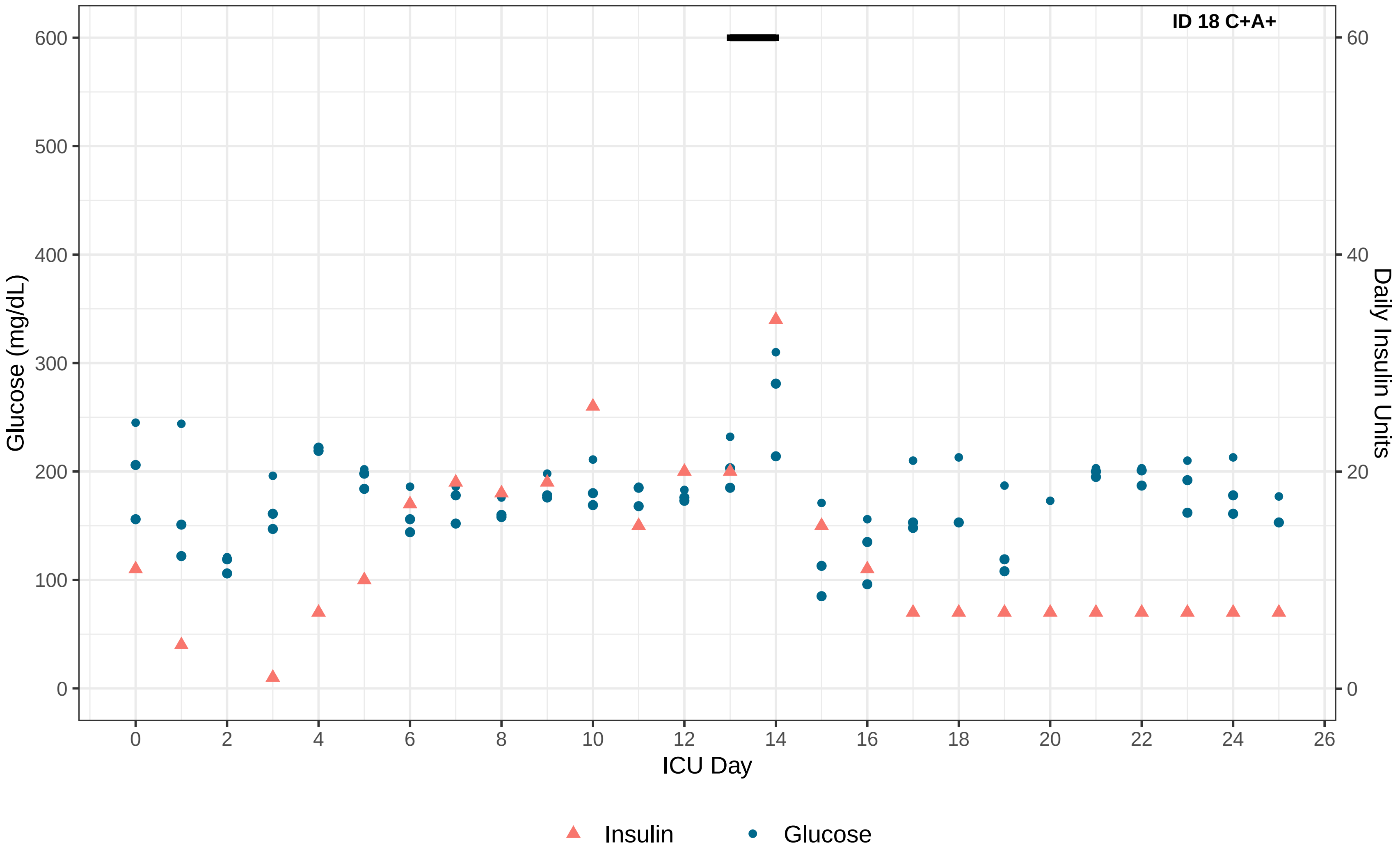

ID 18 C+A+
Glucose (mg/dL)
Daily Insulin Units
ICU Day
0
100
200
300
400
500
600
0
20
40
60
0
2
4
6
8
10
12
14
16
18
20
22
24
26
Insulin
Glucose

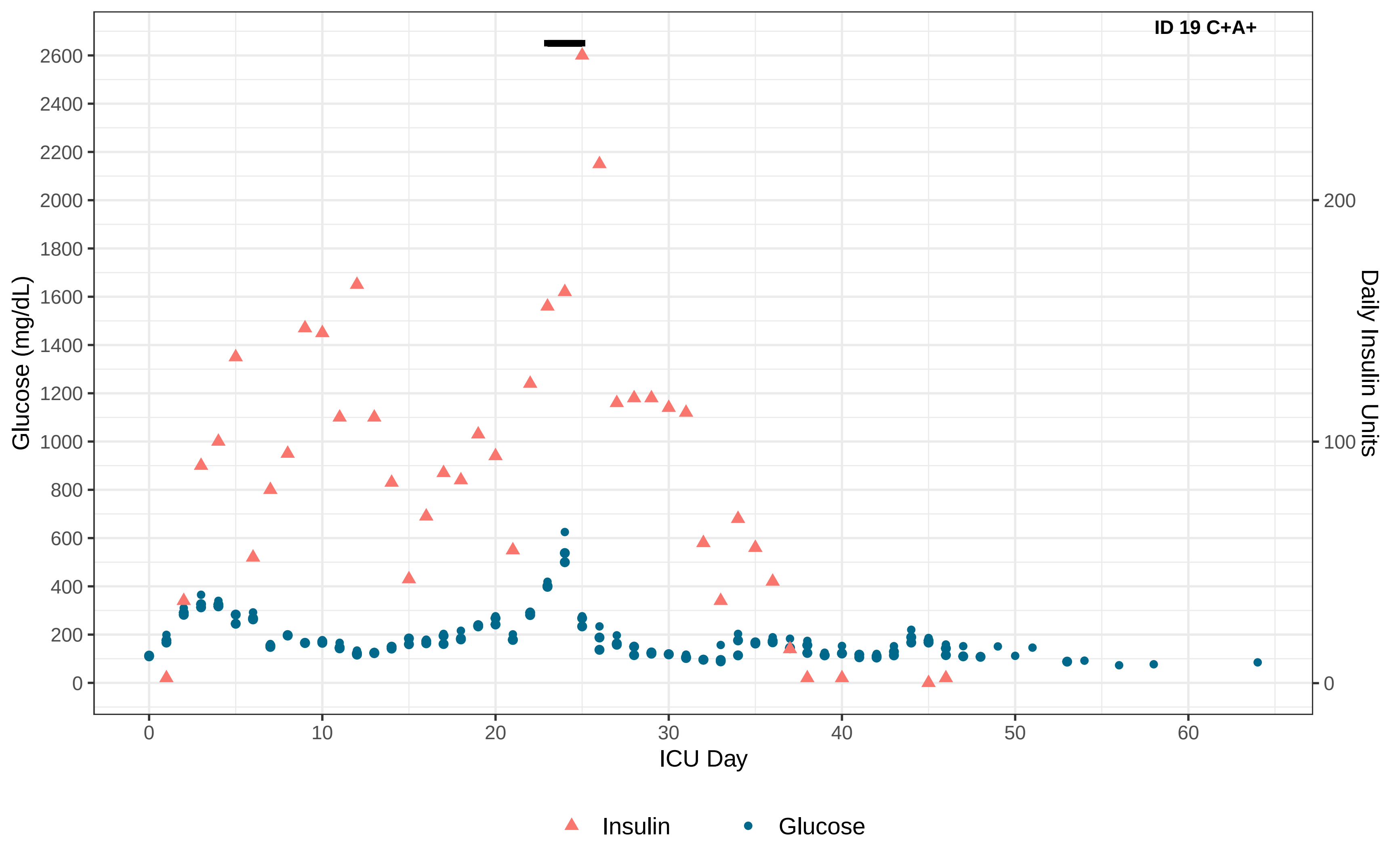

ID 19 C+A+
Glucose (mg/dL)
2600
2400
2200
2000
1800
1600
1400
1200
1000
800
600
400
200
0
Daily Insulin Units
200
100
0
ICU Day
0
10
20
30
40
50
60
Insulin
Glucose

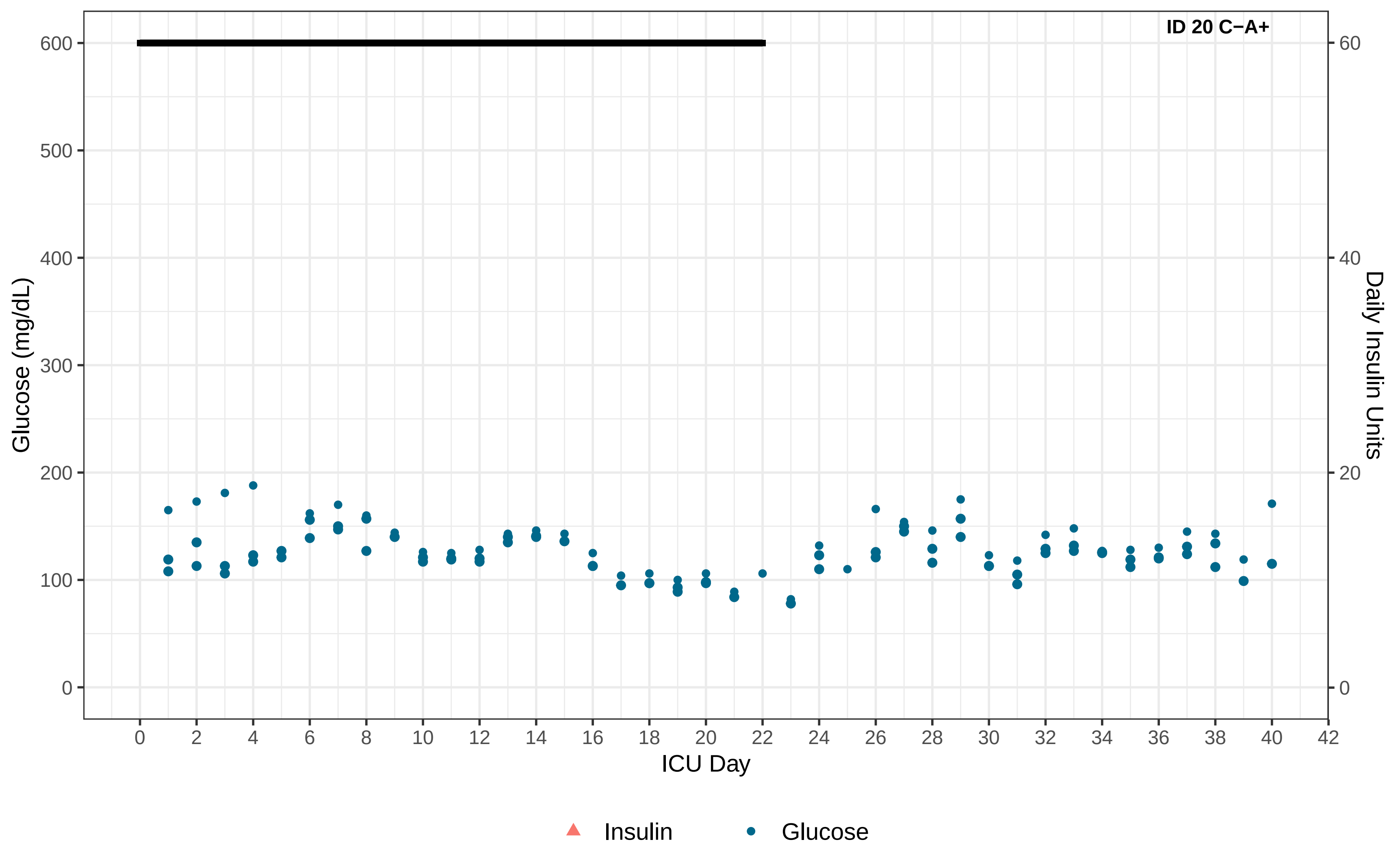
ID 20 C−A+
Glucose (mg/dL)
Daily Insulin Units
ICU Day
0
100
200
300
400
500
600
0
20
40
60
0
2
4
6
8
10
12
14
16
18
20
22
24
26
28
30
32
34
36
38
40
42
Insulin
Glucose

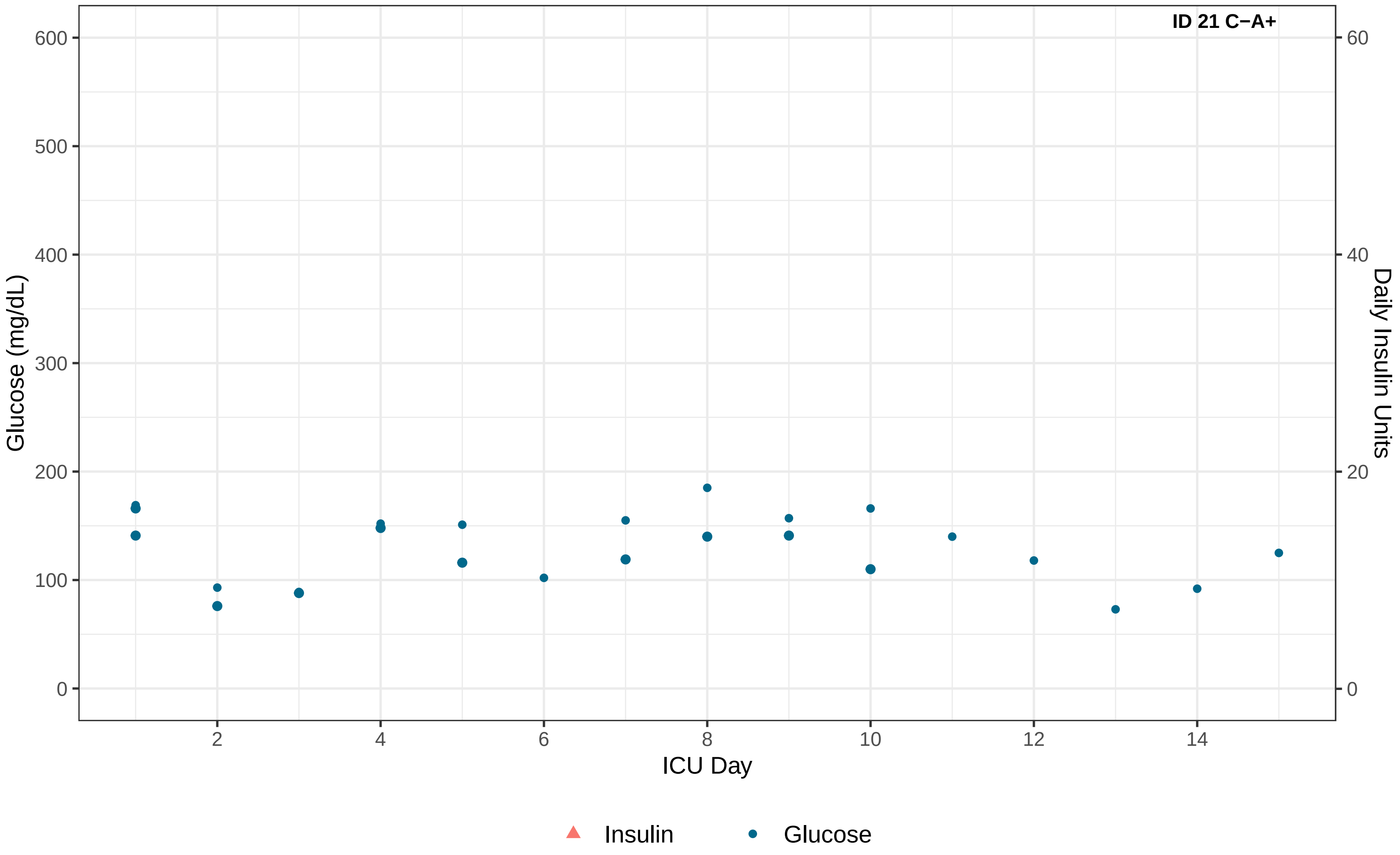

ID 21 C−A+
Glucose (mg/dL)
Daily Insulin Units
ICU Day
0
100
200
300
400
500
600
0
20
40
60
2
4
6
8
10
12
14
Insulin
Glucose

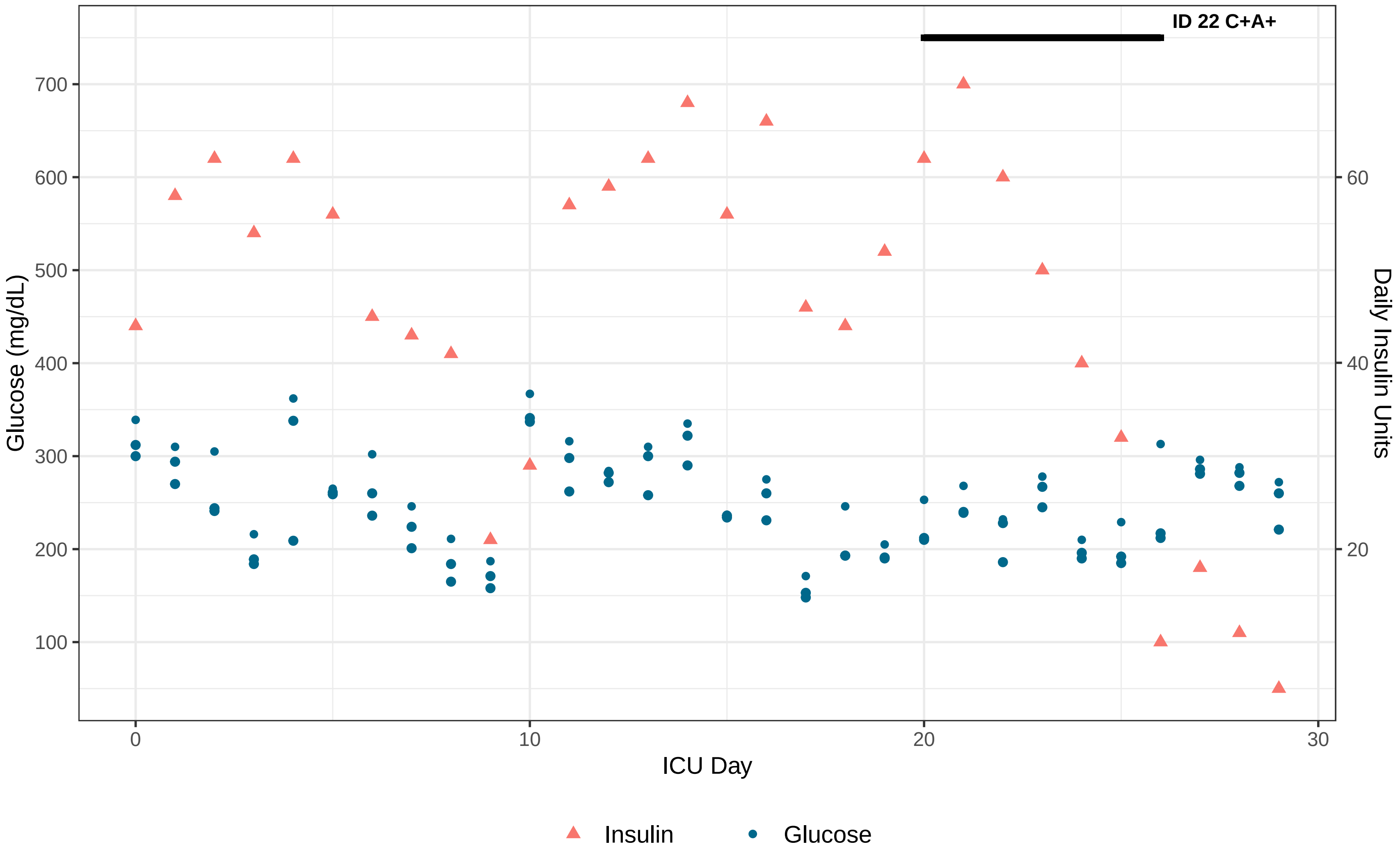

ID 22 C+A+
Glucose (mg/dL)
700
600
500
400
300
200
100
Daily Insulin Units
60
40
20
0
10
20
30
ICU Day
Insulin
Glucose

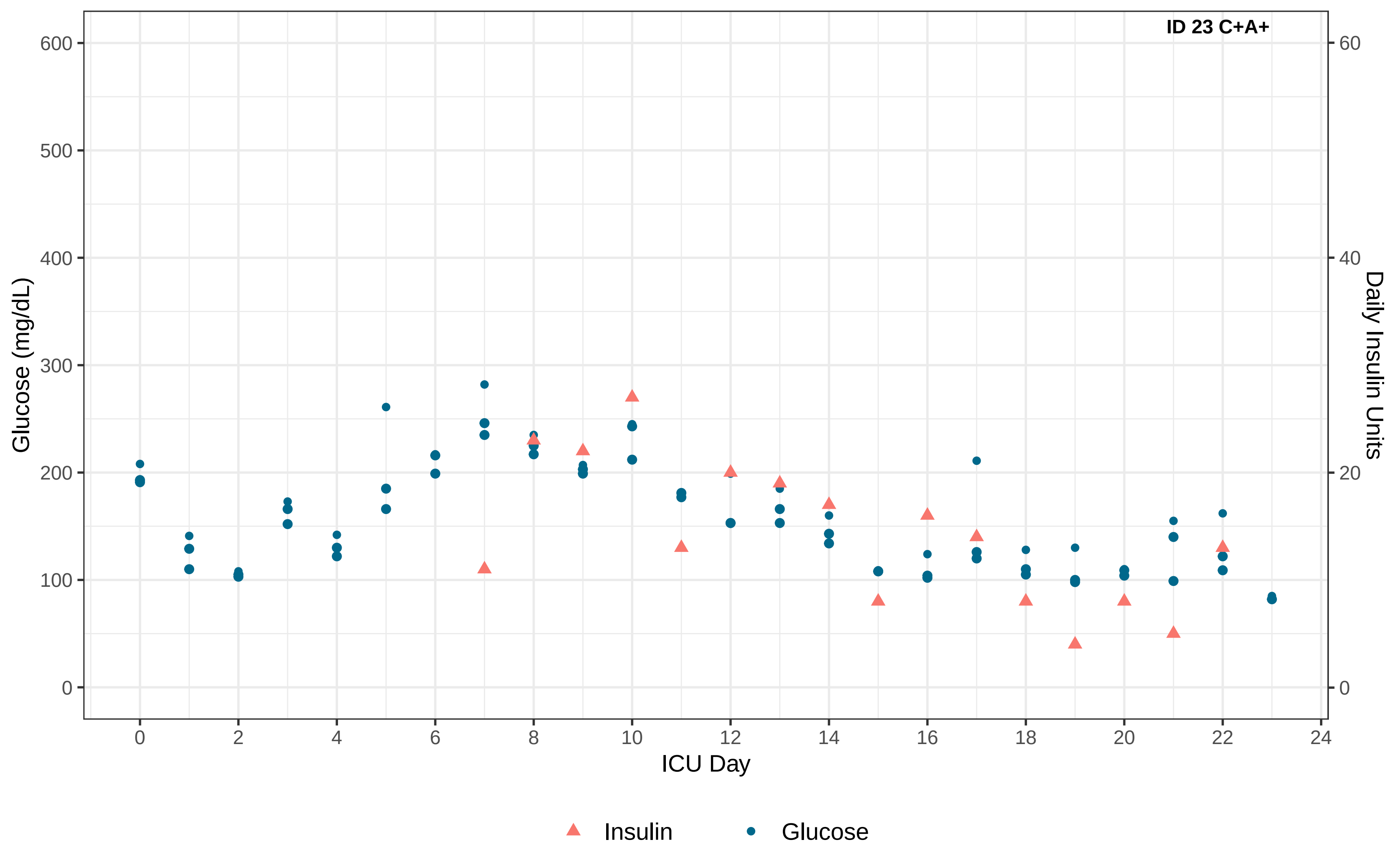
ID 23 C+A+
Glucose (mg/dL)
Daily Insulin Units
ICU Day
0
100
200
300
400
500
600
0
20
40
60
0
2
4
6
8
10
12
14
16
18
20
22
24
Insulin
Glucose

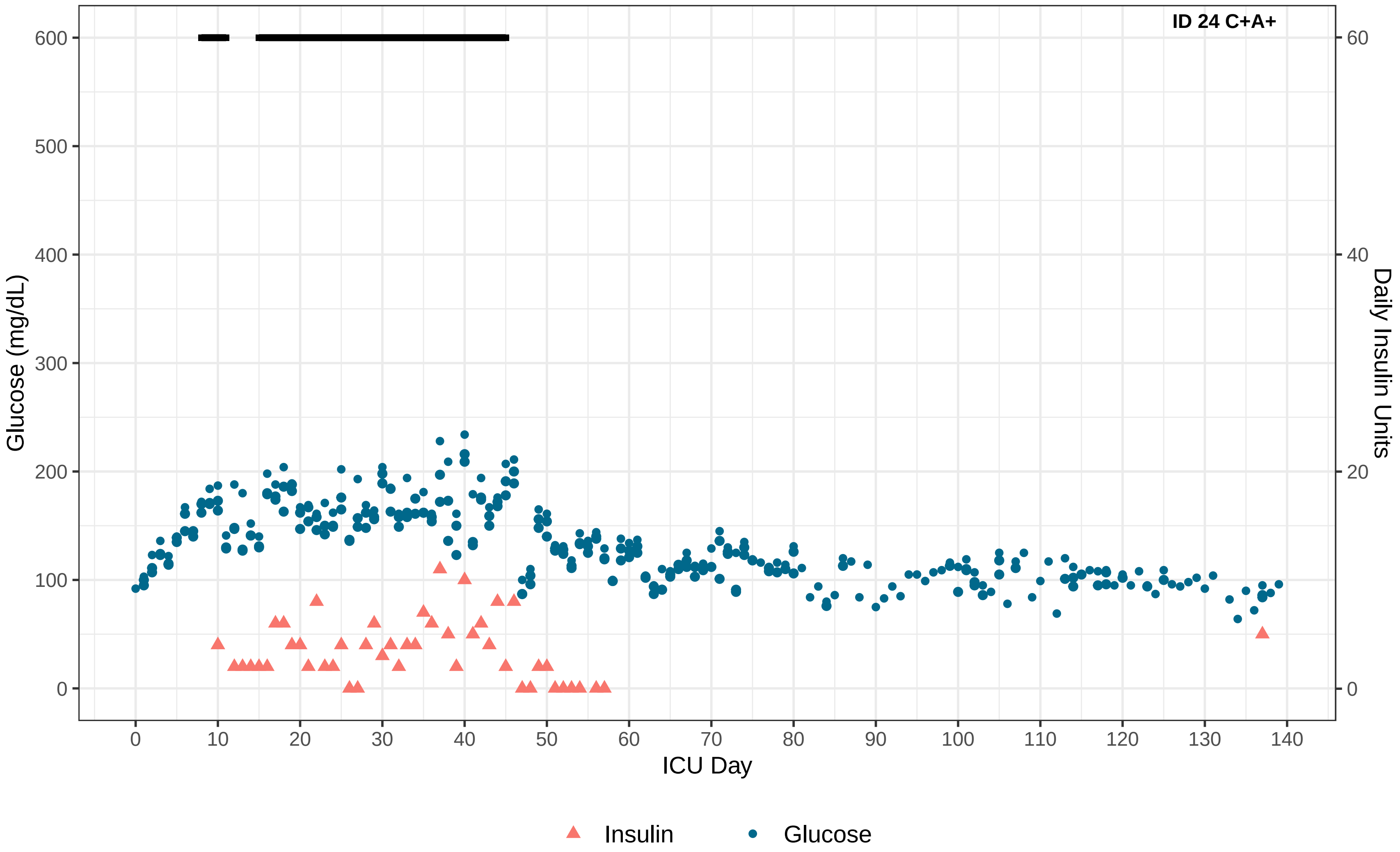

ID 24 C+A+
Glucose (mg/dL)
Daily Insulin Units
ICU Day
0
100
200
300
400
500
600
0
20
40
60
0
10
20
30
40
50
60
70
80
90
100
110
120
130
140
Insulin
Glucose

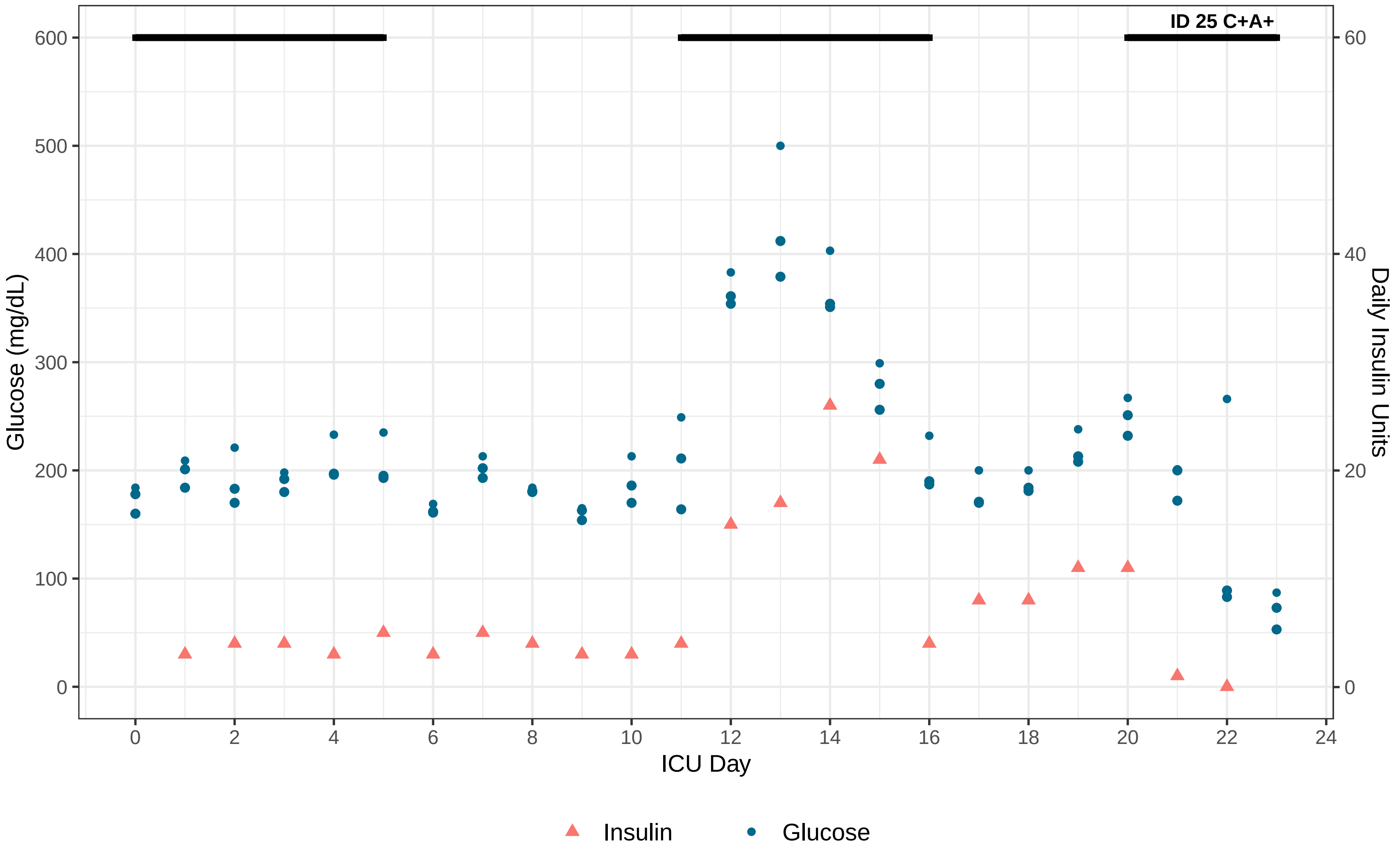
ID 25 C+A+
Glucose (mg/dL)
Daily Insulin Units
ICU Day
0
100
200
300
400
500
600
0
20
40
60
0
2
4
6
8
10
12
14
16
18
20
22
24
Insulin
Glucose

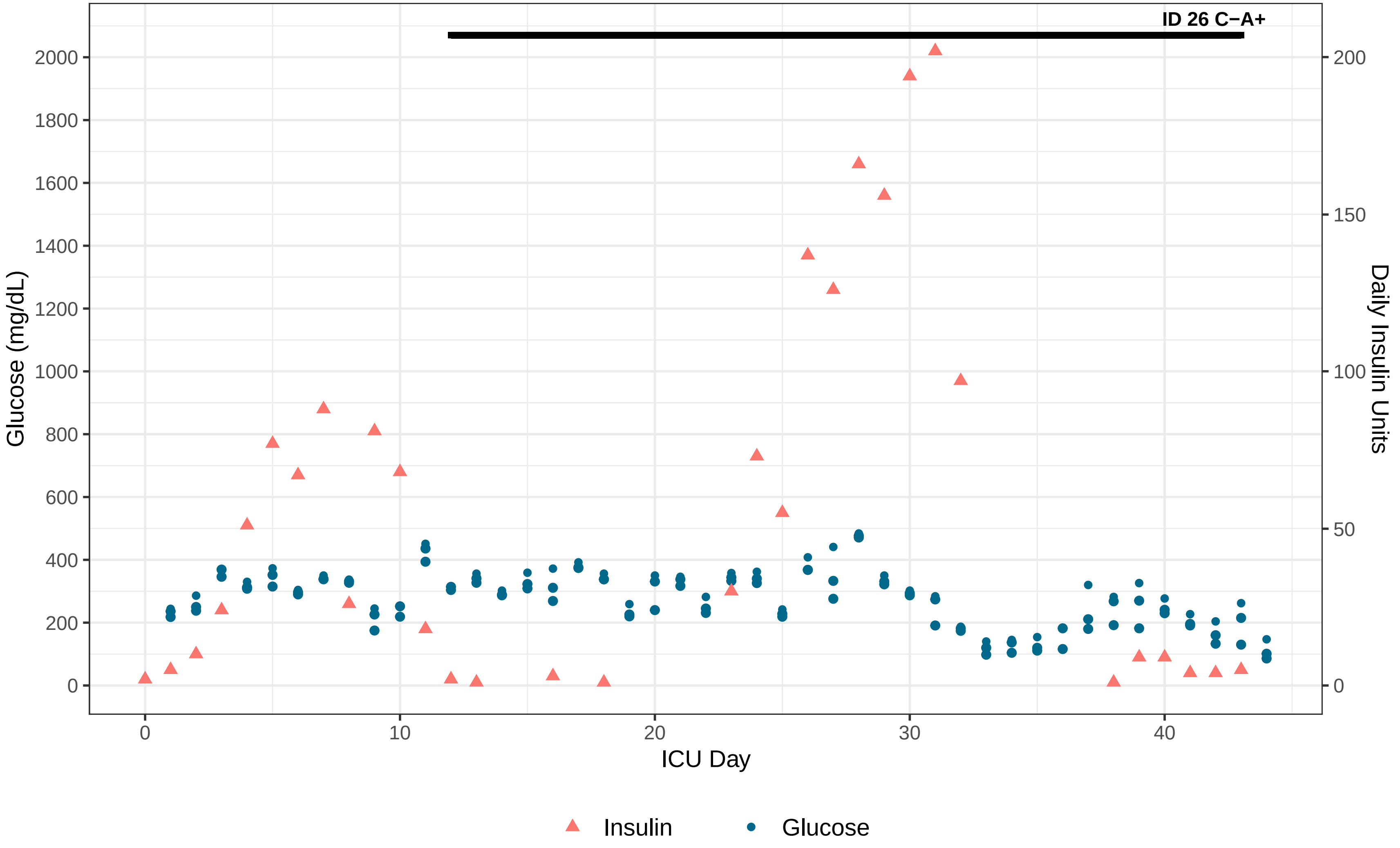

ID 26 C−A+
Glucose (mg/dL)
Daily Insulin Units
ICU Day
2000
1800
1600
1400
1200
1000
800
600
400
200
0
200
150
100
50
0
0
10
20
30
40
Insulin
Glucose

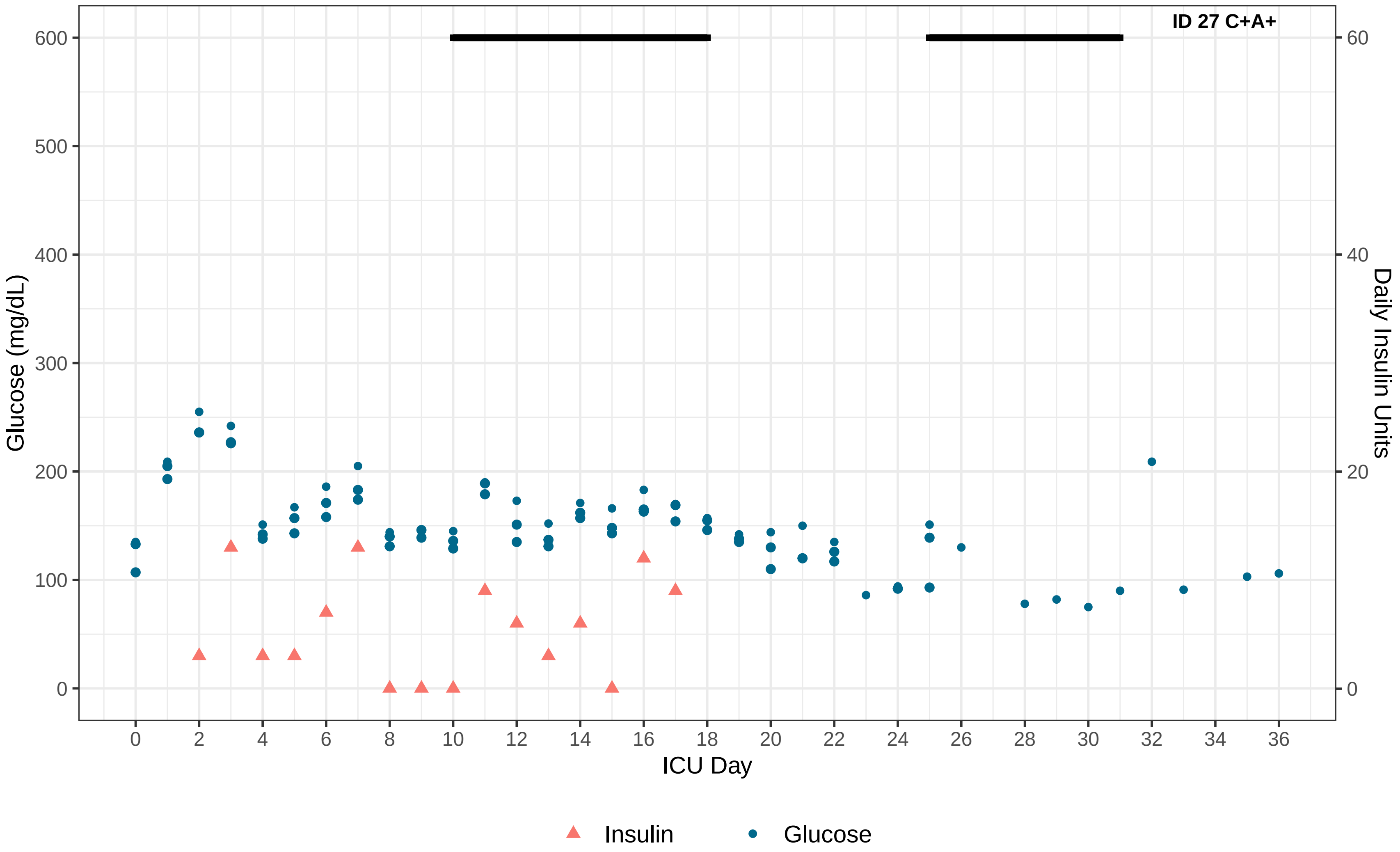

ID 27 C+A+
Glucose (mg/dL)
Daily Insulin Units
ICU Day
0
100
200
300
400
500
600
0
20
40
60
0
2
4
6
8
10
12
14
16
18
20
22
24
26
28
30
32
34
36
Insulin
Glucose

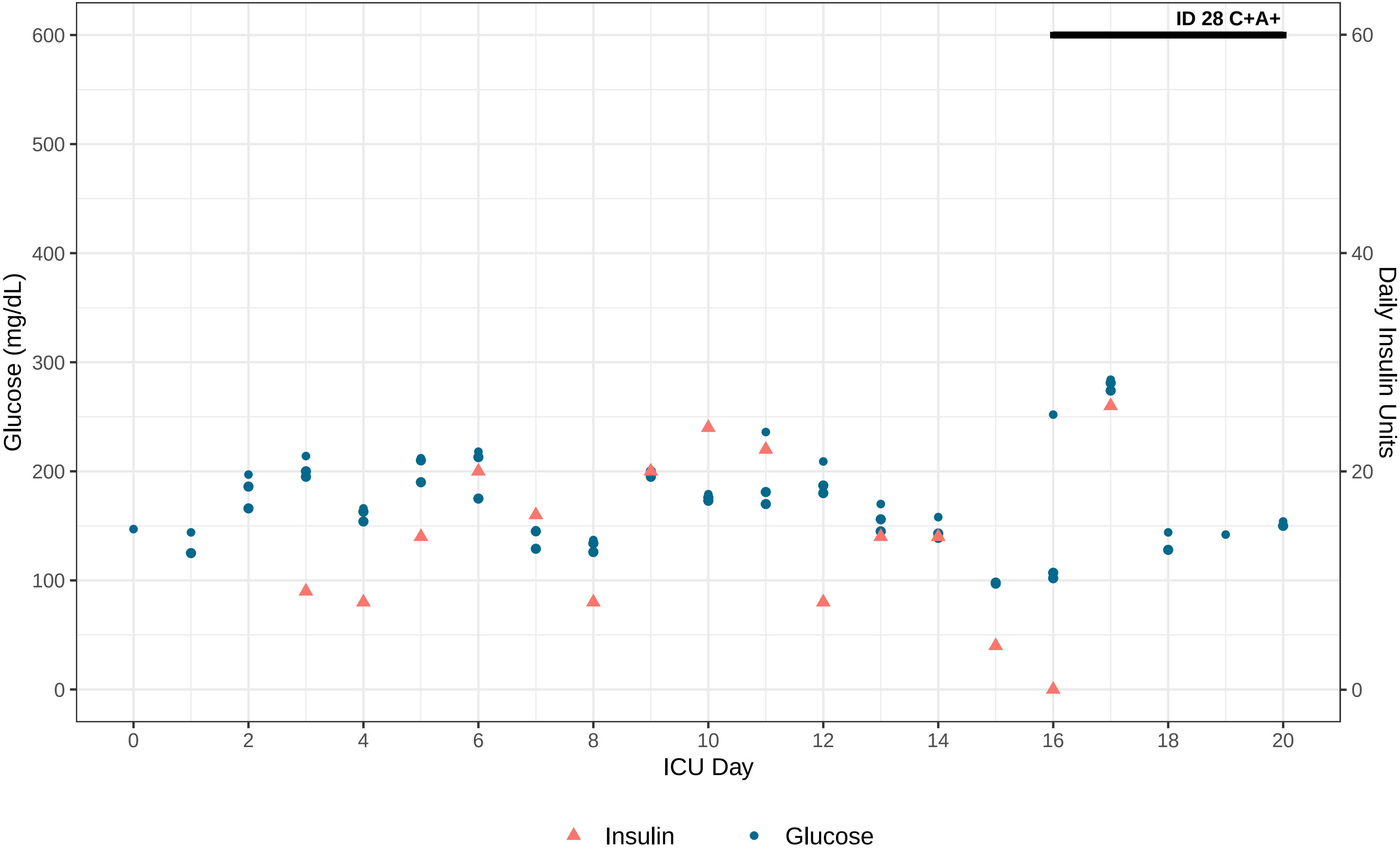

ID 28 C+A+
Glucose (mg/dL)
Daily Insulin Units
ICU Day
600
500
400
300
200
100
0
60
40
20
0
0
2
4
6
8
10
12
14
16
18
20
Insulin
Glucose

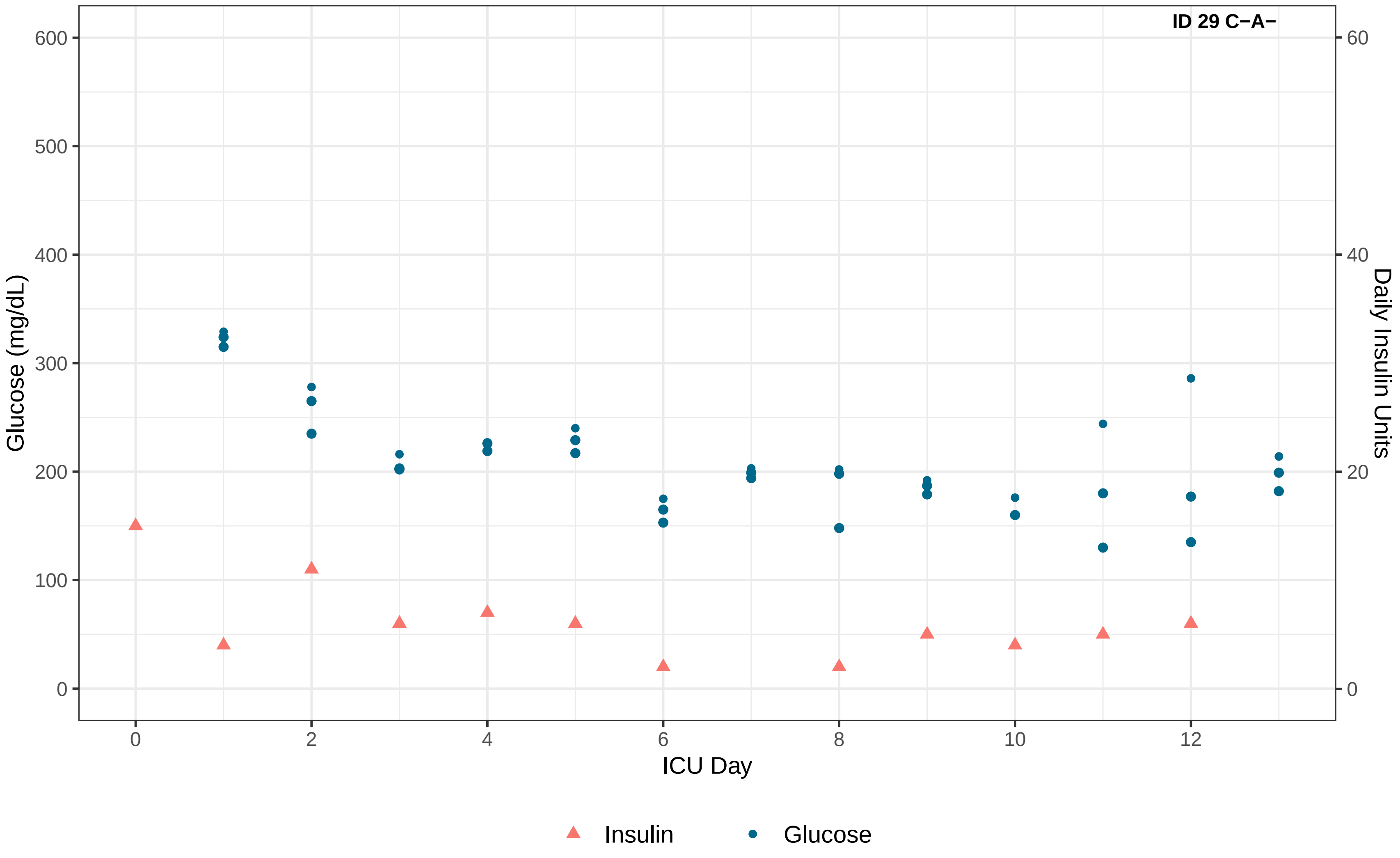

ID 29 C−A−
Glucose (mg/dL)
Daily Insulin Units
ICU Day
0
100
200
300
400
500
600
0
20
40
60
0
2
4
6
8
10
12
Insulin
Glucose

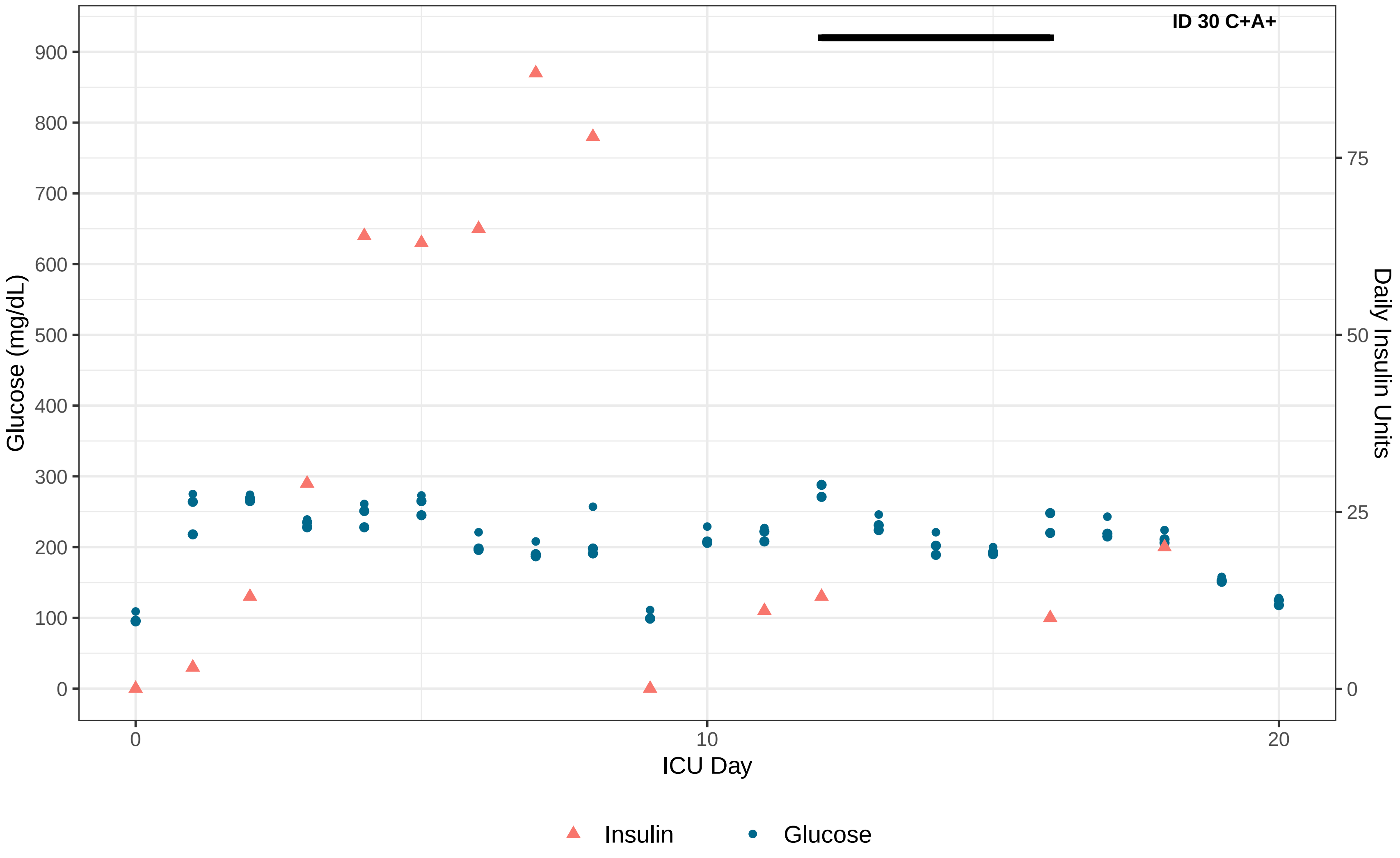

ID 30 C+A+
Glucose (mg/dL)
Daily Insulin Units
ICU Day
0
100
200
300
400
500
600
700
800
900
0
25
50
75
0
10
20
Insulin
Glucose

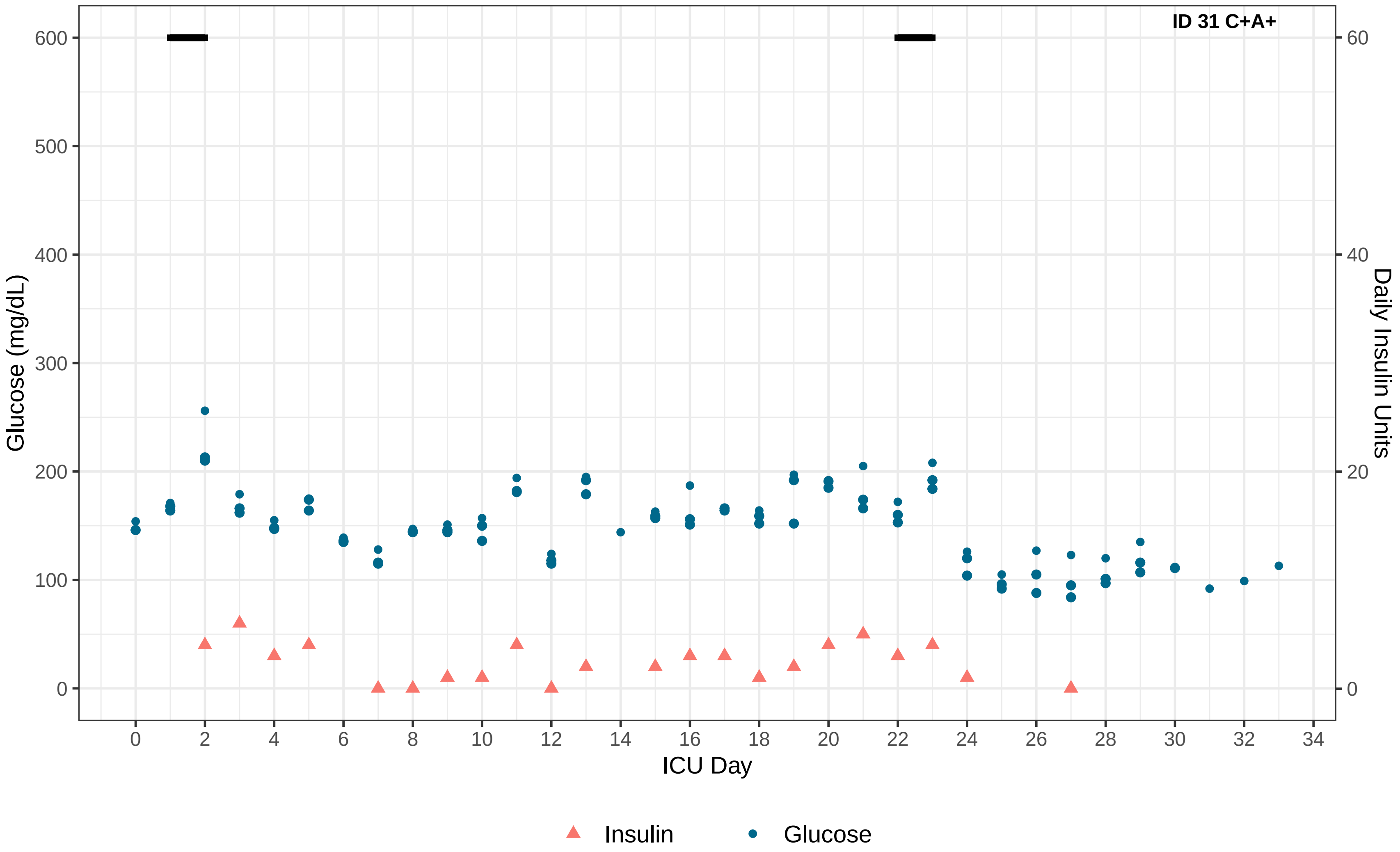
ID 31 C+A+
Glucose (mg/dL)
Daily Insulin Units
ICU Day
0
100
200
300
400
500
600
20
40
60
2
4
6
8
10
12
14
16
18
20
22
24
26
28
30
32
34
Insulin
Glucose

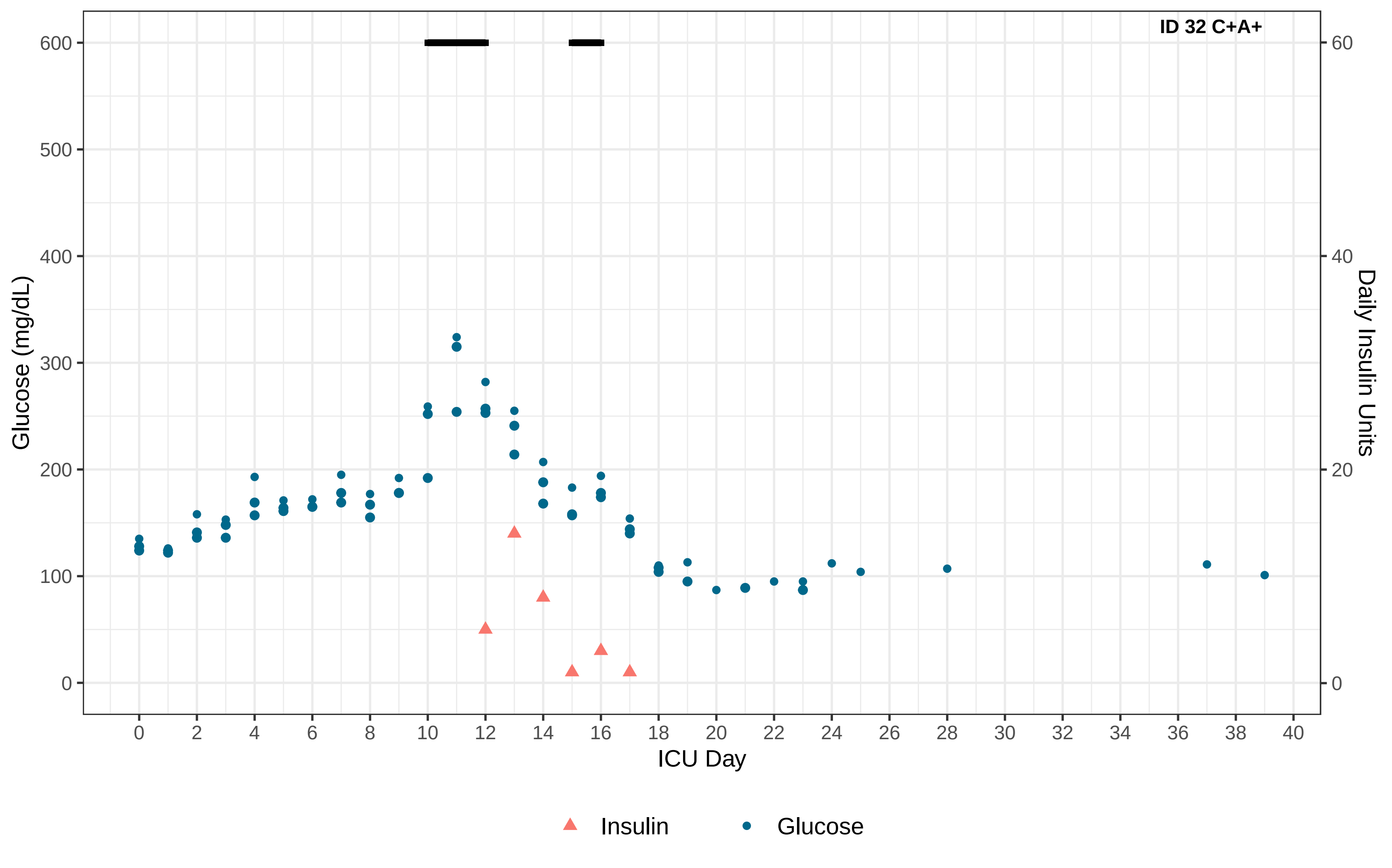

ID 32 C+A+
Glucose (mg/dL)
Daily Insulin Units
ICU Day
0
100
200
300
400
500
600
0
20
40
60
0 2 4 6 8 10 12 14 16 18 20 22 24 26 28 30 32 34 36 38 40
Insulin
Glucose

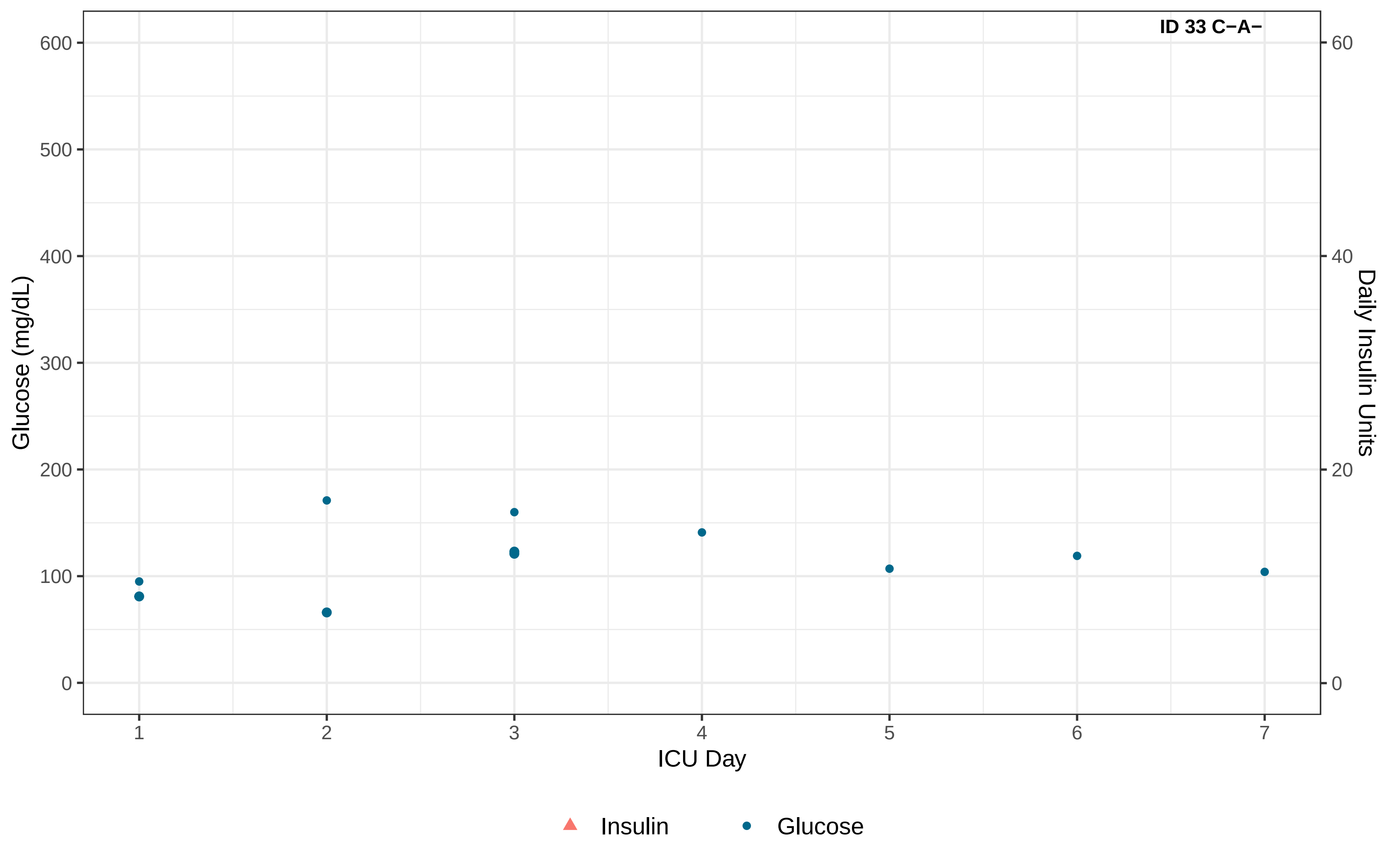

ID 33 C−A−
600
500
400
300
200
100
0
Glucose (mg/dL)
60
40
20
0
Daily Insulin Units
1
2
3
4
5
6
7
ICU Day
Insulin
Glucose

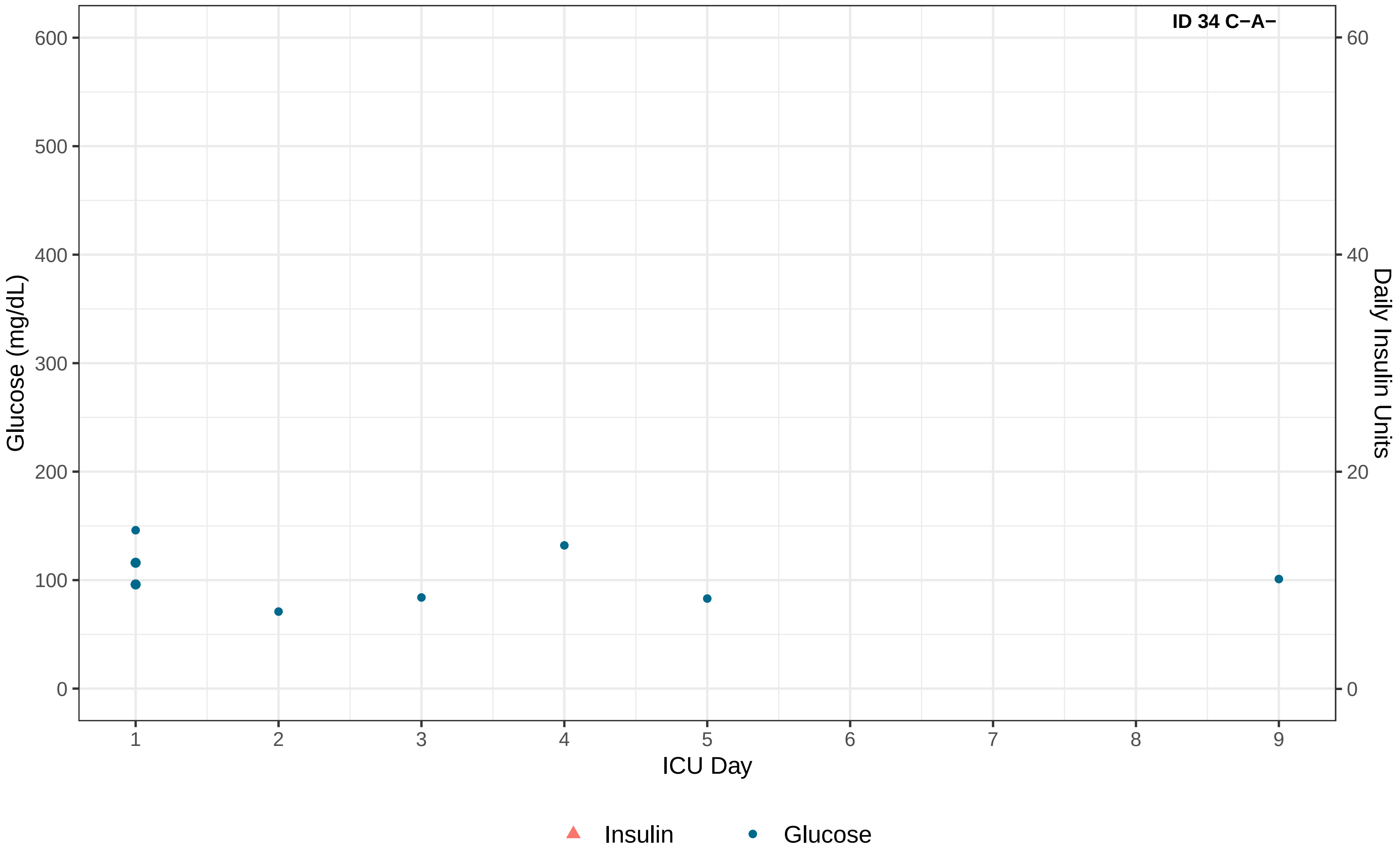
ID 34 C−A−
Glucose (mg/dL)
600
500
400
300
200
100
0
Daily Insulin Units
60
40
20
0
1
2
3
4
5
6
7
8
9
ICU Day
Insulin
Glucose

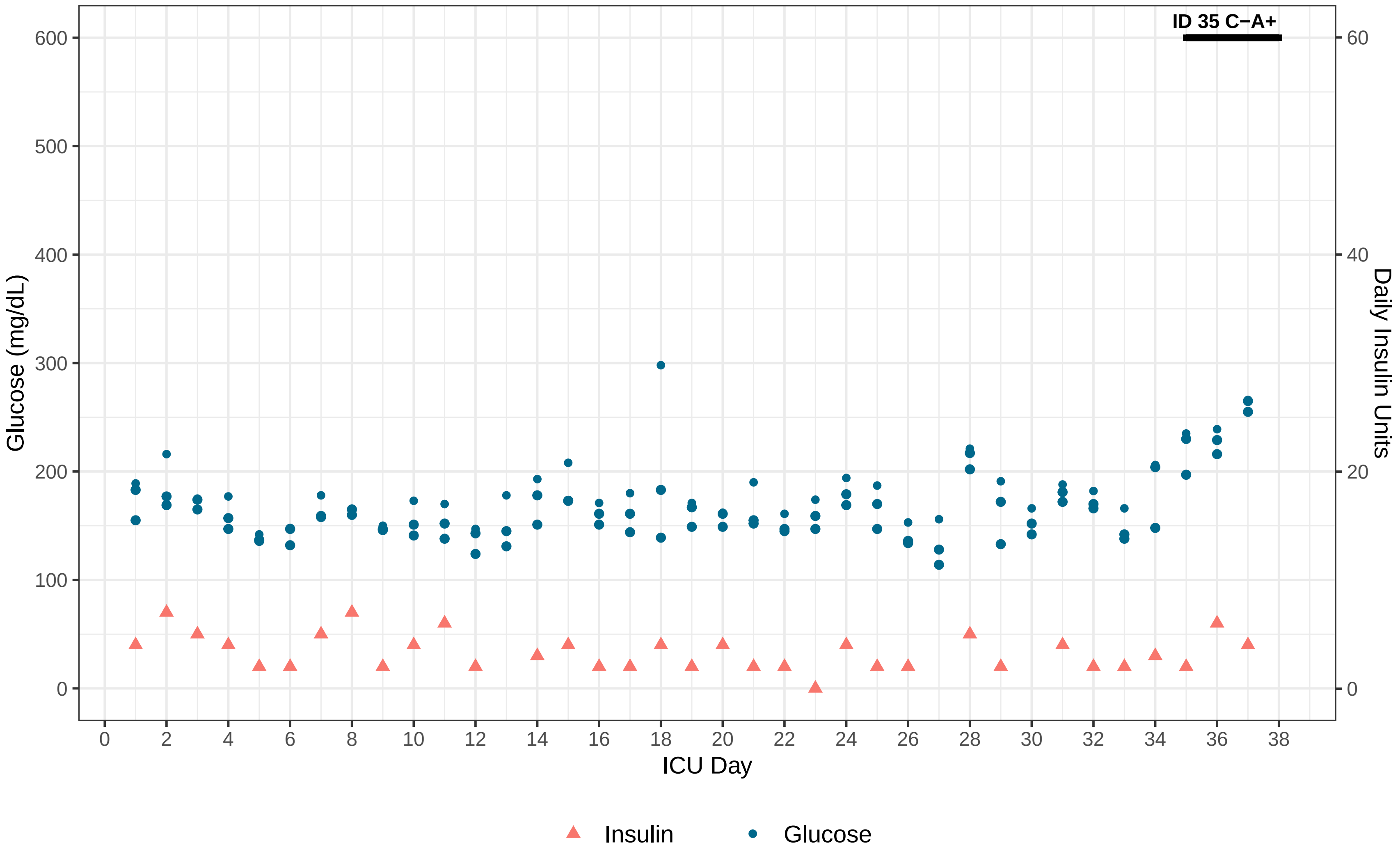
ID 35 C−A+
Glucose (mg/dL)
Daily Insulin Units
ICU Day
0
100
200
300
400
500
600
0
20
40
60
0
2
4
6
8
10
12
14
16
18
20
22
24
26
28
30
32
34
36
38
Insulin
Glucose

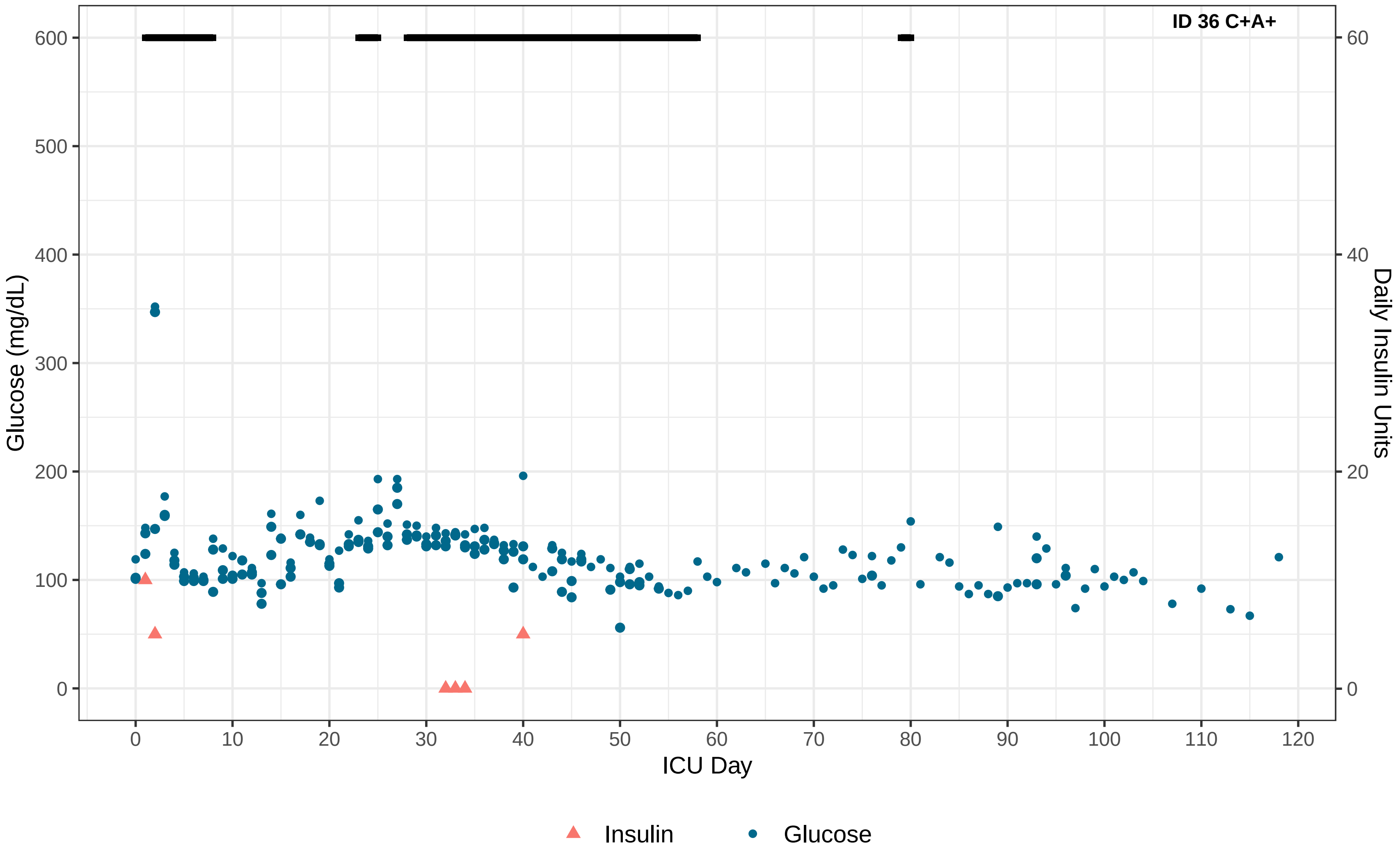
ID 36 C+A+
Glucose (mg/dL)
Daily Insulin Units
ICU Day
0
100
200
300
400
500
600
20
40
60
10
30
50
70
80
90
110
120
Insulin
Glucose

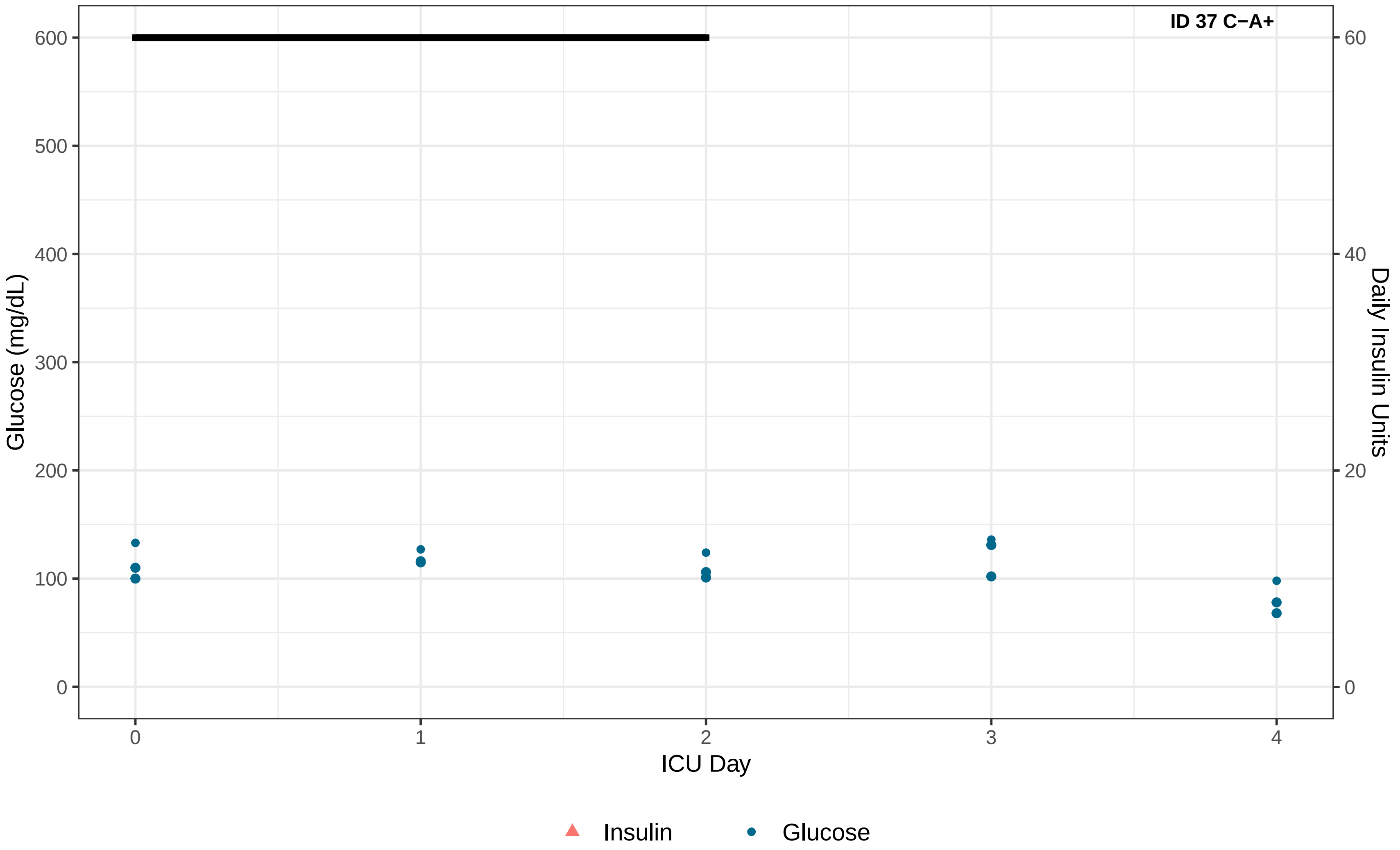

ID 37 C−A+
Glucose (mg/dL)
Daily Insulin Units
ICU Day
0
100
200
300
400
500
600
0
20
40
60
0
1
2
3
4
Insulin
Glucose

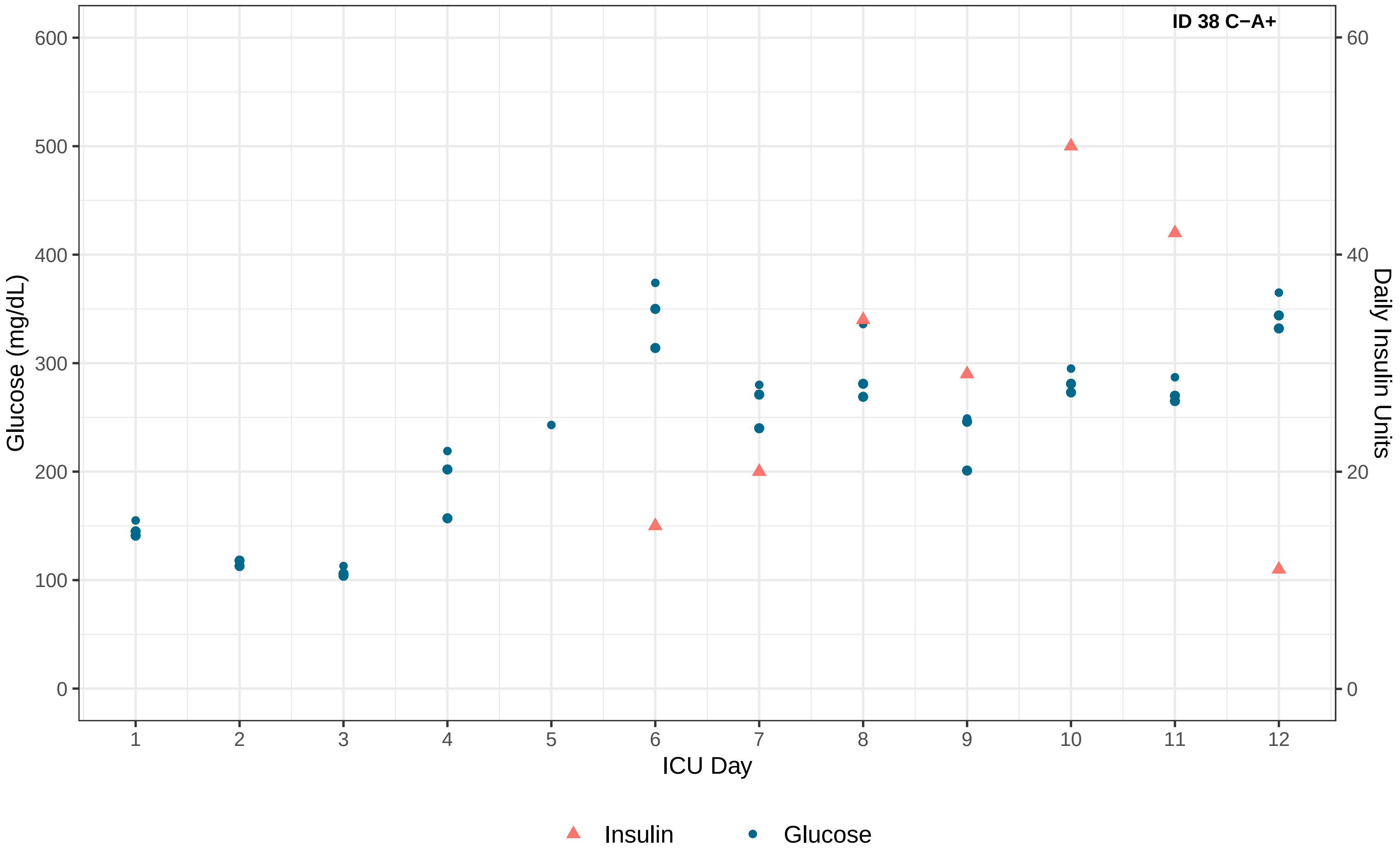

ID 38 C−A+
Glucose (mg/dL)
Daily Insulin Units
ICU Day
0
100
200
300
400
500
600
0
20
40
60
1
2
3
4
5
6
7
8
9
10
11
12
Insulin
Glucose

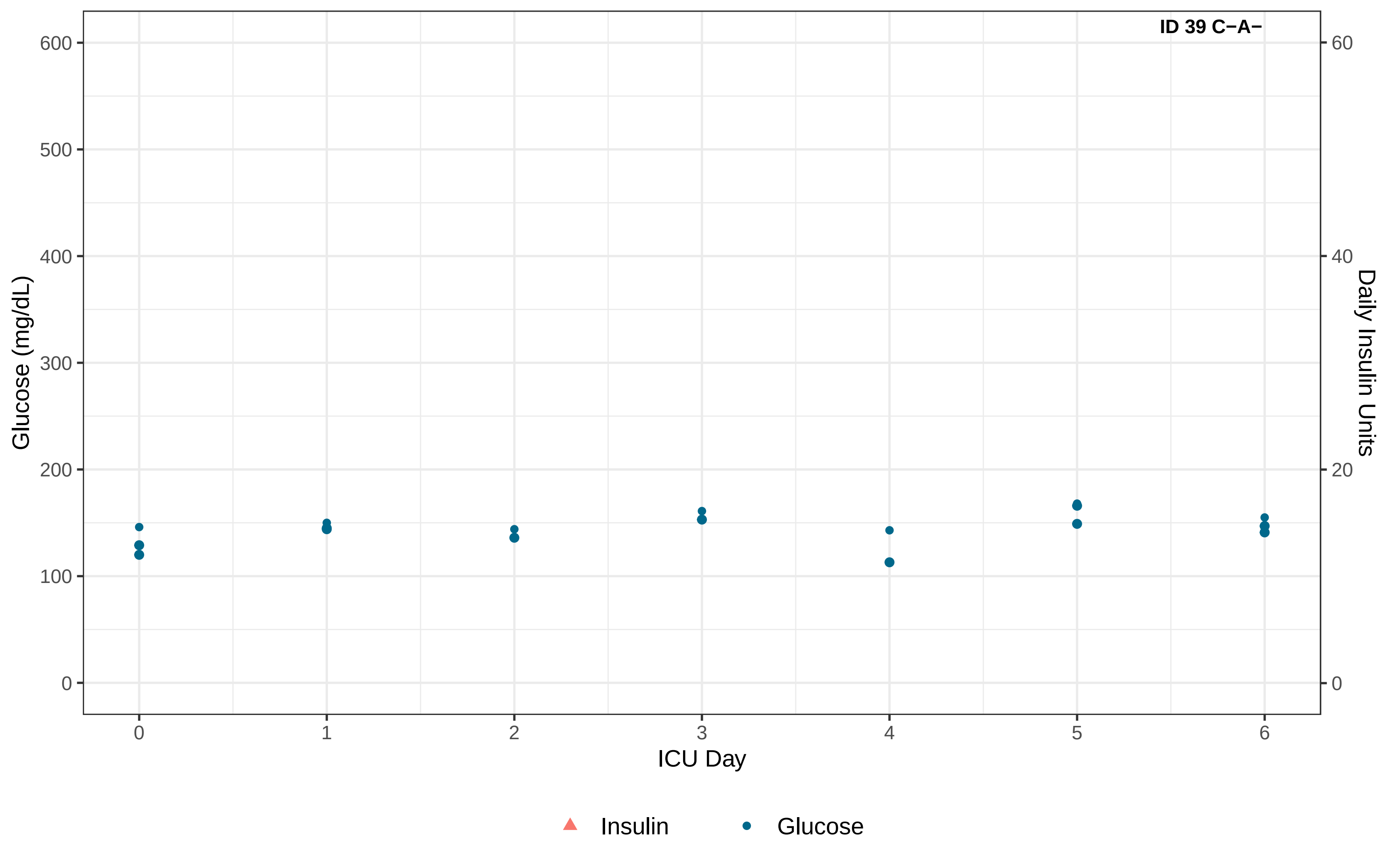
ID 39 C−A−
Glucose (mg/dL)
600
500
400
300
200
100
0
Daily Insulin Units
60
40
20
0
0
1
2
3
4
5
6
ICU Day
Insulin
Glucose

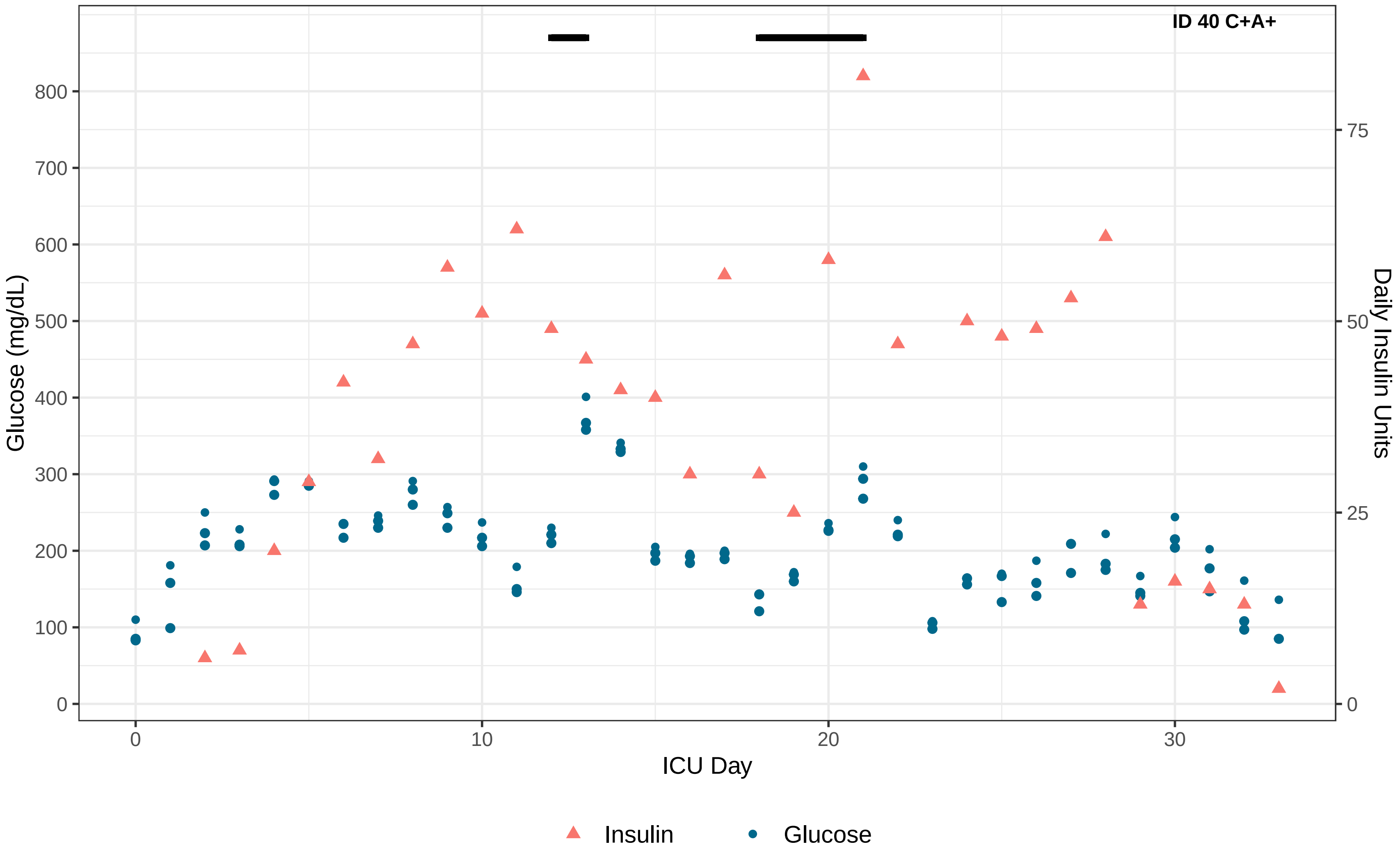
ID 40 C+A+
Glucose (mg/dL)
0
100
200
300
400
500
600
700
800
Daily Insulin Units
0
25
50
75
ICU Day
0
10
20
30
Insulin
Glucose

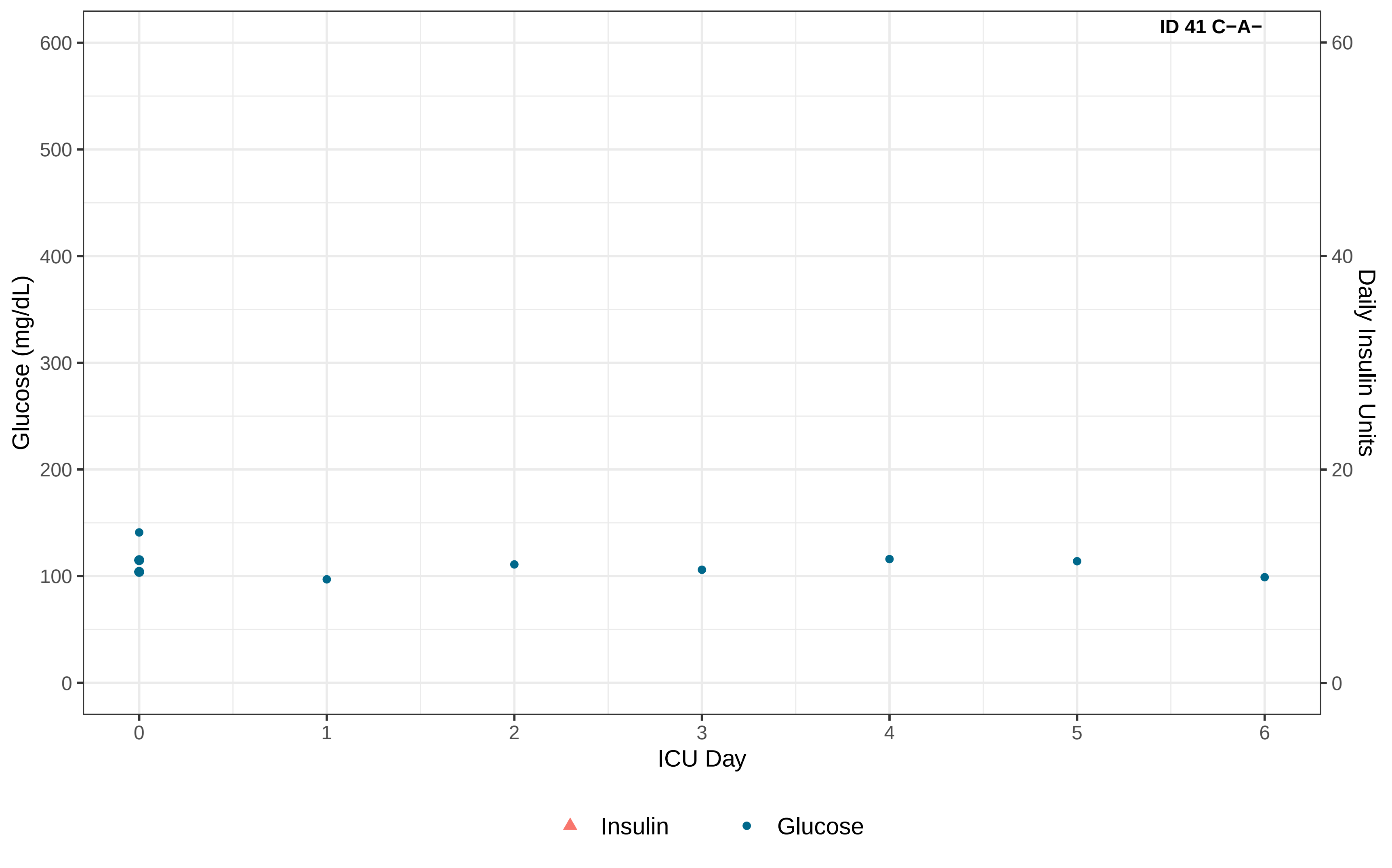
ID 41 C−A−
Glucose (mg/dL)
600
500
400
300
200
100
0
Daily Insulin Units
60
40
20
0
0
1
2
3
4
5
6
ICU Day
Insulin
Glucose

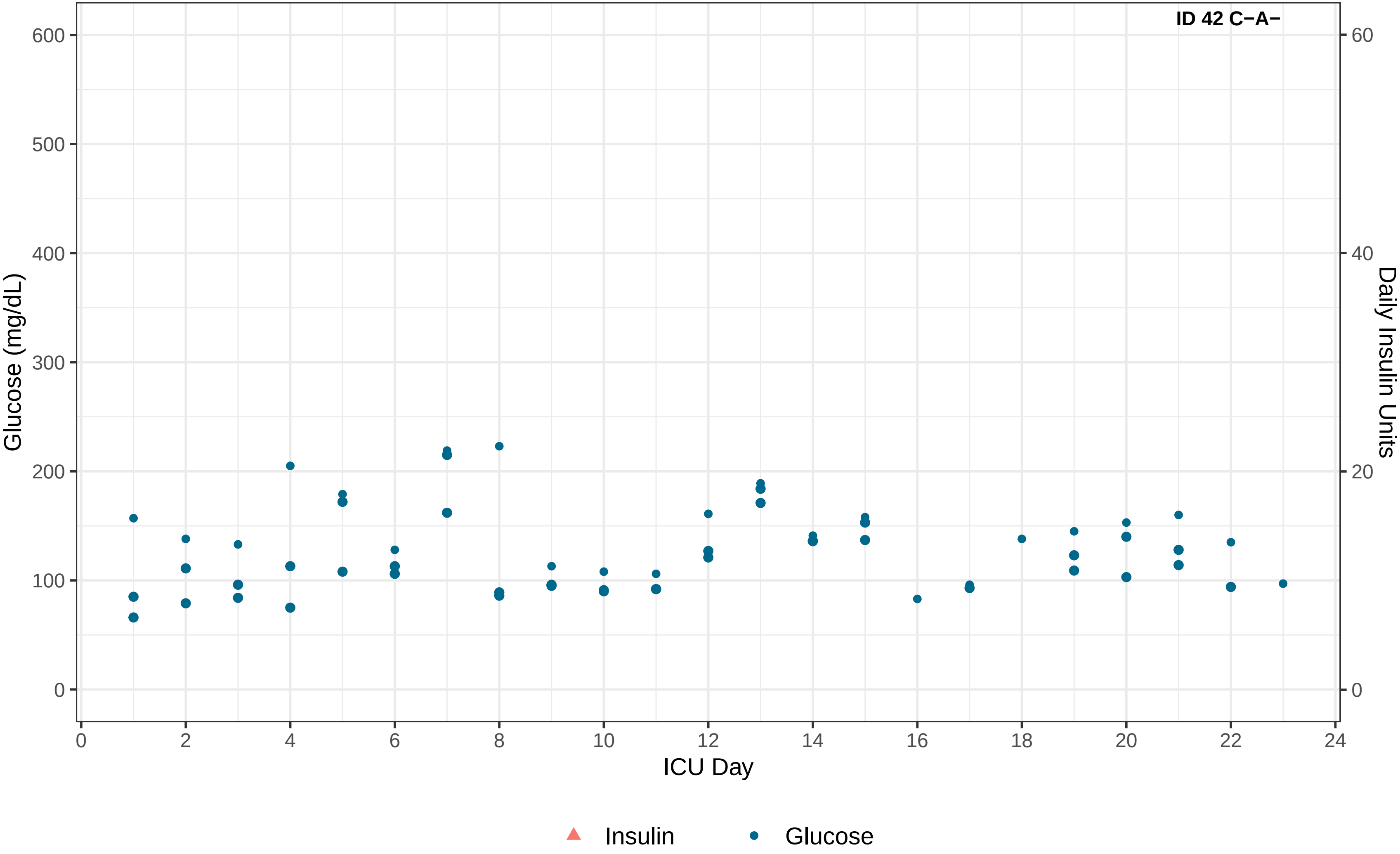
ID 42 C−A−
Glucose (mg/dL)
Daily Insulin Units
ICU Day
0
100
200
300
400
500
600
0
20
40
60
0
2
4
6
8
10
12
14
16
18
20
22
24
Insulin
Glucose

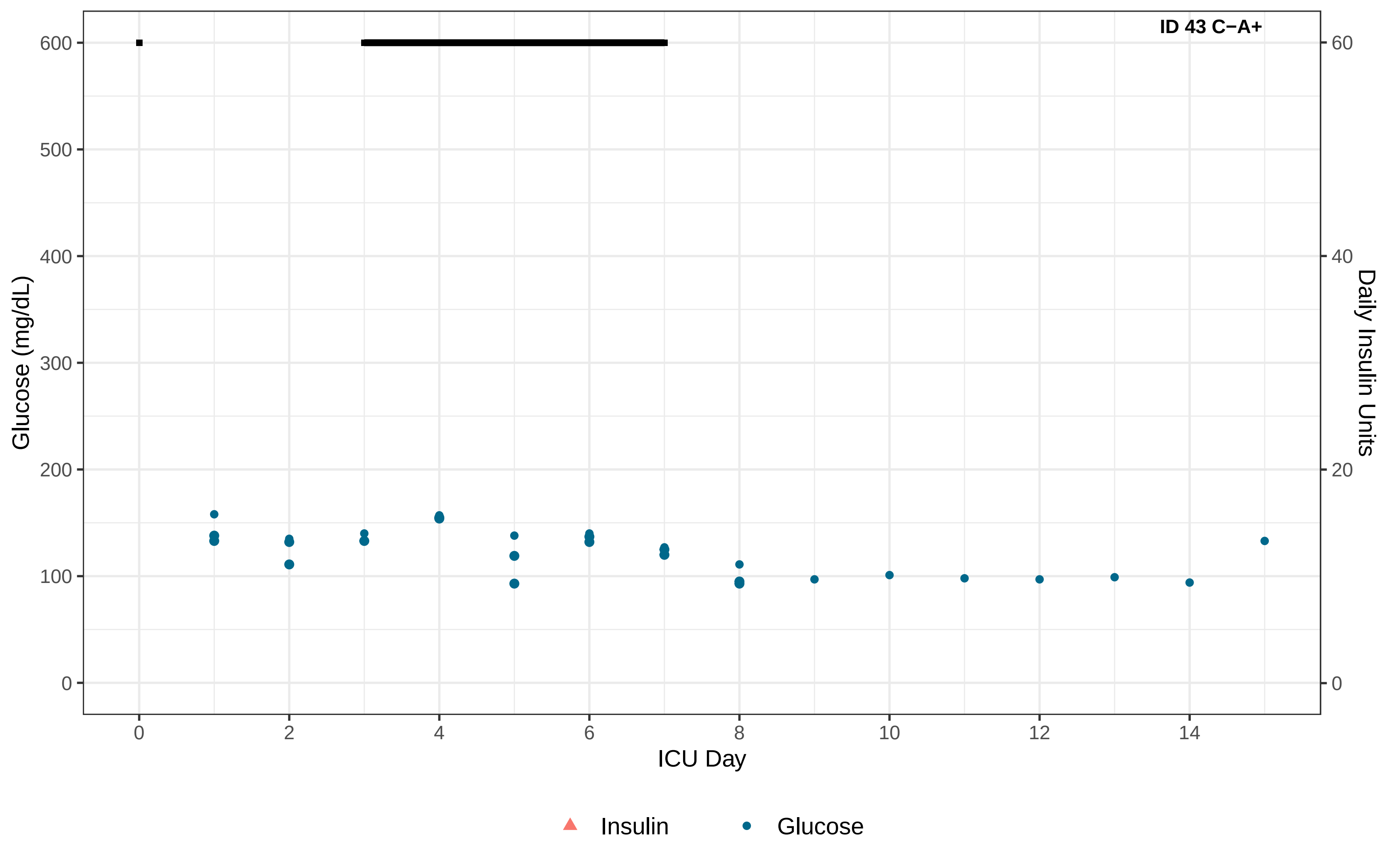
ID 43 C−A+
Glucose (mg/dL)
Daily Insulin Units
ICU Day
0
100
200
300
400
500
600
0
20
40
60
0
2
4
6
8
10
12
14
Insulin
Glucose

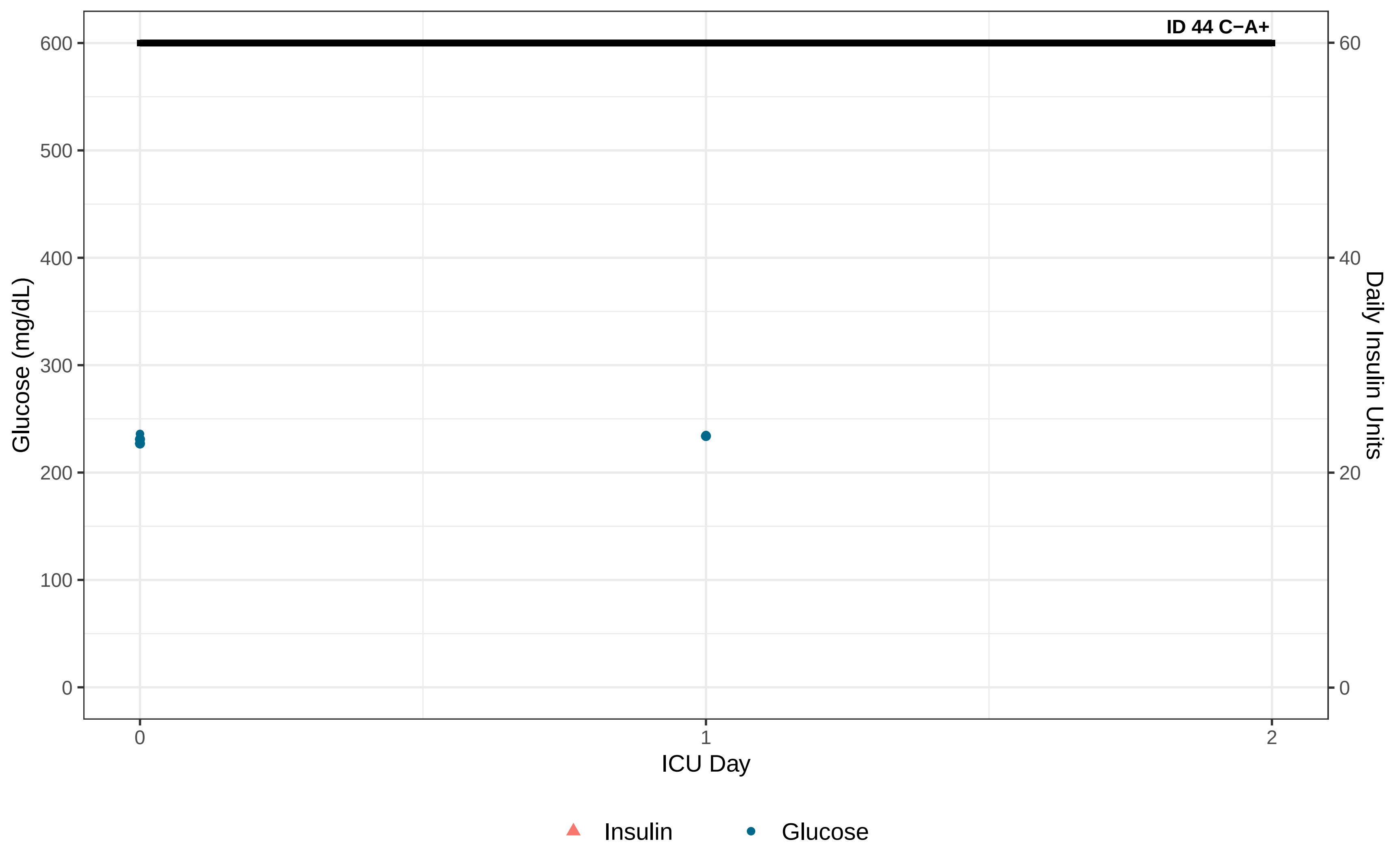

ID 44 C−A+
Glucose (mg/dL)
600
500
400
300
200
100
0
Daily Insulin Units
60
40
20
0
ICU Day
0
1
2
Insulin
Glucose

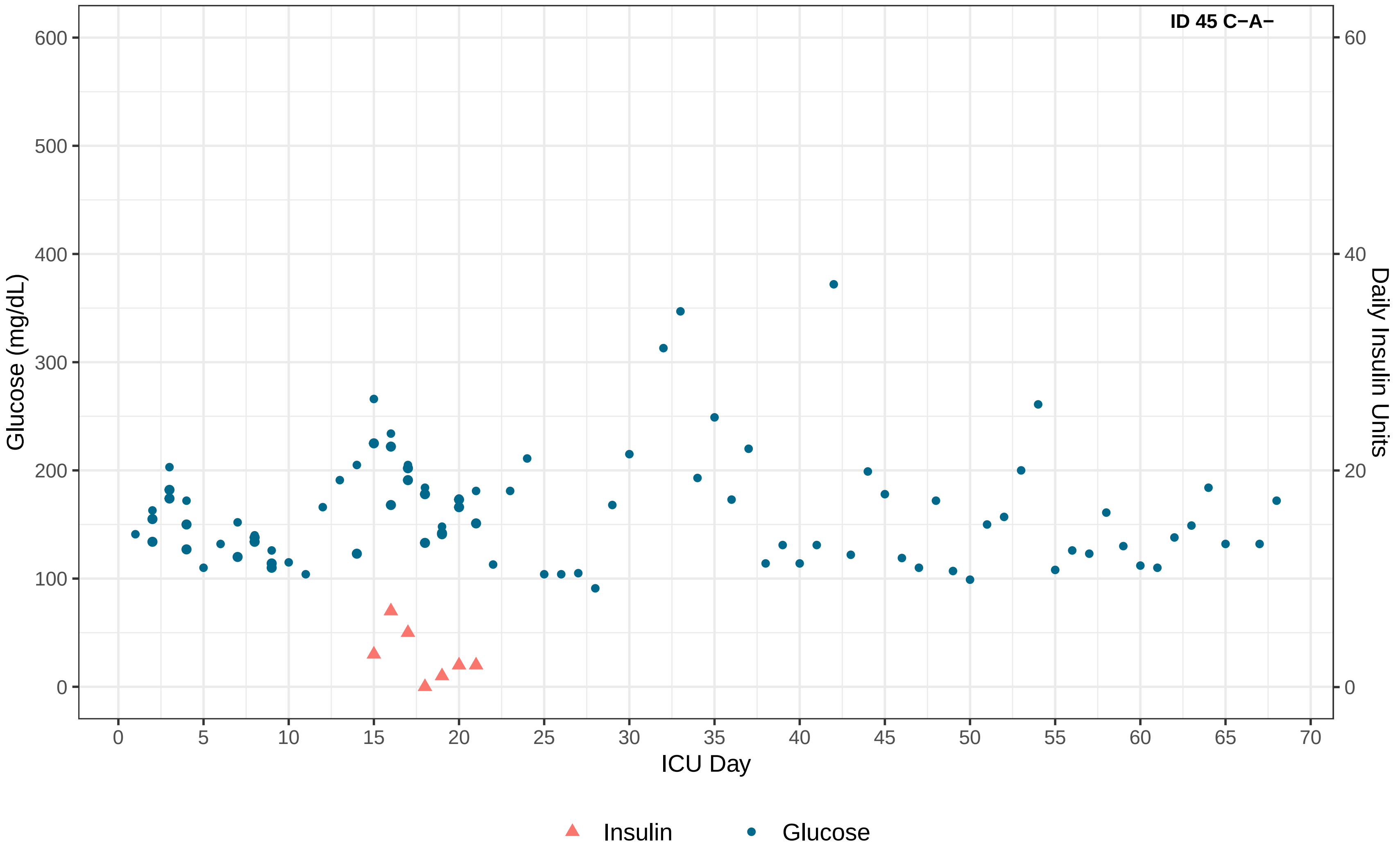

ID 45 C−A−
Glucose (mg/dL)
Daily Insulin Units
ICU Day
0
100
200
300
400
500
600
0
20
40
60
0
5
10
15
20
25
30
35
40
45
50
55
60
65
70
Insulin
Glucose

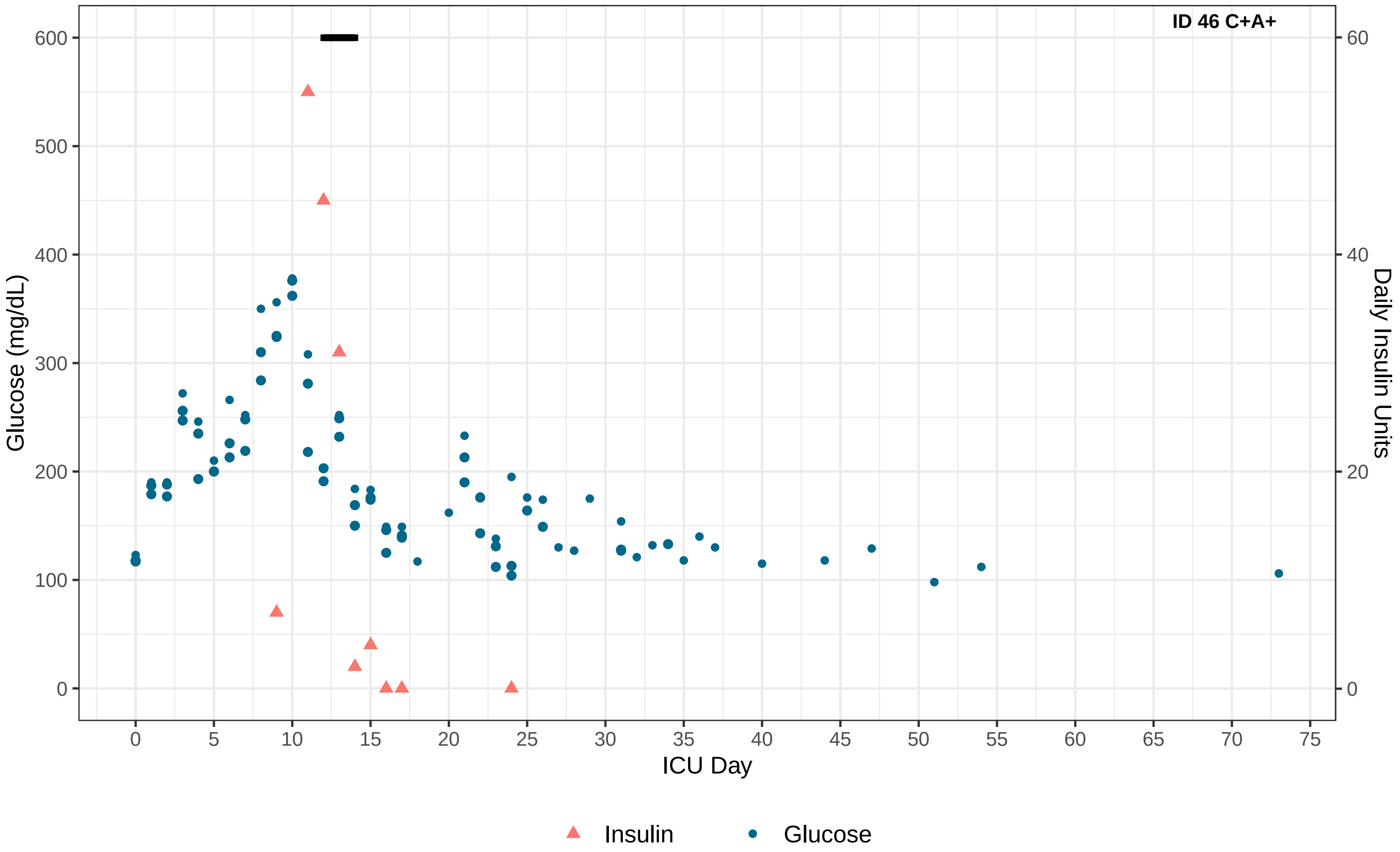
ID 46 C+A+
Glucose (mg/dL)
Daily Insulin Units
ICU Day
0
100
200
300
400
500
600
0
20
40
60
0
5
10
15
20
25
30
35
40
45
50
55
60
65
70
75
Insulin
Glucose

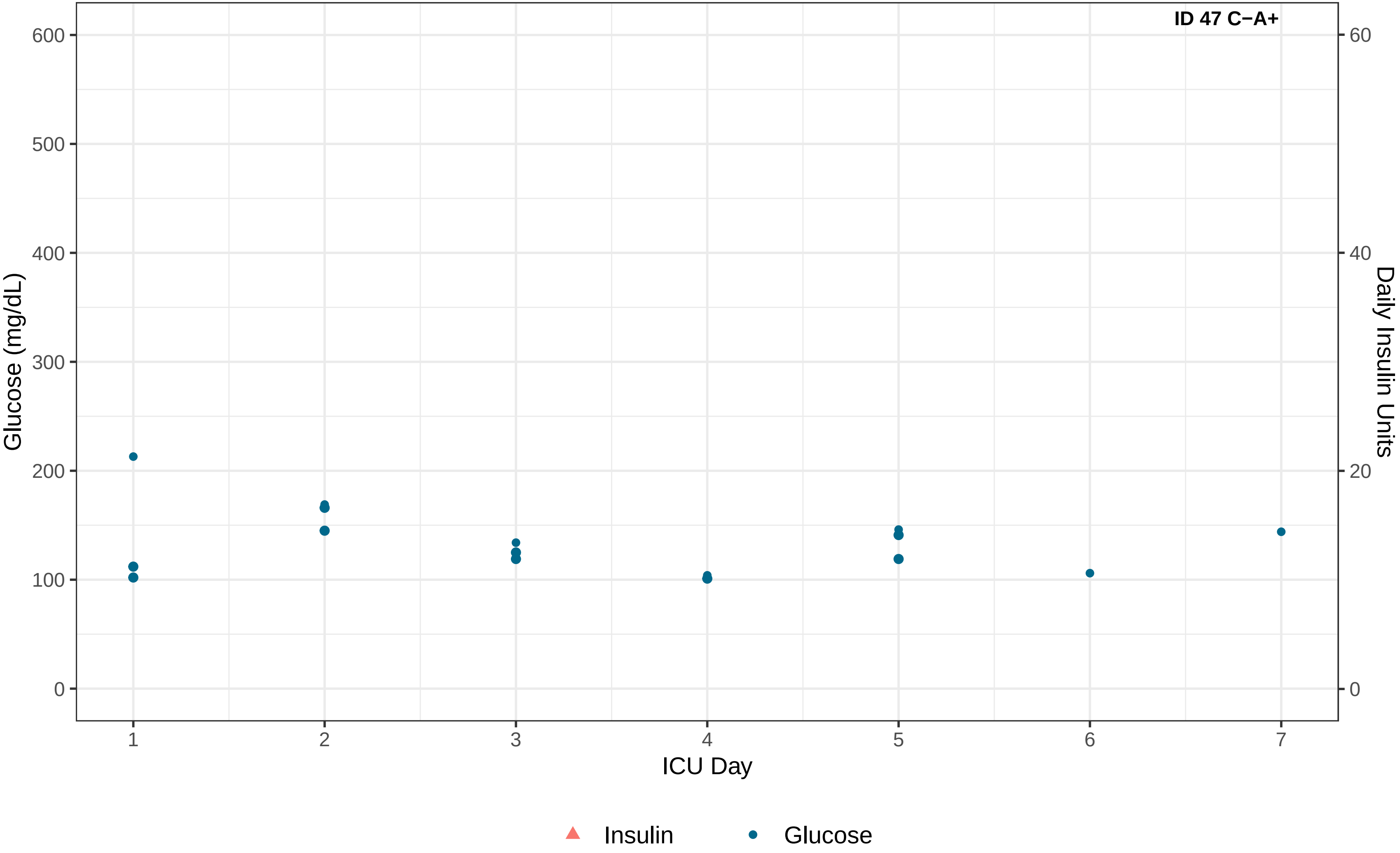
ID 47 C−A+
Glucose (mg/dL)
600
500
400
300
200
100
0
Daily Insulin Units
60
40
20
0
1
2
3
4
5
6
7
ICU Day
Insulin
Glucose

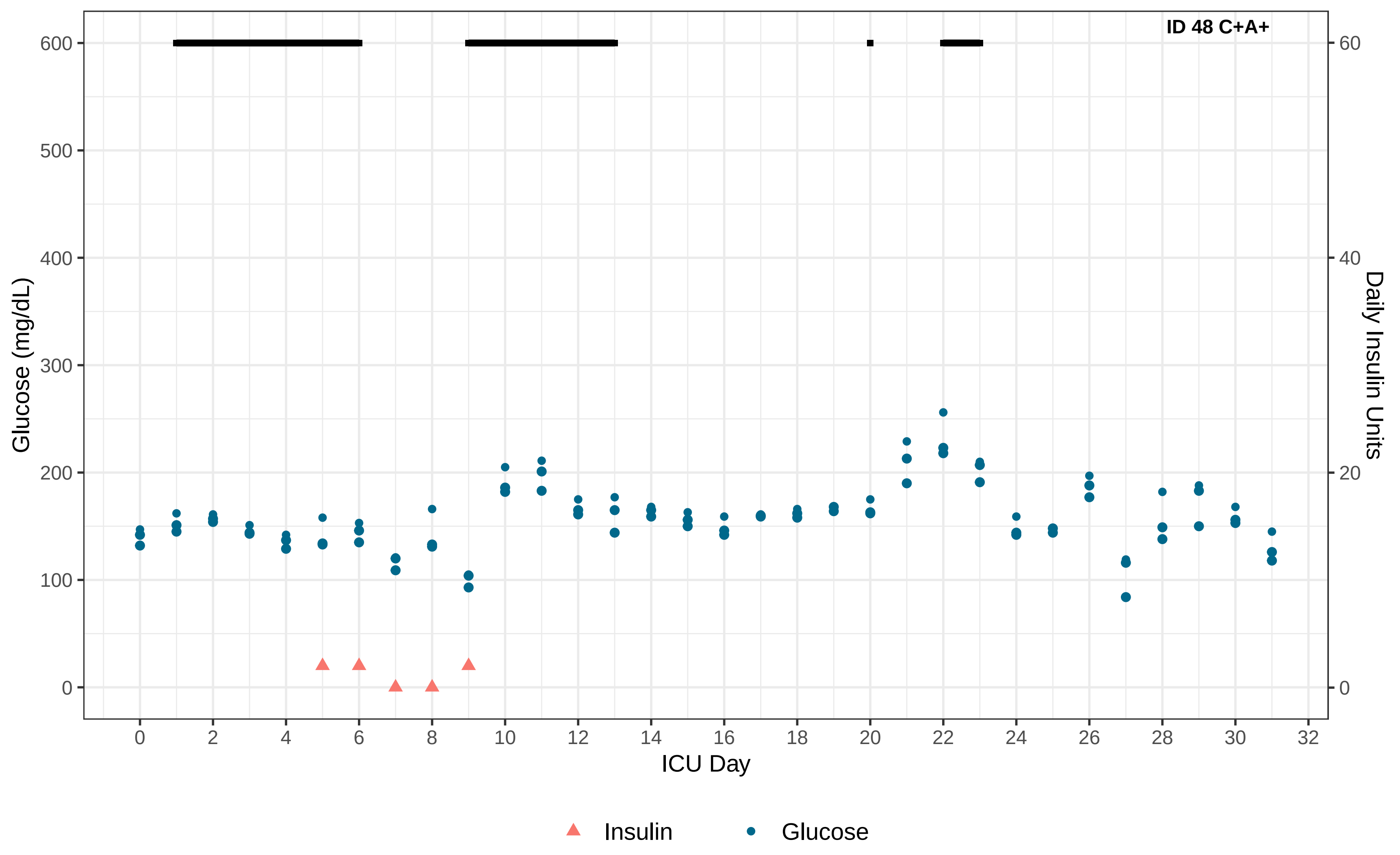

ID 48 C+A+
Glucose (mg/dL)
Daily Insulin Units
ICU Day
600
500
400
300
200
100
0
60
40
20
0
0 2 4 6 8 10 12 14 16 18 20 22 24 26 28 30 32
Insulin
Glucose

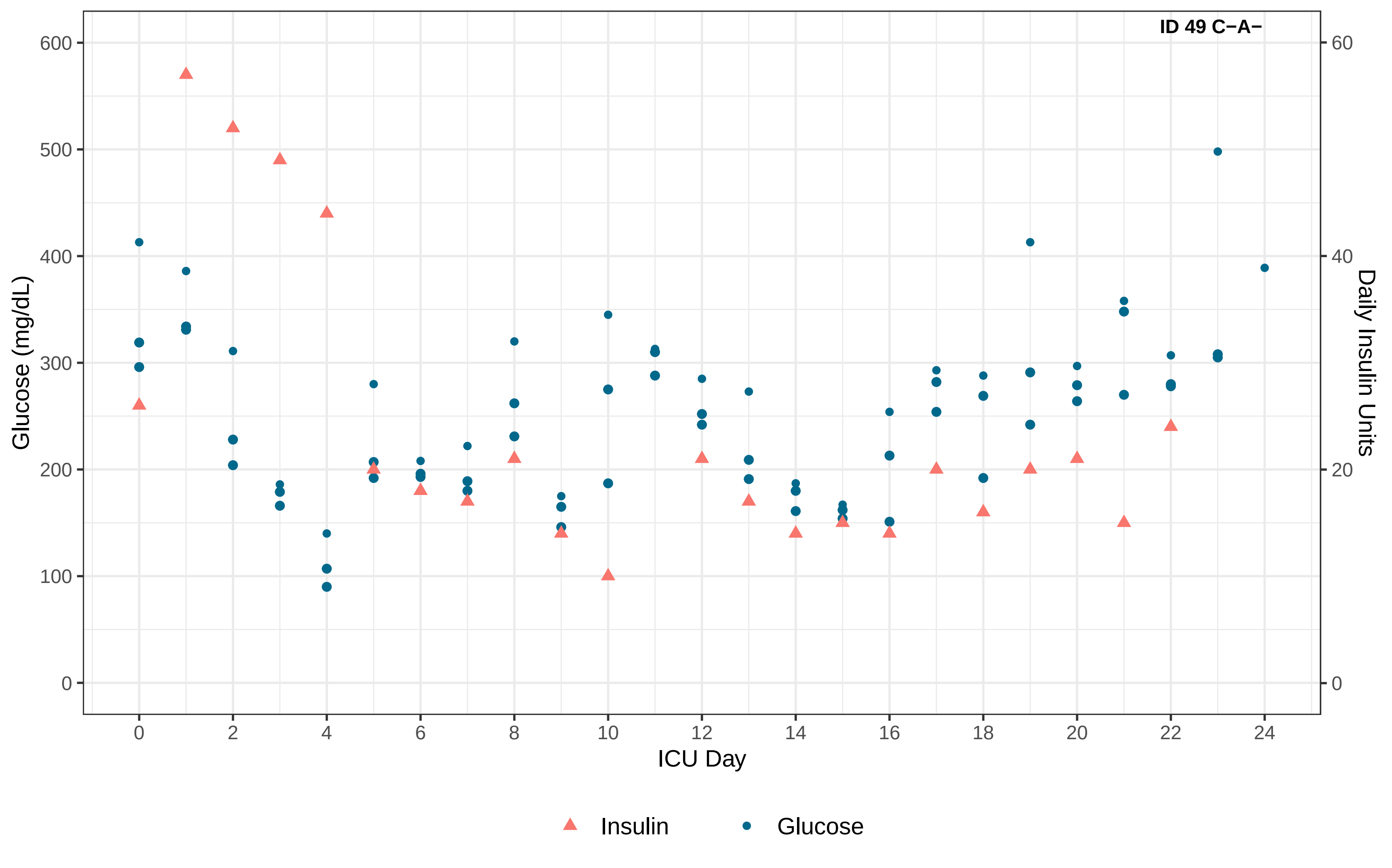

ID 49 C−A−
Glucose (mg/dL)
Daily Insulin Units
ICU Day
0
100
200
300
400
500
600
0
20
40
60
0
2
4
6
8
10
12
14
16
18
20
22
24
Insulin
Glucose

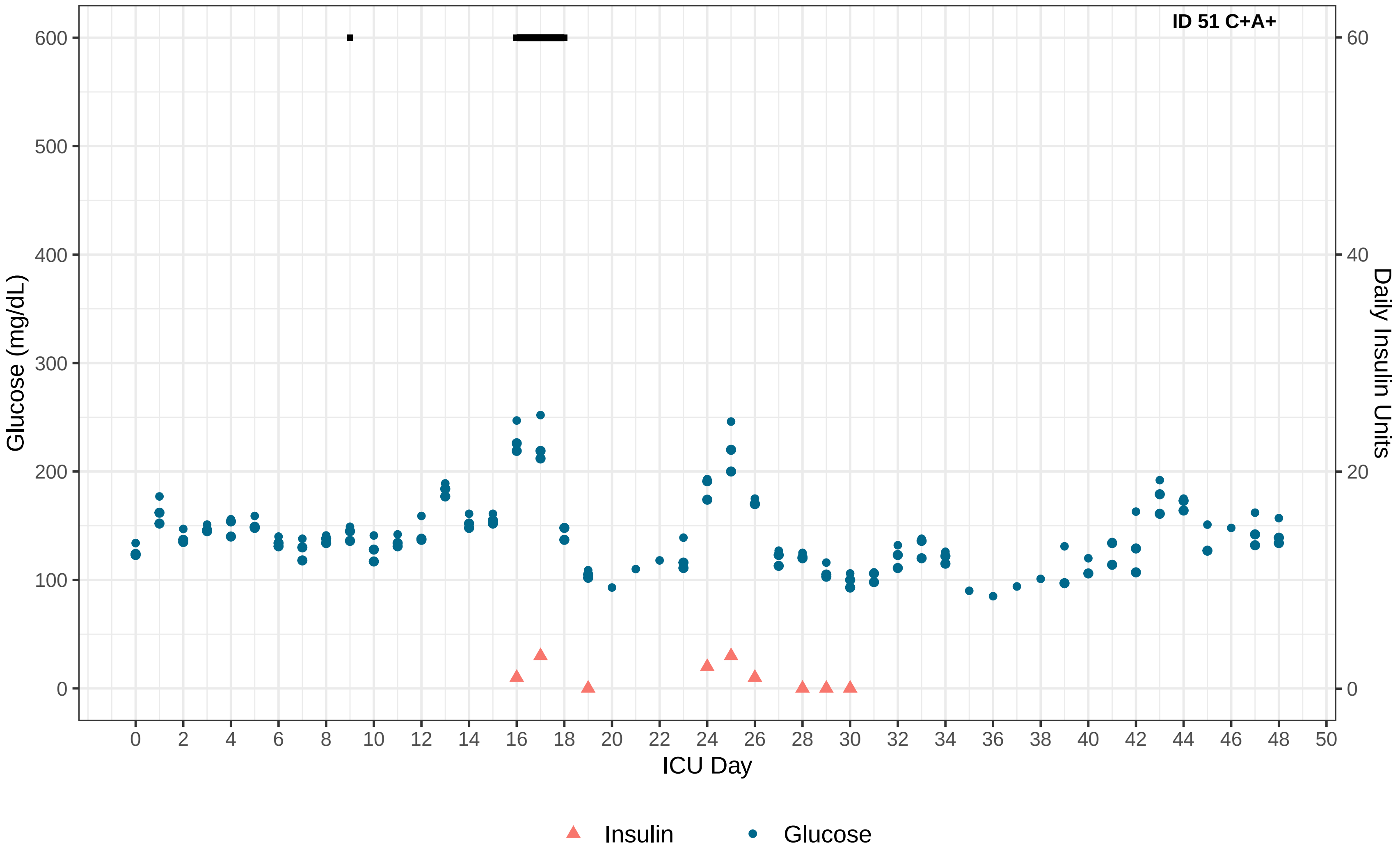
ID 51 C+A+
Glucose (mg/dL)
Daily Insulin Units
ICU Day
0
100
200
300
400
500
600
0
20
40
60
0 2 4 6 8 10 12 14 16 18 20 22 24 26 28 30 32 34 36 38 40 42 44 46 48 50
Insulin
Glucose

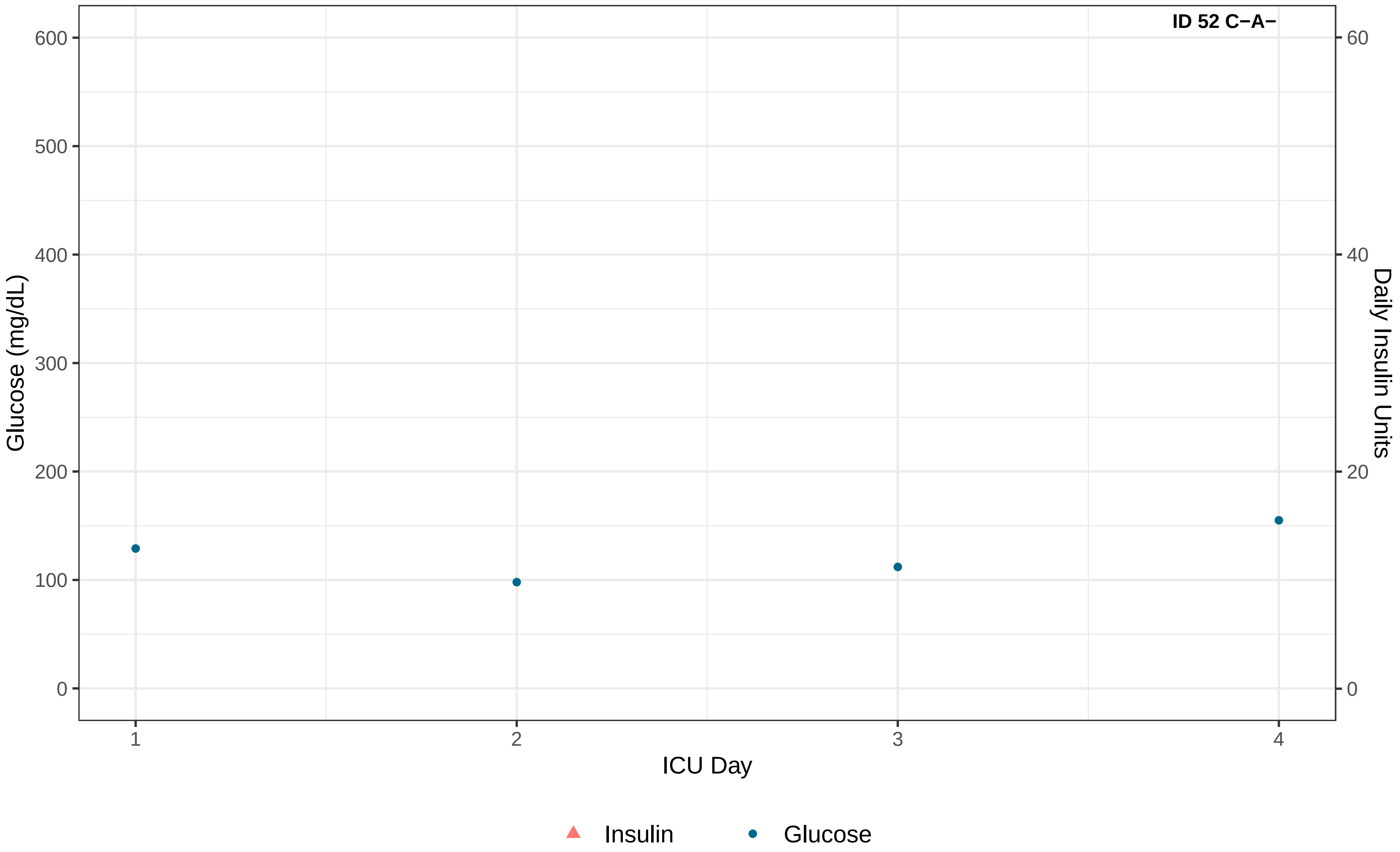
ID 52 C−A−
Glucose (mg/dL)
600
500
400
300
200
100
0
Daily Insulin Units
60
40
20
0
1
2
3
4
ICU Day
Insulin
Glucose

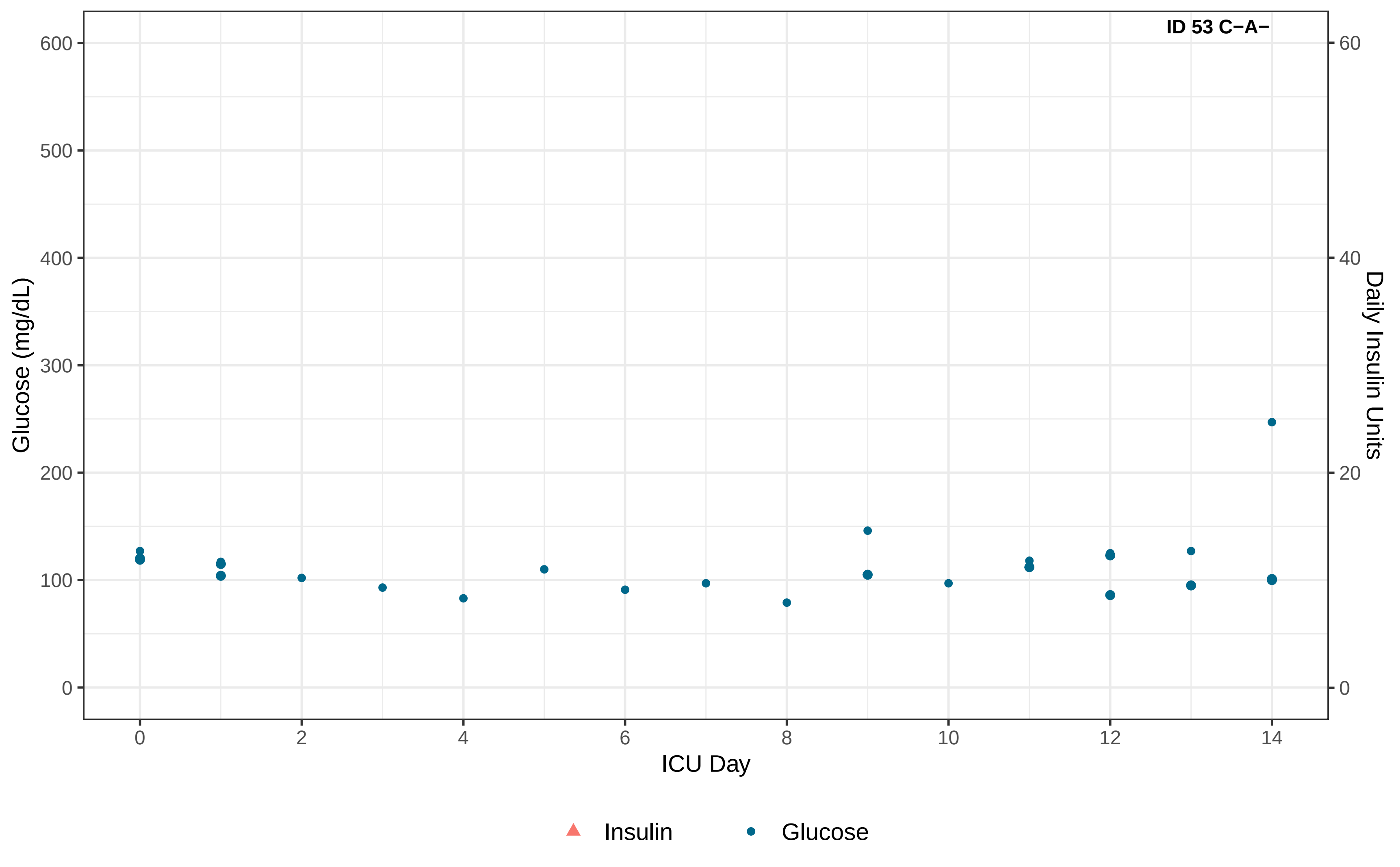

ID 53 C−A−
Glucose (mg/dL)
600
500
400
300
200
100
0
Daily Insulin Units
60
40
20
0
0
2
4
6
8
10
12
14
ICU Day
Insulin
Glucose

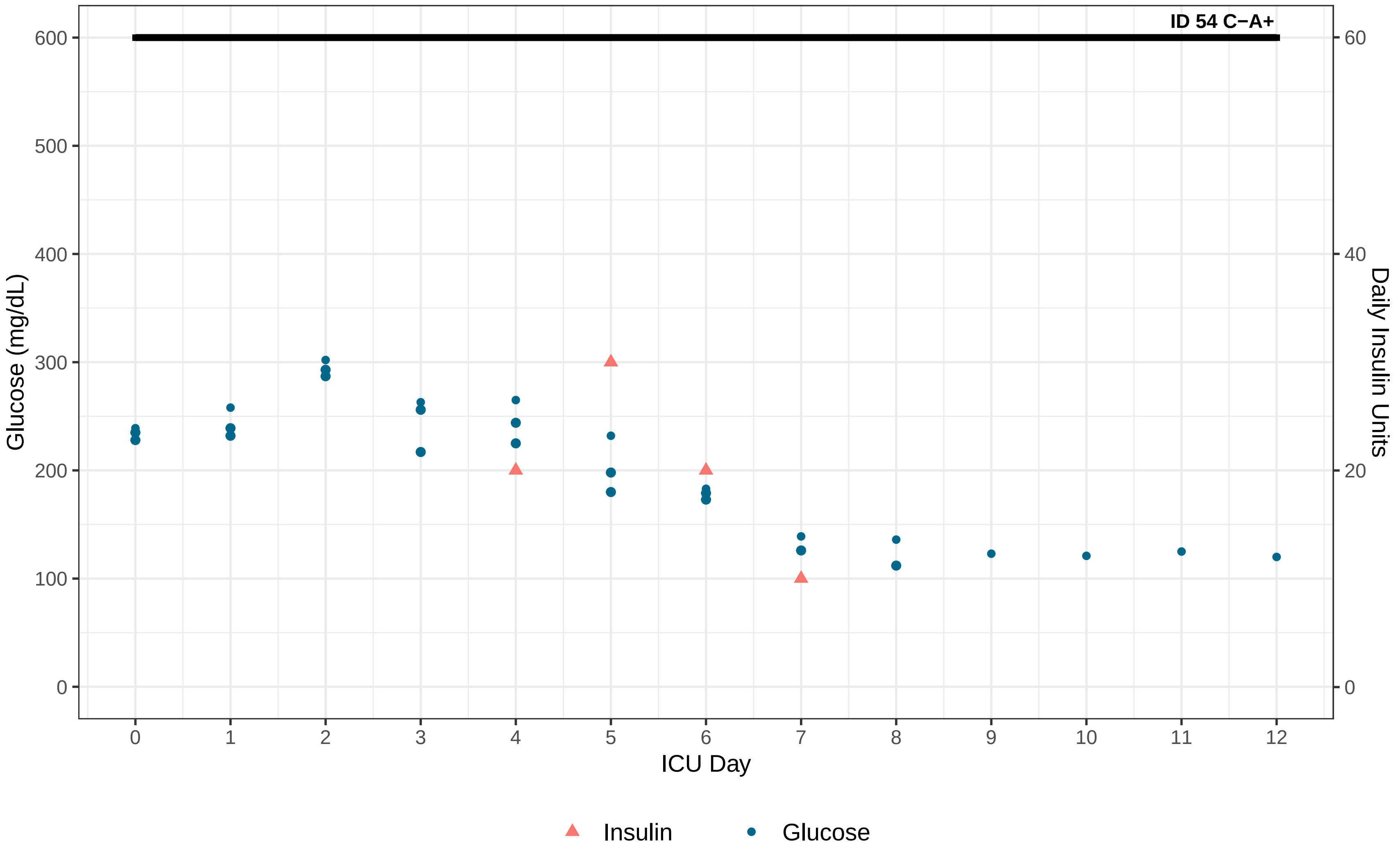

ID 54 C−A+
Glucose (mg/dL)
Daily Insulin Units
ICU Day
0
100
200
300
400
500
600
0
20
40
60
0
1
2
3
4
5
6
7
8
9
10
11
12
Insulin
Glucose

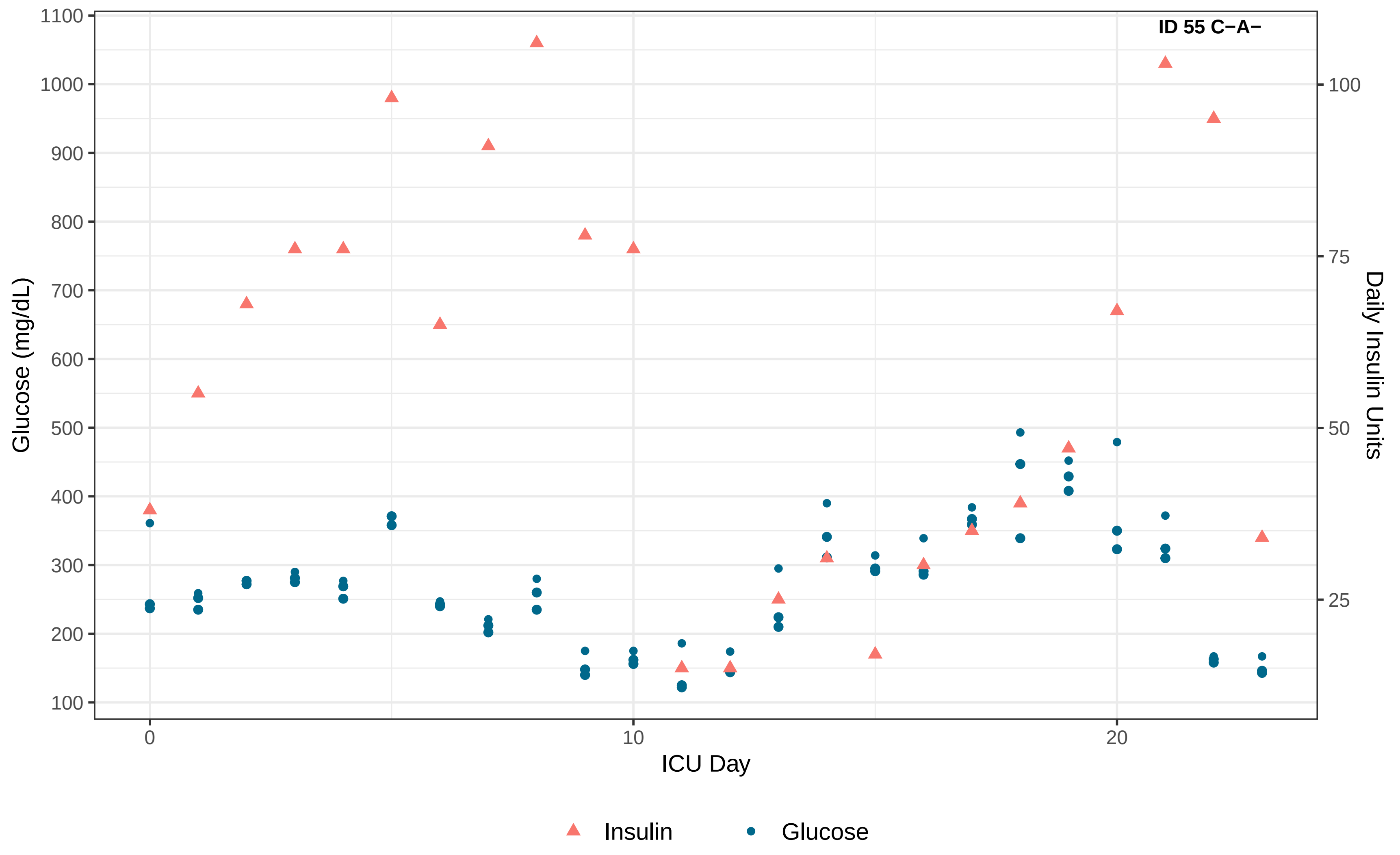
ID 55 C−A−
Glucose (mg/dL)
Daily Insulin Units
ICU Day
1100
1000
900
800
700
600
500
400
300
200
100
100
75
50
25
0
10
20
Insulin
Glucose

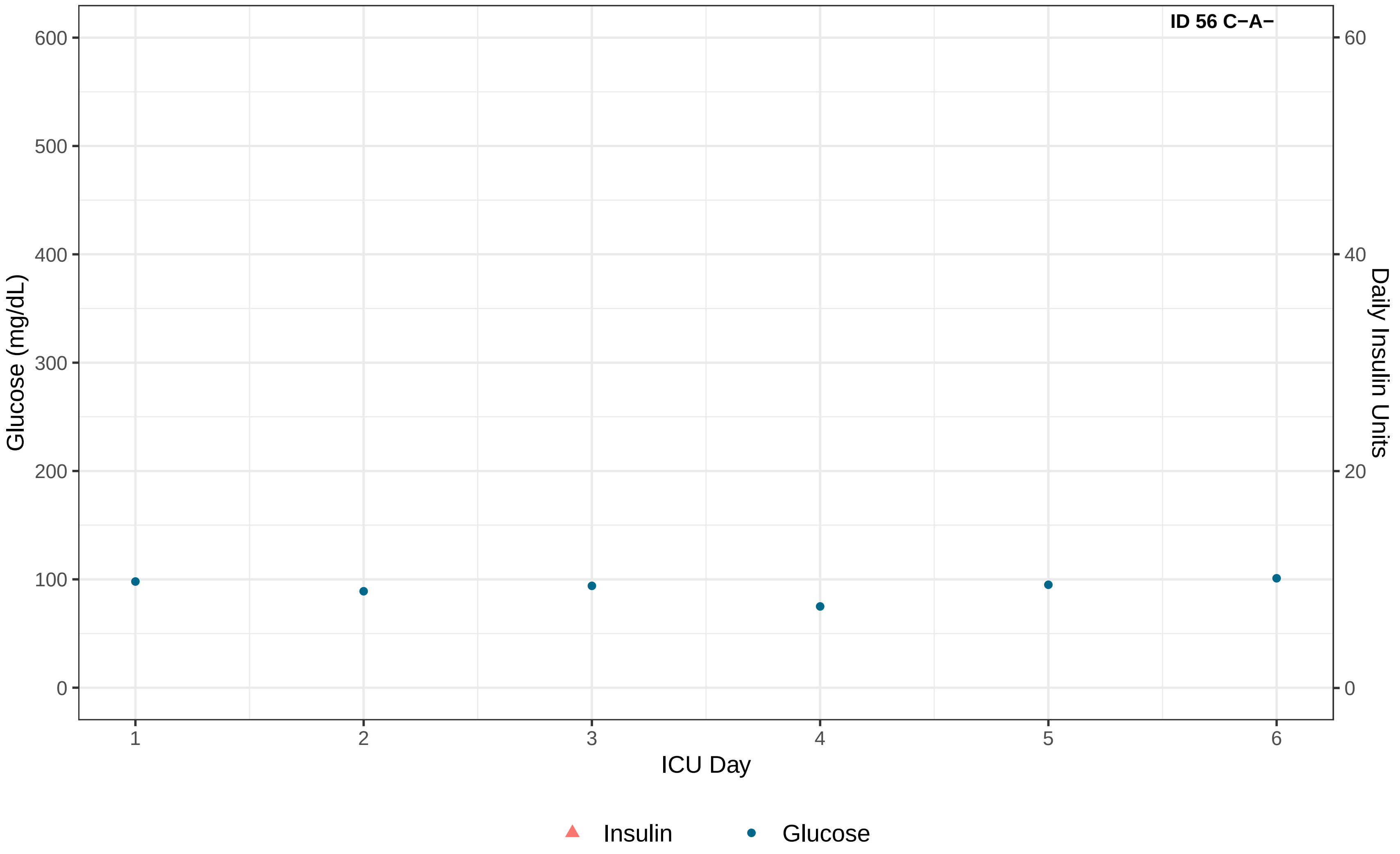
ID 56 C−A−
Glucose (mg/dL)
600
500
400
300
200
100
0
Daily Insulin Units
60
40
20
0
1
2
3
4
5
6
ICU Day
Insulin
Glucose

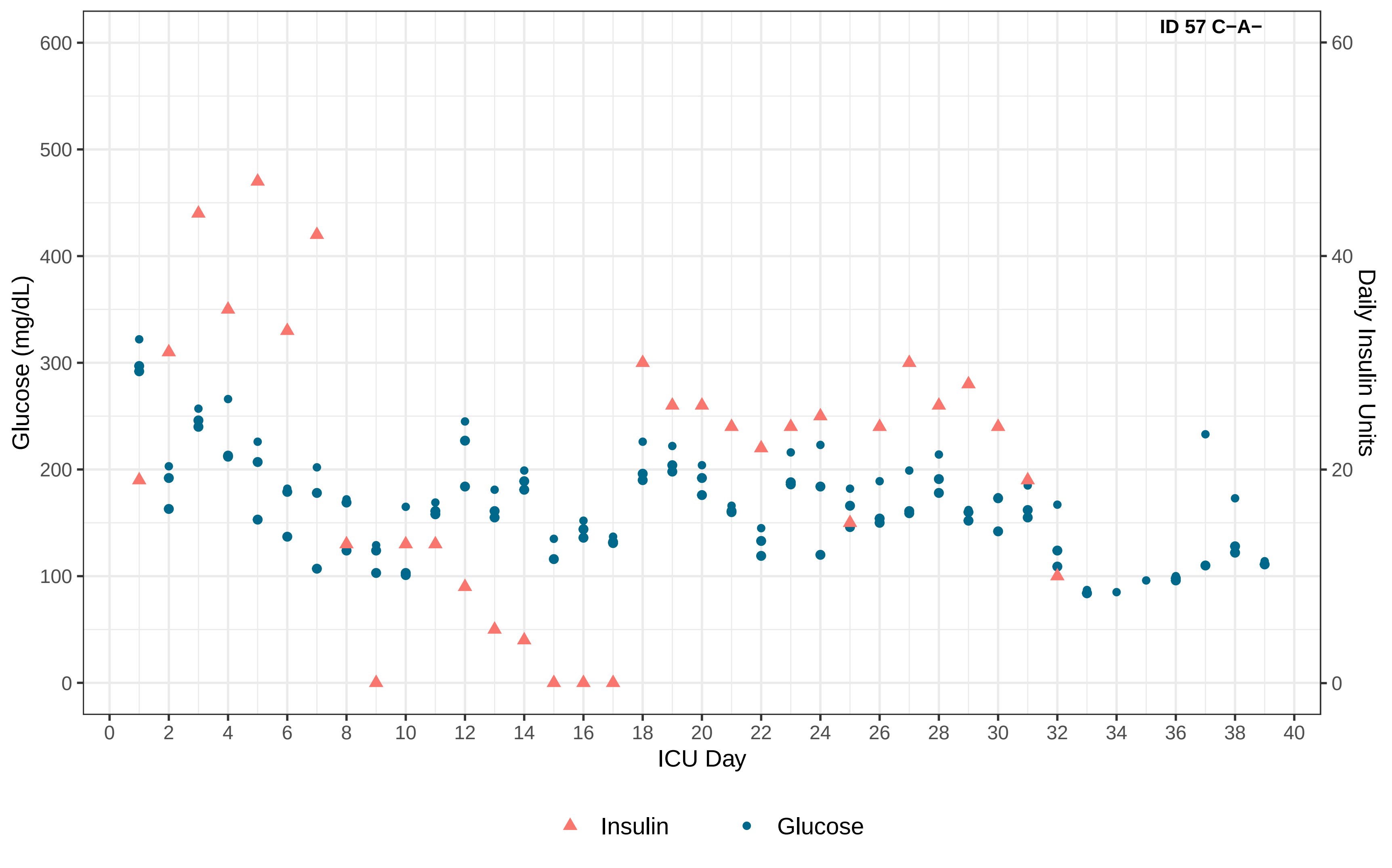

ID 57 C−A−
Glucose (mg/dL)
Daily Insulin Units
ICU Day
0
100
200
300
400
500
600
0
20
40
60
0 2 4 6 8 10 12 14 16 18 20 22 24 26 28 30 32 34 36 38 40
Insulin
Glucose

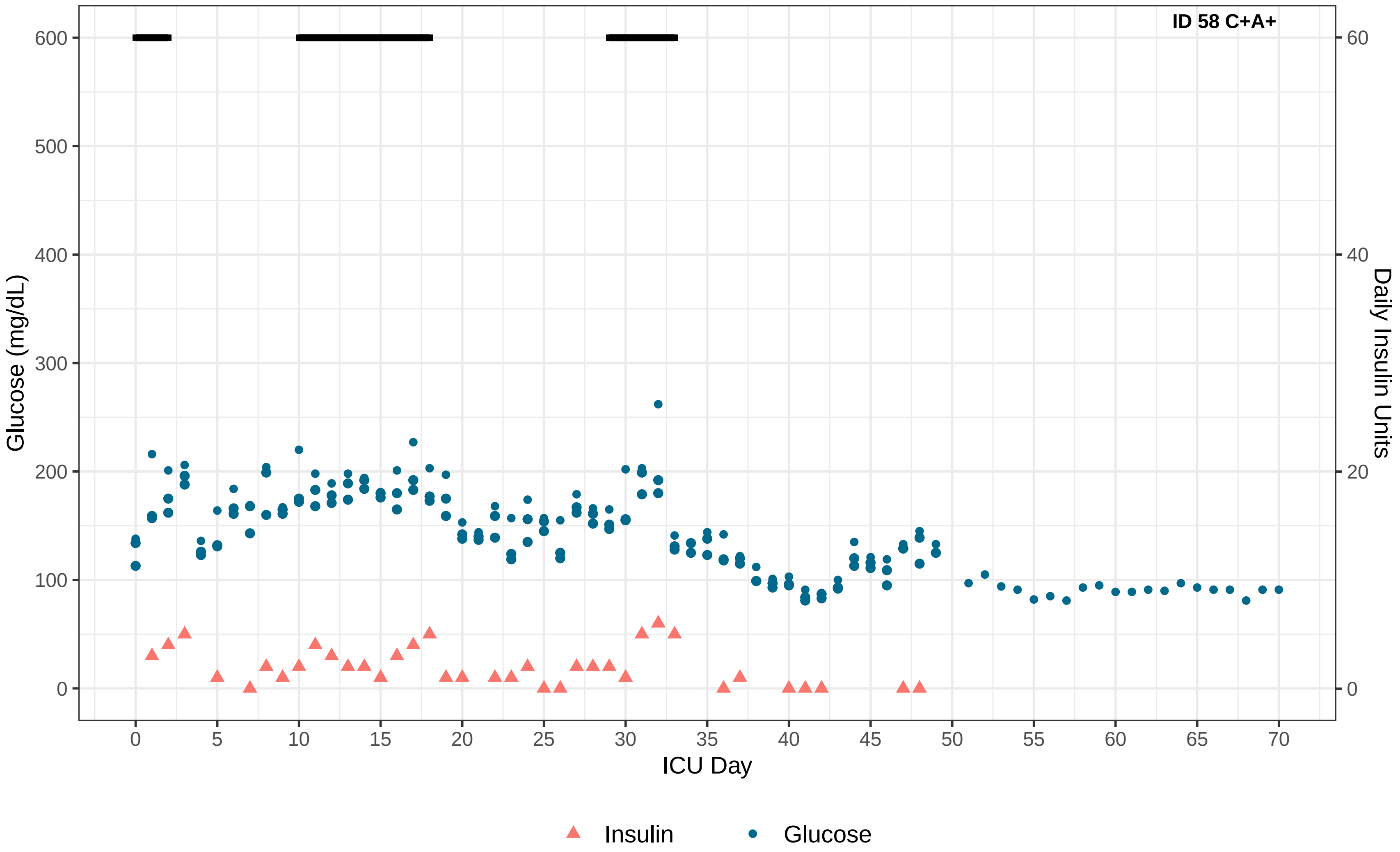
ID 58 C+A+
Glucose (mg/dL)
Daily Insulin Units
ICU Day
0
100
200
300
400
500
600
0
20
40
60
0
5
10
15
20
25
30
35
40
45
50
55
60
65
70
Insulin
Glucose

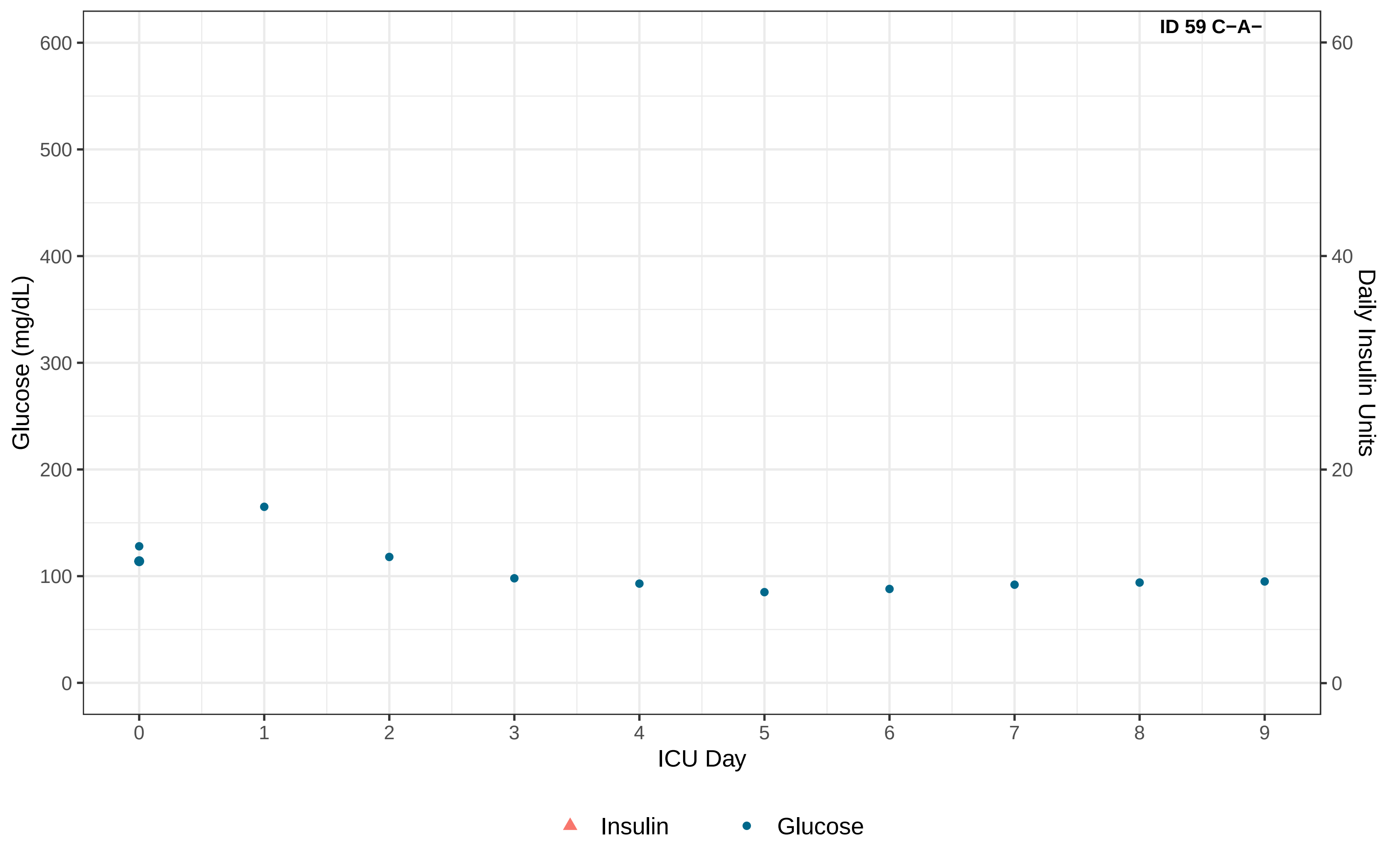

ID 59 C−A−
Glucose (mg/dL)
600
500
400
300
200
100
0
Daily Insulin Units
60
40
20
0
0
1
2
3
4
5
6
7
8
9
ICU Day
Insulin
Glucose

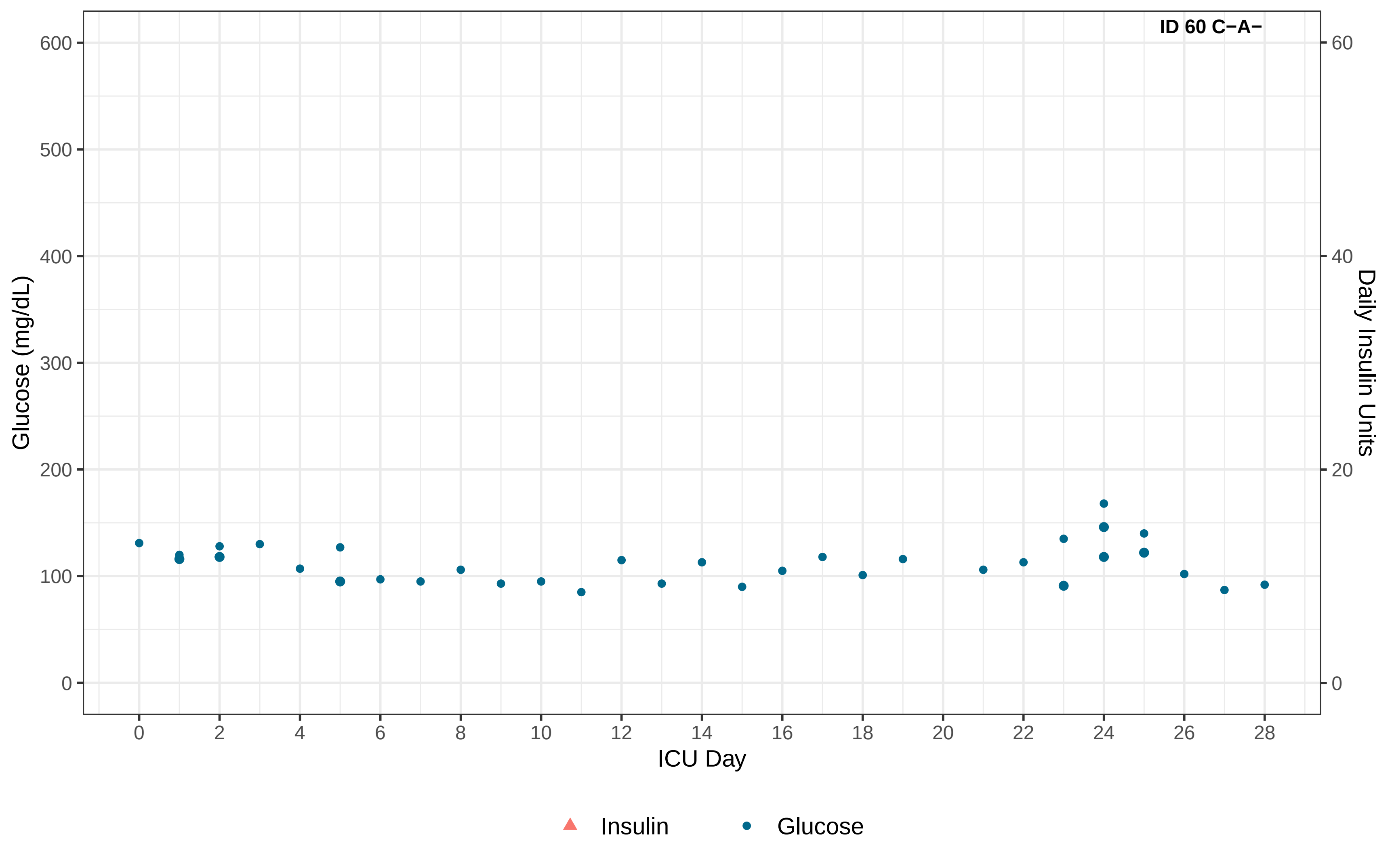

ID 60 C−A−
Glucose (mg/dL)
Daily Insulin Units
ICU Day
0
100
200
300
400
500
600
0
20
40
60
0
2
4
6
8
10
12
14
16
18
20
22
24
26
28
Insulin
Glucose

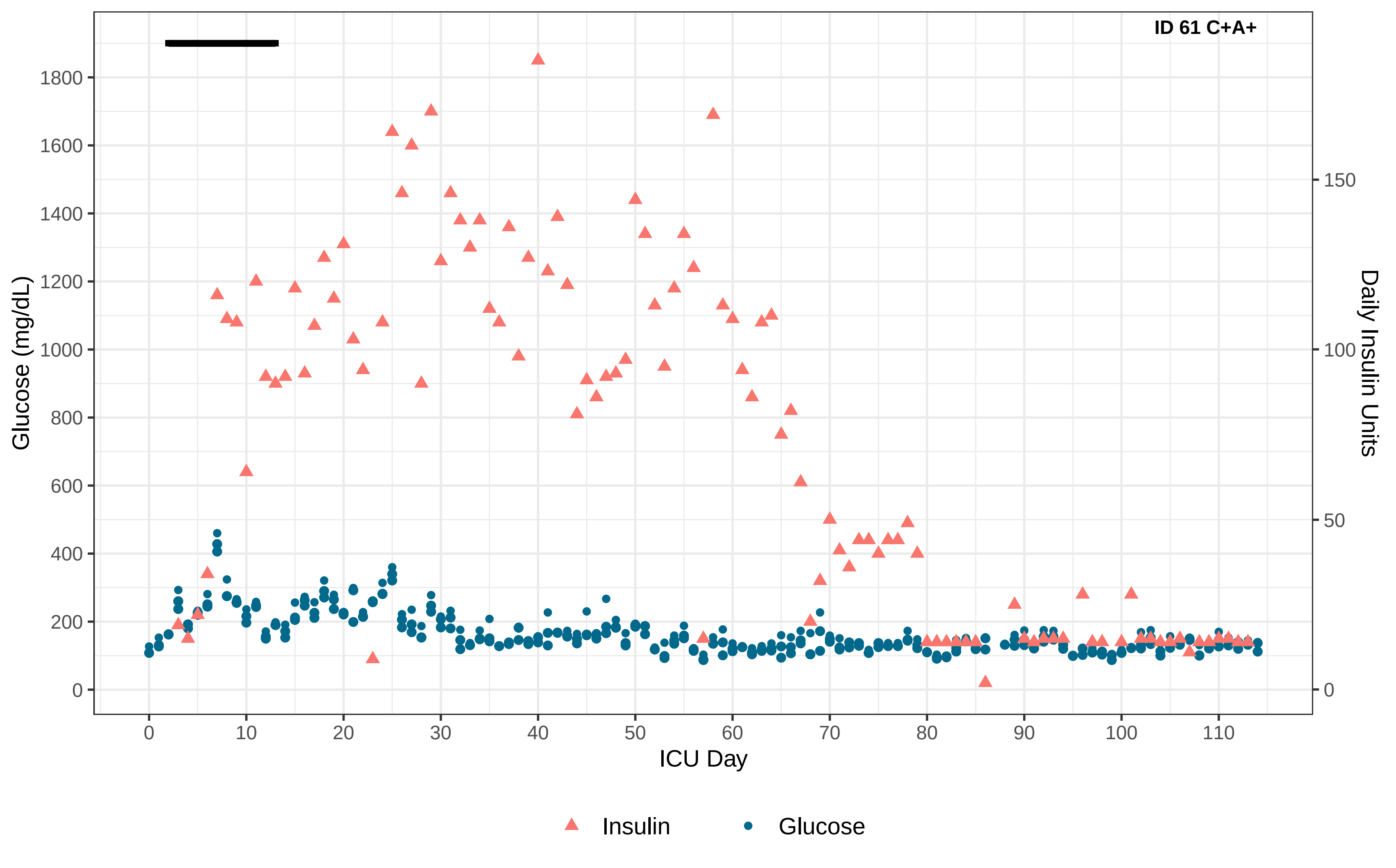

ID 61 C+A+
Glucose (mg/dL)
Daily Insulin Units
ICU Day
Insulin
Glucose

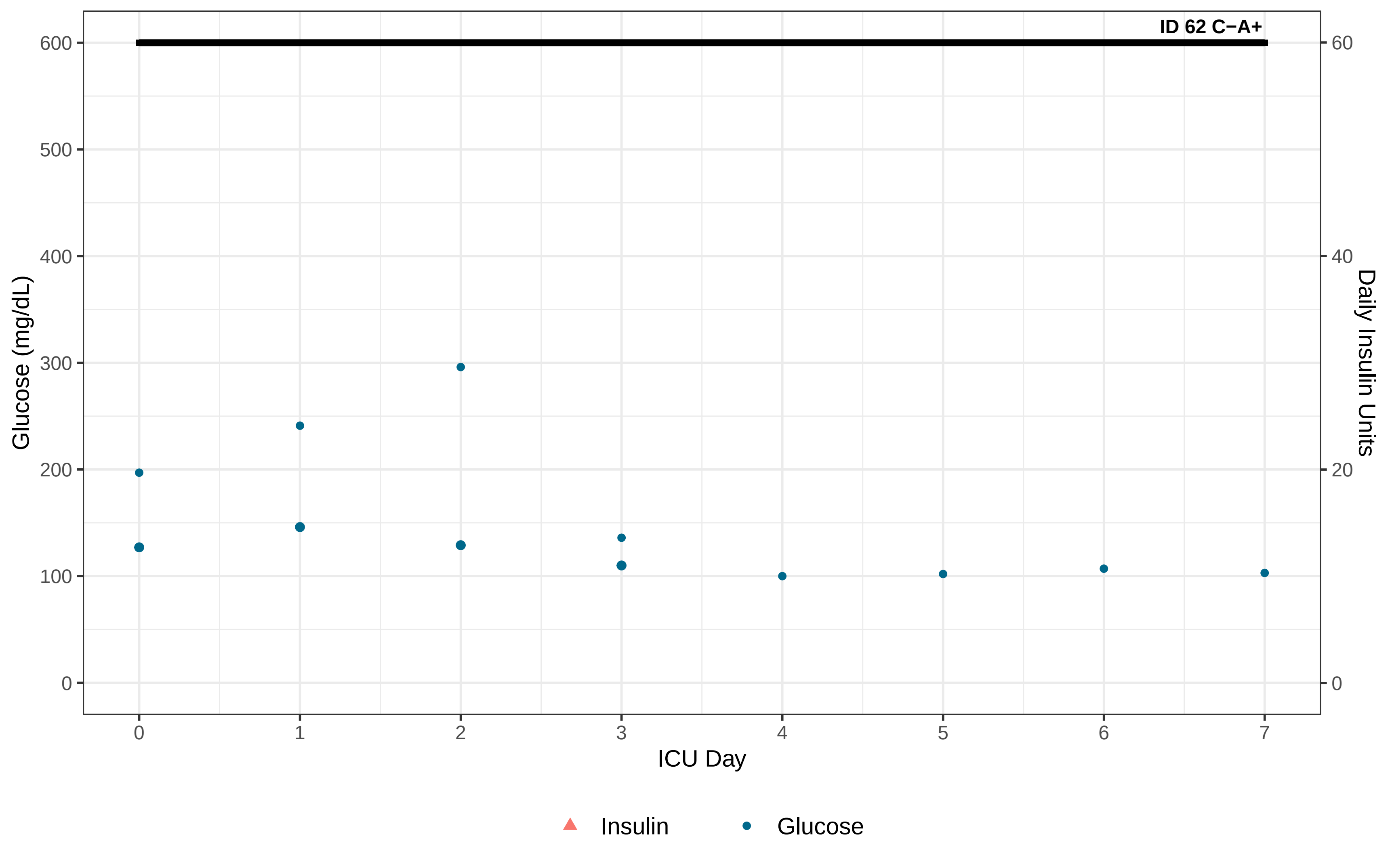

ID 62 C−A+
Glucose (mg/dL)
Daily Insulin Units
0
100
200
300
400
500
600
0
20
40
60
0
1
2
3
4
5
6
7
ICU Day
Insulin
Glucose

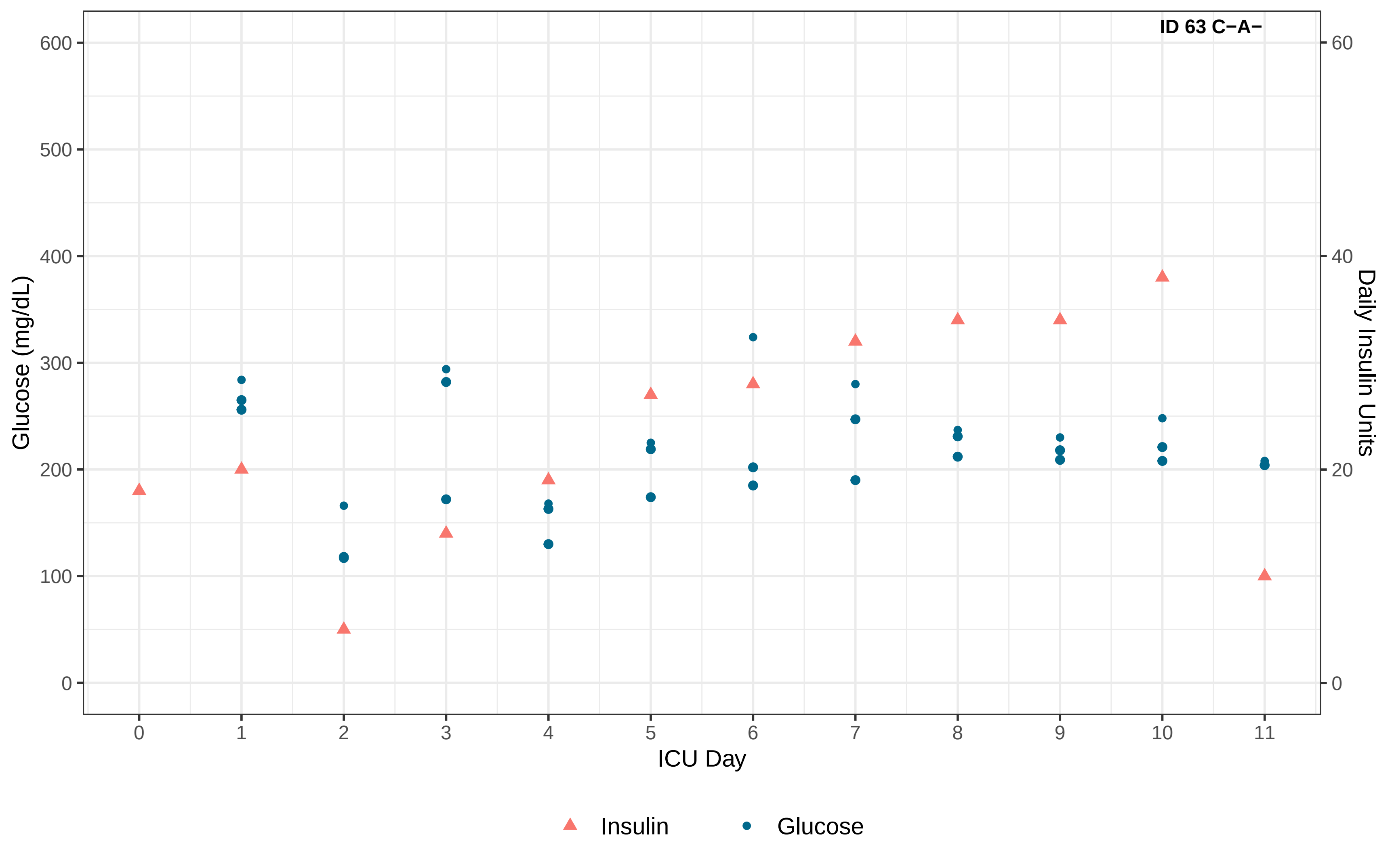
ID 63 C−A−
Glucose (mg/dL)
Daily Insulin Units
ICU Day
0
100
200
300
400
500
600
0
20
40
60
0
1
2
3
4
5
6
7
8
9
10
11
Insulin
Glucose

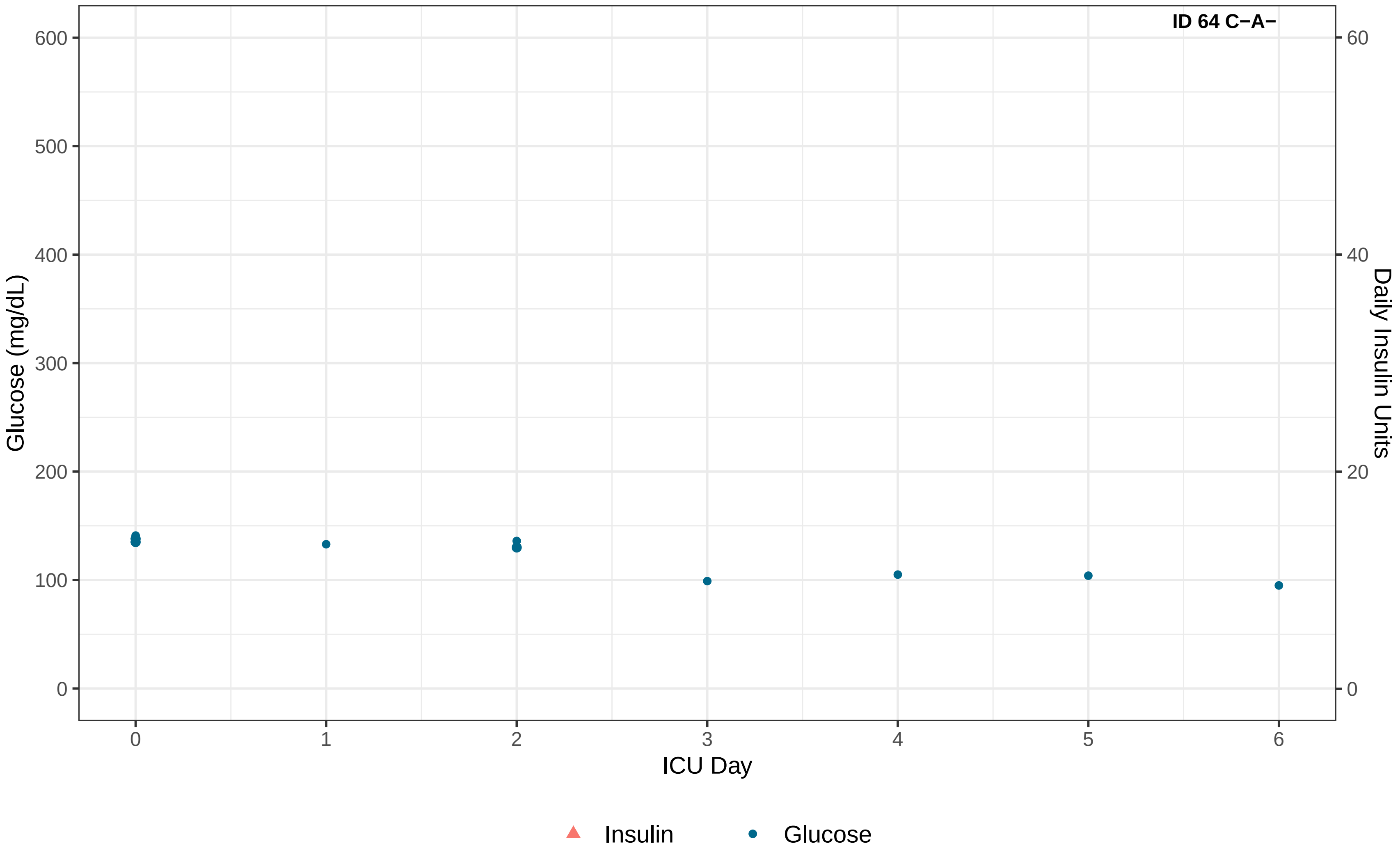

ID 64 C−A−
Glucose (mg/dL)
Daily Insulin Units
ICU Day
0
100
200
300
400
500
600
0
20
40
60
0
1
2
3
4
5
6
Insulin
Glucose

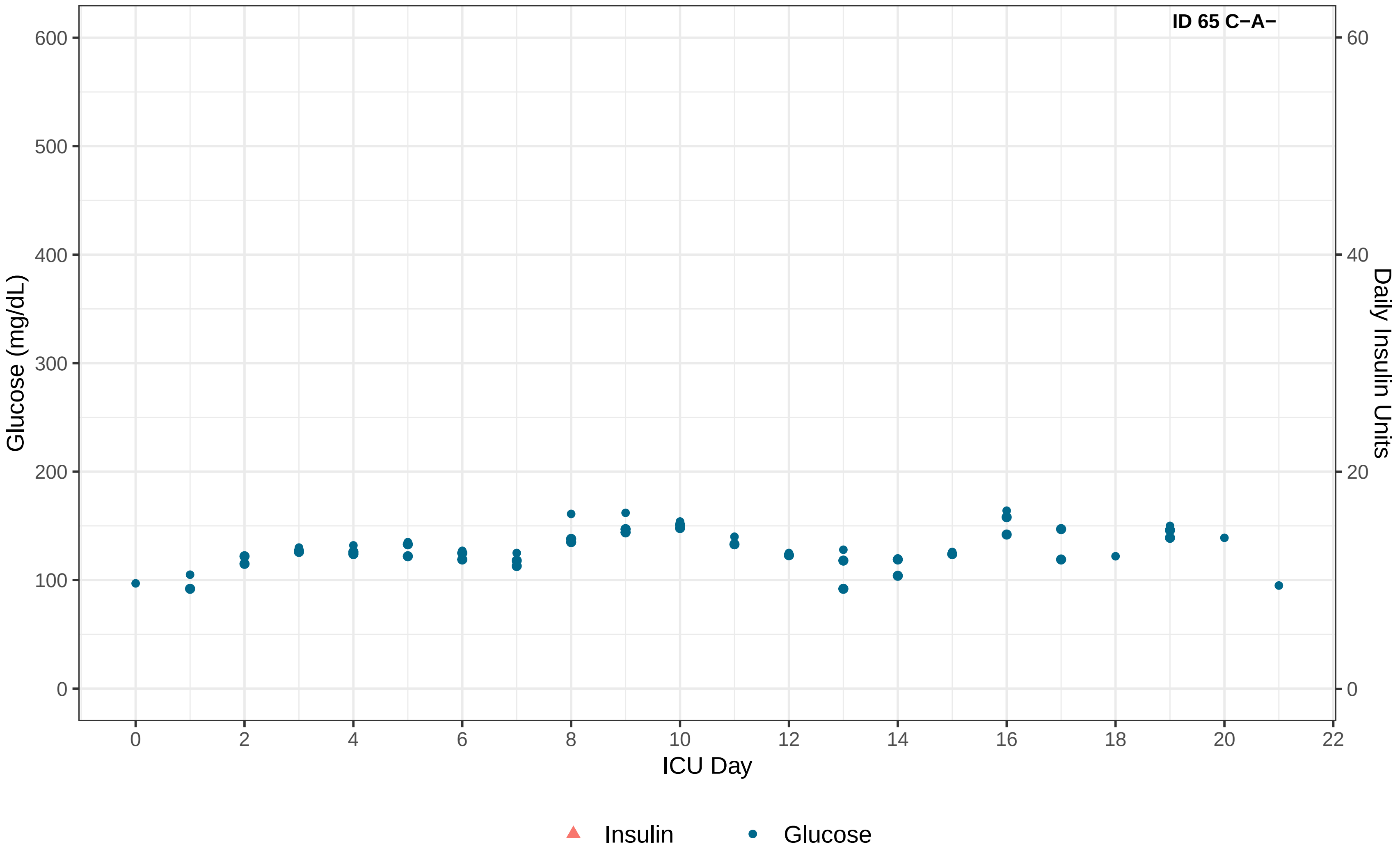

ID 65 C−A−
Glucose (mg/dL)
Daily Insulin Units
ICU Day
0
100
200
300
400
500
600
0
20
40
60
0
2
4
6
8
10
12
14
16
18
20
22
Insulin
Glucose

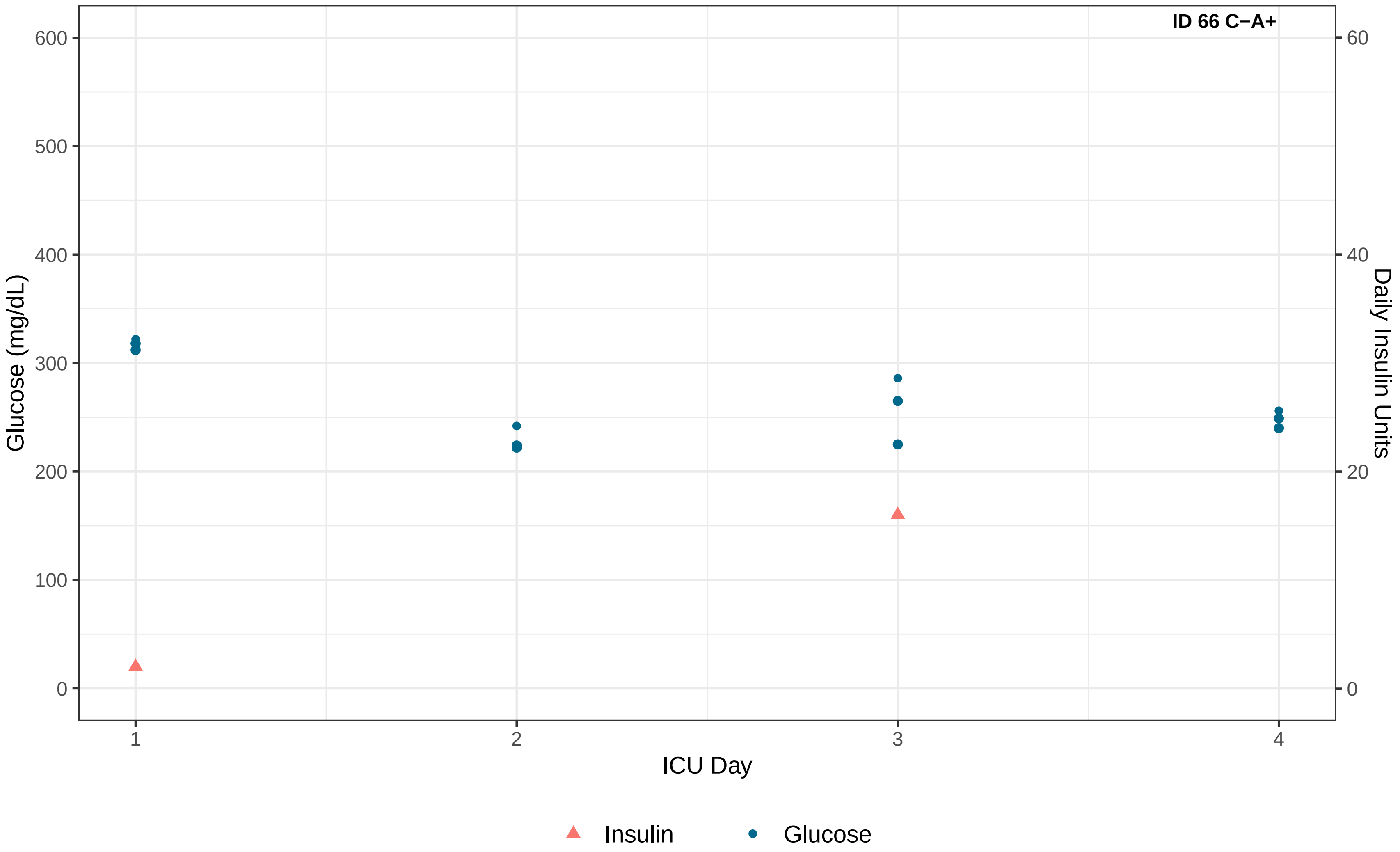
ID 66 C−A+
Glucose (mg/dL)
0
100
200
300
400
500
600
Daily Insulin Units
0
20
40
60
ICU Day
1
2
3
4
Insulin
Glucose

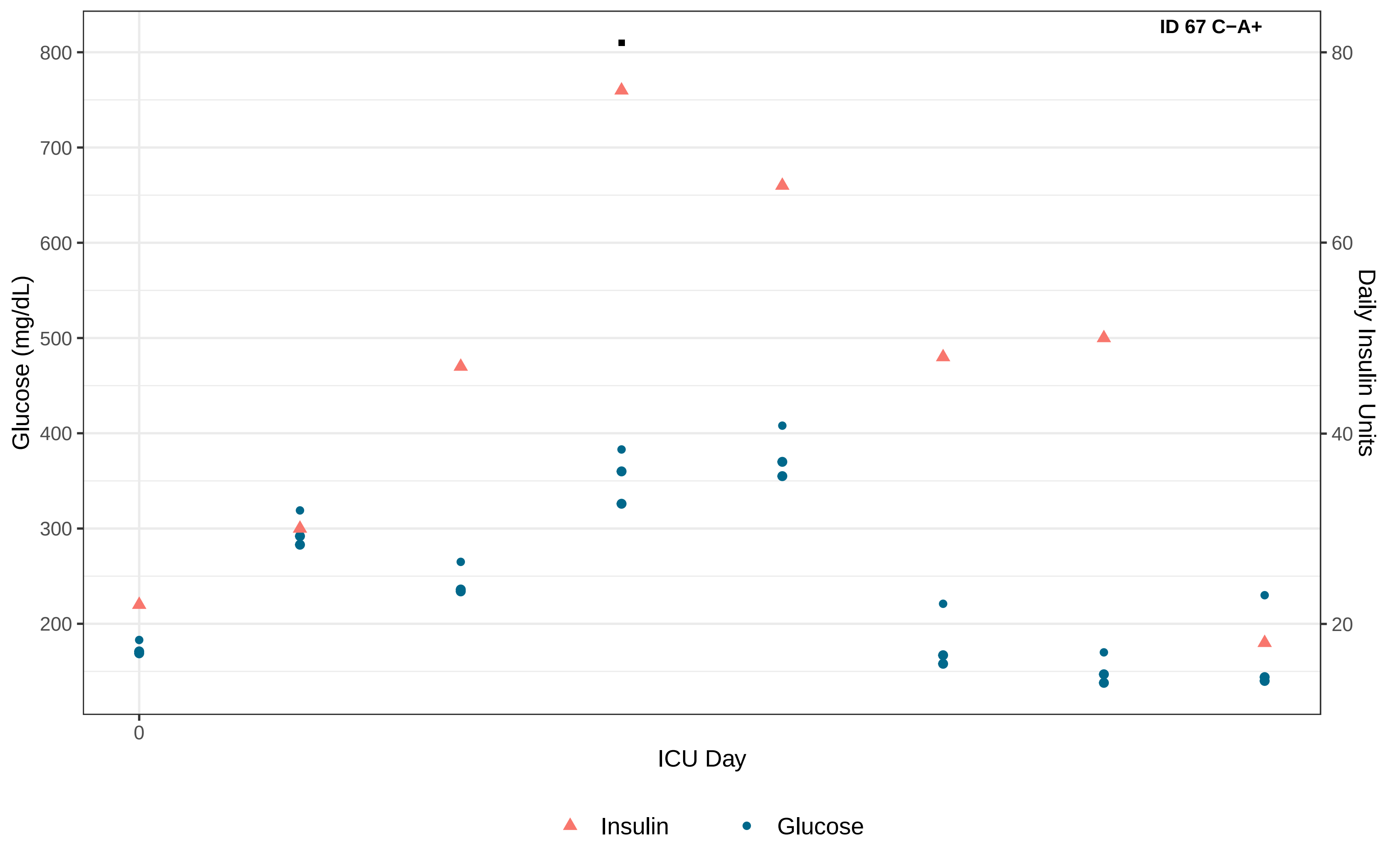
ID 67 C−A+
Glucose (mg/dL)
800
700
600
500
400
300
200
Daily Insulin Units
80
60
40
20
0
ICU Day
Insulin
Glucose

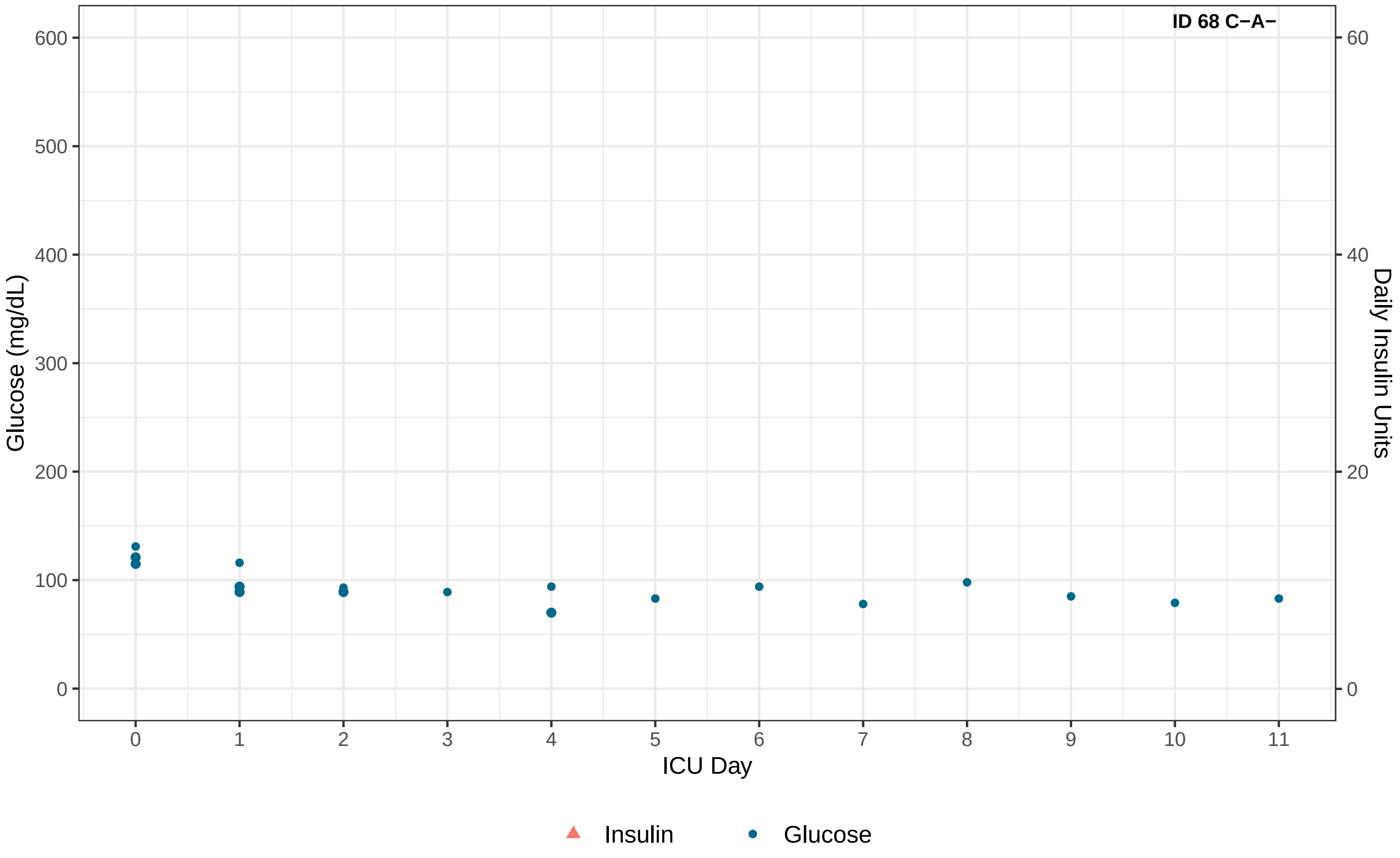

ID 68 C−A−
Glucose (mg/dL)
Daily Insulin Units
ICU Day
0
100
200
300
400
500
600
0
20
40
60
0
1
2
3
4
5
6
7
8
9
10
11
Insulin
Glucose

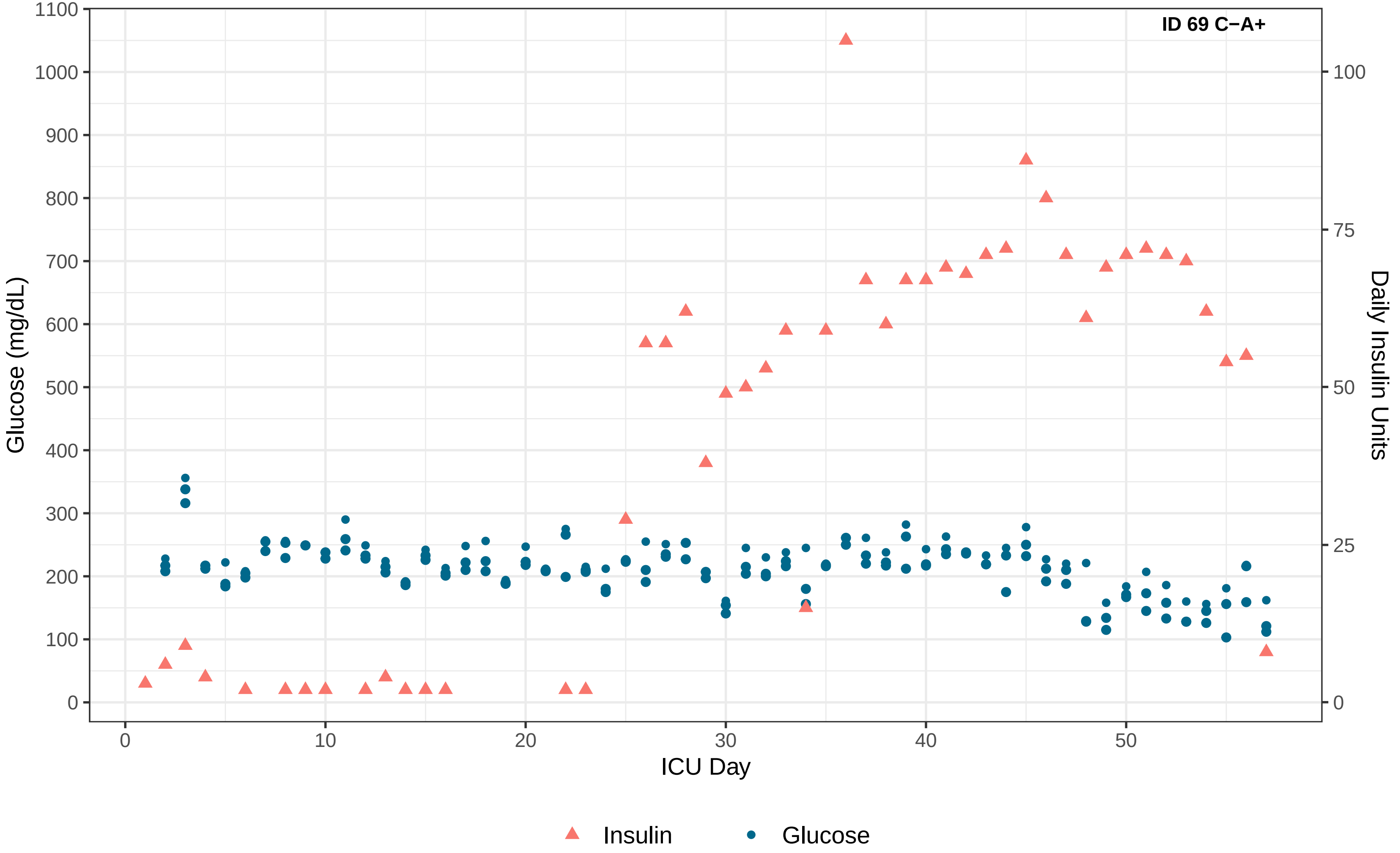
ID 69 C−A+
Glucose (mg/dL)
Daily Insulin Units
ICU Day
0
100
200
300
400
500
600
700
800
900
1000
1100
0
25
50
75
100
0
10
20
30
40
50
Insulin
Glucose

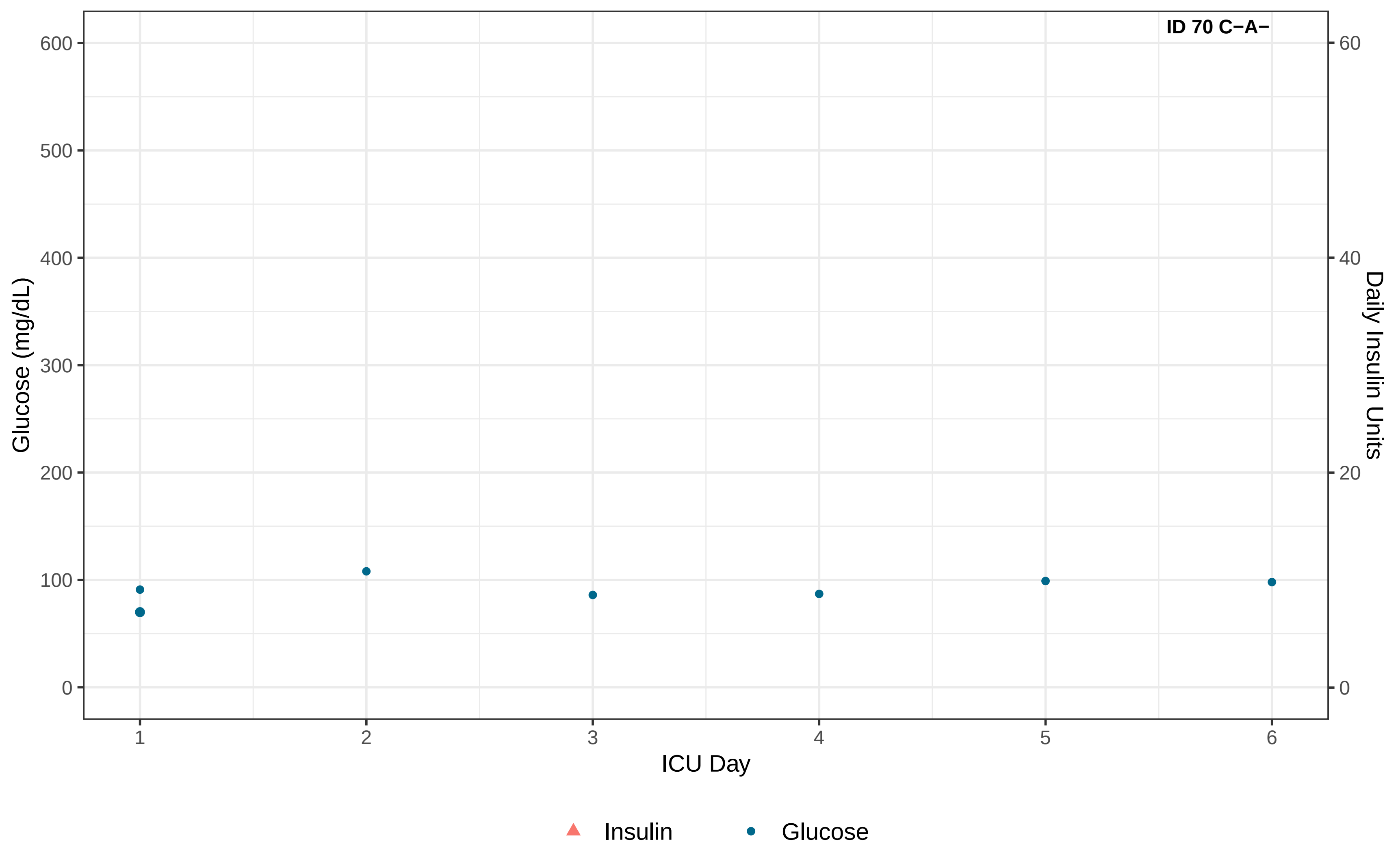
ID 70 C−A−
Glucose (mg/dL)
Daily Insulin Units
ICU Day
0
100
200
300
400
500
600
0
20
40
60
1
2
3
4
5
6
Insulin
Glucose

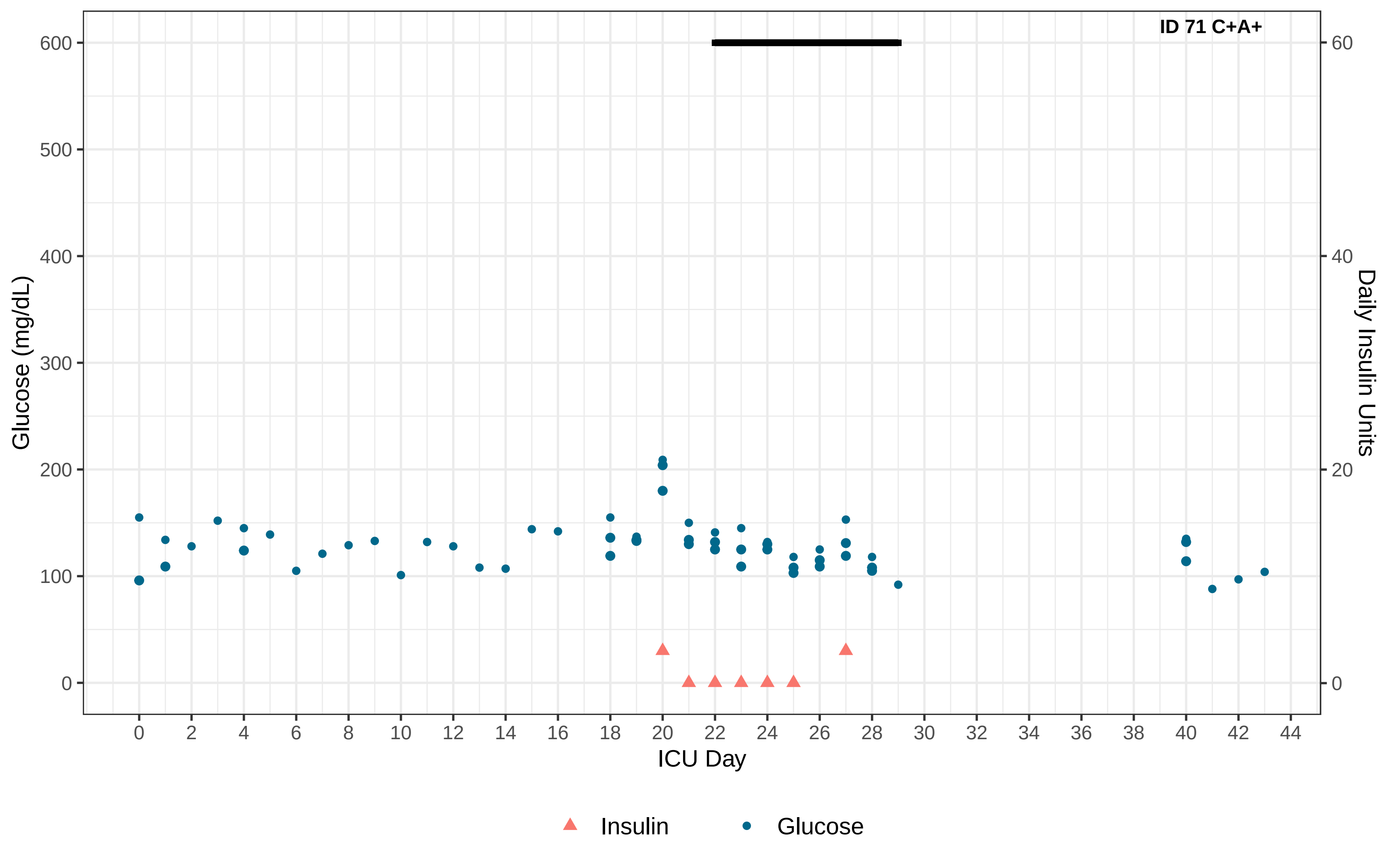

ID 71 C+A+
Glucose (mg/dL)
Daily Insulin Units
ICU Day
0
100
200
300
400
500
600
20
40
60
2
4
6
8
10
12
14
16
18
22
24
26
28
30
32
34
36
38
42
44
Insulin
Glucose

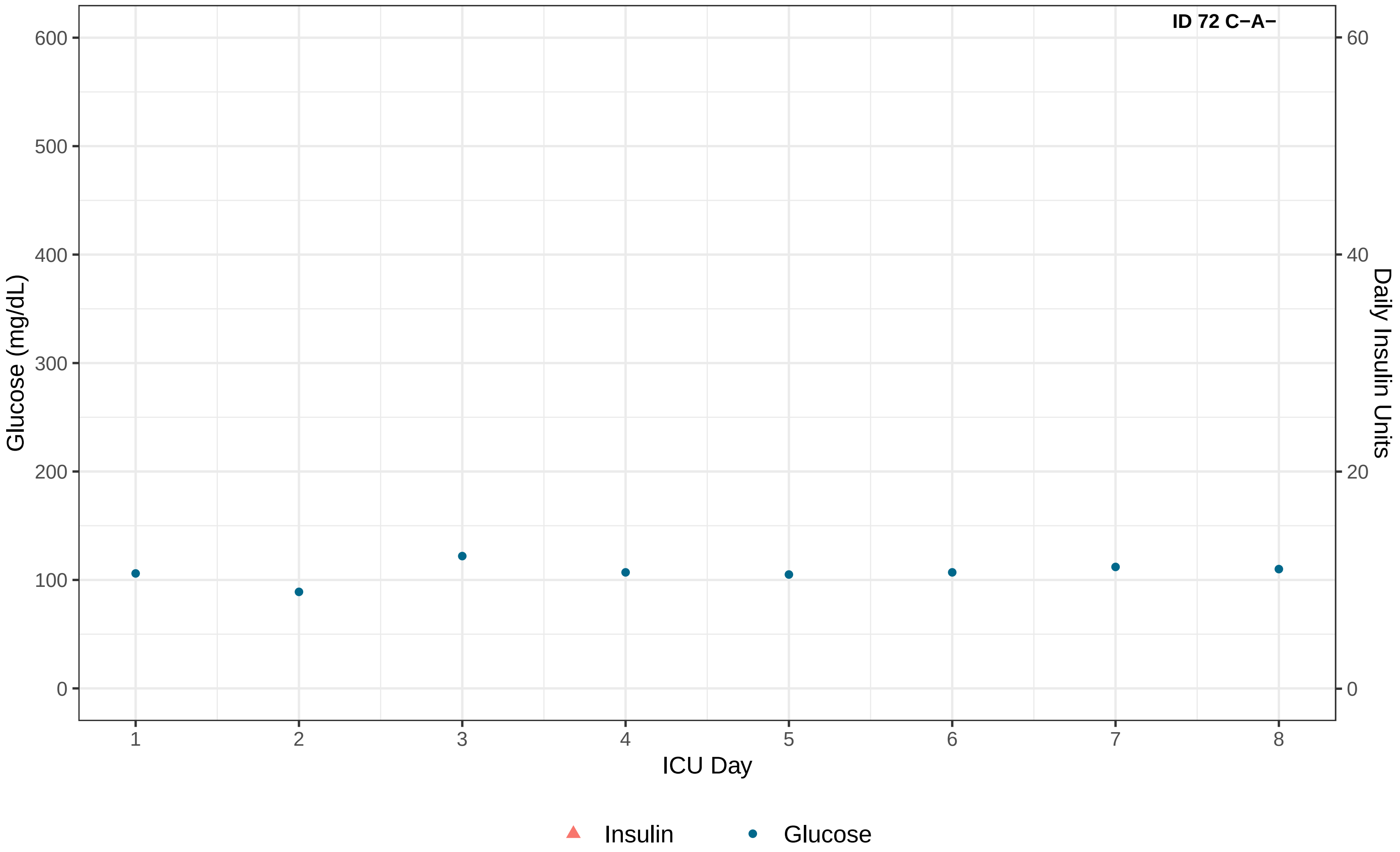
ID 72 C−A−
Glucose (mg/dL)
600
500
400
300
200
100
0
Daily Insulin Units
60
40
20
0
1
2
3
4
5
6
7
8
ICU Day
Insulin
Glucose

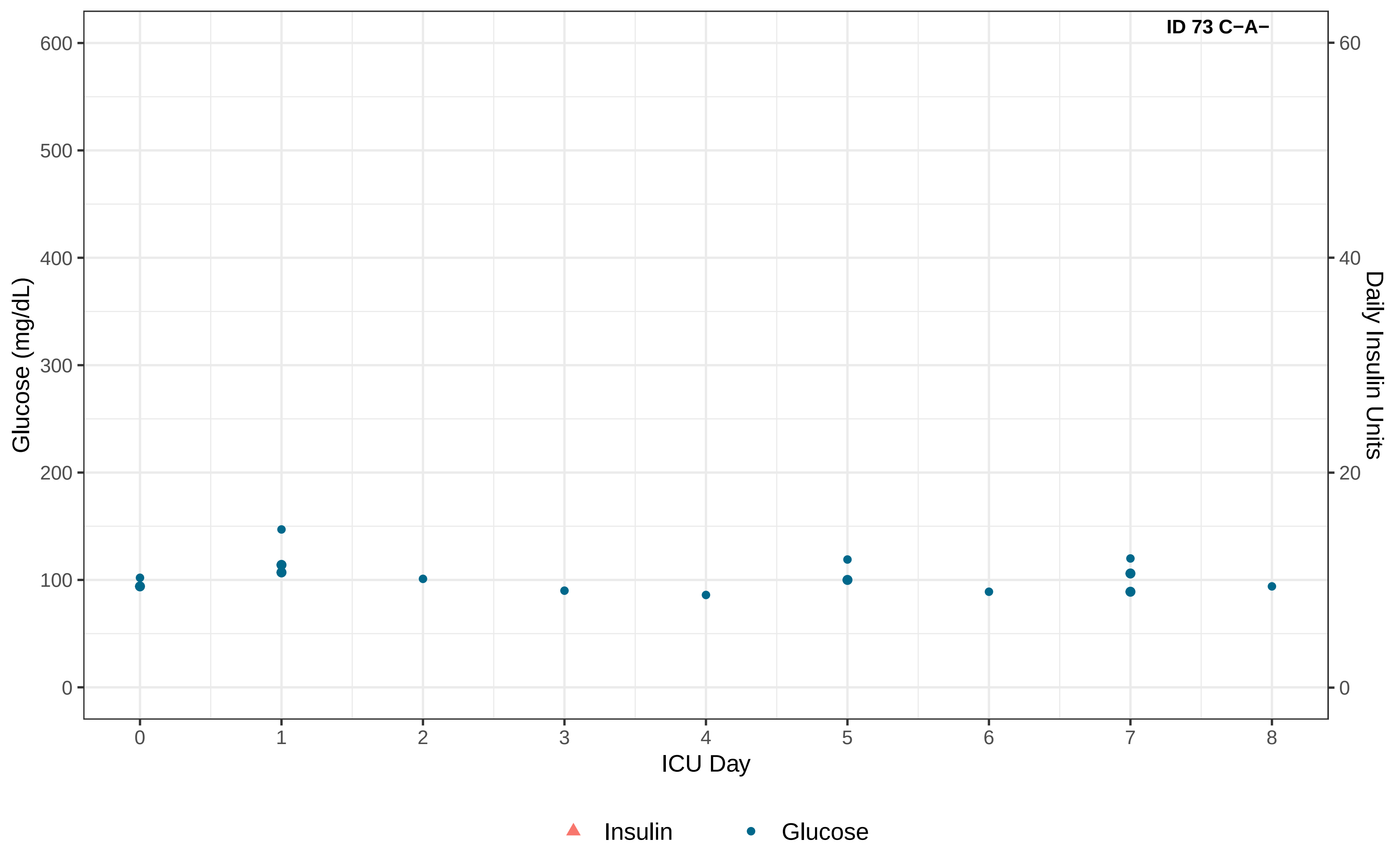

ID 73 C−A−
Glucose (mg/dL)
600
500
400
300
200
100
0
Daily Insulin Units
60
40
20
0
ICU Day
0
1
2
3
4
5
6
7
8
Insulin
Glucose

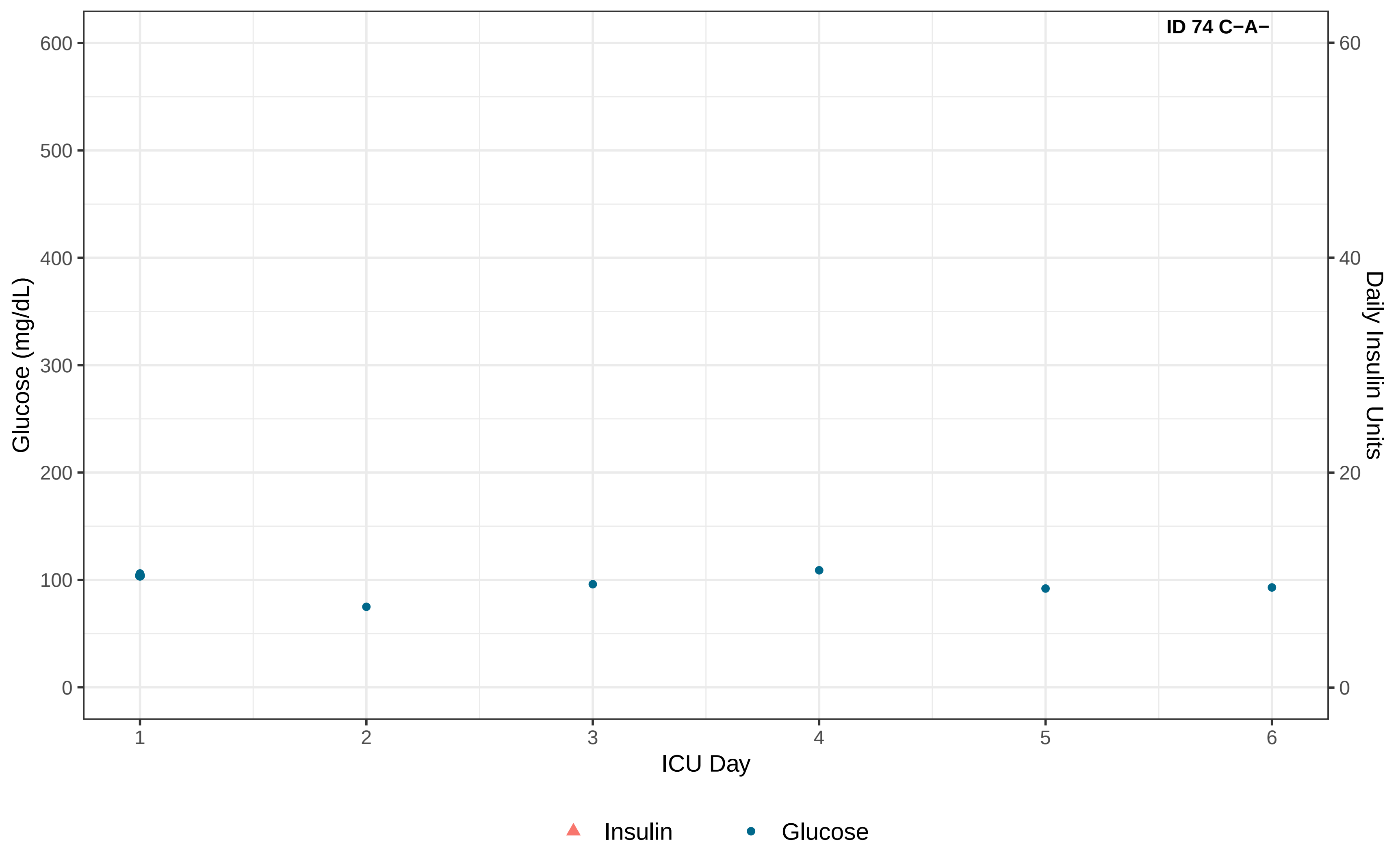
ID 74 C−A−
Glucose (mg/dL)
Daily Insulin Units
ICU Day
0
100
200
300
400
500
600
0
20
40
60
1
2
3
4
5
6
Insulin
Glucose

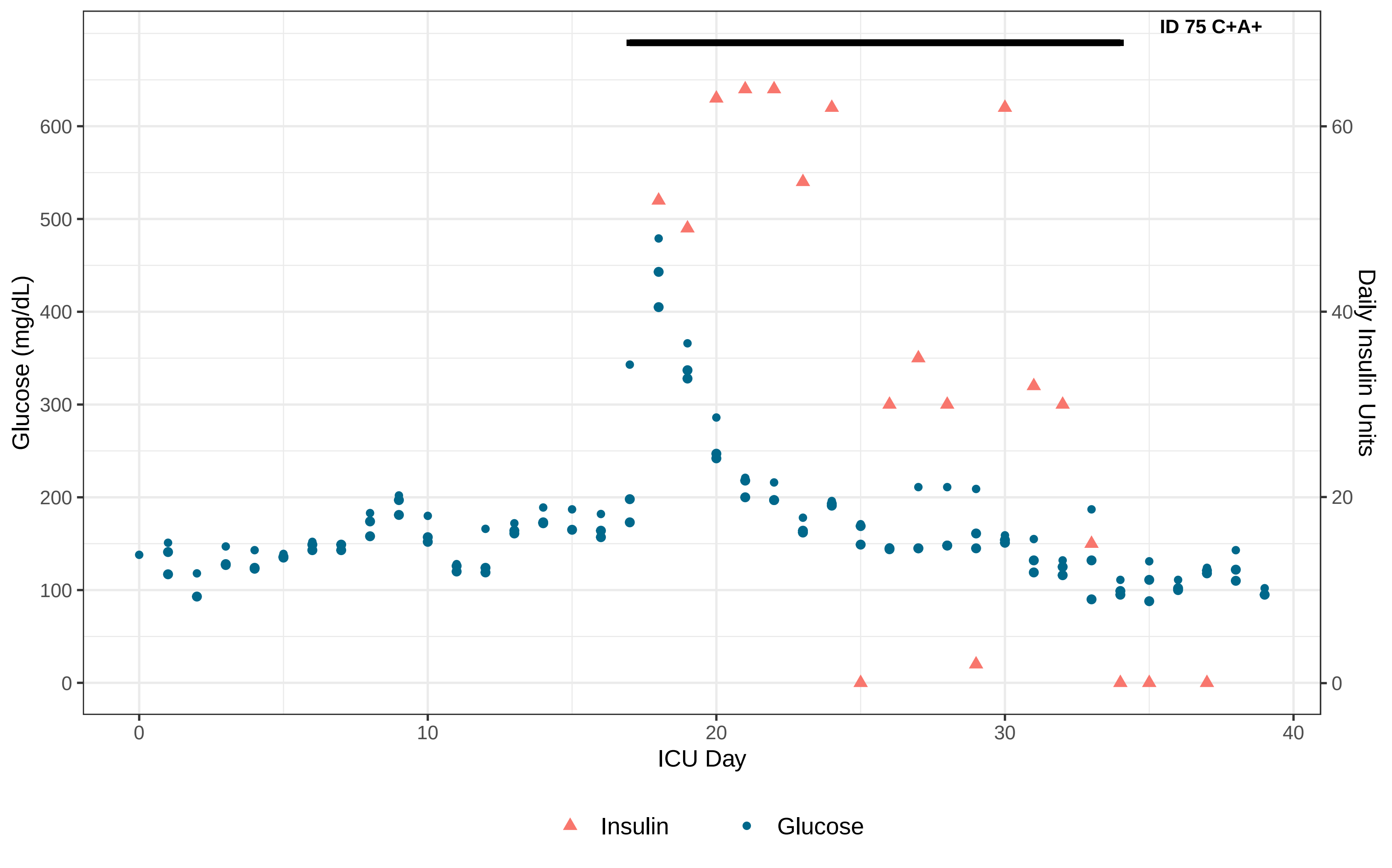

ID 75 C+A+
Glucose (mg/dL)
0
100
200
300
400
500
600
Daily Insulin Units
0
20
40
60
ICU Day
0
10
20
30
40
Insulin
Glucose

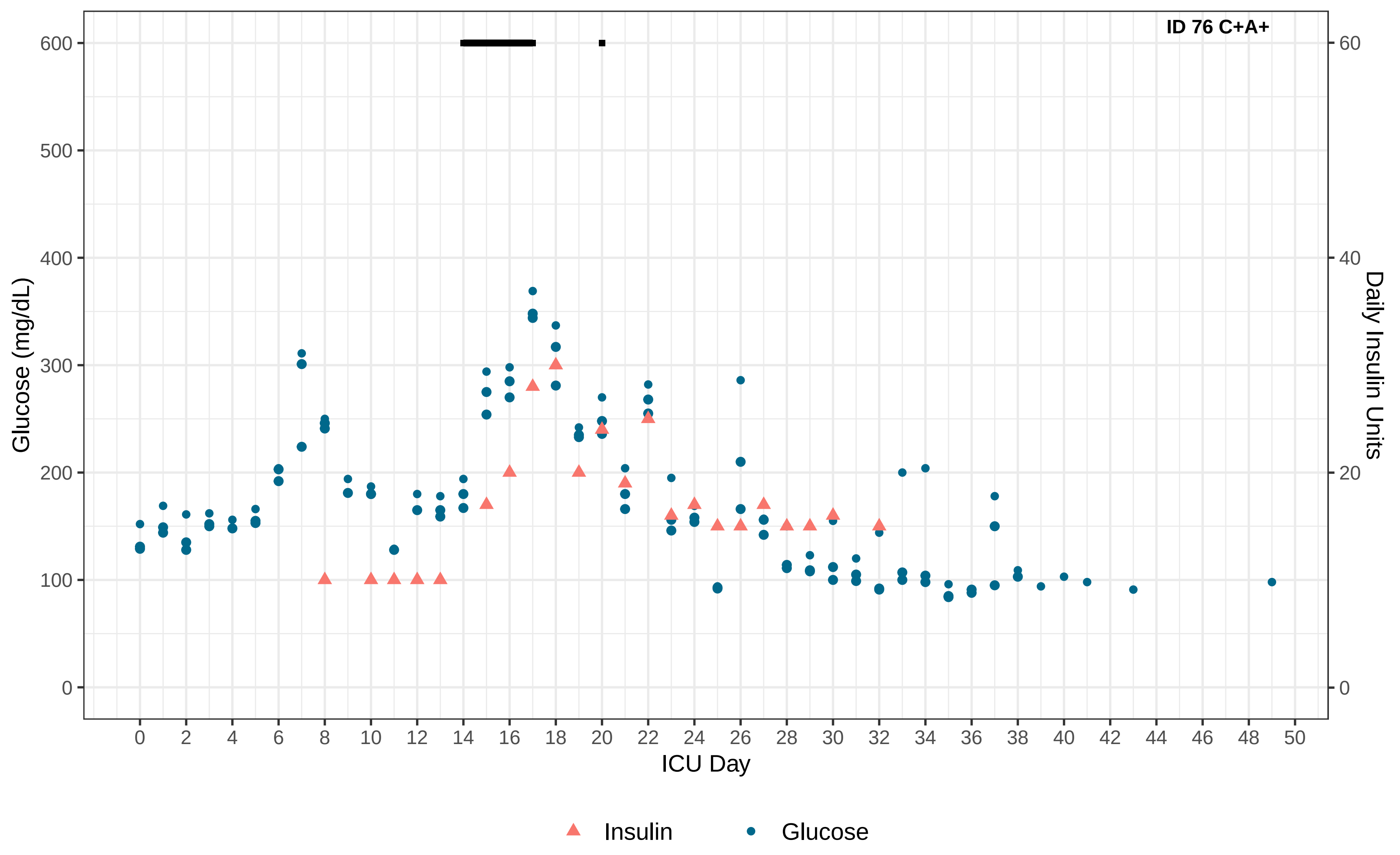

ID 76 C+A+
Glucose (mg/dL)
Daily Insulin Units
ICU Day
0
100
200
300
400
500
600
0
20
40
60
0 2 4 6 8 10 12 14 16 18 20 22 24 26 28 30 32 34 36 38 40 42 44 46 48 50
Insulin
Glucose

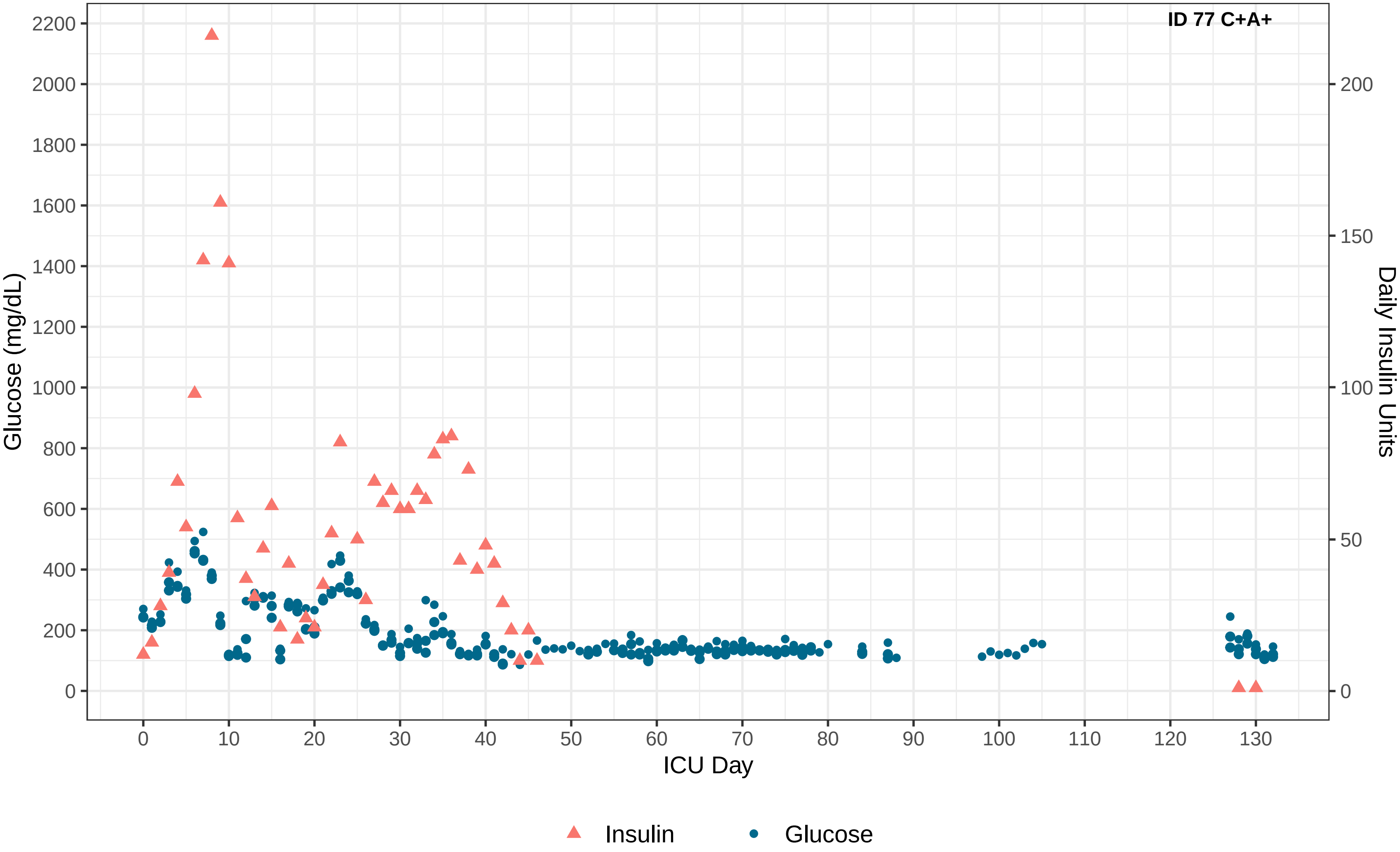
ID 77 C+A+
Glucose (mg/dL)
Daily Insulin Units
ICU Day
0
200
400
600
800
1000
1200
1400
1600
1800
2000
2200
0
50
100
150
200
0
10
20
30
40
50
60
70
80
90
100
110
120
130
Insulin
Glucose

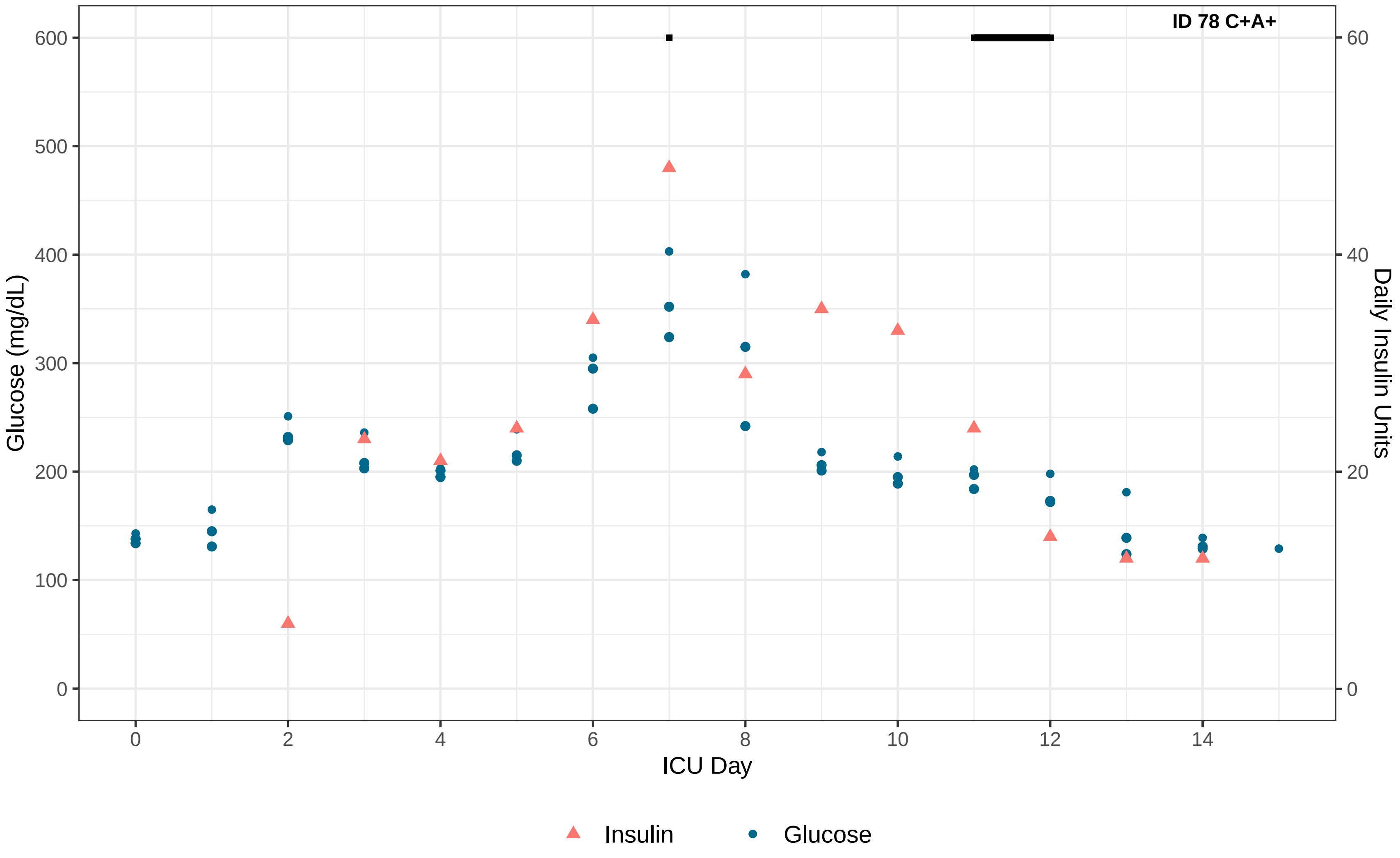
ID 78 C+A+
Glucose (mg/dL)
Daily Insulin Units
ICU Day
0
100
200
300
400
500
600
0
20
40
60
0
2
4
6
8
10
12
14
Insulin
Glucose

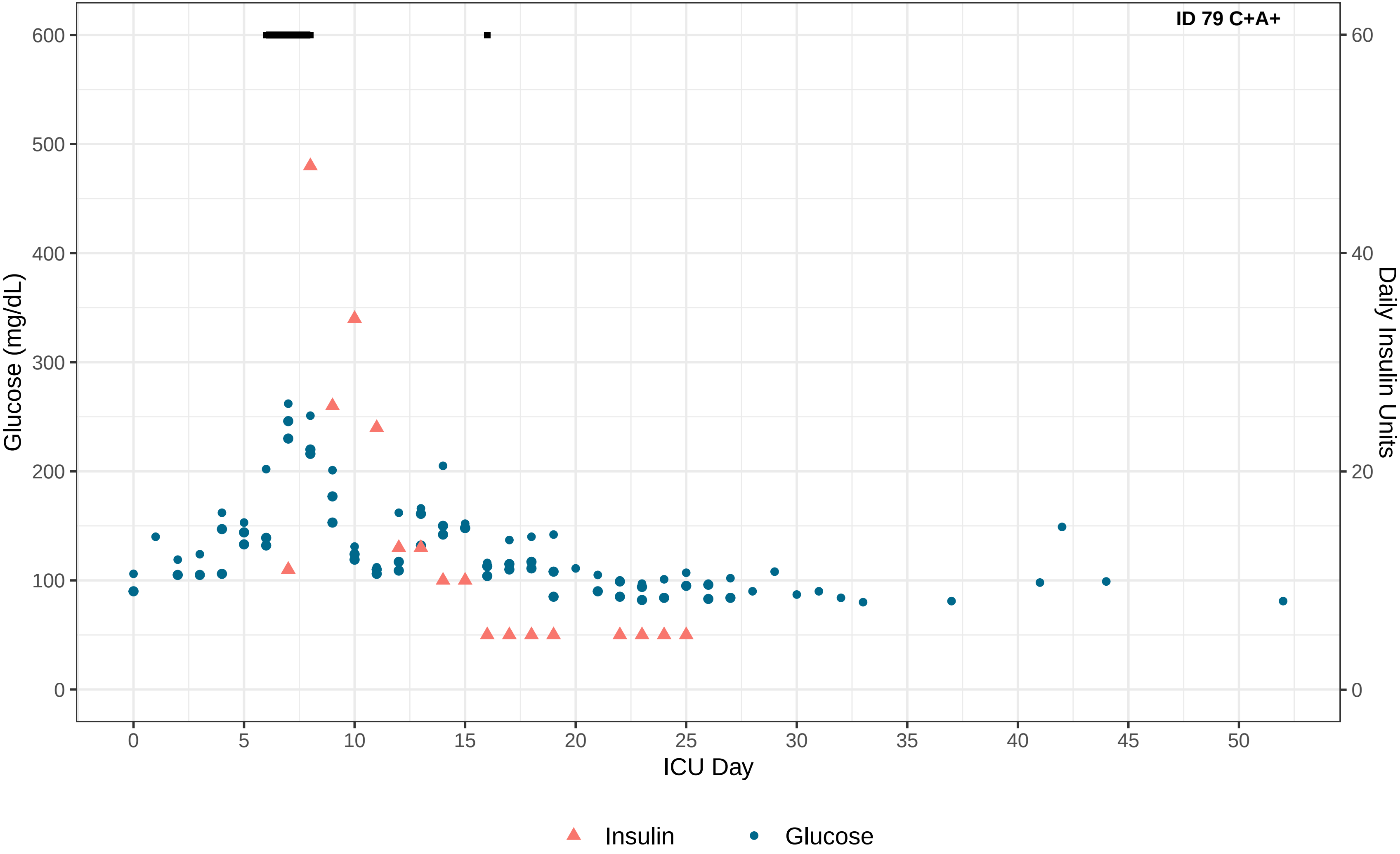

ID 79 C+A+
Glucose (mg/dL)
600
500
400
300
200
100
0
Daily Insulin Units
60
40
20
0
0
5
10
15
20
25
30
35
40
45
50
ICU Day
Insulin
Glucose

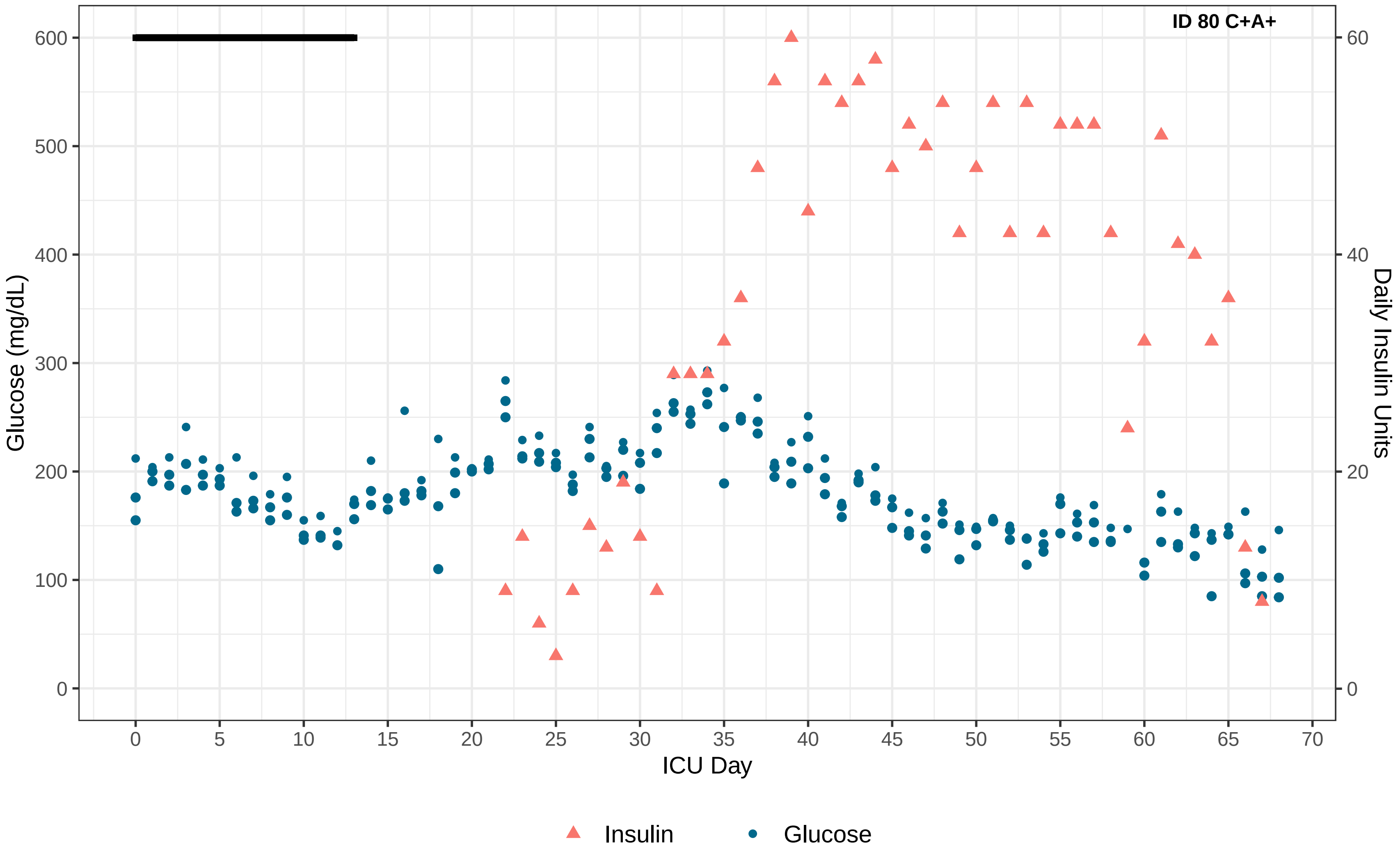
ID 80 C+A+
Glucose (mg/dL)
Daily Insulin Units
ICU Day
0
100
200
300
400
500
600
0
20
40
60
0
5
10
15
20
25
30
35
40
45
50
55
60
65
70
Insulin
Glucose

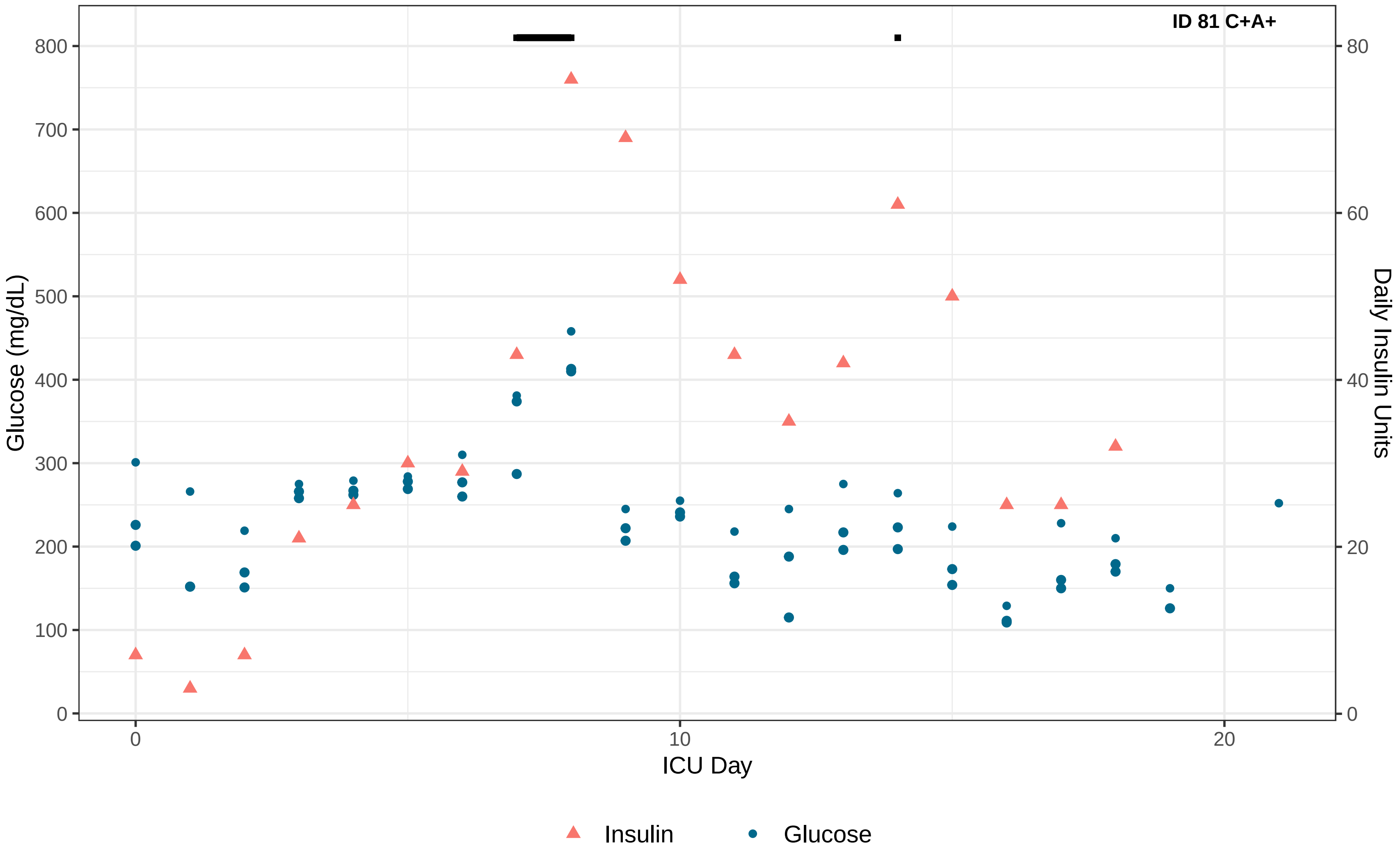

ID 81 C+A+
Glucose (mg/dL)
0
100
200
300
400
500
600
700
800
Daily Insulin Units
0
20
40
60
80
ICU Day
0
10
20
Insulin
Glucose

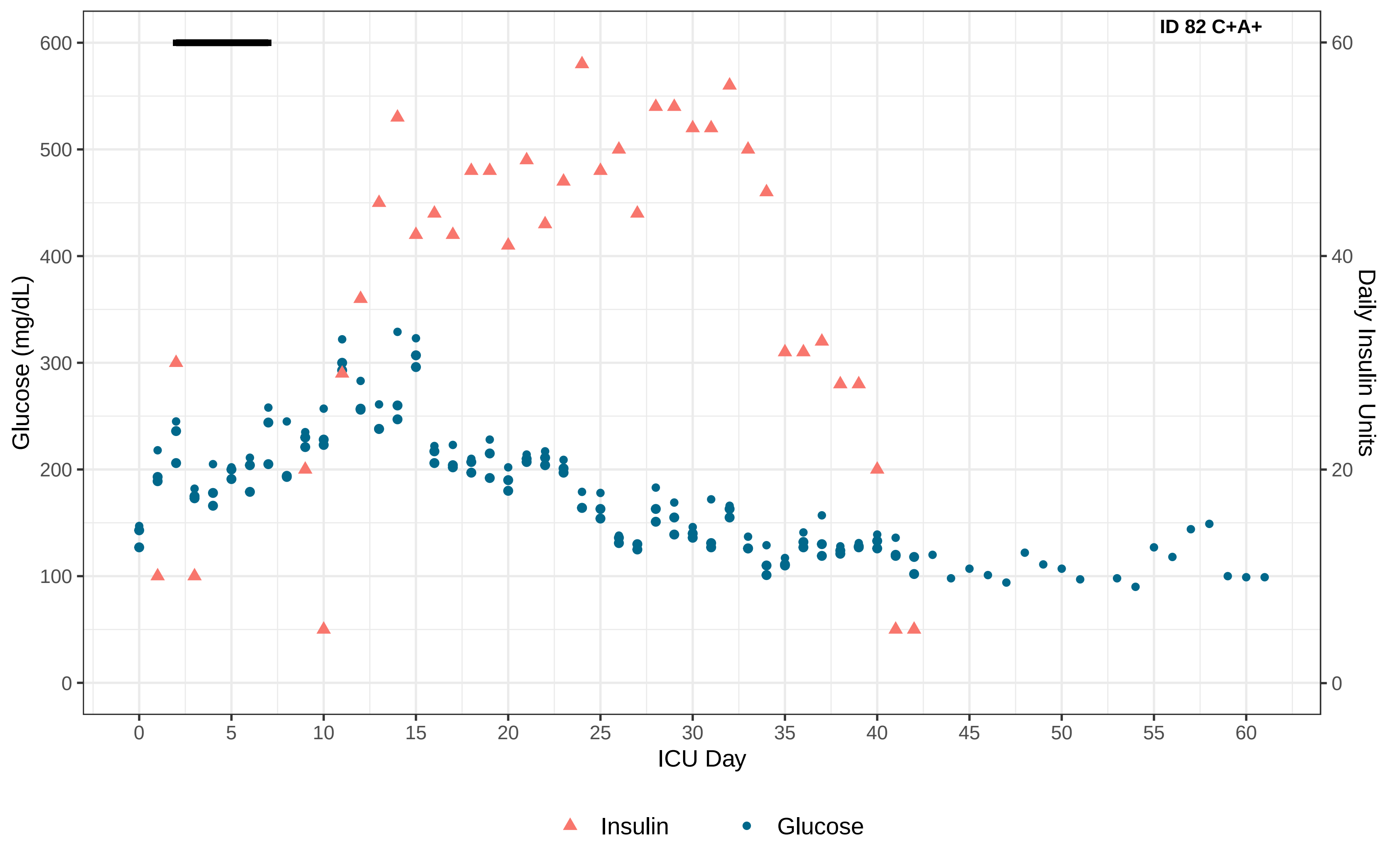

ID 82 C+A+
Glucose (mg/dL)
Daily Insulin Units
ICU Day
0
100
200
300
400
500
600
0
20
40
60
0
5
10
15
20
25
30
35
40
45
50
55
60
Insulin
Glucose

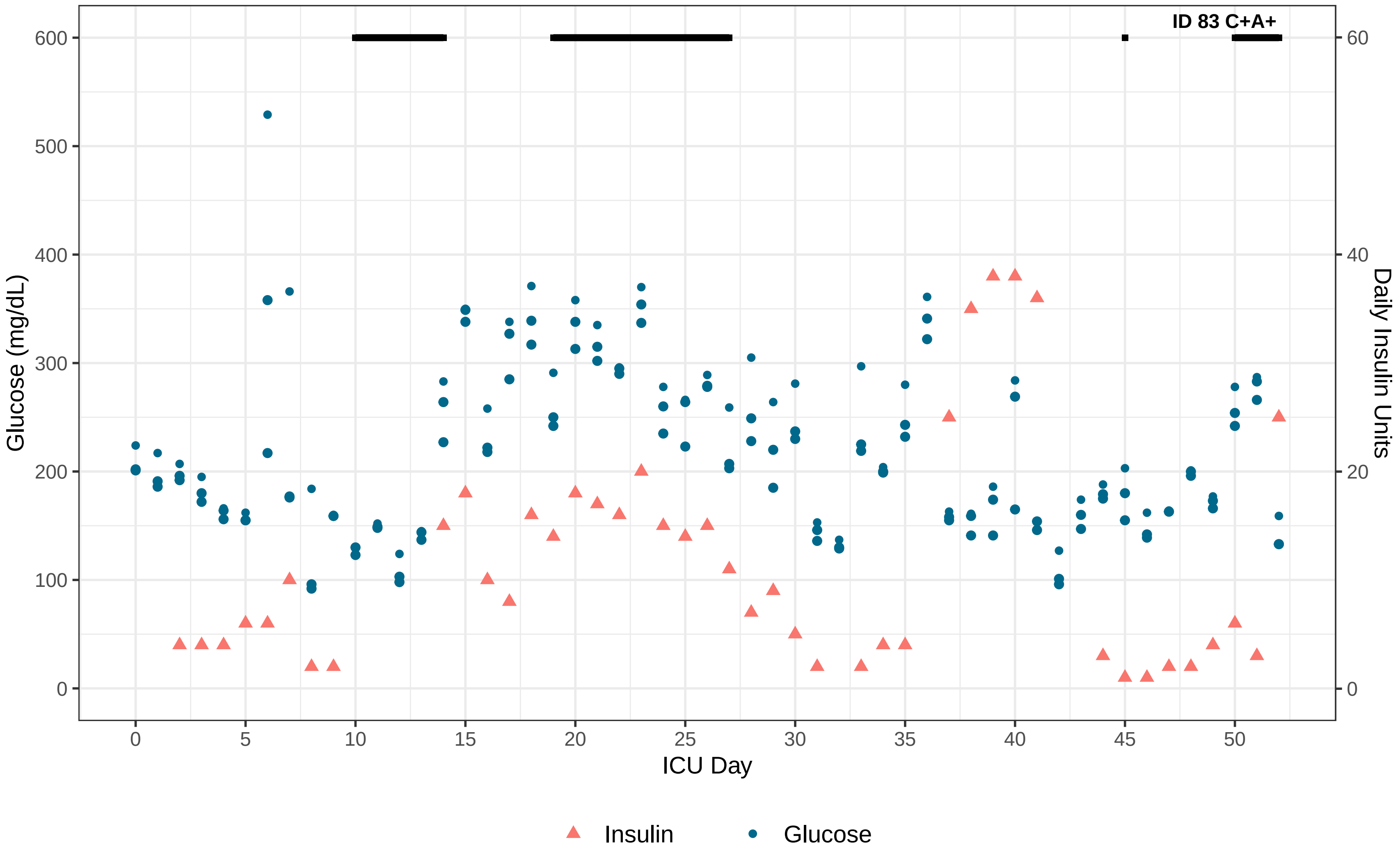

ID 83 C+A+
Glucose (mg/dL)
Daily Insulin Units
ICU Day
0
100
200
300
400
500
600
0
20
40
60
0
5
10
15
20
25
30
35
40
45
50
Insulin
Glucose

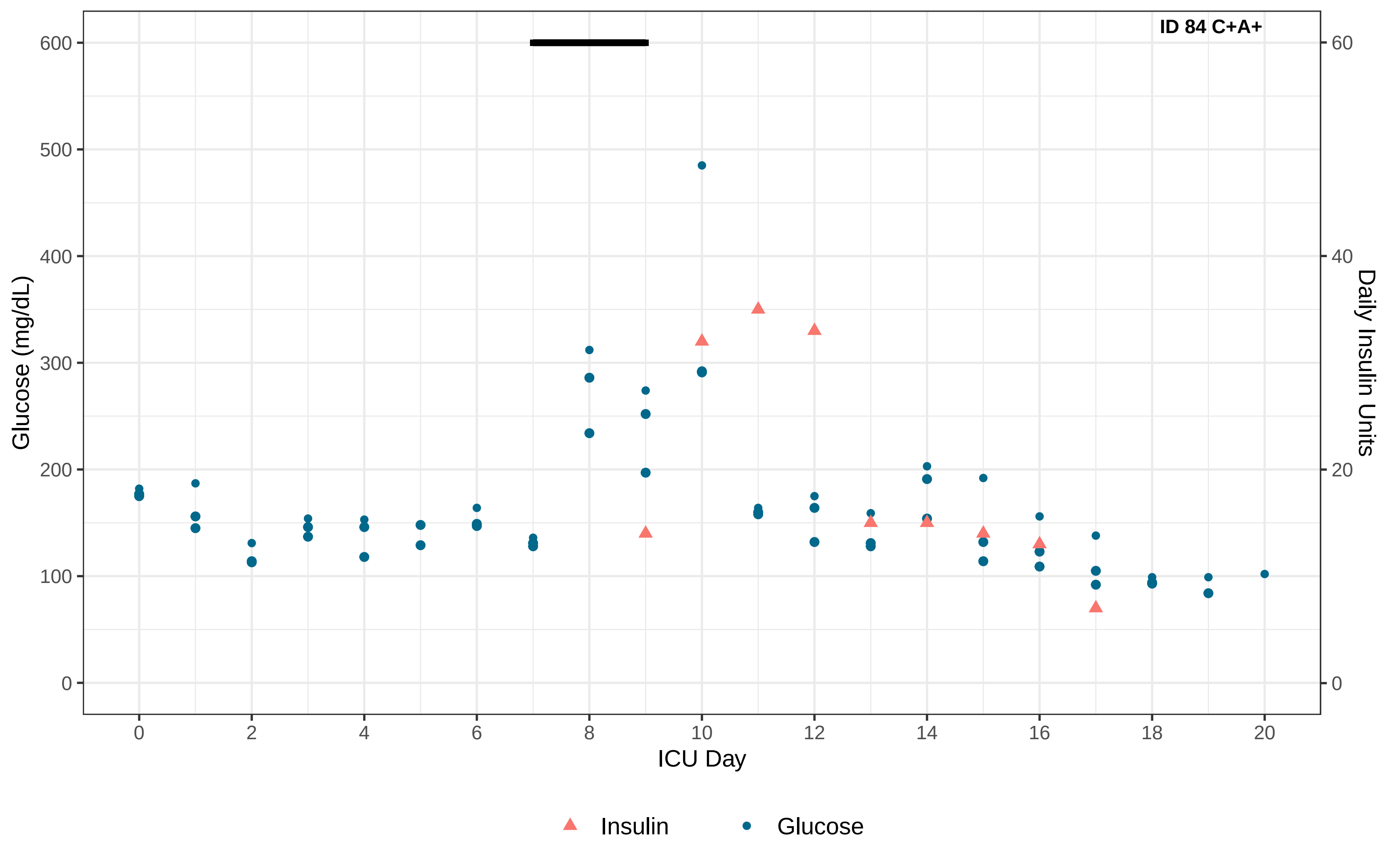

ID 84 C+A+
Glucose (mg/dL)
Daily Insulin Units
ICU Day
0
100
200
300
400
500
600
0
20
40
60
0
2
4
6
8
10
12
14
16
18
20
Insulin
Glucose

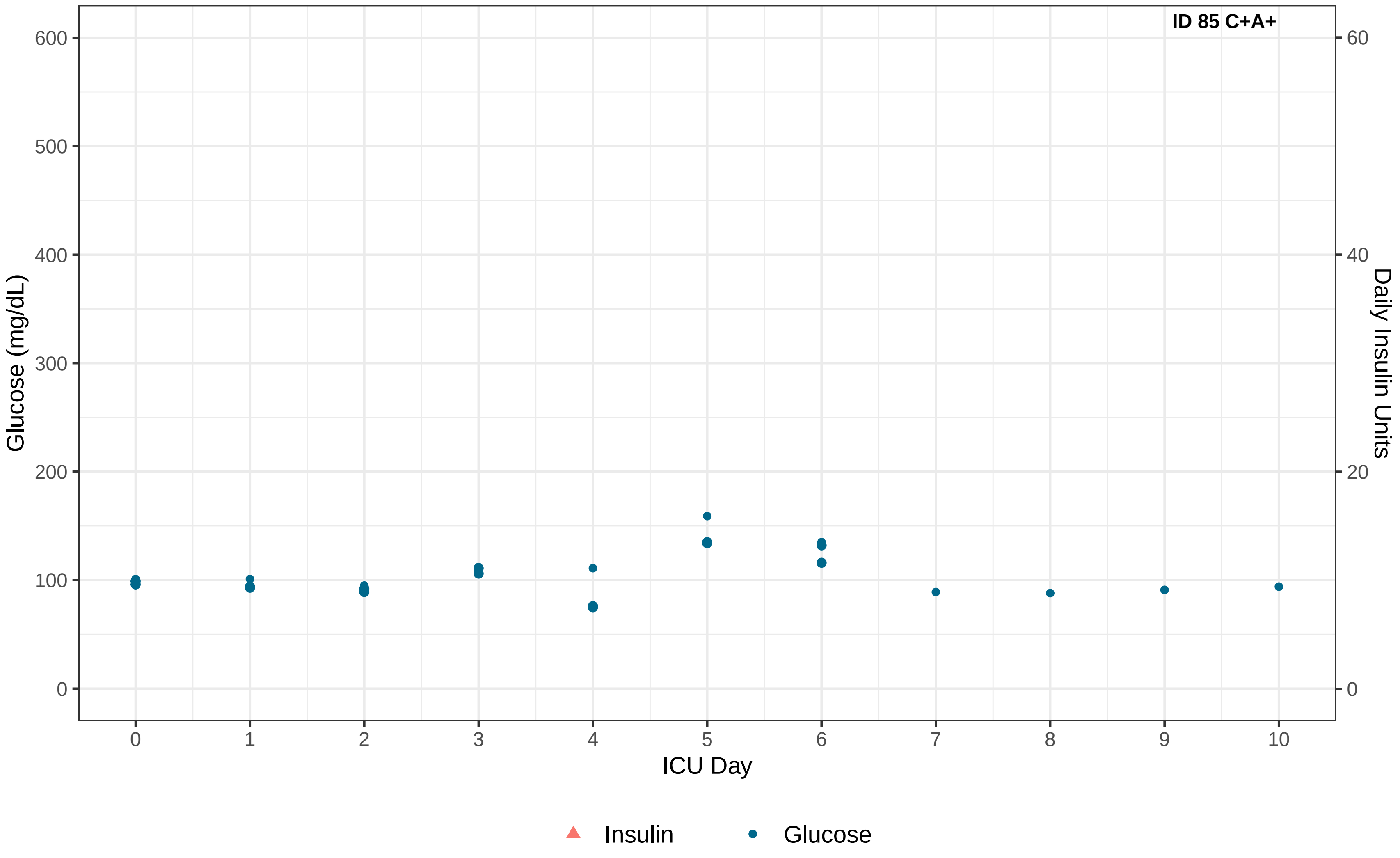

ID 85 C+A+
Glucose (mg/dL)
0
100
200
300
400
500
600
Daily Insulin Units
0
20
40
60
ICU Day
0
1
2
3
4
5
6
7
8
9
10
Insulin
Glucose

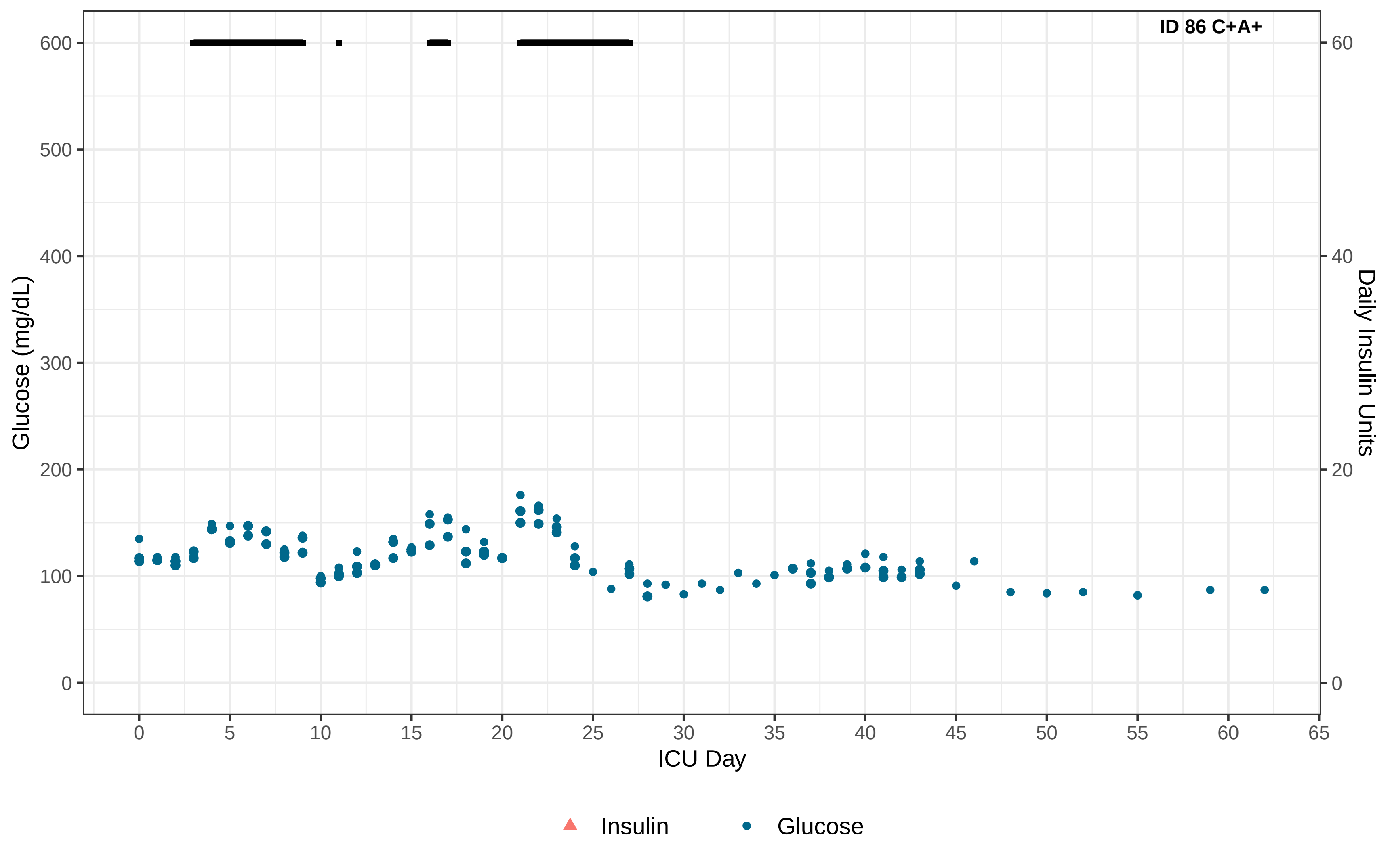

ID 86 C+A+
Glucose (mg/dL)
Daily Insulin Units
ICU Day
0
100
200
300
400
500
600
0
20
40
60
0
5
10
15
20
25
30
35
40
45
50
55
60
65
Insulin
Glucose

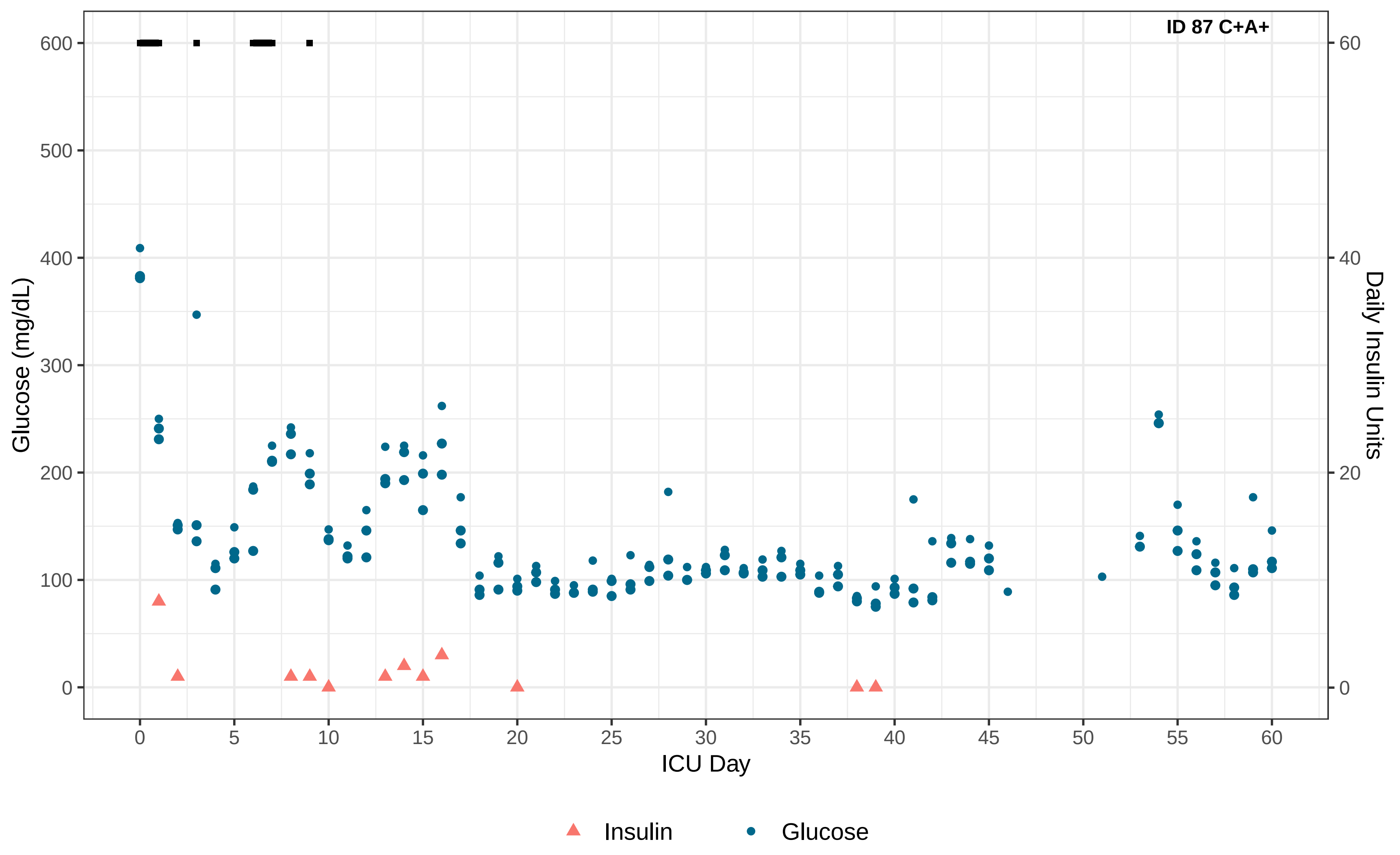

ID 87 C+A+
Glucose (mg/dL)
Daily Insulin Units
ICU Day
0
100
200
300
400
500
600
0
20
40
60
0
5
10
15
20
25
30
35
40
45
50
55
60
Insulin
Glucose

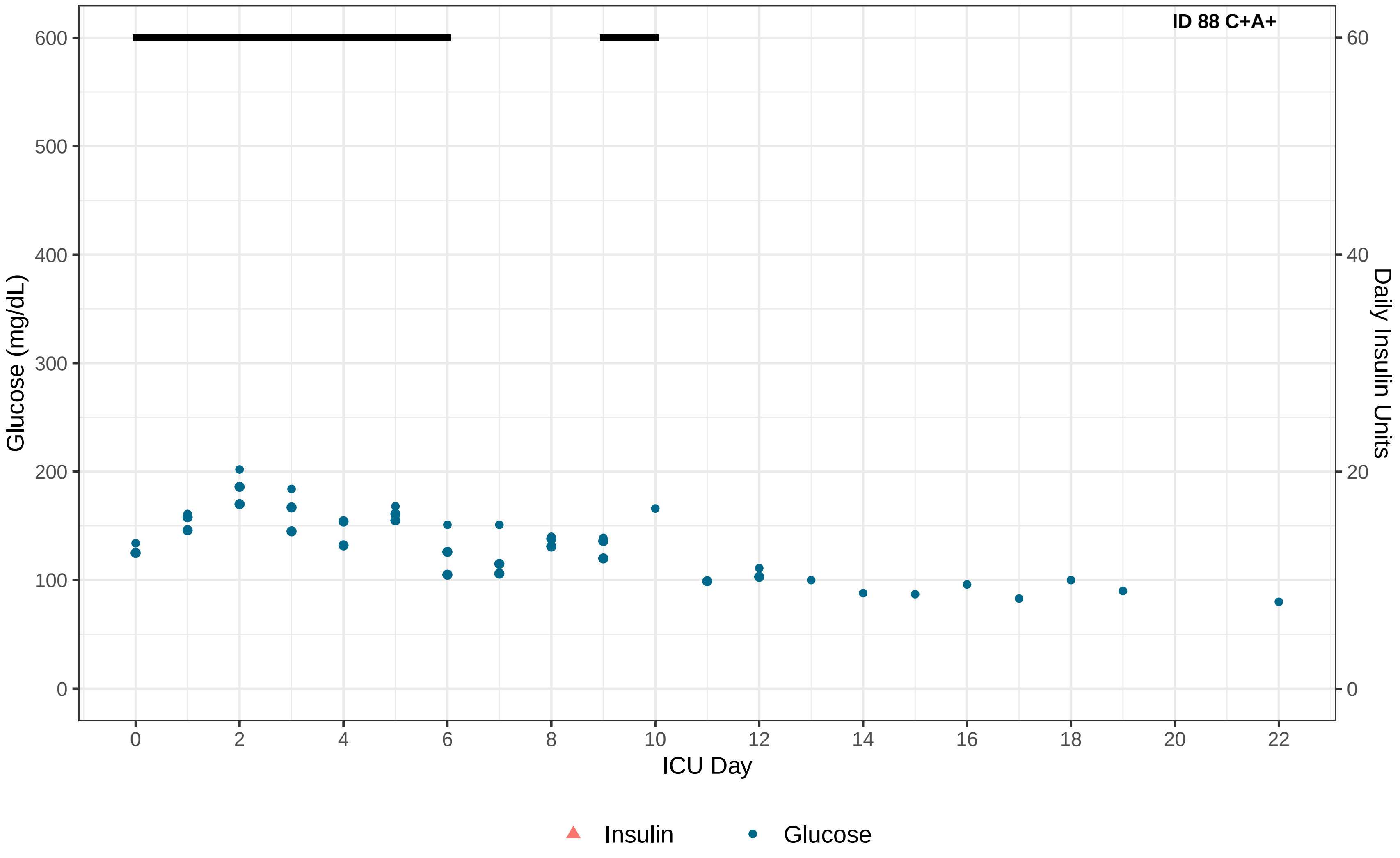

ID 88 C+A+
Glucose (mg/dL)
Daily Insulin Units
ICU Day
0
100
200
300
400
500
600
0
20
40
60
0
2
4
6
8
10
12
14
16
18
20
22
Insulin
Glucose

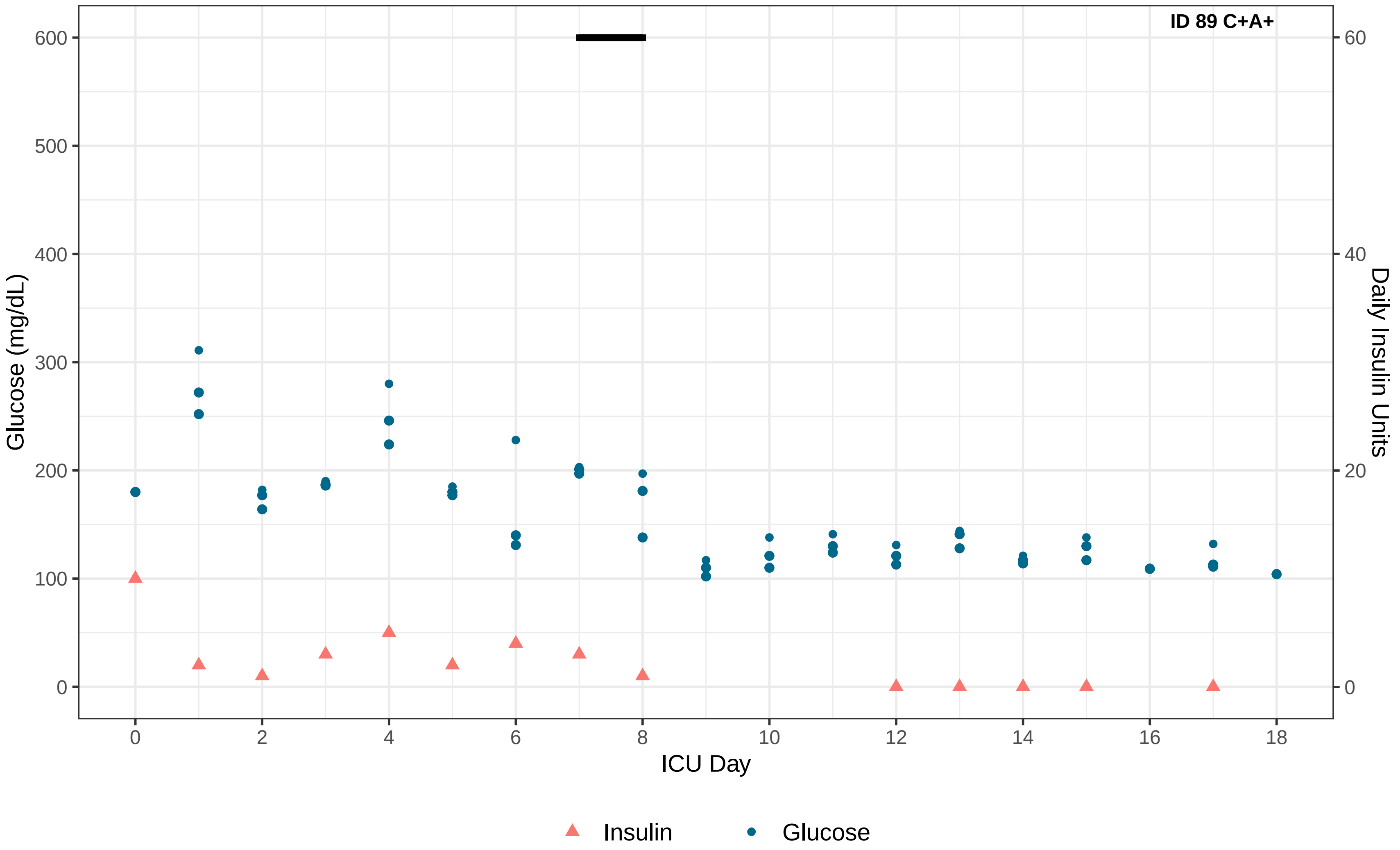
ID 89 C+A+
Glucose (mg/dL)
600
500
400
300
200
100
0
Daily Insulin Units
60
40
20
0
0
2
4
6
8
10
12
14
16
18
ICU Day
Insulin
Glucose

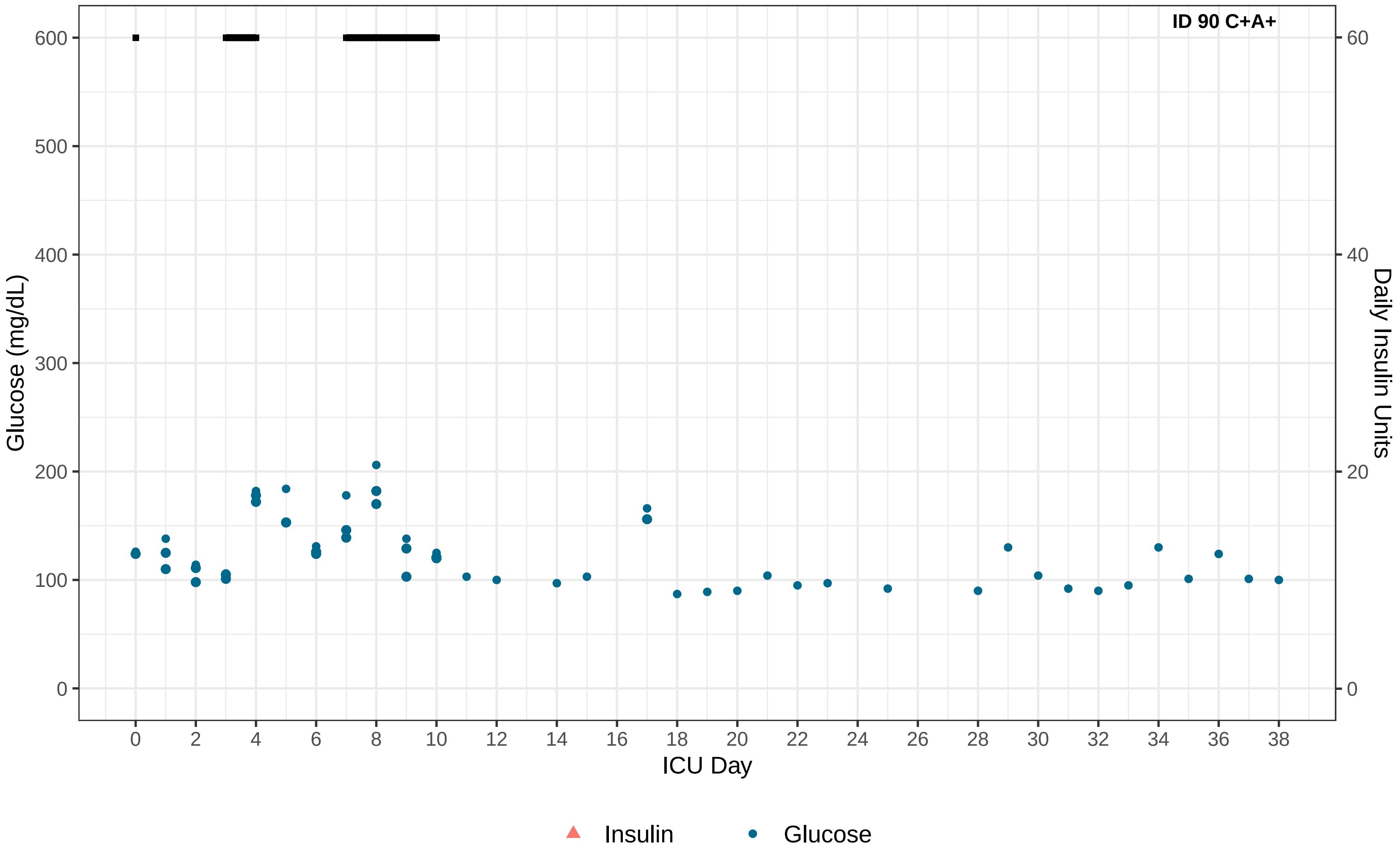

ID 90 C+A+
Glucose (mg/dL)
Daily Insulin Units
ICU Day
0
100
200
300
400
500
600
0
20
40
60
0
2
4
6
8
10
12
14
16
18
20
22
24
26
28
30
32
34
36
38
Insulin
Glucose

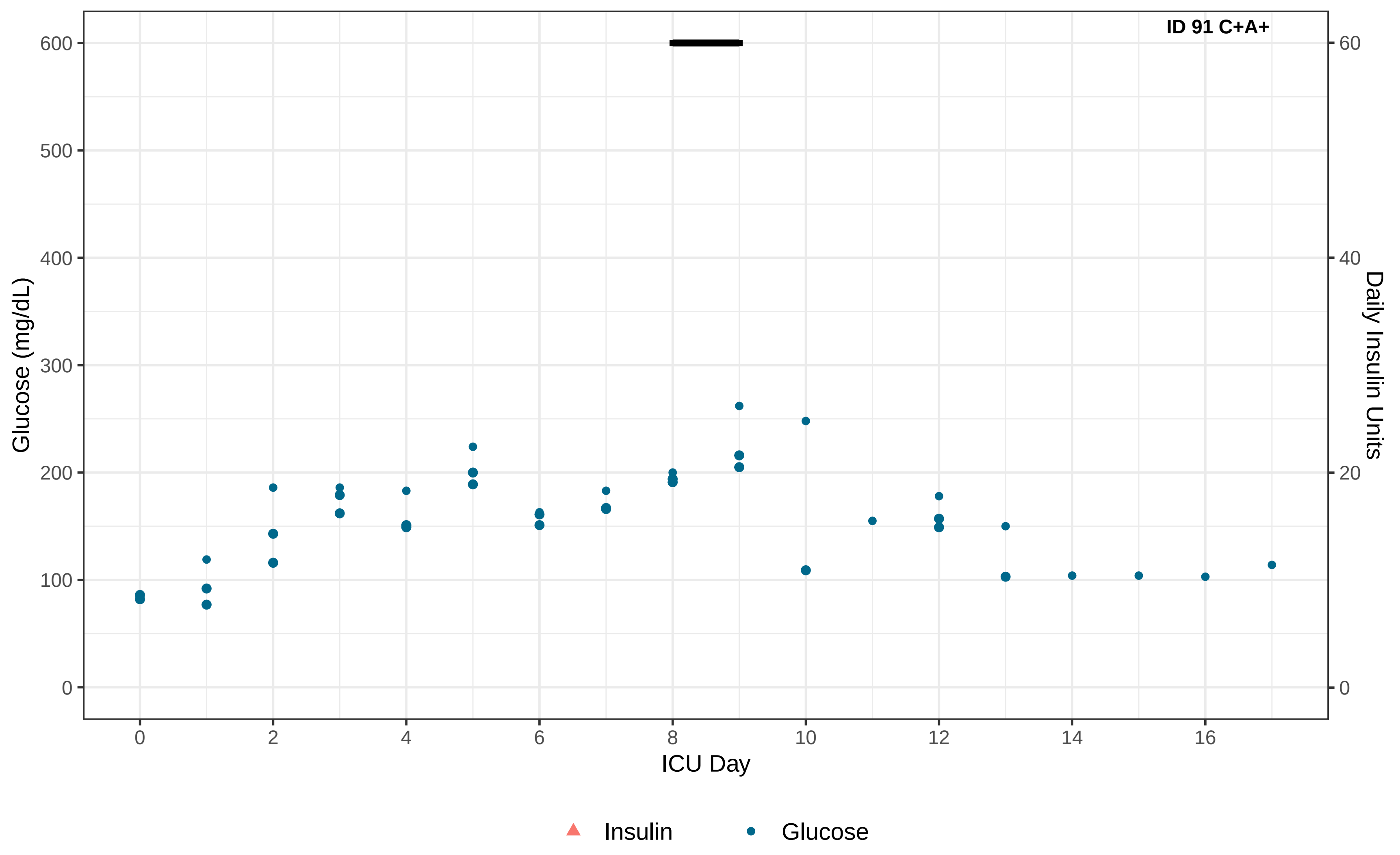

ID 91 C+A+
Glucose (mg/dL)
Daily Insulin Units
ICU Day
0
100
200
300
400
500
600
0
20
40
60
0
2
4
6
8
10
12
14
16
Insulin
Glucose

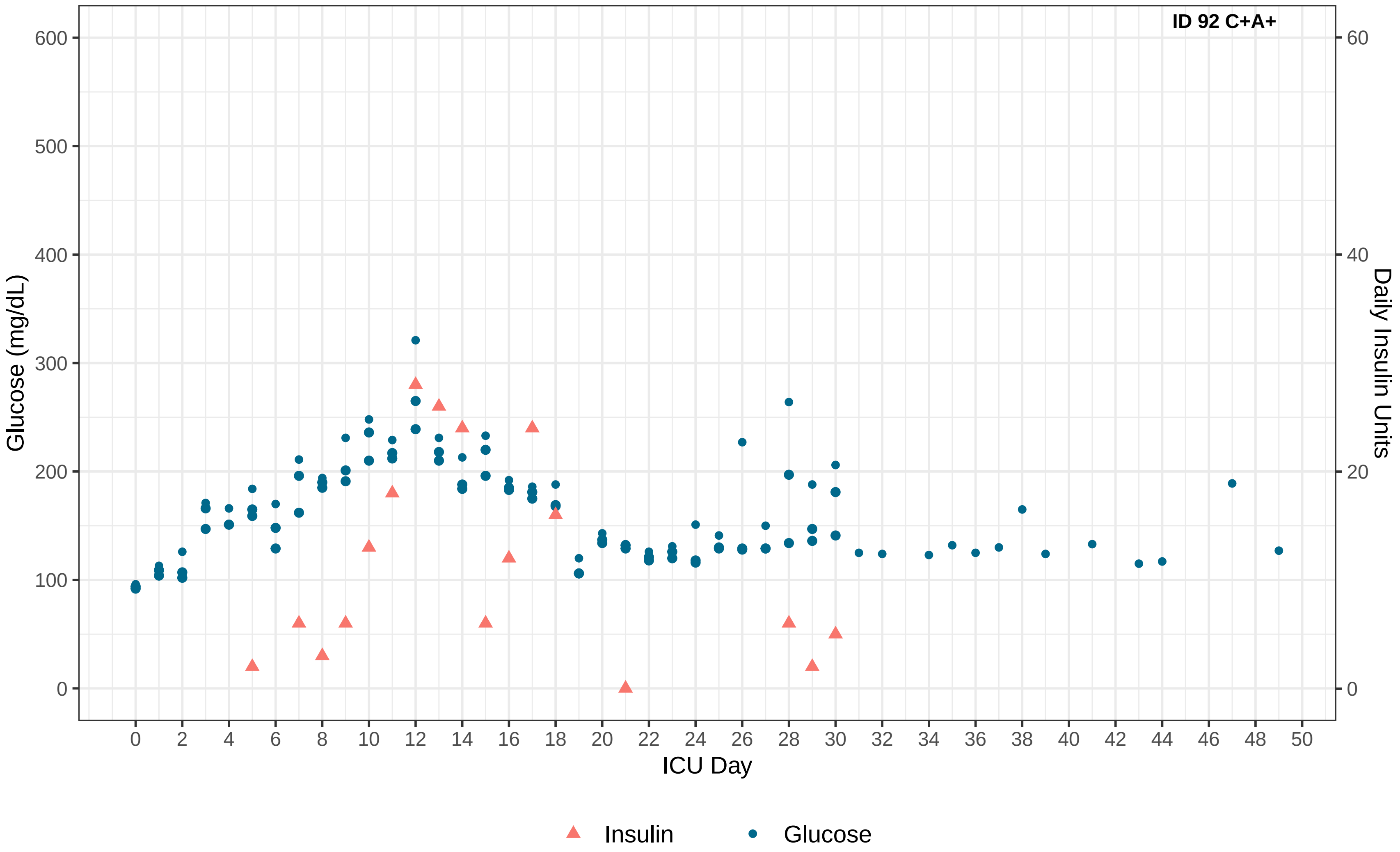
ID 92 C+A+
Glucose (mg/dL)
Daily Insulin Units
ICU Day
0
100
200
300
400
500
600
20
40
60
2
4
6
8
10
12
14
16
18
20
22
24
26
28
30
32
34
36
38
40
42
44
46
48
50
Insulin
Glucose

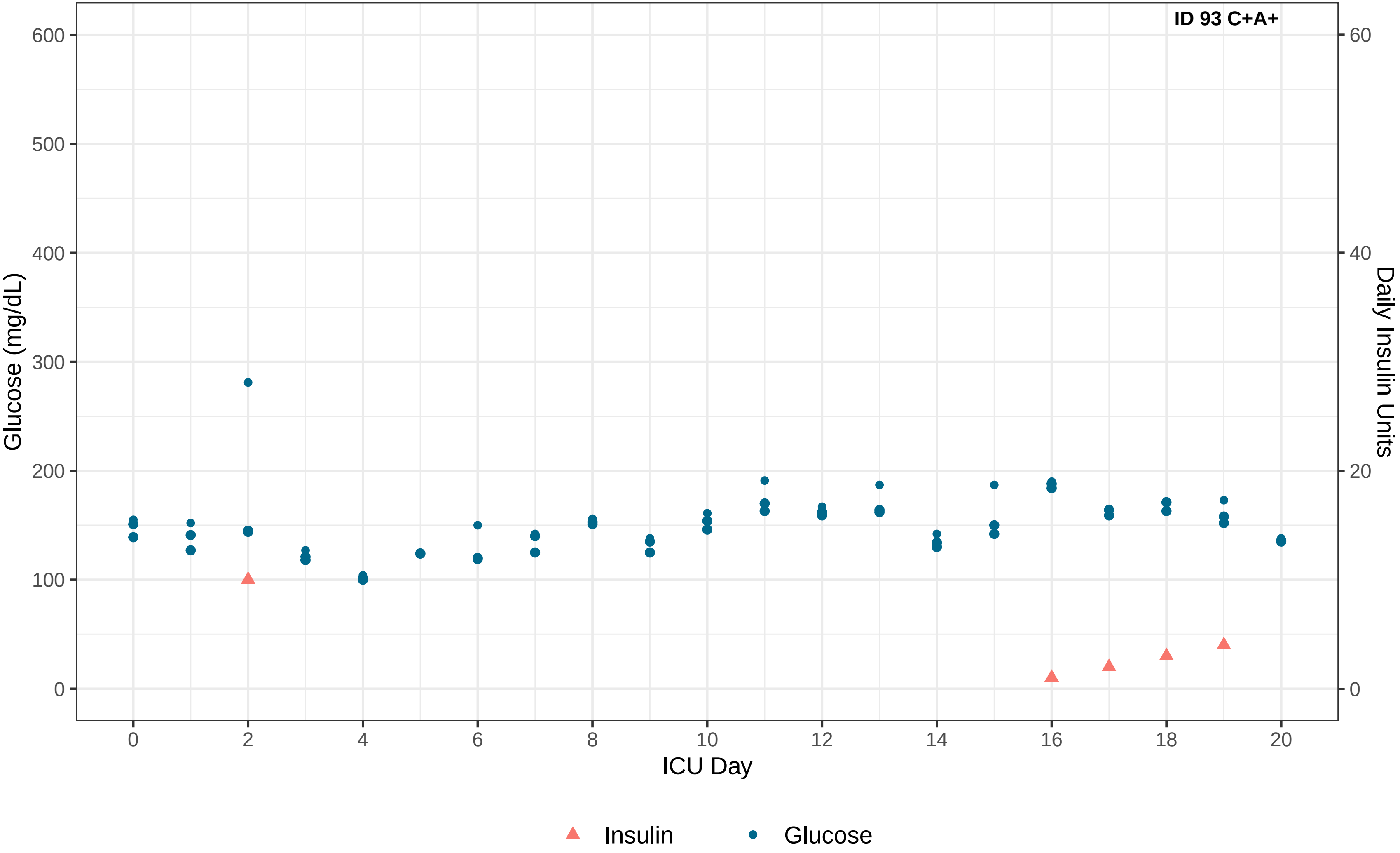
ID 93 C+A+
Glucose (mg/dL)
0
100
200
300
400
500
600
Daily Insulin Units
0
20
40
60
ICU Day
0
2
4
6
8
10
12
14
16
18
20
Insulin
Glucose

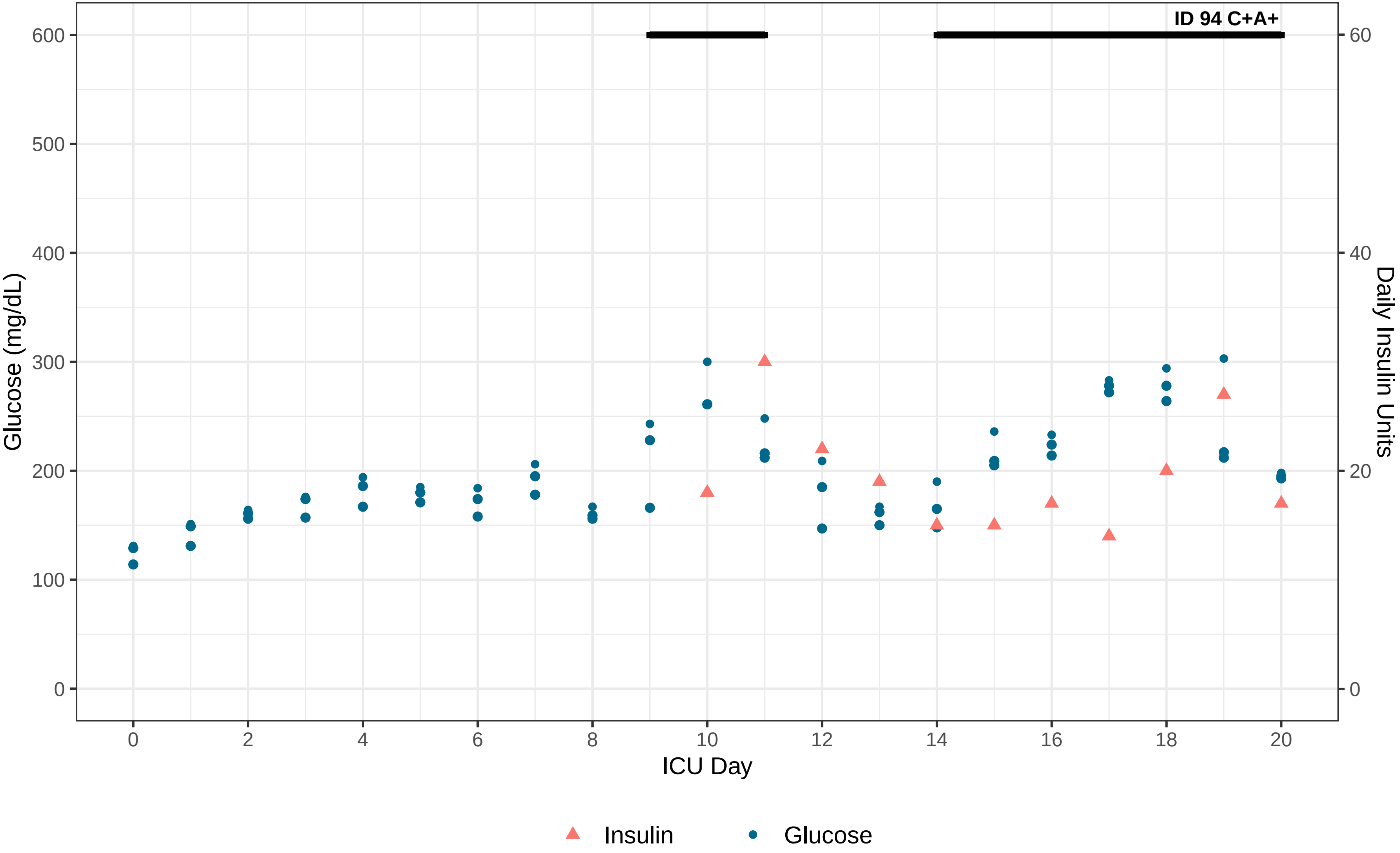
ID 94 C+A+
Glucose (mg/dL)
Daily Insulin Units
ICU Day
0
100
200
300
400
500
600
0
20
40
60
0
2
4
6
8
10
12
14
16
18
20
Insulin
Glucose

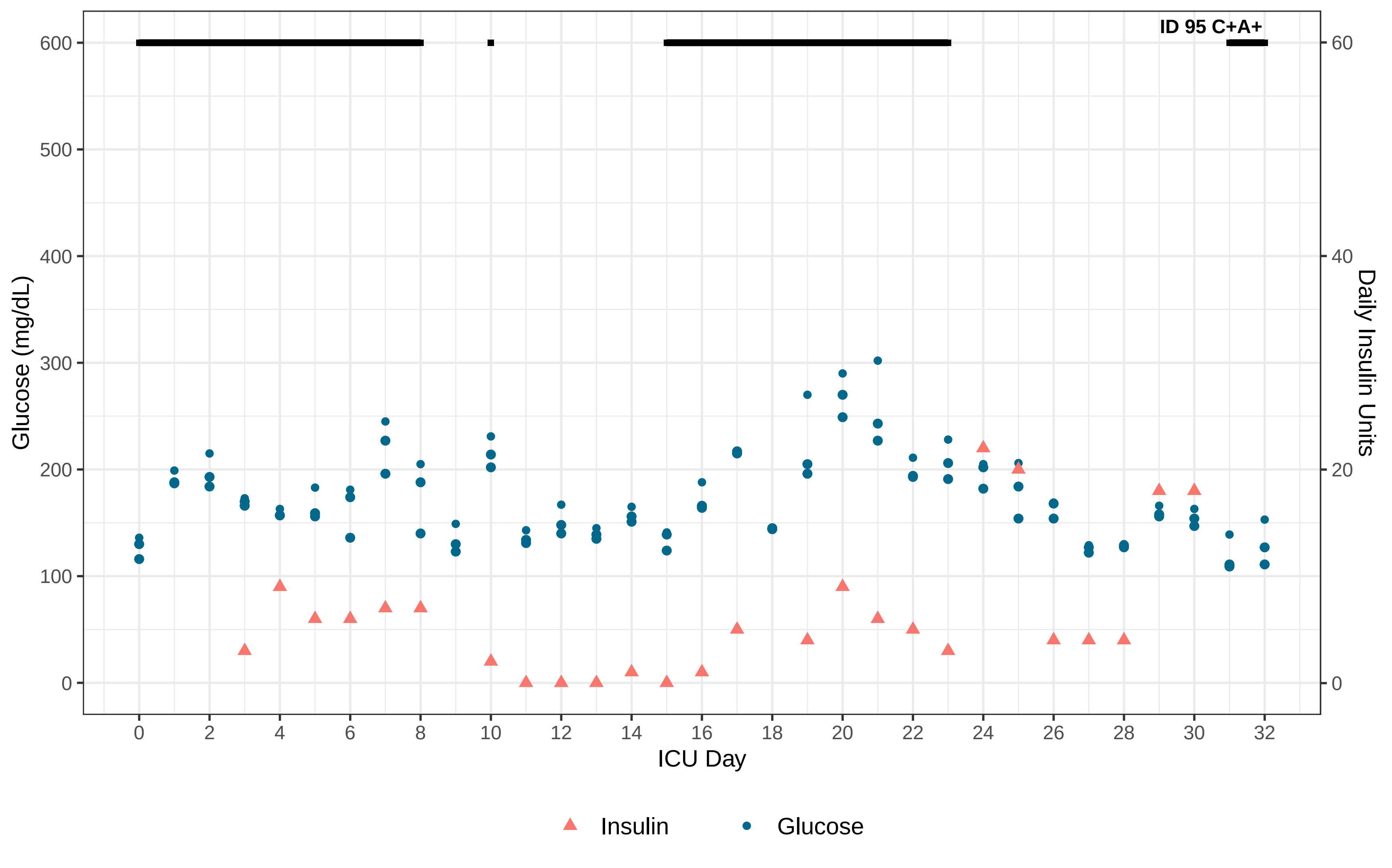

ID 95 C+A+
Glucose (mg/dL)
Daily Insulin Units
ICU Day
0
100
200
300
400
500
600
0
20
40
60
0 2 4 6 8 10 12 14 16 18 20 22 24 26 28 30 32
Insulin
Glucose

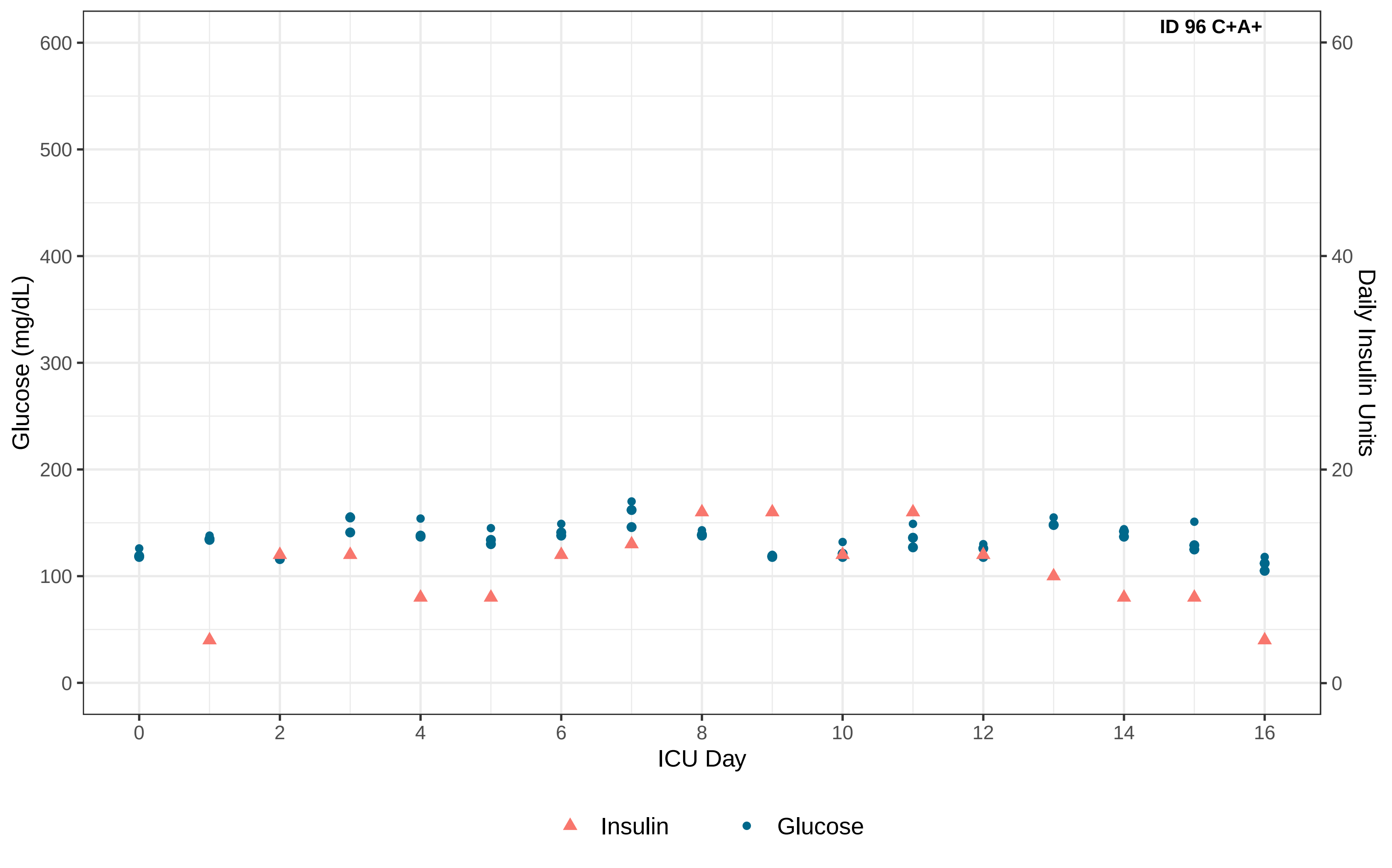

ID 96 C+A+
Glucose (mg/dL)
Daily Insulin Units
ICU Day
0
100
200
300
400
500
600
0
20
40
60
0
2
4
6
8
10
12
14
16
Insulin
Glucose

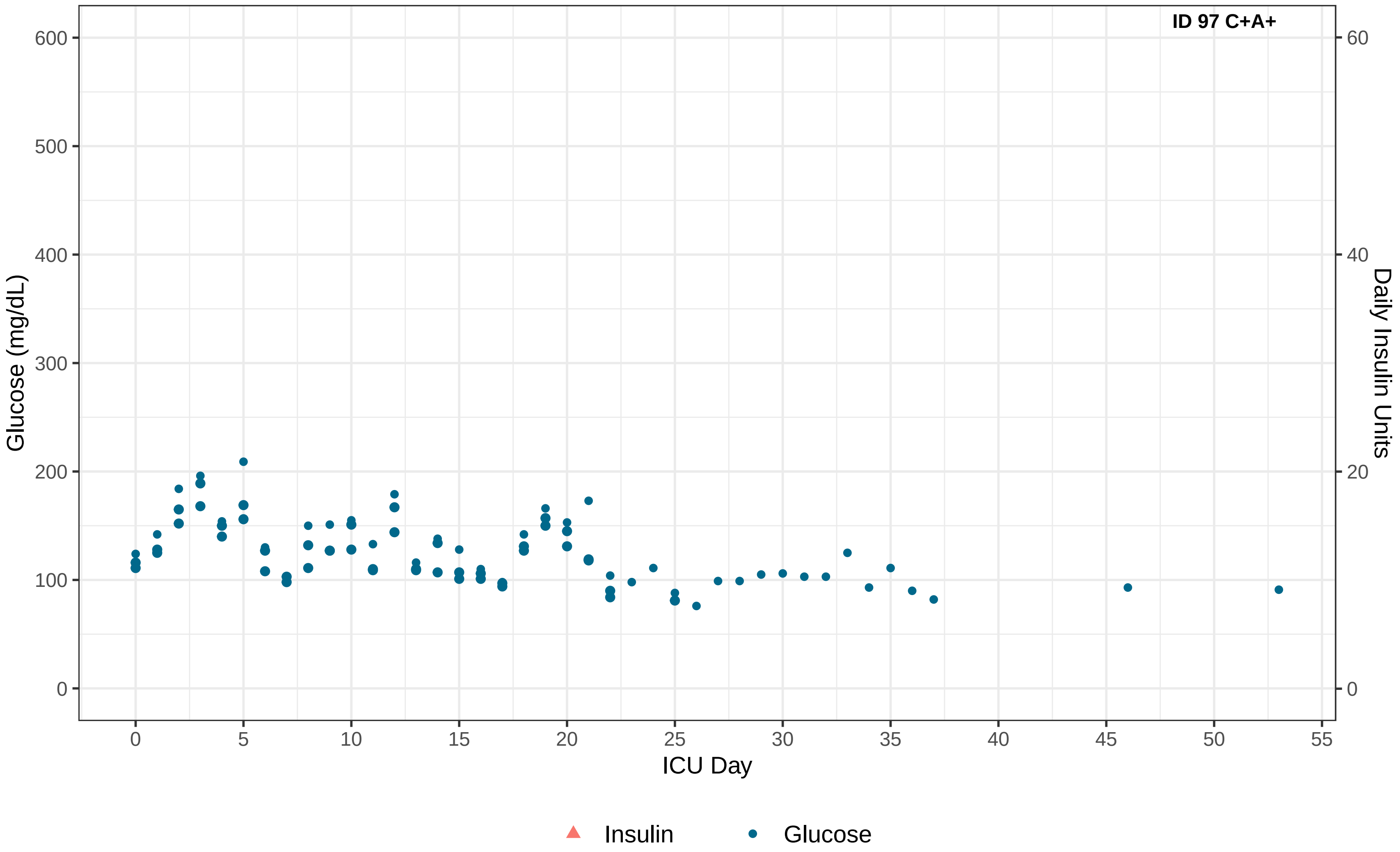

ID 97 C+A+
Glucose (mg/dL)
Daily Insulin Units
ICU Day
0
100
200
300
400
500
600
0
20
40
60
0
5
10
15
20
25
30
35
40
45
50
55
Insulin
Glucose

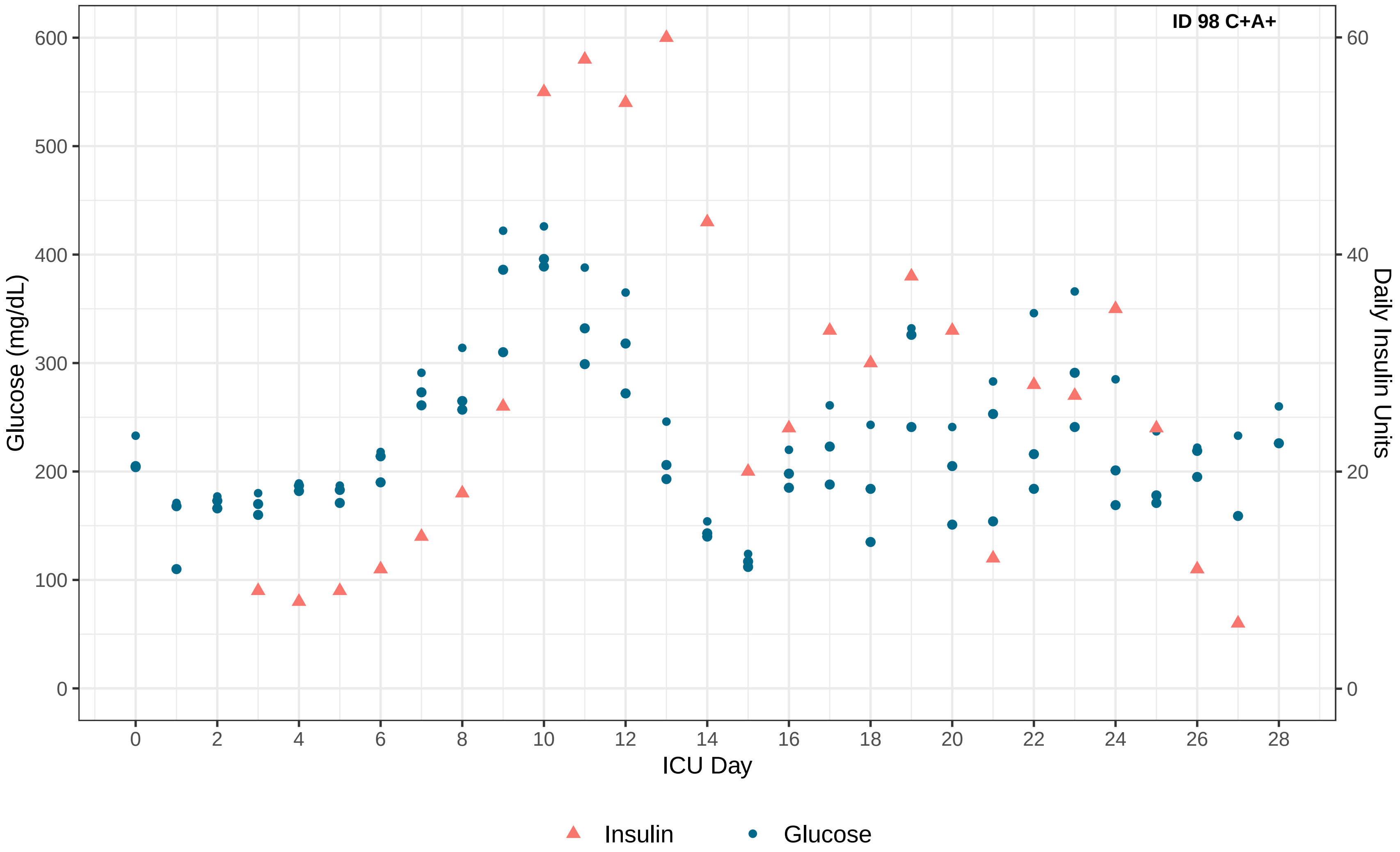
ID 98 C+A+
Glucose (mg/dL)
Daily Insulin Units
ICU Day
0
100
200
300
400
500
600
0
20
40
60
0 2 4 6 8 10 12 14 16 18 20 22 24 26 28
Insulin
Glucose

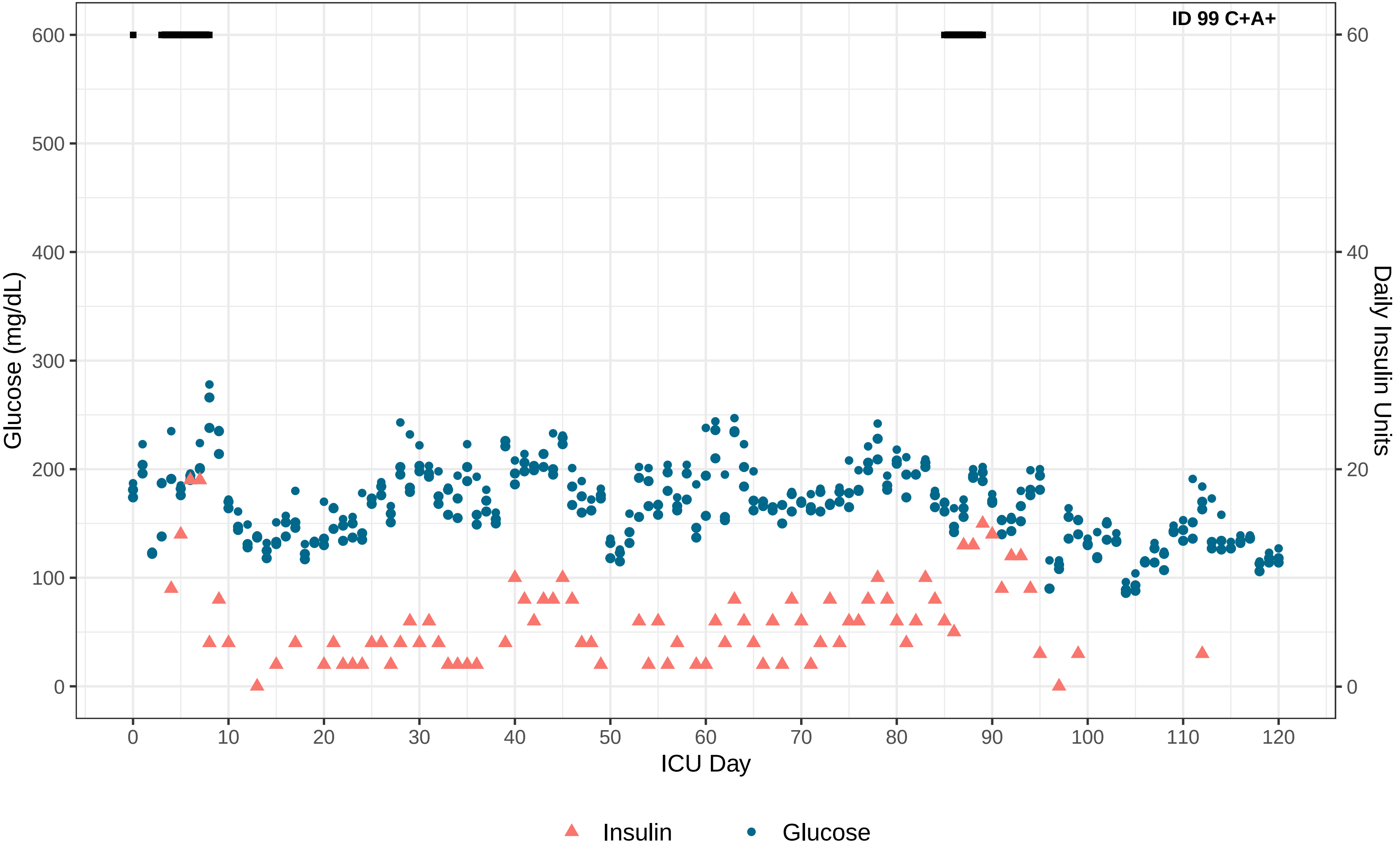

ID 99 C+A+
Glucose (mg/dL)
600
500
400
300
200
100
0
Daily Insulin Units
60
40
20
0
0
10
20
30
40
50
60
70
80
90
100
110
120
ICU Day
Insulin
Glucose

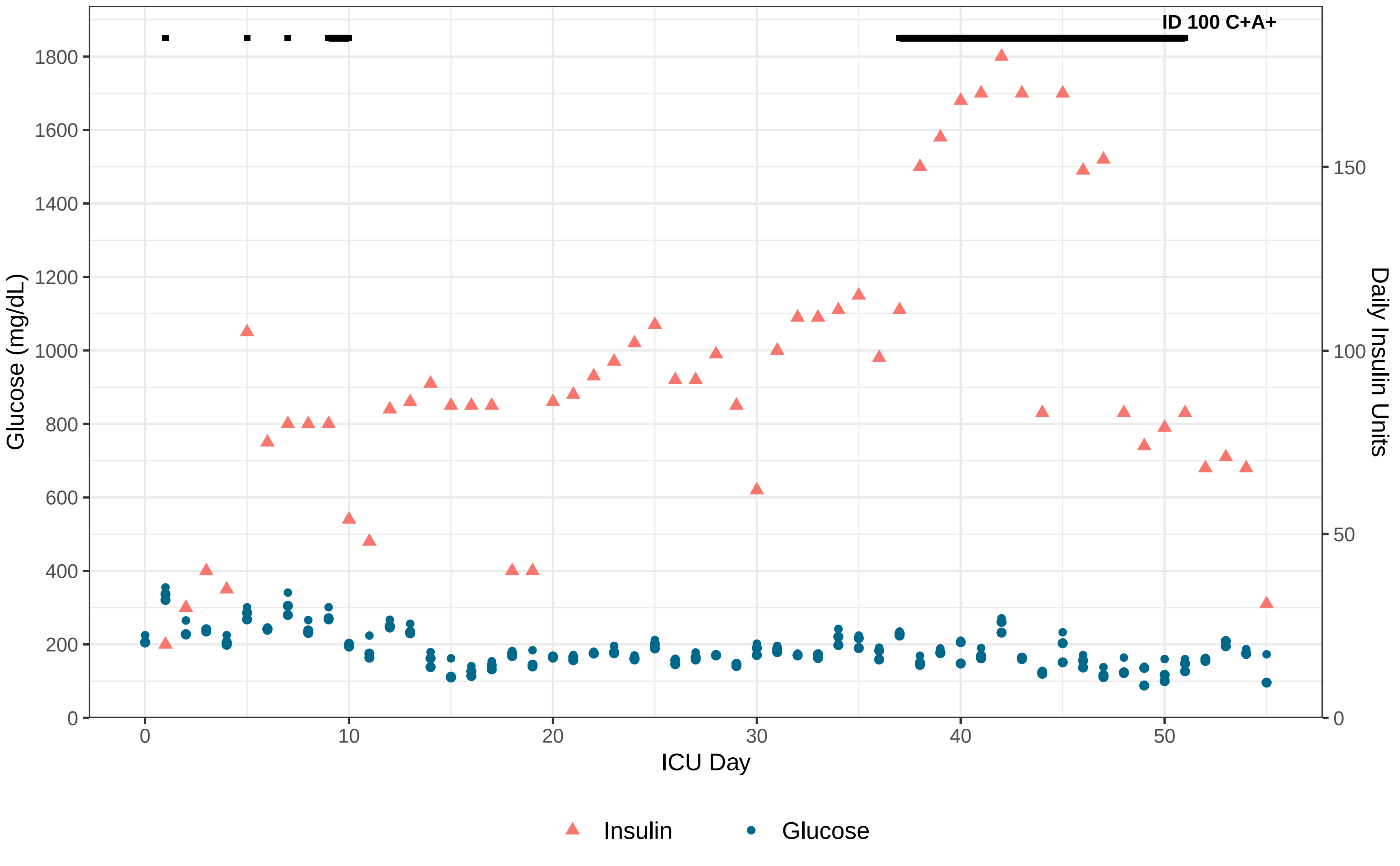
ID 100 C+A+
Glucose (mg/dL)
0
200
400
600
800
1000
1200
1400
1600
1800
Daily Insulin Units
0
50
100
150
ICU Day
0
10
20
30
40
50
Insulin
Glucose

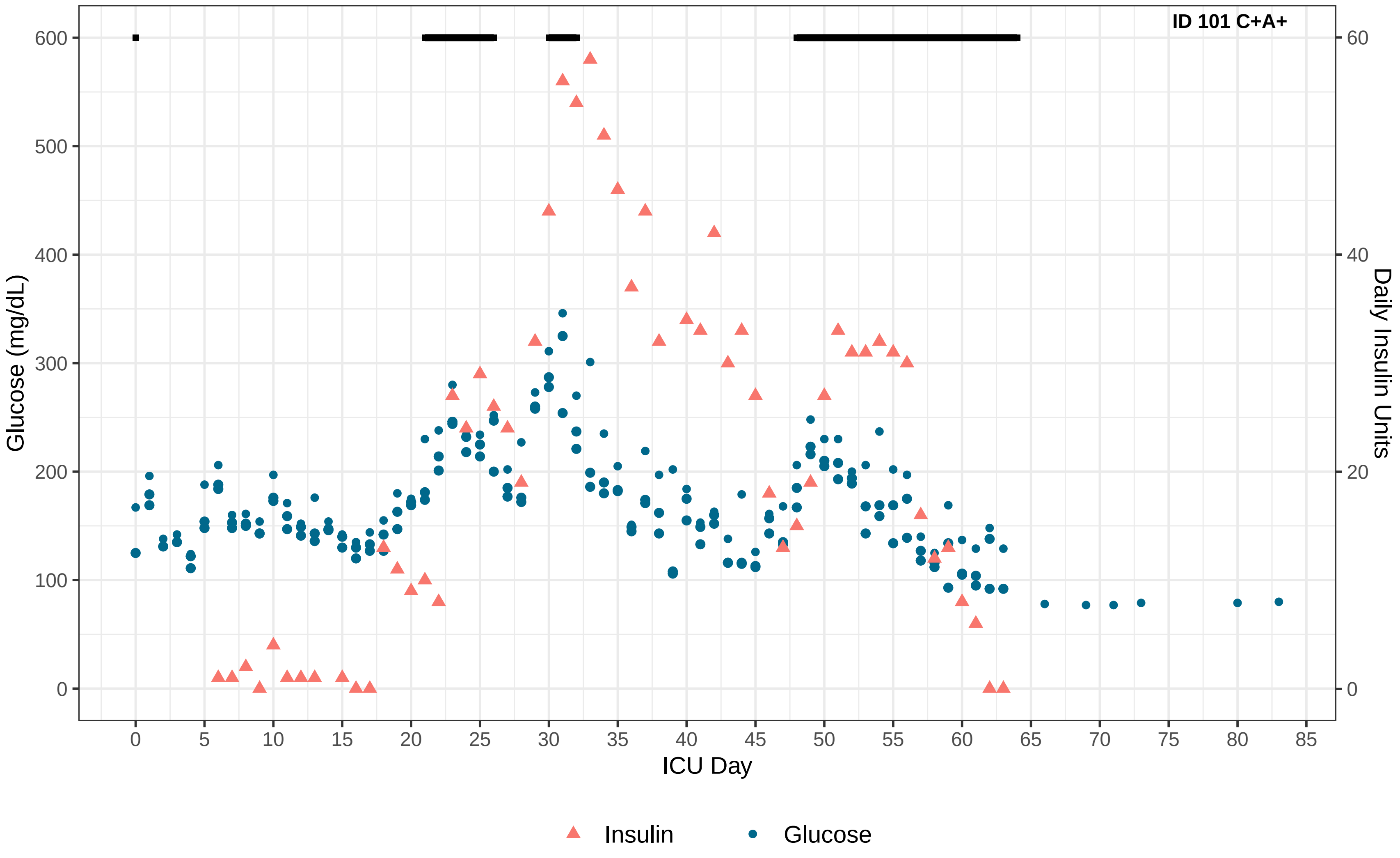
ID 101 C+A+
Glucose (mg/dL)
Daily Insulin Units
ICU Day
0
100
200
300
400
500
600
0
20
40
60
0
5
10
15
20
25
30
35
40
45
50
55
60
65
70
75
80
85
Insulin
Glucose
